# Non-generative readout with few-label calibration versus LoRA fine-tuning of a large language model for ASA physical status classification: accuracy, speed, cost and label efficiency in three data sources

**DOI:** 10.64898/2026.10.20.26366028

**Authors:** Yutong Shi, Yang Wang, Alimujiang Simayi, Li Qu, Guiping Xu

## Abstract

**Background:** The American Society of Anesthesiologists Physical Status (ASA-PS) class underpins perioperative risk communication, but its assignment is subjective. Generative large language models (LLMs) are slow and costly, and LoRA fine-tuning needs site-specific labels and a separately maintained adapter for every site. Non-generative decision models return option probabilities in one forward pass, but their comparison with LoRA in anesthesia, and the number of local labels they need, is unstudied.

**Objective:** To compare, on one open base model, generative zero-shot prompting, a non-generative readout, the readout with cutpoint calibration on a few local labels, and task LoRA for four-class ASA-PS (I, II, III, IV–V) in accuracy, speed and label requirement.

**Methods:** Retrospective study, not preregistered. MOVER (57,473 operations) was the development source, the local hospital’s 2025–2026 operations (n = 2,753) the temporally external validation set, and VitalDB (n = 6,242) a second site. The base model was Qwen3.5-9B; input was preoperative structured variables (T1) or T1 plus diagnosis text (T2). The primary metric was quadratic-weighted κ against each site’s recorded ASA class, with patient-level cluster-bootstrap intervals; the non-inferiority margin was 0.05. Cost means tokens and GPU time.

**Results:** In the local validation set the readout reached κ = 0.460 versus 0.227 for generative zero-shot prompting with a requested rationale (difference +0.234, 95

**Conclusions:** The non-generative readout needs no training and ran about 64 times faster than generative prompting with a rationale. A few hundred local labels bring it close to LoRA in κ without training and storing a per-site adapter, although κ of 0.52–0.58 is not evidence of human-level performance. LoRA keeps a higher κ once several hundred or more labels are available, and higher exact agreement. Non-inferiority holds for the prespecified analyses and sits at the margin in stricter sensitivity analyses, so it should be read as conditional.

## 1. Introduction

The ASA-PS class summarizes a patient’s preoperative systemic disease burden and is used for perioperative risk communication, resource allocation, quality assessment and research stratification [1]. It depends on the rater’s judgment, raters disagree with one another [2], and institutions differ in habit. An automated grading method should therefore take each institution’s existing records as the reference and answer a practical question: how many local annotations are needed before it can be used at a new site? Put differently, when a new site has only k labels, should one calibrate a frozen readout or train a site-specific low-rank adapter?

Large language models have been applied to ASA-PS grading and related perioperative prediction. On 21,049 MOVER operations, GPT-4o with few-shot and feature prompting reached 66% accuracy against 54% for LightGBM [3]; a retrieval-augmented LLaMA-3.1-8B reached an ASA micro-F1 of 0.841 on 24,491 cases against 0.827 for XGBoost [4]. Other studies compared language models with clinicians in small samples [5, 6], and a commentary pointed to unresolved validity threats in LLM-based ASA grading [7]. The generative route has three deployment costs. Output is decoded token by token, so latency and expense grow with output length. Generated text can contradict the input: clinical documentation tasks showed a 1.47% hallucination rate and a 3.45% omission rate [8], and under adversarial prompts containing fabricated details six models hallucinated in 50%–82% of cases [9]. Classification also requires parsing text into a class, and token probabilities are not calibrated class probabilities; in an evaluation of diagnostic probabilities from electronic health records, logit-based estimates were worse than conventional classifiers, and the authors called for better calibration [10].

To raise task accuracy and allow local deployment, small open models are commonly fine-tuned with LoRA. In perioperative complication detection, the micro-F1 of a 4B model rose from 0.28 to 0.64, with LoRA contributing 0.103 [11]; in ASA-PS grading, a LoRA-tuned LLaMA-3 reached a micro-F1 of 0.780, below the 0.815 of XGBoost in the same study [12]. This route has costs too. It needs a labeled, site-specific training set, and each site must train and store its own adapter. The compute is not trivial for most institutions: each fine-tuning run of the clinical language model NYUTron took about 6 hours on 8 A100 GPUs, which the authors considered out of reach for most research groups [13]. A model fine-tuned on single-institution data can degrade in external populations, and joint multi-institution adaptation is limited by data-sharing restrictions, which has prompted sequential adapter training and federated LoRA schemes [14, 15].

Non-generative (System One) decision models offer another route that changes how the model gives its answer. Given a typed input, a question and candidate options, the model returns option probabilities in one forward pass, generates no text, and cannot answer outside the given closed set [16]. We use Jev for the hosted, closed decision model (base model and training details undisclosed) and OpenJev for an open implementation compatible with its request format. Recasting classification as scoring predefined candidate words with a language model was proposed earlier for few-shot text classification [17, 18]; the readout used here belongs to the same family, differing in that site adaptation is done with ordinal cutpoints instead of fine-tuning. The task description and options are written into the request in natural language, so the model works without task-specific training. Published evaluations report advantages in speed and cost: median latency of 0.27–0.31 s and a total cost of about US$0.08 for 2,823 items on medical question-answering and case-reasoning benchmarks [16]; and US$0.20 to screen 4,527 records for a systematic review, against US$2.59 and US$33.45 for two generative LLMs on the same task [19]. Open-source work shows that reading the output logits at the option letters of an off-the-shelf instruction-tuned model gives comparable decision ability, so fine-tuning is optional rather than required [20, 21], and an open decision model needs about 0.1 s per query [22]. The limits are also documented: performance approaches mainstream models on text-dominated tasks but is weaker on numeric-dominated ones [23]; it falls below frontier generative models on medical examinations and complex diagnostic cases [16]; and the claim of calibrated probabilities needs testing on clinical data, since a preregistered evaluation found lower accuracy than comparators on synthetic multi-symptom patients and zero probability assigned to the correct diagnosis in 6.2% of cases [24].

These evaluations mostly address medical question answering, diagnostic reasoning or literature screening. In a search of PubMed, arXiv and Semantic Scholar (October 2026) we found no study that compared a non-generative readout with a LoRA adapter on the same base model for ASA-PS grading in real electronic health records, in terms of speed, training cost and number of local labels, or that tested whether calibration on a few local labels can close the gap between the zero-shot readout and LoRA.

We therefore compared, on one open base model, generative zero-shot prompting, a non-generative readout (including OpenJev, an open System One implementation), ordinal cutpoint calibration on a few local labels, and task-specific LoRA. The outcome was four-class ASA-PS accuracy; speed, cost and label requirement were recorded alongside. Data came from MOVER, the local hospital and VitalDB, with two input types. The prespecified hierarchy of hypotheses was: H1, the non-generative readout is superior to generative zero-shot prompting; H2, the readout calibrated on 1,000 local labels is non-inferior (margin 0.05) to LoRA trained on 1,000 labels; H3, LoRA trained on all local labels is superior to the calibrated readout. We also tested whether the readout’s output can be steered by natural-language guidance, and quantified the token consumption and training cost of each approach.

## 2. Methods

### 2.1. Study design, reporting guideline and registration

This is a retrospective diagnostic/decision-model comparison, reported according to TRIPOD-LLM [25] (study type: LLM evaluation with task fine-tuning; task: classification); the item-by-item self-check is in Supplementary S1. No outcome was annotated by hand: the outcome is the ASA class recorded in each site’s anesthesia record (the manual check in Section 2.7 served only as methodological quality control). No separate study protocol was published; the analysis plan was kept as an internal frozen document (Supplementary S2). Patients and the public were not involved. **The study was not preregistered.** The analysis plan was frozen before each stage of data analysis (file hash in Supplementary S2), but some analyses were added after earlier results were seen; all deviations and post hoc analyses are listed in Section 2.10.

### 2.2. Data sources

#### MOVER

(operating-room database of the University of California, Irvine, USA) [26]: the patient table holds 64,354 operation IDs; 57,504 operations had an ASA class of I–V, of which 57,473 (38,245 patients) were in adults aged 18 years or older. Patients were randomly split into a development set (80%, 45,994 operations) and a test set (20%, 11,479 operations). Evaluation used a random patient-level subset of the test set (seed 20261018, 4,000 operations, 2,606 patients), chosen because of compute limits. MOVER was used under its data use agreement. Operation dates range over 2017–2023 (mainly 2018–2022).

#### Local hospital

(People’s Hospital of Xinjiang Uygur Autonomous Region; abdominal and pelvic surgery): the anesthesia overview contained 8,852 operations, of which 7,348 (6,828 patients) had both an anesthesia start time and an ASA class. Excluding patients under 18 years left 7,332 operations; requiring a preanesthesia visit within 30 days before surgery left 7,079 operations (6,804 patients) from 2021–2026 (one 2020 operation was assigned to the development set). By year, 2021–2024 formed the development set (4,326 operations) and 2025–2026 the temporally external validation set (2,753 operations, 2,640 patients); no patient appears in both.

#### VitalDB

(Seoul National University Hospital, public database) [27]: 6,388 noncardiac operations; after excluding missing ASA and ASA 6, 6,242 operations (5,961 patients) remained and were split by patient into a training set (70%, 4,357 operations) and a test set (30%, 1,885 operations, 1,788 patients). The data are licensed CC BY-NC-SA 4.0 and were used for non-commercial academic purposes; operations took place between August 2016 and June 2017 [27].

Inclusion, exclusion and splits are shown in Fig. 2, cohort characteristics in Table 1, and comparisons between development and evaluation splits in Supplementary Table S12. Table 1 covers all included operations of each source; the evaluation sets used in the Results (local validation set, MOVER evaluation subset, VitalDB test set) have different denominators, which are stated where used.

**Fig. 1.**
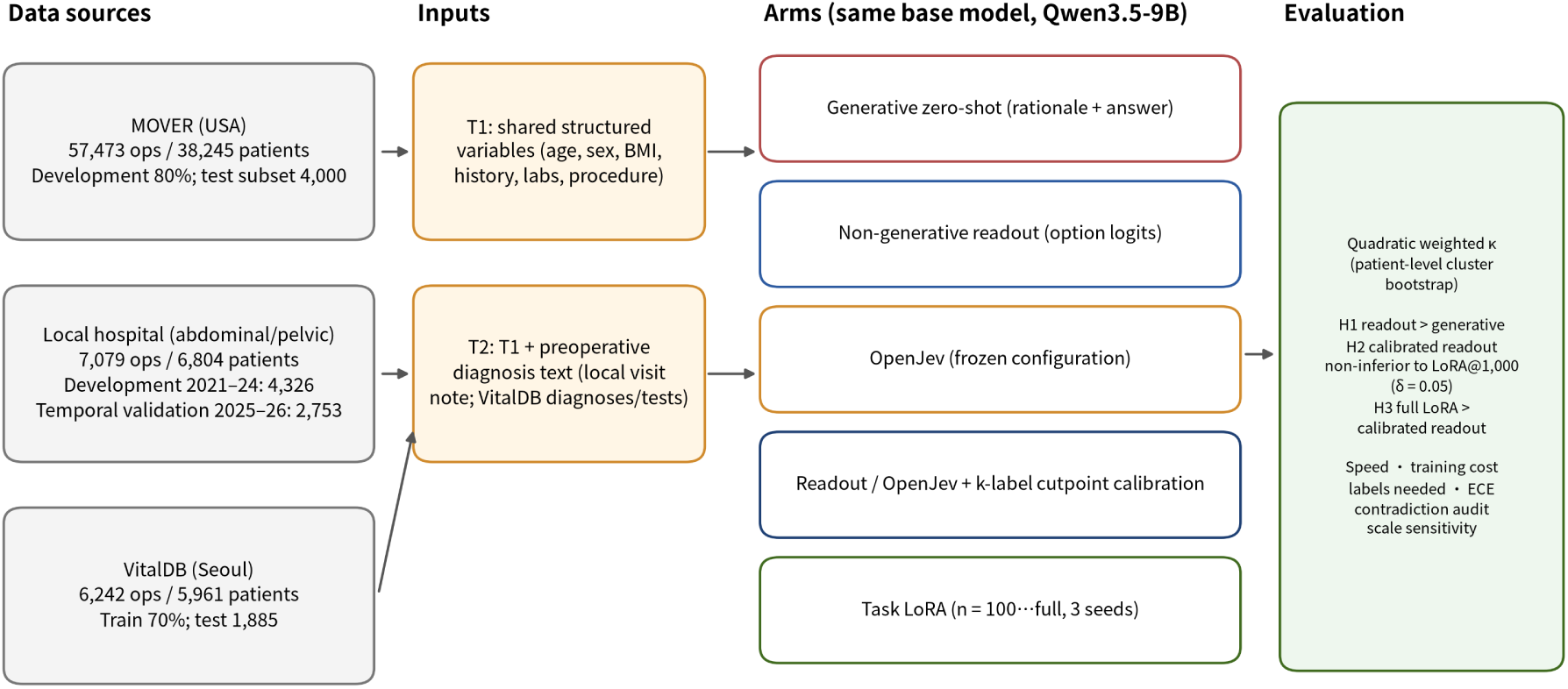
Study design and data flow. Three data sources (MOVER, local hospital, VitalDB) enter through T1 (shared structured variables) or T2 (plus preoperative diagnosis text); on one base model, Qwen3.5-9B, generative zero-shot prompting, the non-generative readout, OpenJev, cutpoint calibration with k labels and task LoRA are compared; H1–H3 are assessed by quadratic-weighted κ against the ASA class in each site’s anesthesia record, and speed, training cost and label requirement are reported. T1: the shared variables listed in Section 2.3; T2: T1 plus preoperative diagnosis text.

**Fig. 2.**
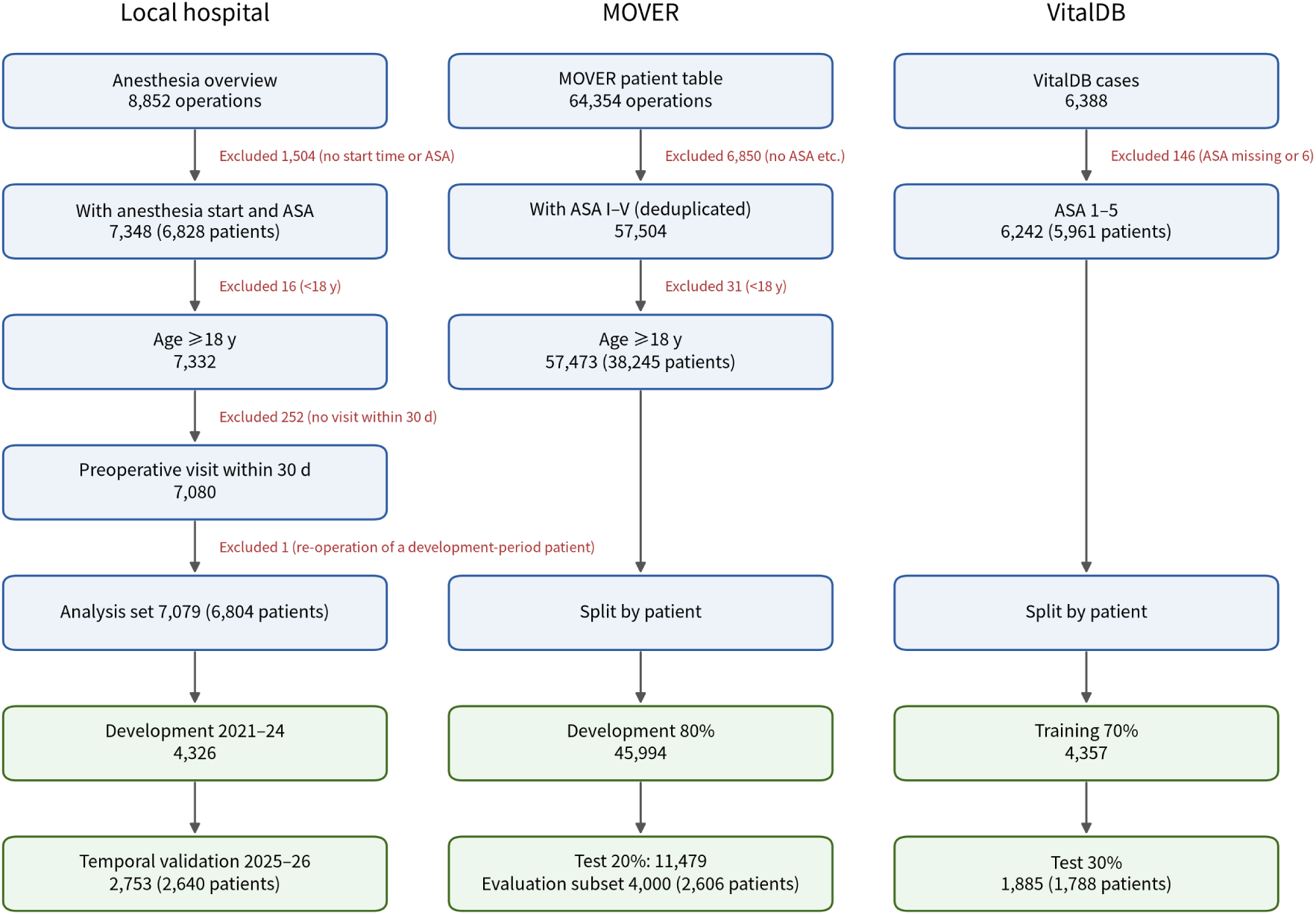
Flow of inclusion, exclusion and splitting. Stepwise exclusions and sample sizes (operations; patients in parentheses) from raw records to development and evaluation sets in the three data sources.

**Table 1.** Cohort characteristics of the three data sources.

| Data source | Operations | Patients | Age, median (IQR) | Female | BMI, median (IQR) | Inpatient | Emergency | ASA I/II/III/IV–V (%) |
| --- | --- | --- | --- | --- | --- | --- | --- | --- |
| MOVER (UCI operating rooms, USA) | 57,473 | 38,245 | 57 (41–68) | 47.7% | 27 (23–31) | 64.4% | — | 5.1 / 31.4 / 51.2 / 12.2 |
| Local hospital (abdominal and pelvic surgery, 2021–26) | 7,079 | 6,804 | 54 (46–65) | 68.6% | 25 (22–28) | 100% | 18.7% | 1.3 / 46.0 / 50.0 / 2.6 |
| VitalDB (Seoul National University Hospital, noncardiac surgery) | 6,242 | 5,961 | 59 (48–68) | 49.4% | 23 (21–25) | — | 11.6% | 28.7 / 59.3 / 11.3 / 0.8 |

Note: ASA mix is given as percentages for I / II / III / IV–V (class V merged into IV); “—” means the source lacks the variable; IQR, interquartile range; BMI, body mass index. Table 1 covers all included operations of each source; the evaluation sets used in the Results have different denominators (local validation set 2,753, MOVER evaluation subset 4,000, VitalDB test set 1,885; H1 additionally restricted to the subset with parsable generative output). Comparisons between development and evaluation splits are in Supplementary Table S12.

### 2.3. Outcome and inputs

#### Outcome

the ASA class in each site’s operative anesthesia record, merged into four classes (I, II, III, IV–V; class V merged into IV). The record is the assessment made by that site’s anesthesiologist in routine care. We did not relabel it and make no claim about its agreement with a “true” ASA class.

#### Input

only preoperatively available information was used. T1 comprises the structured variables common to all three sources, expressed in one English template: age, sex, weight, BMI, inpatient or outpatient status, the most recent preoperative blood pressure and heart rate, hypertension, coronary disease, diabetes history, preoperative creatinine, hemoglobin, albumin, and planned procedure name (missing variables are written as “unknown”). T2 adds preoperative diagnosis text to T1: for the local hospital this is the raw preanesthesia visit note (ASA wording masked); for VitalDB, diagnosis, procedure name, department, approach, anesthesia type, emergency flag, electrocardiogram, pulmonary function and further preoperative laboratory results; for MOVER, T2 uses the undated diagnosis history, which may include postoperative diagnoses and is therefore exploratory only. T1 is an English template whereas the local T2 text is Chinese, so the two differ in language, format and information content at once. To separate these, the T1 template was also translated into Chinese (variable names and values) and read out once with an English and once with a Chinese prompt shell in the local validation set (Section 3.5). **Pre-processing and missing data**: local creatinine was converted from μmol/L to mg/dL (divided by 88.4), and hemoglobin and albumin from g/L to g/dL (divided by 10); ASA wording in the local visit notes was masked (two notes contained it, none remained after masking); local procedure names were kept in the original Chinese. Missing values were not imputed but written as “unknown” in the template, so the model and the cutpoint calibration face missingness directly (MOVER availability of preoperative creatinine, hemoglobin and albumin is about 40%, 42% and 31%, and BMI is missing in 20.7%; VitalDB has no preoperative blood pressure or heart rate values and no coronary disease flag). Class imbalance (low share of ASA I and IV–V) was not handled by resampling or weighting; the metric is quadratic-weighted κ, which penalizes adjacent-class errors less.

### 2.4. Comparison arms (one base model, Qwen3.5-9B [28]; readout and generative zero-shot in fp16, LoRA in 4-bit NormalFloat, NF4)

1. **Generative zero-shot**: the model was prompted to give a rationale of at most three sentences followed by a final answer in a fixed format, and the parsed answer served as the hard label; greedy decoding, at most 140 new tokens; unparsable outputs were counted as missing and their proportion reported. This arm’s prompt asks for a rationale and does not list the options, which differs from the readout prompt, so H1 compares the full generative usage with a closed-set readout and is not a single-factor comparison of interfaces. Because of compute limits, the generative arm was run only on 1,000 randomly drawn local validation operations (seed 20261019) and on the first 2,000 operations of the MOVER evaluation subset; 999 and 1,978 were parsable, and the other 1 and 22 were excluded as missing. Comparability of subset and full set is addressed in Section 3.2.
2. **Non-generative readout (in-house)**: the same prompt followed by the options (A. ASA I; B. ASA II; C. ASA III; D. ASA IV–V); the logits of the four option letters at the last position were passed through softmax to give closed-set probabilities, in a single forward pass with no text generation. The primary analysis used the natural order above (I→IV–V), fixed before any result was seen. To test the influence of option order, six orders (natural, reversed, three cyclic shifts and one random order) were each read out once, the positional probabilities were mapped back to classes and averaged, and the cutpoints were refitted (Section 3.9).
3. **OpenJev (open System One implementation)**: GitHub30/OpenJev, MIT license, commit aae11e1, with two compatibility patches (the `quantization_config` argument for 4-bit loading; `enable_thinking=False` in the chat template, because otherwise the thinking tokens of the Qwen3.5 template degrade the output). Prompt and question type were chosen and frozen on an independent tuning set of 800 MOVER development operations, from eight predefined configurations (full prompt texts in the supplement): an anesthesia system prompt, one-sentence option descriptions and a score question type (2-fold cross-validation with calibrated κ as the criterion; scores of all eight configurations in Supplementary S5).
4. **Cutpoint calibration**: for any zero-shot arm, the expected score E = Σ k·p_k was computed, and three cutpoints were fitted on k local labeled cases by coordinate search to maximize quadratic-weighted κ; the remaining cases were graded by these cutpoints. No model parameter was updated.
5. **Task LoRA**: 4-bit NF4 quantization (QLoRA [29]), LoRA [30] rank 16, α = 32, dropout 0.05, on all attention and feed-forward linear layers; AdamW, learning rate 1×10^−4^ (one-cycle cosine), effective batch size 16, one epoch (in line with a published LoRA study on the same task, where the validation set had converged after one epoch and more epochs gave no clear gain [12]); the loss was cross-entropy on the logits of the four option letters. Local training label numbers were n = 100, 300, 1,000 and all; for n ≤ 1,000 and for the full local set three seeds were used (20261017, 11, 12), and the same three seeds for MOVER at 3,000 and 12,000.
6. **Supplementary arms**: XGBoost (same structured features); retrieval-based few-shot kNN-8; OpenJev+LoRA (decision model plus LoRA, trained with its calibrated scoring rule; the loss variant was chosen on the independent MOVER tuning set); scale sensitivity (Qwen3.5-27B, Qwen3.8-27B, 4-bit); and an fp16 LoRA control (local n = 1,000, seed 11, the same labels as the 4-bit LoRA; the fp16 base model was split by layer across two cards, same hyperparameters, fp16 automatic mixed precision with gradient scaling).

#### 2.4.1. Prompt design and data used

The readout and generative prompts consist of a one-sentence role statement plus the input template and options. Prompt variants were compared only on 1,500 random MOVER development operations (three prompts; Section 3.9 and Supplementary S6). A single trial run on 200 random local development operations measured speed and output form (not a performance evaluation); the sealed local validation set and VitalDB took no part in prompt selection. Prompts were frozen before the evaluation sets were touched. All models were run from 17 to 20 October 2026 using the weights of Qwen/Qwen3.5-9B from the Hugging Face hub (post-trained version, Apache-2.0), downloaded on 16 October 2026; the SHA-256 checksums of the weight files accompany the supplement.

#### 2.4.2. Natural-language guidance (readout arm, no training, no labels)

A paragraph of natural-language guidance was added before the readout prompt, everything else unchanged, and four conditions were compared in the local validation set (n = 2,753): no guidance; the official ASA 2020 definitions with examples; the local recording habit (based on the principal investigator’s description of local practice: ASA I is rarely recorded, apparently healthy patients undergoing oncologic or major abdominal surgery are mostly recorded as II, older patients with organic changes mostly as III, the rest being general clinical common sense); and strict adherence to the guideline (opposite in direction to the former). The wording of the four paragraphs was fixed before validation results were seen and never changed; 800 random local development operations were used only to check direction. Inputs were T1 and T2. The interpretation rule fixed in advance was: if the “local habit” guidance moves the predicted class mix close to the original records and raises κ, natural language can replace part of the labels; if the mix changes but κ does not rise, the output can be steered without becoming more accurate; if the mix does not change, the property is not supported. In addition, under each guidance condition cutpoints were fitted on 100 development labels (60 repetitions) to see whether guidance and calibration add up.

### 2.5. Obtaining the calibration labels

Local calibration labels came from 1,000 random development operations (seed 11), MOVER labels from 1,000 random operations in the development split (seed 11), and VitalDB labels from 1,000 random training-set operations (seed 11). For the label-number curves, k {25, 50, 100, 200, 500, 1,000} labels were drawn from this pool with 100 repetitions (60 to 100 for VitalDB). LoRA training labels were drawn from the same development set by seed: the 1,000 training labels of seed 11 are identical to the calibration pool (checked for both the local hospital and MOVER), whereas the training pools of the other two seeds overlap partly with the calibration pool (22%–24% locally, about 2% in MOVER). The cutpoint search grid used by the curve script is coarser (61 points, 2 passes); at k = 1,000 it gives a local κ of 0.540, 0.006 above the 0.534 of the finer grid of the main analysis (121 points, 3 passes), a difference due to search precision; the abstract and the text use 0.534 throughout. The prespecified contrast used seed 20261017, whose training pool differs from the calibration pool and is therefore not label-matched; the label-matched contrast is seed 11. To estimate the uncertainty from label sampling, 50 independent 1,000-label pools were drawn from the local development set, and 50 pools from a further 5,000 operations of the MOVER training split; cutpoints were fitted in each and evaluated on the evaluation set.

### 2.6. Speed and cost

Cost was measured in three ways. Token consumption was counted with the tokenizer of the same base model, split into input tokens and, for generative arms, output (token-by-token decoded) tokens: input tokens were taken from the first 1,000 operations each of the local validation set and the MOVER subset, and generative output tokens were measured on the first 96 operations of the local generative subset (thinking mode off, as in the generative arm). Training cost was recorded both as the number of tokens processed in training (labels × input tokens per case, one epoch) and as actual GPU time. Tokens were not converted into floating-point operations or money.

The hardware was two NVIDIA GeForce RTX 2080 Ti cards with expanded memory (Turing, compute capability 7.5; 11 GB at the factory, 22,528 MiB per card as reported by nvidia-smi; linked by NVLink; configuration supplied by the authors). All main-analysis runs used a single card, the second being used for parallel queuing; only the supplementary fp16 LoRA control used cross-card placement with layers split across the two cards (model parallelism, not tensor parallelism). The architecture does not support bf16, so fp16 was used throughout; the linear-attention layers of Qwen3.5 ran without optimized kernels (flash-linear-attention and causal-conv1d were not installed, so the slower reference implementation was used). On a single card, we measured the median and 95th percentile of per-case latency at batch size 1, and throughput and peak memory at batch size 8; training cost is actual GPU time. To separate the effects of quantization and adapter, latency was also measured for the 4-bit readout (no adapter) and the 4-bit LoRA. “Cost” in this paper means computational cost (tokens and GPU time) and excludes annotation, personnel and monetary costs. OpenJev’s shared-prefix scoring path is incompatible with the linear-attention layers of Qwen3.5 and fell back on this machine to full-sequence scoring per candidate (about 0.8–1.3 s per case), so its latency is not compared with the design speed.

### 2.7. Screening generative output for contradictory content

One investigator carried out this check; there was no external reviewer beyond the review guide. A rule-based method flagged assertions in the generative rationale that contradict the structured input (negation handled clause by clause); 40 flagged rationales were drawn at random and read one by one to estimate the true contradiction rate. The method covers only structured fields and its recall is unknown. In addition, the saved rationales are truncated at the first 400 characters (77.5% of local and 90.4% of MOVER rationales were truncated), so the check covers only the first 400 characters of each. The resulting proportion is a lower bound.

### 2.8. Metrics and statistical analysis

The primary metric was quadratic-weighted κ; secondary metrics were accuracy, adjacent accuracy (prediction within one class of the record), the proportion of errors of two or more classes, macro-averaged area under the curve (AUC, one-versus-rest), expected calibration error (ECE) and Brier score (raw probabilities and after single-parameter temperature scaling). Intervals used a patient-level cluster bootstrap (B = 1,000), with paired differences computed on the same resamples. The non-inferiority margin for H2 was 0.05 (lower bound of the 95% CI of the difference > −0.05; non-inferiority is a one-sided test at α = 0.025, for which the lower bound of the two-sided 95% interval is the criterion), and H3 tests superiority. H1→H2→H3 were tested in a fixed sequence; all other analyses are secondary or exploratory and carry no confirmatory inference. The margin has no clinical anchor: in a published sample of 101 orthopedic patients, inter-clinician quadratic-weighted κ for ASA-PS was 0.713 [5], so 0.05 is about 7% of it (0.713 serves only as an order-of-magnitude reference, not as external justification); we therefore also report results for δ = 0.03, 0.05 and 0.10. For LoRA, seed-wise results and seed means are reported; the paired difference of seed means is the mean of the per-seed κ differences. The prespecified LoRA seed for pairing was 20261017; results of the other seeds are reported as robustness analyses. Analysis code was written in Python (scikit-learn, XGBoost, PEFT).

### 2.9. Investigator visibility and data roles

The MOVER development set was used to choose prompts, hyperparameters and the calibration method; the temporally external local validation set was evaluated after the plan was frozen. One point needs disclosure: predictions of the in-house readout and of XGBoost (trained on MOVER) for the 7,079 local operations had been generated before it was decided to “run MOVER first”, and the κ of 1,500 of them had been inspected; the settings of these methods had been fixed on MOVER and were not adjusted on the basis of local results.

### 2.10. Deviations and post hoc analyses

1. Three-cutpoint calibration was added after multinomial logistic regression calibration was seen to lower κ in MOVER; in MOVER it is exploratory, and in the local validation set it is a prespecified confirmation.
2. The OpenJev arm, OpenJev+LoRA, scale sensitivity and the VitalDB replication were all added after the main results of the in-house readout were known, so the expected direction cannot be regarded as an independent prior.
3. An early internal file called the in-house readout “generative zero-shot”; this was corrected and a true generative zero-shot arm was added.
4. VitalDB T2 inputs and the target-site LoRA control were added after the zero-shot and calibration curves of that database were known.
5. Generative zero-shot was run on T1 only; there is no like-for-like generative comparator for T2.
6. Option-order permutation averaging, the Chinese T1 template, label-pool matching and label-sampling sensitivity, δ sensitivity, break-even, subgroups and the 4-bit readout latency were robustness or exploratory analyses added after internal review; they do not change the prespecified hierarchy of hypotheses.
7. OpenJev tuning was done only on the independent MOVER tuning set; the local validation set took no part in any selection.
8. The natural-language guidance experiment was added after the main results were known to test the expectation that the output can be steered by natural language; the four paragraphs were fixed before validation results were seen, but the “local habit” paragraph is based on the principal investigator’s description of local practice, i.e., a domain expert’s account of local custom, and it told the model that the population includes oncologic and major abdominal surgery.

## 3. Results

### 3.1. Cohort characteristics

Table 1 gives the cohort characteristics of the three sources. The local hospital performs abdominal and pelvic surgery; 68.6% of its patients are women, and the ASA I / II / III / IV–V mix is 1.3% / 46.0% / 50.0% / 2.6%. In MOVER, ASA III accounts for 51.2% and IV–V for 12.2%; in VitalDB, ASA I accounts for 28.7%, II for 59.3% and III for only 11.3%. The three mixes differ markedly.

### 3.2. Non-generative readout versus generative zero-shot (H1)

In 1,000 randomly drawn local validation operations, 999 were parsable (1 excluded); the uncalibrated readout (T1) reached κ = 0.460 against 0.227 for generative zero-shot, a difference of +0.234 (95% CI 0.201 to 0.264). In the first 2,000 operations of the MOVER evaluation subset, 1,978 were parsable (22 excluded), and the values were 0.473 against 0.249, a difference of +0.224 (0.199 to 0.247). The share of unparsable generative outputs was 1.1% (MOVER) and 0.1% (local). H1 holds in both sources. H1 is a paired comparison on the same cases and is internally valid. Comparability of subset and full set is shown in Supplementary Table S25: the readout κ was 0.476 and 0.470 for the first and last 2,000 MOVER operations (evaluation subset 0.473), with a similar mix; the local random subset is slightly lighter (ASA II 49.0%, III 46.8%, against 45.9% and 49.9% outside the subset), with a readout κ of 0.461 against 0.432 outside the subset and 0.443 in the whole validation set, so the absolute κ of the local H1 is about 0.018 higher than in the full set. The prompts of the two arms differ (the generative arm asks for a rationale and does not list options), so the difference reflects the combined effect of the full generative usage versus a closed-set readout.

### 3.3. Speed and training cost

#### Token consumption (same Qwen3.5-9B tokenizer)

The readout prompt averaged 171 input tokens in local T1 (MOVER 161); after one forward pass, the logits of the four option letters at the last position were read, and 0 tokens were generated. The generative zero-shot prompt had 187 input tokens (MOVER 177) and required token-by-token output; on the first 96 operations of the local generative subset, the measured average was 89 output tokens (median 87, 10th–90th percentile 73–104, all ending in “Final answer”). For 10,000 cases, readout and LoRA inference each process about 1.71 M tokens with 0 decoding steps, and generative prompting about 2.76 M tokens, of which 0.89 M are sequentially executed decoding steps; the readout’s token total is about 62% of the generative one. The gap between 7,067 ms and 111 ms comes mainly from these roughly 89 sequential decoding steps, each limited by memory bandwidth, and cannot be derived directly from the token totals. One-time label-related consumption: readout plus 1,000-label calibration needs forward scoring of 1,000 cases, about 0.17 M tokens (100 labels: about 0.017 M); LoRA training at n = 1,000 processes the same 0.17 M tokens but needs backpropagation and parameter updates (the full local set of 4,326 cases is about 0.74 M tokens), and each site must store one adapter. These token counts are estimated from training input length and exclude the extra computation of model loading and gradient checkpointing; MOVER generative output tokens were not re-measured.

Table 2 lists speed and cost. At batch size 1, the median per-case latency was 111 ms for the readout (95th percentile 118 ms) and 7,067 ms for generative zero-shot (9,105 ms), about 64 times (the prompt asks for a rationale, and the roughly 89 tokens of sequential generation account for most of this; earlier evaluations had already reported that non-generative decision models are faster and cheaper than generative LLMs [16, 19, 22], and the 64-fold figure here is what was measured under this prompt). Projected inference time for 10,000 operations is about 18.5 min for the readout (14 min at batch size 8) and about 19.6 h for generative prompting (3.5 h at batch size 8). The 4-bit LoRA had a latency of 179 ms; at the same 4-bit precision, the readout without adapter took 156 ms, so the adapter adds about 23 ms (about 15%); the lower latency of the fp16 readout (111 ms, re-measured 114 ms) differs mainly through the dequantization overhead of 4-bit, not through the method itself (Supplementary Table S21). The fp16 LoRA run across two cards (adapter not merged, including inter-card transfer) had a batch-1 p50 latency of 160 ms (Supplementary Table S22). The extra cost of calibrating the readout with k labels is scoring k cases (about 11 s for k = 100, about 2 min for k = 1,000) plus a cutpoint fit of under 1 s on a CPU; LoRA training on the 9B model, 4-bit, one 2080 Ti, takes about 6.7 min for n = 1,000, about 20 min for n = 3,000 and about 81 min for n = 12,000 (each plus 3–4 min of loading); the 27B model takes about 23 min and 69 min for n = 1,000 and 3,000. A LoRA adapter has to be trained and stored separately at each site. Fig. 3 shows speed against κ and κ against label number.

**Fig. 3.**
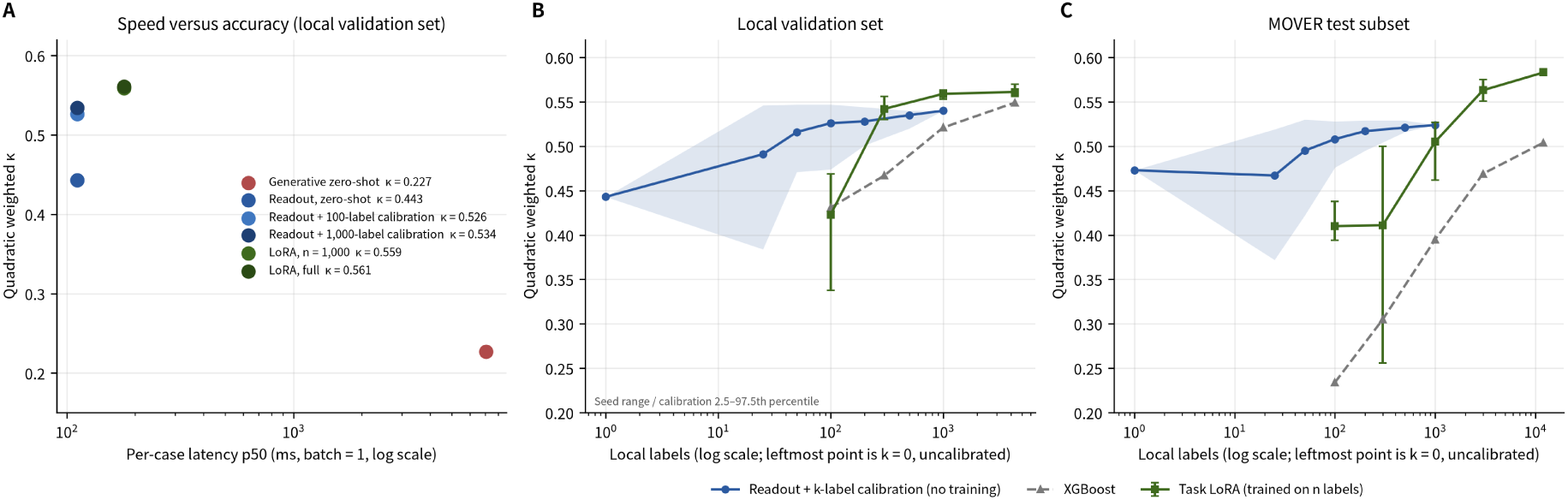
Speed, accuracy and label efficiency. A: median per-case inference latency (log axis, batch size 1) against κ for each approach in the local validation set (generative zero-shot n = 999, others n = 2,753). B, C: κ against the number of local labels (local validation set; MOVER test subset n = 4,000). Blue line and shading: mean and 2.5–97.5 percentiles of the readout calibrated with k labels (leftmost point k = 0, uncalibrated); green line: three-seed mean of task LoRA with seed range as error bars; dashed gray line: XGBoost. κ, quadratic-weighted κ; LoRA, low-rank adaptation.

**Fig. 4.**
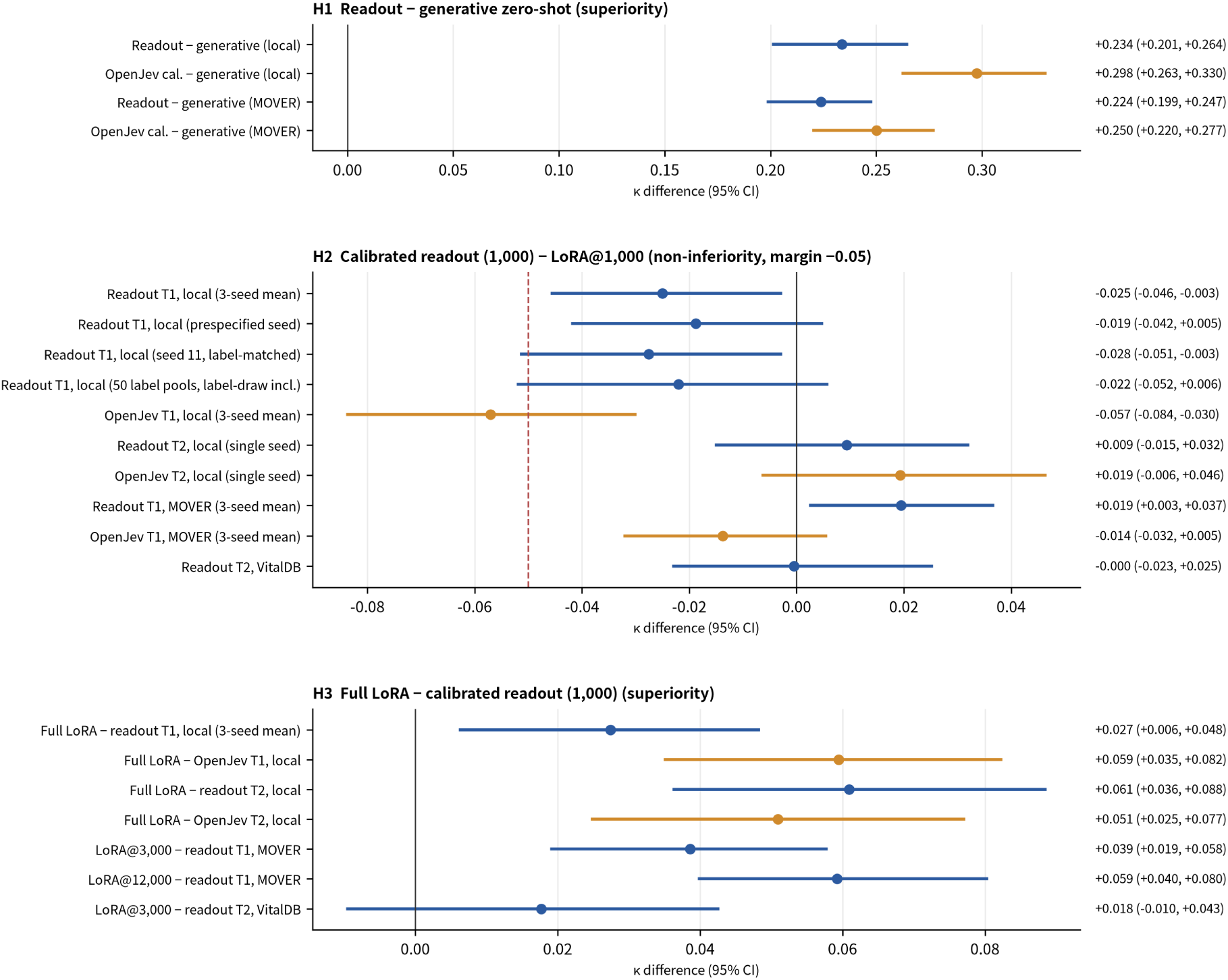
Forest plot of paired differences. Points are κ differences (A − B), lines the 95% CI (patient-level cluster bootstrap, B = 1,000). H1: readout (blue) or OpenJev (orange) minus generative zero-shot; H2: calibrated readout or OpenJev with 1,000 labels minus LoRA@1,000 (“label-matched” means LoRA and calibration use the same 1,000 labels), with the red dashed line at the non-inferiority margin of −0.05; H3: full-set (or 3,000 or 12,000 labels) LoRA minus calibrated readout or OpenJev. T1: structured variables only; T2: plus preoperative diagnosis text. “Three-seed mean”: average of the three LoRA seeds; “single seed”: one seed only.

**Table 2.** Speed and training cost of each approach (9B base model, one modified RTX 2080 Ti with expanded memory (22 GB), 4-bit training)

| Approach | Tokens per case<br>(input + sequential<br>output, local T1) | Per-case<br>latency p50<br>(batch 1) | 10,000 cases<br>(batch 1 / batch<br>8) | Training cost (GPU time) | Local labels<br>needed for $\kappa \geq$<br>0.52 | Inference<br>memory |
| --- | --- | --- | --- | --- | --- | --- |
| Generative<br>zero-shot (fp16) | 187 + 89 | 7,067 ms | 19.6 h / 3.5 h | None | Not reached ($\kappa$<br>0.227) | 18.9 GB |
| Non-generative<br>readout,<br>zero-shot (fp16) | 171 + 0 | 111 ms | 18.5 min / 14<br>min | None | Not reached ($\kappa$<br>0.443) | 18.8 GB |
| Non-generative<br>readout,<br>zero-shot (4-bit,<br>no adapter) | 171 + 0 | 156 ms | 26.0 min / 13.9<br>min | None | Not reached ($\kappa$<br>0.346; 0.514 after<br>calibration) | 8.6 GB |
| Readout +<br>k-label<br>calibration<br>(fp16) | 171 + 0<br>(calibration at $k =$<br>1,000 needs a<br>further 0.17 M<br>tokens of forward<br>passes) | 111 ms | Same | Scoring $k$ cases: about 11 s<br>at $k = 100$, about 2 min at $k$<br>$= 1,000$; three-cutpoint fit $<$<br>1 s (CPU) | About 100 ($\kappa$<br>0.526) | 18.8 GB |
| Task LoRA<br>(fp16, layers<br>split across two<br>cards, $n = 1,000$;<br>supplementary) | 171 + 0 (training<br>processes 0.17 M<br>tokens, including<br>backward pass) | 160 ms<br>(adapter not<br>merged,<br>includes<br>inter-card<br>transfer) | — | 324 s (62 steps) | Measured at $n =$<br>1,000 only ($\kappa$<br>0.553) | 10.4 + 9.5<br>GB (two<br>cards) |
| Task LoRA ($n$<br>labels; 4-bit +<br>adapter) | 171 + 0 (training at<br>$n = 1,000$<br>processes 0.17 M<br>tokens, including<br>backward pass) | 179 ms | 29.8 min / 17.2<br>min | $n = 300$ about 2 min; 1,000<br>about 6.7 min; 3,000 about<br>20 min; 12,000 about 81<br>min (each plus 3–4 min<br>loading); 27B: $n = 1,000$<br>about 23 min, 3,000 about<br>69 min | About 300 ($\kappa$<br>0.542) | 8.7 GB +<br>one adapter<br>per site |
| OpenJev (open<br>System One) | — | Falls back to<br>full-sequence<br>scoring per<br>candidate on<br>this machine,<br>about 0.8–1.3<br>s per case, not<br>representative<br>of design<br>speed | — | None | 0.502 after<br>calibration, not<br>reached | — |

Note: usable accuracy is defined as local validation $\kappa \geq 0.52$ (about 97% of the calibrated-readout plateau of 0.534; it only separates whether the plateau region is reached and is not a clinical threshold); the readout ran in fp16 and LoRA in 4-bit with adapter, so precision and quantization differ and latency cannot be attributed to the method itself; the 4-bit readout row separates the effects of quantization and adapter.

### 3.4. Label efficiency: calibrated readout versus LoRA (H2, H3)

#### Local validation set (T1) (Fig. 3, Table 3)

The uncalibrated readout had κ = 0.443 (95% CI 0.419–0.468); with three cutpoints fitted on local labels, κ was 0.526 with 100 labels and 0.534 (0.508–0.559) with 1,000. The local LoRA (mean of three seeds) had κ = 0.423, 0.542, 0.559 and 0.561 at n = 100, 300, 1,000 and 4,326. With 100 labels the calibrated readout (0.526) was above LoRA (0.423); LoRA needed about 300 labels to match it. XGBoost (trained on the local development set) gave 0.431, 0.467, 0.521 and 0.549 for the same label numbers.

**Table 3.**
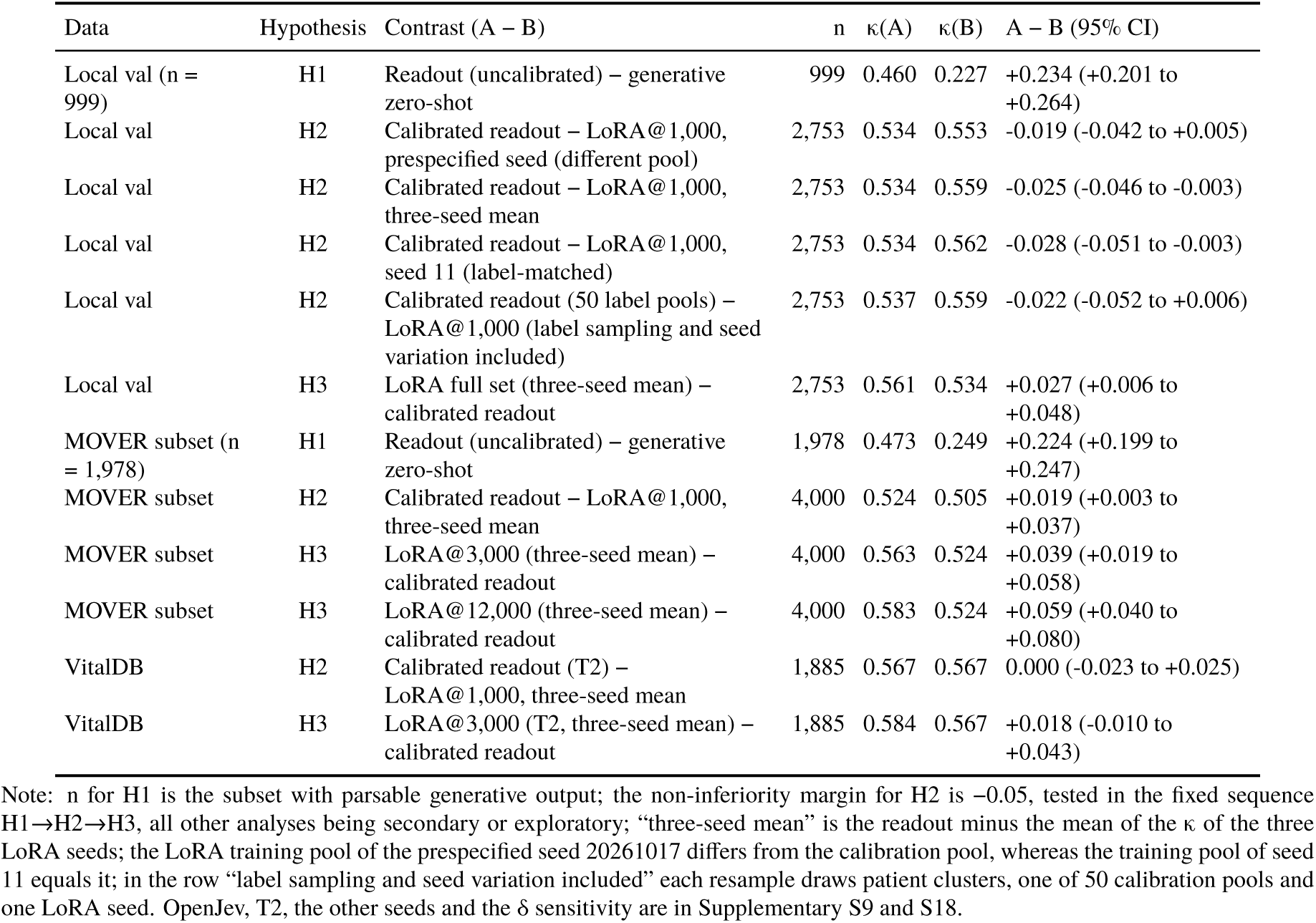
Paired differences in the main analyses (quadratic-weighted κ, A − B; 95% CI by patient-level cluster bootstrap, B = 1,000)

| Data | Hypothesis | Contrast (A – B) | n | $\kappa$(A) | $\kappa$(B) | A – B (95% CI) |
| --- | --- | --- | --- | --- | --- | --- |
| Local val (n = 999) | H1 | Readout (uncalibrated) – generative zero-shot | 999 | 0.460 | 0.227 | +0.234 (+0.201 to +0.264) |
| Local val | H2 | Calibrated readout – LoRA@1,000, prespecified seed (different pool) | 2,753 | 0.534 | 0.553 | -0.019 (-0.042 to +0.005) |
| Local val | H2 | Calibrated readout – LoRA@1,000, three-seed mean | 2,753 | 0.534 | 0.559 | -0.025 (-0.046 to -0.003) |
| Local val | H2 | Calibrated readout – LoRA@1,000, seed 11 (label-matched) | 2,753 | 0.534 | 0.562 | -0.028 (-0.051 to -0.003) |
| Local val | H2 | Calibrated readout (50 label pools) – LoRA@1,000 (label sampling and seed variation included) | 2,753 | 0.537 | 0.559 | -0.022 (-0.052 to +0.006) |
| Local val | H3 | LoRA full set (three-seed mean) – calibrated readout | 2,753 | 0.561 | 0.534 | +0.027 (+0.006 to +0.048) |
| MOVER subset (n = 1,978) | H1 | Readout (uncalibrated) – generative zero-shot | 1,978 | 0.473 | 0.249 | +0.224 (+0.199 to +0.247) |
| MOVER subset | H2 | Calibrated readout – LoRA@1,000, three-seed mean | 4,000 | 0.524 | 0.505 | +0.019 (+0.003 to +0.037) |
| MOVER subset | H3 | LoRA@3,000 (three-seed mean) – calibrated readout | 4,000 | 0.563 | 0.524 | +0.039 (+0.019 to +0.058) |
| MOVER subset | H3 | LoRA@12,000 (three-seed mean) – calibrated readout | 4,000 | 0.583 | 0.524 | +0.059 (+0.040 to +0.080) |
| VitalDB | H2 | Calibrated readout (T2) – LoRA@1,000, three-seed mean | 1,885 | 0.567 | 0.567 | 0.000 (-0.023 to +0.025) |
| VitalDB | H3 | LoRA@3,000 (T2, three-seed mean) – calibrated readout | 1,885 | 0.584 | 0.567 | +0.018 (-0.010 to +0.043) |

Note: n for H1 is the subset with parsable generative output; the non-inferiority margin for H2 is -0.05, tested in the fixed sequence H1→H2→H3, all other analyses being secondary or exploratory; “three-seed mean” is the readout minus the mean of the $\kappa$ of the three LoRA seeds; the LoRA training pool of the prespecified seed 20261017 differs from the calibration pool, whereas the training pool of seed 11 equals it; in the row “label sampling and seed variation included” each resample draws patient clusters, one of 50 calibration pools and one LoRA seed. OpenJev, T2, the other seeds and the $\delta$ sensitivity are in Supplementary S9 and S18.

#### H2

calibrated readout (1,000 labels) minus LoRA@1,000 was −0.019 (95% CI −0.042 to +0.005) for the prespecified seed 20261017 and −0.025 (−0.046 to −0.003) for the mean of the three seeds. The LoRA training pool of the prespecified seed differs from the calibration pool, so the comparison is not label-matched. The label-matched contrast (seed 11, LoRA and calibration using the same 1,000 labels) was −0.028 (−0.051 to −0.003); seed 12, whose training pool partly overlaps the calibration pool, gave −0.029 (−0.051 to −0.006). Both lower bounds fall slightly below −0.05. To include label sampling in the uncertainty, 50 independent 1,000-label pools were drawn from the development set and cutpoints fitted in each: the mean κ of the calibrated readout was 0.537 (between-pool SD 0.006, range 0.514–0.546) and that of LoRA for the three seeds 0.553–0.563. Each resample drew patient clusters, one calibration pool and one LoRA seed together, and the H2 difference was −0.022 (−0.052 to +0.006), with a lower bound slightly below −0.05. At δ = 0.05, H2 therefore holds for the prespecified seed and the seed mean and sits at the margin for the label-matched and label-sampling analyses; at δ = 0.03 none holds, and at δ = 0.10 all hold (Supplementary Table S18). To test whether 4-bit quantization affects this contrast, an LoRA with fp16 base model was run once across two cards (local n = 1,000, seed 11, same labels as the 4-bit LoRA): κ = 0.553 (0.525–0.580), accuracy 0.741, macro AUC 0.845; against the 4-bit LoRA (0.562, 0.747, 0.846) the difference was −0.008 (−0.021 to +0.006). With this fp16 LoRA as the comparator, the label-matched H2 difference was −0.019 (−0.045 to +0.006), with a lower bound above −0.05 (one seed and one label pool, exploratory; Supplementary Table S22). H2 therefore holds in the prespecified analysis, the three-seed mean and the label-matched fp16 LoRA analysis, and sits at the margin in the label-matched 4-bit LoRA and label-sampling analyses (lower bounds −0.051 and −0.052). If the readout is also quantized to 4-bit, the local difference is −0.045 (−0.069 to −0.022), which does not meet the margin, whereas MOVER gives +0.016 (−0.002 to +0.034), which does (Section 3.9, Supplementary Table S23); H2 thus depends on running the readout in fp16, which is feasible on one card with 22 GB.

#### H3

full-set LoRA minus calibrated readout was +0.027 (three-seed mean, +0.006 to +0.048); per seed +0.024, +0.036 and +0.022, with a lower bound of −0.001 for seed 12. H3 holds, but the effect is small.

#### MOVER subset

the uncalibrated readout had κ = 0.473 and 0.524 after calibration with 1,000 labels; LoRA had 0.410, 0.411, 0.505, 0.563 and 0.583 at n = 100, 300, 1,000, 3,000 and 12,000. Calibrated readout minus LoRA@1,000 was +0.019 for the seed mean (+0.003 to +0.037), and −0.003, +0.062 and −0.001 per seed; LoRA@3,000 and LoRA@12,000 exceeded the calibrated readout by 0.039 (0.019 to 0.058) and 0.059 (0.040 to 0.080). The between-seed range of LoRA at n = 300 was 0.256–0.500, showing marked variability in small samples. In MOVER, 50 independent 1,000-label pools drawn from a further 5,000 training-split operations gave a mean calibrated-readout κ of 0.525 (between-pool SD 0.004, range 0.513–0.530), and the H2 difference including label sampling and seed variation was +0.020 (−0.020 to +0.080).

#### Exact agreement and adjacent-class errors (Table 4)

The gap in κ is smaller than the gap in case-level correctness. In the local validation set, the accuracy of the zero-shot readout, the readout calibrated on 1,000 labels and LoRA (n = 1,000, mean of three seeds) was 0.563, 0.666 and 0.746; adjacent accuracy (|difference| ≤ 1) was 0.984, 0.991 and 0.999; and errors of two or more classes were 1.6%, 0.9% and 0.1%. In the MOVER subset, accuracy of the calibrated readout and of LoRA (n = 1,000) was 0.515 and 0.600, and errors of two or more classes 3.4% and 1.3%. Cutpoints are fitted to maximize κ, so non-inferiority in κ does not imply non-inferiority in accuracy.

**Table 4.**
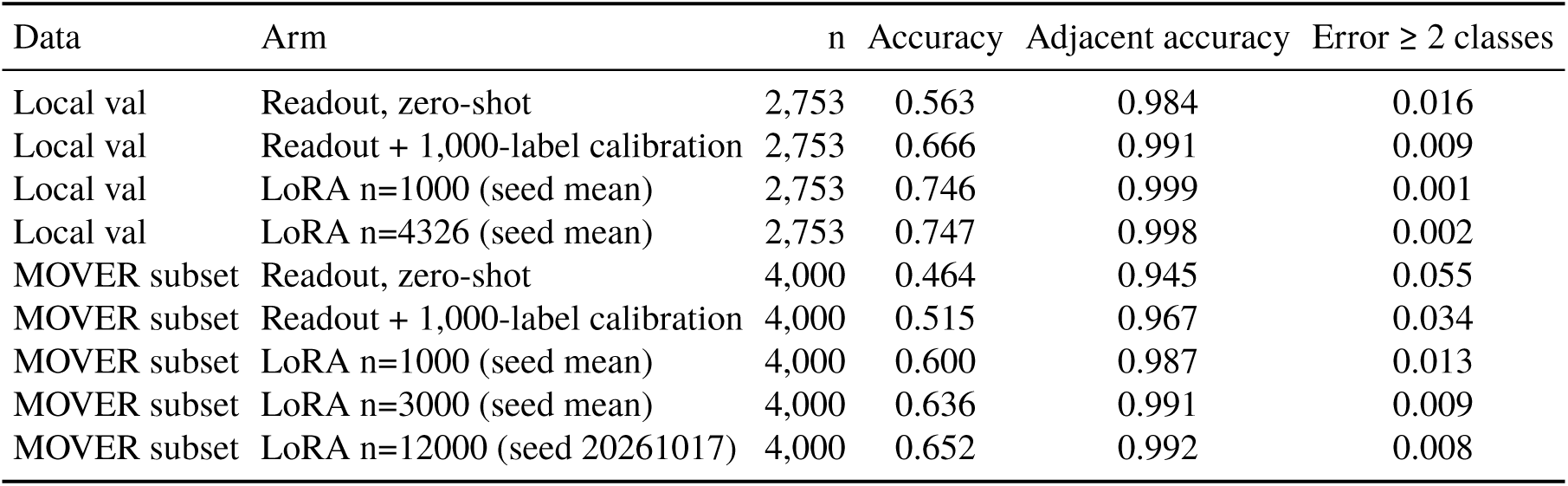
Accuracy and adjacent-class errors (cutpoints fitted to maximize κ; LoRA accuracy is the three-seed mean, MOVER n = 12,000 is the single seed 20261017)

#### Crossover and decision rule

The κ curves of the calibrated readout and of LoRA cross at about 270 labels locally (per seed 259, 264, 318), at about 1,000–1,800 in MOVER (per seed 958, 986, 1,832; the calibration curve is extrapolated as a plateau beyond k = 1,000) and at about 270–1,800 in VitalDB (T2; per seed 268, 996, 1,775). In terms of accuracy, LoRA crosses earlier: locally at n = 100 the accuracy difference between LoRA and the calibrated readout (pool 11) was +0.011 (−0.006 to +0.029), i.e., no difference (against a mean of 0.693 over 50 pools, LoRA’s 0.677 is even lower); at n = 300 it was +0.071 (+0.056 to +0.086) and at n = 1,000 +0.080 (+0.065 to +0.095). In MOVER at n = 100, 300 and 1,000 it was +0.062 (+0.045 to +0.083), +0.047 (+0.033 to +0.064) and +0.085 (+0.068 to +0.102), so LoRA’s accuracy is already higher at n = 100 (Supplementary Tables S19, S25). Thus, for κ as the target, below about 300 labels (local) to 1,000 (MOVER, VitalDB) the calibrated readout is no worse than LoRA; for case-level accuracy as the target, LoRA is better from about 300 labels locally and from 100 labels in MOVER.

The calibration curve flattens at about 100 labels: in MOVER 0.467, 0.495, 0.508, 0.517, 0.521 and 0.524 (k = 25, 50, 100, 200, 500, 1,000; zero-shot 0.473); locally 0.491, 0.516, 0.526, 0.528, 0.535 and 0.540 (zero-shot 0.443). These curves are subsets of the same 1,000-label pool, so the points are not independent.

### 3.5. Dependence on implementation and input information

The frozen OpenJev reached κ = 0.314 uncalibrated and 0.502 calibrated (macro AUC 0.761) on local T1, below the in-house readout (0.443; 0.534). H2 does not hold for OpenJev: calibrated OpenJev minus LoRA@1,000 was −0.051 (−0.080 to −0.022; three-seed mean −0.057, −0.084 to −0.030). In MOVER, calibrated OpenJev had κ = 0.491, differing from the in-house readout by +0.018 (−0.001 to +0.037) and from LoRA@1,000 by a seed-mean −0.014 (−0.032 to +0.005). With preoperative diagnosis text (T2), the local uncalibrated readout rose from 0.443 to 0.505, OpenJev reached 0.522 uncalibrated and 0.555 calibrated (macro AUC 0.834), and local T2 LoRA reached 0.536 at n = 1,000 and 0.606 on the full set. Under T2, calibrated OpenJev minus LoRA@1,000 was +0.019 (−0.006 to +0.046), so H2 holds (the T2 diagnosis text is in Chinese, and part of this difference may come from language matching; see below); full-set LoRA minus calibrated OpenJev was +0.051 (+0.025 to +0.077). The calibrated κ of the in-house readout under T2 was 0.545, +0.009 (−0.015 to +0.032) above LoRA T2@1,000. T2 has no like-for-like generative comparator (Section 2.10). To separate language from information content, the T1 template was translated into Chinese (the local procedure names were originally Chinese). With an English prompt shell, the Chinese T1 template had an uncalibrated and calibrated κ of 0.462 and 0.516 (macro AUC 0.788), close to the English template (0.443, 0.534, 0.794); when the prompt shell was also in Chinese, the uncalibrated κ rose to 0.512, the calibrated κ was 0.530 and the macro AUC rose to 0.821, already close to T2 with diagnosis text (0.505, 0.545, 0.833, English prompt shell). Most of the gain in macro AUC from T1 to T2 (0.794→0.833) can therefore be explained by matching of language and prompt shell (0.794→0.821), and the increment from the diagnosis text itself is small (0.821→0.833); the differences of OpenJev and LoRA between T1 and T2 likewise should not be attributed to information content alone. LoRA and OpenJev were not rerun with the Chinese T1 template (Supplementary Table S20).

A decision-model implementation with LoRA brought no extra gain. OpenJev+LoRA minus task LoRA averaged −0.011 over six (n, seed) combinations in MOVER; locally at n = 1,000 the three seeds gave −0.004, −0.005 and −0.004, and on the full set +0.001, −0.016 and +0.014.

### 3.6. Second site: VitalDB (target-site replication, exploratory)

The VitalDB test set has 1,885 operations (1,788 patients) with an ASA I / II / III / IV–V mix of 28.3 / 58.8 / 11.7 / 1.2%, which differs markedly from MOVER and the local hospital; this part is an exploratory replication only. Calibration labels and LoRA training labels both came from the site’s own training split.

With diagnosis and examination inputs (T2), zero-shot readout κ was 0.380 and frozen OpenJev 0.370; calibration with 100 target-site labels (60-run average) gave 0.545 and 0.549, and with 1,000 labels 0.567 and 0.582 (Fig. 5). Target-site LoRA (T2, three-seed mean and range) was 0.282 (0.061–0.444), 0.470 (0.365–0.570), 0.567 (0.532–0.602) and 0.584 (0.561–0.599) at n = 100, 300, 1,000 and 3,000. Calibrated readout (1,000 labels) minus LoRA@1,000 was 0.000 for the seed mean (−0.023 to +0.025), and +0.035 (+0.004 to +0.070), −0.001 (−0.027 to +0.028) and −0.035 (−0.063 to −0.008) per seed; for OpenJev the same contrast was +0.015 (−0.008 to +0.037). LoRA@3,000 minus calibrated readout was +0.018 (−0.010 to +0.043), an interval that includes 0. The difference between the calibrated readout and LoRA@1,000 varied from −0.035 to +0.035 across seeds because the between-seed κ range of LoRA at this site is wide (0.532–0.602); the +0.035 of the prespecified seed should not be quoted on its own.

**Fig. 5.**
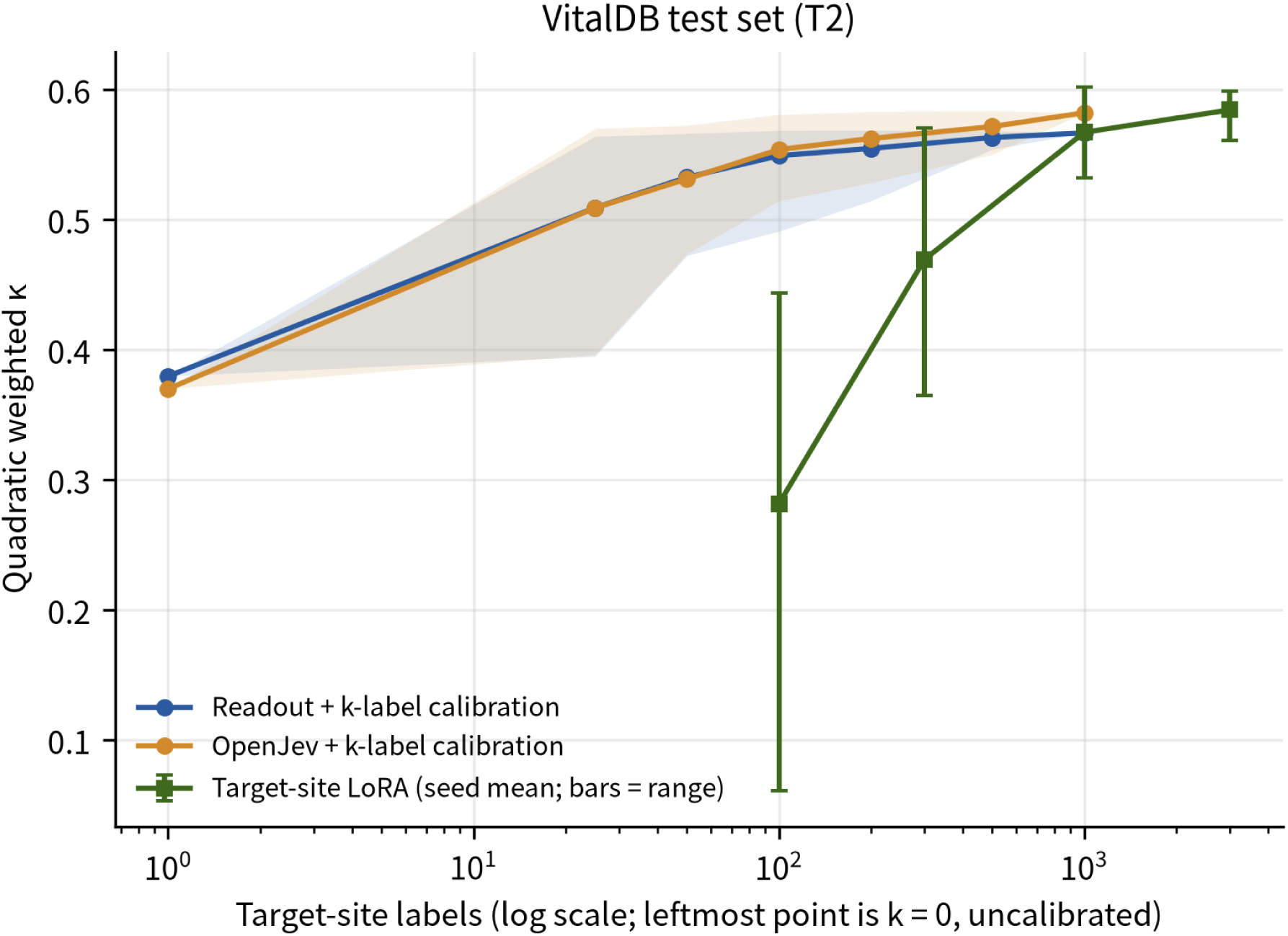
Label efficiency at the target site in VitalDB (T2, test set n = 1,885). Blue and orange lines: κ of the readout and of the frozen OpenJev after calibration with k target-site labels (k drawn from a 1,000-label pool in the training split, 100 repetitions per k, shading the 2.5–97.5 percentiles, leftmost point k = 0, uncalibrated); green line: three-seed mean of the target-site LoRA with seed range as error bars.

With structured variables only (T1), the zero-shot readout had κ = 0.357 and OpenJev 0.366; after calibration on 1,000 labels, 0.507 and 0.505. The target-site LoRA (one seed) reached 0.500 and 0.548 at n = 1,000 and 3,000; calibrated readout minus LoRA@1,000 was +0.007 (−0.024 to +0.039), and LoRA@3,000 minus calibrated readout was +0.041 (+0.010 to +0.072).

### 3.7. Natural-language guidance

Guidance moved the predicted class mix of the readout in the direction of its wording (Fig. 6A, B). With structured variables only (T1), all guidance lowered κ: 0.443 without guidance, 0.312 with the official ASA definitions (difference −0.131, 95% CI −0.150 to −0.113), 0.272 with the local habit (−0.171, −0.195 to −0.146) and 0.187 with strict adherence to the guideline (−0.256, −0.275 to −0.237). The “local habit” guidance labeled 85.2% of cases as class II, reduced ASA I to 0 and left only 14.6% in class III (48.8% in the original records); the mix shifted without approaching the original records.

**Fig. 6.**
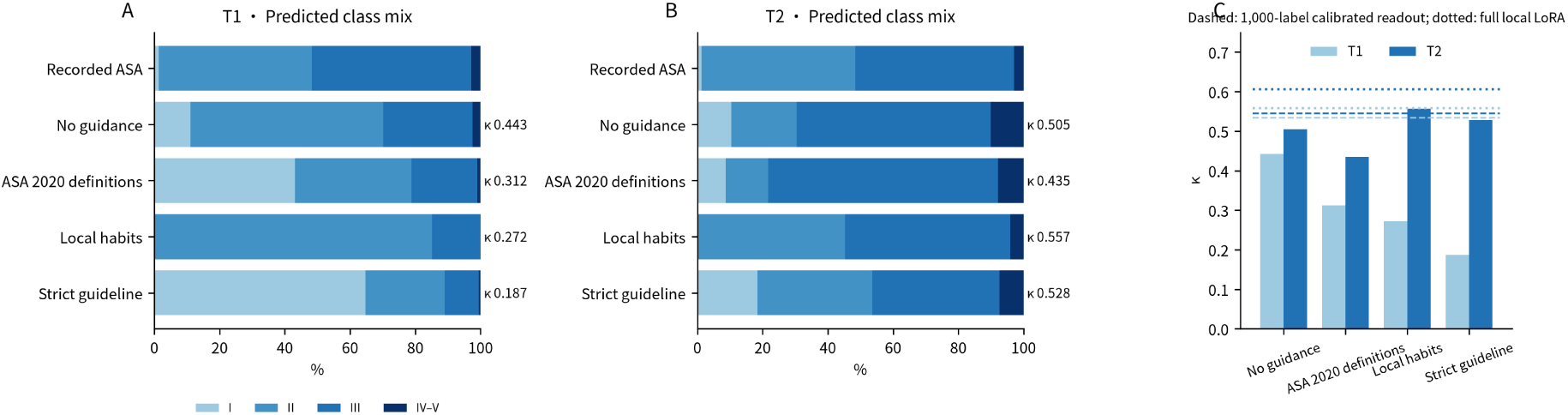
Effect of natural-language guidance on the predicted class mix and κ of the readout (local validation set, n = 2,753). A, B: predicted class mix (I, II, III, IV–V) and corresponding κ for the original records and four guidance conditions, with T1 and T2 input; C: κ of each condition, the dashed line being the readout calibrated on 1,000 labels and the dotted lines the full-set local LoRA (light: T1, dark: T2). The wording of the guidance was fixed before validation results were seen.

With preoperative diagnosis text (T2) the result differed. The “local habit” guidance brought the predicted mix (I / II / III / IV–V = 0.0 / 45.3 / 50.6 / 4.1%) close to the original records (1.3 / 47.0 / 48.8 / 2.9%) and raised κ from 0.505 to 0.557 (+0.052, +0.028 to +0.077). This level does not differ from the readout calibrated on 1,000 labels (0.545; +0.012, −0.011 to +0.036) or from local LoRA T2@1,000 (0.536; +0.021, −0.005 to +0.047), and is below full-set local LoRA (0.606; −0.049, −0.075 to −0.022). Under T2, “strict adherence to the guideline” gave 0.528 (+0.023, +0.002 to +0.044) and the official ASA definitions 0.435 (−0.070, −0.087 to −0.052).

When 100 labels were used for calibration after guidance, κ under T2 for the four conditions was 0.531–0.550 (no guidance 0.531, official definitions 0.531, local habit 0.532, strict guideline 0.550), a small spread. The gain from guidance overlaps with the threshold adjustment that a few labels provide, and the two do not add up.

### 3.8. Probability calibration and contradictory content in generative output

For raw zero-shot probabilities, the ECE in MOVER was 0.092 (95% CI 0.077–0.110) for the in-house readout and 0.151 (0.135–0.169) for OpenJev; locally 0.030 (0.017–0.047) and 0.140 (0.122–0.161). After single-parameter temperature scaling, MOVER gave 0.047 (0.038–0.066) and 0.049 (0.034–0.068), and the local hospital 0.072 (0.055–0.090) and 0.075 (0.060–0.096); the local ECE of the in-house readout went from 0.030 to 0.072, i.e., it did not improve. The ECE of task LoRA (n = 1,000) was 0.051 (0.028–0.074) in MOVER and 0.040 (0.027–0.064) locally, and 0.027 (0.012–0.050) and 0.023 (0.013–0.048) after scaling. Intervals for the Brier score are in Supplementary S4. The calibration of readout probabilities points in different directions at the two sites: locally the raw ECE of the readout (0.030) is below that of LoRA (0.040), whereas in MOVER it is the reverse (0.092 versus 0.051); in some settings the readout needs a temperature parameter.

The rule-based screen of generative rationales flagged 8.0%–8.6% of entries; of the 40 flagged samples read by hand, 25% (10/40) truly contradicted the structured input, all being cases where the input recorded “no hypertension” and the rationale said “controlled hypertension” or “history of hypertension”; four more (10%) described a BMI of 27.9–29.2 as obese. From this, the proportion of rationales with such contradictions is estimated at about 2% (95% CI about 1%–3%). The estimate covers only structured fields and should be read as a lower bound. The non-generative readout outputs no narrative text, its output is limited to four options, and its unparsable rate is 0; this property is guaranteed by construction and is not an empirical estimate.

### 3.9. Robustness and sensitivity

#### Base-model scale

the κ of the in-house readout was 0.489 for Qwen3.5-27B and 0.473 for Qwen3.5-9B, a difference of +0.015 (−0.006 to +0.038); for the frozen OpenJev (4-bit, first 2,000 MOVER operations) it was 0.387 for 27B and 0.444 for 9B, a difference of −0.057 (−0.080 to −0.033). The zero-shot readout of Qwen3.8-27B reached κ = 0.115 only (strongly biased toward ASA I), rising to 0.590 after LoRA@3,000. Under LoRA, 27B was 0.02–0.03 higher than 9B. A larger base model did not reliably improve zero-shot accuracy.

#### Option order and label format

the uncalibrated κ of the in-house readout varies widely with option order. In the local validation set, the natural order, the reversed order, the three cyclic shifts and one random order gave 0.443, 0.258, 0.331, 0.142, 0.493 and 0.505; in the MOVER evaluation subset 0.473, 0.245, 0.315, 0.222, 0.449 and 0.435. The natural order is the default of the readout script, was fixed before any result was seen, is not the highest locally and is the highest in MOVER. Cutpoint calibration absorbed most of the variation: after fitting cutpoints for each order on the same 1,000-label pool, κ was 0.518–0.534 locally and 0.481–0.524 in MOVER. After averaging the probabilities of the six orders, the uncalibrated κ was 0.396 (local) and 0.414 (MOVER), the calibrated κ 0.531 and 0.523, the macro AUC 0.785 and 0.745 (natural order 0.794 and 0.762), and the ECE 0.024 and 0.091 (natural order 0.030 and 0.092). The difference between the order-averaged calibrated readout and the three-seed mean of LoRA@1,000 was −0.028 (−0.051 to −0.006) locally and +0.018 (−0.000 to +0.035) in MOVER, in line with −0.025 and +0.019 for the natural order, so the H2 conclusion does not depend on the order chosen (Supplementary Table S20). For OpenJev, κ under numeric, letter and Roman-numeral label formats was 0.355–0.384 (MOVER) and 0.259–0.276 (local, 1,000 random operations), and 10%–16% of predictions flipped between formats.

#### 4-bit quantized readout

4-bit (NF4) quantization degraded the zero-shot readout. In the local validation set, uncalibrated κ fell from 0.443 to 0.346 (difference −0.097, −0.114 to −0.081), macro AUC from 0.794 to 0.739, with an agreement of 0.588 with the fp16 predictions; after calibration on 1,000 labels, κ fell from 0.534 to 0.514 (−0.020, −0.033 to −0.005). In the MOVER evaluation subset, uncalibrated κ fell from 0.473 to 0.378 (−0.095, −0.112 to −0.080), macro AUC from 0.762 to 0.733, agreement 0.622; after calibration the values were 0.521 and 0.524, a difference of −0.003 (−0.015 to +0.009). Calibration thus recovered most of the loss of the 4-bit readout, leaving a gap of 0.020 locally. At the same 4-bit precision, calibrated readout minus LoRA@1,000 was −0.045 locally (three-seed mean; −0.069 to −0.022) and −0.047 (seed 11, label-matched; −0.073 to −0.023), and +0.016 in MOVER (−0.002 to +0.034) (Supplementary Table S23).

#### Official definitions in the prompt

on 1,500 MOVER training operations, writing the official ASA definitions into the readout prompt lowered κ from 0.454 to 0.415 and 0.325; the calibrated κ of the eight OpenJev prompt configurations ranged from 0.448 to 0.501, a small spread.

### 3.10. Subgroups (exploratory)

Descriptive results by sex and age in the local validation set are in Supplementary Table S17. The κ of the calibrated readout and of LoRA@1,000 (three-seed mean) was 0.526 and 0.542 in women (n = 1,966), 0.413 and 0.410 in men (n = 787), and 0.484 and 0.480 in patients under 65 years (n = 2,129). In the group aged 65 years or older (n = 624) the ASA mix is concentrated (78.2% class III, no class I), κ is limited by the range of values and is not interpreted; only accuracy is reported (calibrated readout 0.625, LoRA 0.786). The share of ASA III is higher among men and patients aged 65 or older. Sample sizes are limited, and no inference is drawn between subgroups.

## 4. Discussion

### 4.1. Principal findings

In both MOVER and the local hospital, the non-generative readout of one base model was clearly better than generative zero-shot prompting (κ difference about 0.22–0.23; H1 holds). The readout generates no token per case, whereas generative prompting with a rationale decodes about 89 tokens on average, and per-case latency of the former is about 1/64 of the latter; the factor depends on the generative prompt asking for a rationale, and the conclusion that non-generative decision models are faster than generative LLMs is already supported by earlier evaluations [16, 19, 22]. Measured by quadratic-weighted κ, fitting three cutpoints on about 100 local labels brings the readout close to its plateau (local 0.526), at which point LoRA reaches only 0.423; the two curves cross at about 270 labels locally and at about 1,000–1,800 in MOVER. With several thousand labels, LoRA has the higher κ (local full set +0.027, MOVER with 12,000 labels +0.059; H3 holds).

The prespecified H2 holds: the difference between the calibrated readout and LoRA@1,000 was −0.019 and −0.025 for the prespecified seed and the three-seed mean, with lower bounds above −0.05, and −0.019 (lower bound −0.045) with an fp16 LoRA as the label-matched comparator. In stricter sensitivity analyses, the 4-bit LoRA with the same labels gave −0.028 (lower bound −0.051), and including label sampling and seed variation −0.022 (lower bound −0.052), slightly below the margin. The prespecified seed and the calibration pool are not the same, and after label matching the lower bound is −0.051. The point estimate always favors LoRA, by about 0.02–0.03, and all analyses hold at δ = 0.10. With matched precision, readout and LoRA both in fp16 (label-matched) meet the margin, whereas both in 4-bit fail locally (−0.045, lower bound −0.069) and pass in MOVER (+0.016): running the readout in fp16 is one of the conditions for H2. With diagnosis text (T2), both readout implementations meet non-inferiority; in the second site, VitalDB, the seed-mean difference is 0.000 (−0.023 to +0.025), but the between-seed κ range of LoRA at that site is wide. The direction is not the same across the three sites: the local hospital favors LoRA, MOVER favors the readout, and VitalDB shows no difference. Non-inferiority refers to quadratic-weighted κ; in exact agreement LoRA is higher (local n = 1,000: 0.746 versus 0.666), see Section 4.3.

### 4.2. Why a few cutpoints approach LoRA: thresholds and discrimination

The gap of the zero-shot readout has two parts. The first is a shift in thresholds and priors. ASA-PS carries institutional habit, and calibration on 100 local development labels raised κ from 0.443 to 0.526, so a sizeable part of the gap can be closed by three numbers. The second is discrimination: cutpoint calibration does not change the ranking, so the macro AUC stays at 0.794 whereas LoRA reaches 0.846–0.857; this part takes more labels to learn, which is why LoRA’s advantage appears only after several hundred to several thousand labels. Earlier evaluations that read LLM judgments from token probabilities also found that logit-based probability estimates are worse than conventional classifiers trained on structured data and recommended better calibration [10]; here the calibration is done with three ordinal cutpoints, and we distinguish what calibration can and cannot repair.

How much of LoRA’s gain comes from ASA discrimination and how much from fitting a site’s recording habit cannot be separated in this study. The reference is the ASA class recorded at each site, which contains rating habit and label noise; in a published sample of 101 orthopedic patients the inter-clinician quadratic-weighted κ was 0.713 [5], showing that the rating itself carries non-negligible disagreement. We had no independent human rating and do not try to estimate a human ceiling: reporting agreement with the record and with independent raters side by side would need two or more raters on the same data, which is beyond this design. What can be said is that LoRA is unstable with very few labels (between-seed κ range at MOVER n = 300: 0.256–0.500), consistent with small-sample fitting under noisy labels, and that three-cutpoint calibration has few parameters and is less affected by noise.

Compared with published inter-physician agreement, the κ here falls in the lower-middle range. Reported κ between physicians, or between physicians and a reference, varies widely: under favorable conditions or after training, weighted κ between anesthesiologists is 0.62–0.85 (inter-clinician 0.713 [5]; preoperative screening versus day of surgery 0.62, 0.85 after an intervention [31]; two independent anesthesiologists 0.76 [32]; against a reference 0.63 [33]), Fleiss κ is 0.55 [34], and for cross-specialty rating or against a reference panel 0.21–0.43 [33, 35]; the pooled quadratic-weighted κ of AI against clinicians is 0.69 (13 studies, 402,336 cases) [36]. On the external data here, the calibrated readout in MOVER has κ = 0.524 and LoRA 0.505–0.583; in VitalDB (T2) the calibrated readout has 0.567 and LoRA 0.567–0.584; locally the values are 0.534 and 0.559–0.562. These values are above the κ of cross-specialty rating and below the weighted κ between trained anesthesiologists (0.62–0.85) and the pooled AI value of 0.69, which is the lower-middle part of published inter-physician agreement. This comparison has clear limits: the studies differ in population, rating method (real or fictitious cases), weighting of κ and ASA mix, and κ itself varies with prevalence and range; the κ here is the agreement of a model with a single recording rater, with no inter-physician agreement on the same data to compare, so we draw no inference of “human-level performance” (Supplementary Table S24).

The way the input is presented is a larger lever than the method. With diagnosis text, the zero-shot readout rose locally from 0.443 to 0.505 (+0.062), full-set LoRA from 0.561 to 0.606 (+0.048), and calibrated frozen OpenJev from 0.502 to 0.555. But once the T1 template was translated into Chinese with a Chinese prompt shell, the uncalibrated readout κ was already 0.512 and the macro AUC rose from 0.794 to 0.821 (0.833 for T2), indicating that most of this gain comes from matching of language and prompt shell and that the increment from the diagnosis text itself is small; template format and length still differ between T1 and T2, and LoRA and OpenJev were not rerun under the Chinese T1, so this decomposition is preliminary. A larger base model did not reliably help zero-shot performance (in-house readout 27B versus 9B +0.015, OpenJev 4-bit 27B versus 9B −0.057). The frozen OpenJev was weaker on T1 with structured variables only (local uncalibrated κ 0.314) and clearly better after diagnosis text was added, which fits the earlier observation that decision models are stronger on text-dominated tasks, weaker on numeric-dominated ones, and may overstate probabilities [22, 23]; a preregistered evaluation also noted that such models may assign zero probability to the correct diagnosis [24]. Here the closed set has only four classes and the probabilities serve only to rank cases and place cutpoints, so the influence of this risk is limited, but it was not tested separately.

Option order is a known weakness of the readout: the uncalibrated κ ranged over 0.142–0.505 (local) and 0.222–0.473 (MOVER) across six orders, so a single zero-shot κ should not be taken as an operating characteristic of the method. Cutpoint calibration compressed the differences to 0.518–0.534 (local) and 0.481–0.524 (MOVER), and the order-averaged calibrated readout is almost identical to the natural order, so calibration also corrected the order-induced bias and the H2 conclusion does not depend on order; the price is that uncalibrated, unaveraged zero-shot use is not robust, and averaging needs several forward passes.

### 4.3. Label efficiency and a decision rule: the cost of LoRA relative to the non-generative readout

The data support the comparison in five respects. The first is label need: at 100 local labels the calibrated readout has κ = 0.526 and LoRA 0.423, and LoRA needs about 300 labels to catch up. For training and storage, calibration only needs forward scoring of k annotated cases (about 11 s for k = 100, about 2 min for k = 1,000) followed by a three-cutpoint fit on a CPU, and what each site stores is three numbers; LoRA training on a single RTX 2080 Ti takes about 6.7 min for n = 1,000, about 81 min for n = 12,000 and about 23 min for the 27B base model at n = 1,000, and every site trains and stores an adapter. In inference, the readout takes 111 ms and the 4-bit LoRA with adapter 179 ms, about 1.6 times; about two thirds of that comes from 4-bit quantization (the 4-bit readout takes 156 ms) and the adapter itself adds about 23 ms (about 15%), so the latency difference is mostly the overhead of quantization and adapter, not a difference inherent to the methods. Quantization does not affect the two approaches in the same way: LoRA is trained in 4-bit, and with the same labels fp16 and 4-bit LoRA differ in κ by −0.008 (−0.021 to +0.006), so LoRA’s κ advantage is not caused by quantization; the zero-shot readout has no adaptation, and 4-bit lowers its uncalibrated κ by about 0.10 and its calibrated κ by 0.00–0.02. A less aggressive quantization (fp16) thus lets the readout approach the level of the 4-bit LoRA without training, whereas at the same 4-bit the gap widens locally. With very few labels, LoRA is also less stable: at n = 100–300 the κ range across seeds is wide (MOVER n = 300: 0.256–0.500; VitalDB n = 100: 0.061–0.444), whereas the 2.5–97.5 percentile range of repeated samples of the calibrated readout is much narrower (local k = 100: 0.474–0.547; VitalDB k = 100: 0.491–0.569). In token consumption, LoRA inference equals the readout, and training at n = 1,000 processes about 0.17 M tokens, the same as scoring 1,000 cases for calibration, but with backpropagation and parameter updates.

This matches observations in the literature: fine-tuning the clinical language model NYUTron took about 6 hours on 8 A100 GPUs per run [13]; LoRA reduces the number of trained parameters but still needs a labeled training set, and the adapter is site-specific; in multi-institution settings, sequential or federated adapter training is needed to avoid data sharing [14, 15]. This is a sixth cost of LoRA in multi-site deployment; the evidence comes from the literature, and in this study LoRA was trained separately for each of three data sources and joint multi-site training was not tested. Non-generative readout with cutpoint calibration reduces the site-specific part to three thresholds and leaves the model frozen, so no model file has to be maintained for each site. This advantage has limits: at 9B, in 4-bit and on one card, the absolute training cost of LoRA is not high, so the cost advantage should be stated as relative.

None of this makes LoRA inferior across the board. Beyond κ, LoRA is better in exact agreement and in large errors: in the local validation set the accuracy of LoRA (n = 1,000, three-seed mean) is 0.746, that of the readout calibrated on 1,000 labels is 0.666 (mean of 50 label pools 0.693) and that of the zero-shot readout 0.563; errors of two or more classes are 0.1%, 0.9% and 1.6%. In the MOVER subset, LoRA at n = 1,000 has an accuracy of 0.600 against 0.515 for the calibrated readout, and errors of two or more classes of 1.3% against 3.4%. The gap in κ is small mainly because quadratic weighting penalizes adjacent-class errors lightly and the cutpoints are fitted to maximize κ, not accuracy. “Close to LoRA with a few labels” should therefore be read as close in ordinal agreement (κ), not equal in case-level exact judgment.

This yields a decision rule with label number as the variable, under the assumption that labeling cost, model maintenance cost and data drift are not considered separately. For quadratic-weighted κ as the target: when a new site has fewer than about 300 labels (local) to 1,000 (MOVER, VitalDB), calibrate the readout first, and train LoRA when more labels are available. For case-level exact judgment and large errors as the target, consider LoRA from about 300 labels locally and from 100 labels in MOVER. The two are not exclusive: the calibrated readout can serve as an interim solution until labels reach the crossover, after which one switches. The κ crossovers for MOVER and VitalDB rely on extrapolating the calibration curve beyond k = 1,000 and are an order of magnitude only.

### 4.4. Natural-language guidance: a prior when there are no labels

The output can indeed be steered by natural language, and the predicted class mix moves with the direction of the wording, but the effect depends on input and wording. With structured variables only, all guidance lowers κ. Only when the input includes diagnosis text and the guidance describes the local rating habit accurately does κ rise to the level of 1,000-label calibration (0.557 versus 0.545), and this gain overlaps with the threshold adjustment that a few labels provide, so the two do not add up. Natural-language guidance can thus serve as a way to adjust the output tendency before labels are available, provided a reliable description of local habit exists; unsuitable guidance lowers accuracy.

### 4.5. Intended use and deployment

This is an offline retrospective method comparison and does not evaluate use of the model in a clinical workflow. If used clinically, the intended user would be the anesthesiologist, the setting preoperative assessment support, the level of autonomy assistive, and the result would have to be confirmed by the physician. For missing or abnormal input, the current approach is to write “unknown” in the template and read out as usual; whether to refuse output or prompt manual review was not tested. A practical deployment recipe is: begin with the zero-shot readout in natural order (or averaged over orders), collect about 100–300 local annotations to fit three cutpoints, then decide whether to move to LoRA by the rule in Section 4.3; when a site’s rating habit changes, only the cutpoints need refitting on recent annotations.

### 4.6. Limitations and boundary conditions

#### Reference standard and metric

The reference is the ASA class in each site’s anesthesia record, which carries inter-rater variation and institutional habit; the ceiling of κ is unknown, there was no independent rating, and we make no inference of “human-level performance” (the published inter-clinician κ is 0.713 [5]). Non-inferiority was tested on quadratic-weighted κ and the cutpoints were also fitted to maximize κ; LoRA is better in exact agreement and in errors of two or more classes, and closeness in κ must not be read as equality in case-level judgment. H2 is sensitive to the label pool, the LoRA seed and the implementation, which was addressed through label matching, label sampling and δ sensitivity, but LoRA has only three training pools, so its seed variation is still estimated coarsely.

#### Design and implementation

Cutpoint calibration, the OpenJev arm, scale sensitivity, the VitalDB replication, natural-language guidance and the analyses added after review were all introduced once the main results were known, and the study was not preregistered. The zero-shot readout is sensitive to option order and format; averaging over orders mitigates this, but averaging needs several forward passes, and other prompt presentations were not exhausted. T1 and T2 differ in language, format and information content at once, and the Chinese-T1 sensitivity analysis separates these factors only in part. The core evidence on Jev/System One is mostly preprints or benchmark reports, the base model and training of commercial Jev are undisclosed, and our conclusions concern the open implementation only and are not extrapolated to the commercial model. In the main analysis the readout ran in fp16 and LoRA in 4-bit QLoRA, so the precision is not matched; the fp16 LoRA was run once across two cards, only at local n = 1,000 (seed 11), the 4-bit readout was run locally and in MOVER, and the other sample sizes, seeds and data sources were not rerun. Most LoRA points have only three seeds and the 27B LoRA has one seed; the generative arm asks for a rationale and does not list options, so H1 and the “64-fold” figure compare the full generative usage with a closed-set readout, and an answer-only generative baseline was not run; the generative arm ran only on 1,000 (local) and 2,000 (MOVER) operations. The screen for contradictory generative output was a rule-based pass plus manual sampling, covering only structured fields and the first 400 characters of each rationale, and the resulting proportion is a lower bound; natural-language guidance was tested with one wording per direction, one base model and one site.

#### Generalizability and scope

Speed and cost were measured on a modified RTX 2080 Ti with expanded memory (22 GB) at batch size 1; absolute values cannot be extrapolated to other graphics cards, and only relative comparisons on the same hardware are made. Accuracy metrics such as κ do not depend on the card, but numerical differences between fp16 and other precisions were not tested separately; the readout ran in fp16 and LoRA in 4-bit with adapter, and token counts were not converted into floating-point operations or cost. There is only one base-model family (Qwen3.5), the ASA mix of VitalDB differs greatly from the other two sources, and the MOVER T2 may contain postoperative diagnoses; the subgroup analysis is descriptive with limited sample sizes and allows no conclusion about performance in different populations. The boundary conditions are closed-set classification, text input, site-specific thresholds and noisy record labels; the conclusions do not apply to open-ended generation, multimodal input, or settings with no local label at all (where only the zero-shot readout or natural-language guidance is available).

### 4.7. Conclusion

In this study the non-generative readout needs no training and, against generative zero-shot prompting that asks for a rationale, was about 64 times faster per case and generated no tokens. Fitting three cutpoints on about 100 to 1,000 local labels brought quadratic-weighted κ close to that of LoRA at a small label budget and removed the need to train and store an adapter for every site; the advantage of LoRA is a higher κ (by 0.03–0.06) and higher exact agreement once labels reach several hundred to several thousand. Non-inferiority holds only at the margin, depends on implementation, input information, option presentation, label sampling and LoRA seed, and should be treated as a conditional finding. Future work should evaluate how the approach is used in the clinical workflow in a prospective setting, replicate it on more base models and sites, and use independent ratings to test whether LoRA’s gain reflects discrimination or site fitting.

## Supporting information

Analysis code and aggregate results

## Data Availability

Local hospital data contain patient identifiers and cannot be shared. MOVER is available from its providers under a data use agreement. VitalDB is openly available (doi:10.13026/czw8-9p62). Analysis scripts, prompts and the OpenJev patch are provided in the supplementary code archive.

## Declarations

## Ethics approval

This study was approved by the Ethics Committee of the People’s Hospital of Xinjiang Uygur Autonomous Region. All procedures conformed to the Declaration of Helsinki. Because this study involved retrospective analysis of fully anonymized medical records, the requirement for written informed consent was waived; all patient data were anonymized before extraction and analysis, and access was restricted to the study investigators. MOVER and VitalDB are public databases and were used in accordance with their data use agreements.

## Registration

This was a retrospective study and was not prospectively registered.

## Data and code availability

The local hospital data contain patient information and cannot be shared. MOVER requires a data use agreement from its providers; VitalDB is public (doi:10.13026/czw8-9p62). Analysis scripts, the prompt texts of every configuration and the OpenJev patches are submitted as supplementary material.

## Funding

No external funding.

## Competing interests

All authors declare no competing interests.

## Author contributions (CRediT)

Yutong Shi: methodology, data curation, analysis, writing; Yang Wang: conceptualization, data extraction, verification; Alimujiang Simayi and Li Qu: clinical interpretation, revision; Guiping Xu: clinical interpretation, supervision, guarantor.

## Notes

### Competing Interest Statement

The authors have declared no competing interest.

### Author Declarations

Ethics committee/IRB of the People's Hospital of Xinjiang Uygur Autonomous Region gave ethical approval for this work. All procedures conformed to the Declaration of Helsinki. Because this study involved retrospective analysis of fully anonymized medical records, the requirement for written informed consent was waived; all patient data were anonymized before extraction and analysis, and access was restricted to the study investigators.

