## Supplementary material (S1-S25) for "Non-generative readout with few-label calibration versus LoRA fine-tuning of a large language model for ASA physical status classification: accuracy, speed, cost and label efficiency in three data sources"

2026-10-21

This supplement is submitted together with the main text. The numbering S1–S25 matches the “Supplementary materials list” of the main text. Unless stated otherwise, $\kappa$ is quadratic-weighted $\kappa$ and intervals are 95% confidence intervals from resampling by patient. Tables were generated from the analysis outputs; “Local” denotes the local hospital (People’s Hospital of Xinjiang Uygur Autonomous Region).

### S1 TRIPOD-LLM item self-check

##### TRIPOD-LLM self-check (English manuscript v4, 2026-10-20)

Item texts follow the official TRIPOD-LLM checklist (Gallifant et al., Nat Med 2025;31:60–69; 19 main items, 59 sub-items). Applicable modules: study design D (fine-tuning) + E (evaluation), task C (classification). Items that apply only to H (evaluation in clinical workflow), M (methodology) or QA/IR/DG/SS/MT are marked not applicable. Status: Yes = reported; Partial = partly reported or needs additions; No = not reported; N/A = not applicable. **This is the authors' self-check, not a formal compliance certification.**

| Item | Requirement (abridged) | Reported | Location / note |
|---|---|---|---|
| 2a | Title identifies LLM study, task, population, outcome | Yes | Title names the large language model, ASA physical status classification and the comparison; abstract states population and task |
| 2b | Abstract: medical background, use, rationale | Yes | Abstract, Background |
| 2c | Abstract: objective (development/fine-tuning/evaluation) | Yes | Abstract, Objective |
| 2d | Abstract: study setting | Yes | Abstract, Methods (three data sources) |
| 2e | Abstract: data, splits | Yes | Abstract, Methods |
| 2f | Abstract: LLM name and version | Yes | Qwen3.5-9B (Qwen/Qwen3.5-9B, downloaded 2026-10-16) |
| 2g | Abstract: LLM construction steps (M, D) | Yes | Abstract mentions LoRA and calibration |
| 2h | Abstract: task and input/output | Partial | The four-class ASA task is stated; the input (structured variables / diagnosis text) is given as T1/T2 only |
| 2i | Abstract: evaluation data and whether held out | Partial | Abstract states temporally external validation; the hold-out statement could be one clause longer |
| 2j | Abstract: main results and interpretation | Yes | |
| 2k | Abstract: wider implications or concerns | Yes | Abstract, Conclusions (conditional) |
| 2l | Registration number (H only) | N/A | Declarations: not preregistered |
| 3a | Background, use case, rationale | Yes | Introduction, paragraphs 1–4 |
| 3b | Target population and intended use (E, H) | Partial | Last paragraph of the Introduction and Section 4.5 state an offline method comparison; intended users and workflow are not elaborated because clinical workflow is not evaluated |
| 4 | Objectives (development/fine-tuning/validation) | Yes | Last paragraph of the Introduction, H1–H3 |
| 5a | Source of each dataset and reason for use | Yes | Section 2.2 |
| 5b | Quantitative description of data points and distributions | Yes | Section 2.3, Table 1, Supplementary Table S12 |
| 5c | Earliest/latest text date in development and evaluation data | Yes | MOVER 2017–2023, local hospital 2021–2026, VitalDB August 2016 to June 2017 (per the data paper) |
| 5d | Preprocessing and quality checks | Yes | Section 2.3, Preprocessing and missing data |
| 5e | Handling of missing and imbalanced data; exclusion reasons | Yes | Section 2.2 (exclusion counts), Section 2.3 |
| 6a | LLM name, version, date of last training or inference | Yes | Qwen/Qwen3.5-9B (Hugging Face, post-trained version), downloaded 2026-10-16; run 2026-10-17 to 20; weight checksums in S16 |
| 6b | Architecture, construction steps, hyperparameters (temperature, length limits) | Yes | Section 2.4 (greedy decoding, 140 tokens; LoRA hyperparameters) |

| Item | Requirement (abridged) | Reported | Location / note |
|---|---|---|---|
| 6c | Development process: training, fine-tuning, alignment (D) | Yes | Section 2.4 item 5, Section 2.5, Section 2.8 |
| 6d | Initial and post-processed outputs | Yes | Section 2.4 (hard-label parsing; closed-set probabilities; expected score and cutpoints) |
| 6e | How probabilities are determined and thresholds chosen (C) | Yes | Section 2.4 item 4; the difference in search grid is explained in Section 2.5 |
| 7a | Outcome definition, how predictions are computed, metrics (E) | Yes | Sections 2.3 and 2.8 |
| 7b | Qualification and guidance of subjective raters | Yes | Section 2.7 (a single investigator, stated) |
| 7c | Comparison with other LLMs, humans and benchmarks | Yes | Sections 2.4 and 2.8; no comparison with human reviewers (the outcome is each site's physician record) |
| 8a–8c | Annotation (if any) | N/A | No new annotation in this study; stated in Section 2.1 |
| 9a | Prompt design and selection process | Yes | Sections 2.4.1 and 3.9; full prompts in S6 |
| 9b | Data used to develop prompts | Yes | Section 2.4.1 |
| 10 | Preprocessing for summarization tasks | N/A | Not a summarization task |
| 11 | Instruction fine-tuning/alignment instructions and interface (D) | Partial | LoRA is trained with cross-entropy on the option letters and the prompt template is in S6; the absence of RLHF or a reward model could be stated in Section 2.4 item 5 |
| 12 | Compute or proxy (E, D) | Yes | Sections 2.6 and 3.3, Table 2 |
| 13 | Ethics committee and informed consent | Partial | Declared as a retrospective analysis of routine records of the local hospital; no approval number is reported |
| 14a | Funding and role of funders | Yes | ”No external funding” stated |
| 14b | Conflicts of interest | Yes | ”None” stated |
| 14c | Protocol access or statement that none was written (H) | Yes | Section 2.1 (required for H only) |
| 14d | Registration information or statement of non-registration (H) | Yes | Section 2.1, Declarations |
| 14e | Data availability | Yes | Declarations |
| 14f | Code availability | Yes | Scripts, prompt texts and patches submitted as supplementary material |
| 15 | Patient and public involvement (H) | Yes | Stated in Section 2.1 as not included |
| 16a | Data flow (documents/patients at each step) | Yes | Fig. 2 shows inclusion, exclusion and splits of the three sources with sample sizes at each step |
| 16b | Characteristics per source and split (E, H) | Yes | Table 1, Supplementary Table S12 |
| 16c | Comparison of predictor distributions in development and evaluation data (E) | Yes | Supplementary Table S12 |
| 16d | Numbers of cases and events per analysis (E) | Yes | Table 3 includes n; Sections 3.2–3.4 |
| 17 | Performance by prespecified metrics | Yes | Sections 3.2–3.8; non-prespecified parts are labeled |
| 18 | Results of LLM updates | N/A | No model updates; calibration and LoRA are comparison arms |
| 19a | Overall interpretation, including fairness | Partial | Sections 4.1–4.2; fairness: descriptive subgroups by sex and age (S17), exploratory only and not a basis for inference between populations |

| Item | Requirement (abridged) | Reported | Location / note |
|---|---|---|---|
| 19b | Limitations and their effect on bias, uncertainty, generalizability | Yes | Section 4.6 (reference standard, implementation dependence, post hoc analyses, generalizability and boundary conditions) |
| 19c | Known data challenges in task and domain (E, H) | Yes | Section 4.6 (reference standard, missingness, differences in mix) |
| 19d | Intended use: input, end user, level of autonomy (E, H) | Yes | Section 4.5: users are anesthesiologists, setting preoperative assessment support, assistive autonomy with physician confirmation |
| 19e | Handling of poor-quality or missing input (E, H) | Partial | Section 4.5: missing inputs are written as unknown and read out as usual; whether to refuse output was not tested (stated) |
| 19f | Whether users must take part in input processing (E, H) | Yes | Section 4.5: users do not process input; the physician confirms the output |
| 19g | Next steps for research | Yes | Last sentence of Section 4.7 |

#### S1.1 Summary

- 55 table rows (8a–8c merged into one): Yes 44; Partial 7; N/A 4; No 0.
- Still partial: 2h/2i (abstract could add a clause), 3b (intended use is an offline method comparison only), 11 (LoRA without RLHF/reward model could be stated), 13 (ethics: declared as a retrospective analysis, no approval number reported), 19a (subgroups only descriptive), 19e (refusal policy for missing input not tested).
- Reporting risks outside the checklist: H2 is sensitive to the label pool, the LoRA seed, the implementation and the input language, which is disclosed in the abstract and in Sections 3.4 and 4.1; sensitivity to option order was mitigated by multi-order averaging and calibration (Sections 3.9 and 4.2).

#### S2 Frozen analysis-plan files and hashes

The analysis plan was frozen as a document before the evaluation at the local hospital began; afterwards no method choice, comparison arm or primary metric was changed on the basis of the local validation set. File B is a working document recording methodological revisions made after the first MOVER results; it was not used as the frozen plan. Both files are written in Chinese.

- **File A: "Evaluation plan for the local validation set (frozen version)".** Data roles and splits, comparison arms and fixed hyperparameters, primary metrics and testing hierarchy, and prior exposures that must be disclosed. Frozen on 2026-10-18 23:40.
- **File B: "Study framework v2 (working document)".** Record of methodological adjustments after the first MOVER results, in use from 2026-10-18; checksum of the version at submission below.

SHA-256 (computed on the file content at submission):

```
A  09da7c7feb3435205fcdc8ddc10b664f3faf2c188b5f85bc013a4f3b81943245
B  b31332922a9783004df0fc9bf8683b878905f07207dcf3159c3c1058c924e705
```

### S3 Scores of the eight OpenJev tuning configurations

##### OpenJev configuration tuning (MOVER tuning set, n = 800; all configurations listed)

| System prompt | Option description | Question type | Uncalibrated κ | 3-cutpoint calibrated κ (2-fold) | Macro AUC | Predicted distribution |
|---|---|---|---|---|---|---|
| generic | short | choice | 0.333 | 0.457 | 0.751 | [0, 600, 159, 41] |
| generic | short | score | 0.412 | 0.458 | 0.747 | [0, 424, 333, 43] |
| generic | official | choice | 0.346 | 0.475 | 0.732 | [0, 532, 240, 28] |
| generic | official | score | 0.387 | 0.448 | 0.729 | [0, 506, 261, 33] |
| anes | short | choice | 0.355 | 0.499 | 0.756 | [25, 589, 129, 57] |
| anes | short | score | 0.402 | 0.501 | 0.754 | [11, 505, 229, 55] |
| anes | official | choice | 0.381 | 0.488 | 0.736 | [0, 536, 212, 52] |
| anes | official | score | 0.367 | 0.488 | 0.742 | [0, 536, 228, 36] |

Selected configuration (frozen): system prompt anes; option description short; question type score; uncalibrated κ = 0.402, 2-fold calibrated κ = 0.501, macro AUC = 0.754.

### S4 Calibration curves, temperature scaling and ECE

**Zero-shot readout + cutpoint calibration: curve over the number of calibration labels (LoRA at the same label numbers for comparison)**

#### S4.1 MOVER (sub4000); task LoRA for comparison: n = 100 0.410, 300 0.411, 1000 0.505, 3000 0.563

| Calibration labels k | Mean κ | 2.5% | 97.5% |
|---|---|---|---|
| 0 (uncalibrated argmax) | 0.473 | — | — |
| 25 | 0.467 | 0.372 | 0.519 |
| 50 | 0.495 | 0.422 | 0.530 |
| 100 | 0.508 | 0.476 | 0.528 |
| 200 | 0.517 | 0.495 | 0.529 |
| 500 | 0.521 | 0.516 | 0.529 |
| 1000 | 0.524 | 0.524 | 0.524 |

#### S4.2 Local validation set; local LoRA for comparison: n = 100 0.423, 300 0.542, 1000 0.559, 4326 0.561

| Calibration labels k | Mean κ | 2.5% | 97.5% |
|---|---|---|---|
| 0 (uncalibrated argmax) | 0.443 | — | — |
| 25 | 0.491 | 0.384 | 0.546 |
| 50 | 0.516 | 0.471 | 0.547 |
| 100 | 0.526 | 0.474 | 0.547 |
| 200 | 0.528 | 0.501 | 0.544 |
| 500 | 0.535 | 0.520 | 0.540 |
| 1000 | 0.540 | 0.540 | 0.540 |

Note: the curve script uses a coarser cutpoint search grid (61 points, 2 passes) than the main analysis (121 points, 3 passes), so k = 1000 gives 0.540 locally against 0.534 in the main analysis. The points are subsets of one 1,000-label pool and are not independent.

### S5 Scale sensitivity (including Qwen3.5-27B)

##### Scale sensitivity of the frozen OpenJev (4-bit NF4, MOVER subset n = 2000, 1444 patients)

| Model | κ (argmax) | κ (rounded expectation) | Macro AUC | Predicted distribution (argmax) |
|---|---|---|---|---|
| Qwen3.5-9B | 0.444 | 0.474 | 0.762 | [92, 1240, 390, 278] |
| Qwen3.5-27B | 0.387 | 0.402 | 0.731 | [518, 795, 567, 120] |

- 27B − 9B, κ (argmax): -0.057 (-0.080 ~ -0.033)
- 27B − 9B, κ (rounded expectation): -0.073 (-0.095 ~ -0.049)
- 27B − 9B, macro AUC: -0.031 (-0.041 ~ -0.021, 300 resamples)

### S6 Full prompts, prompt variants and option-order/format sensitivity

This section gives the prompt texts actually used by each comparison arm. Prompts were frozen before the evaluation sets were touched; square brackets mark where the patient input is inserted. T1 is the structured-variable template shared by the data sources, T2 adds preoperative diagnosis text.

#### S6.1 In-house readout and task LoRA (same template)

The readout arm reads the output logits at the last position for the four candidate letters A–D and turns them into closed-set probabilities; the LoRA arm uses the same template for training and inference, with cross-entropy on the one of the four positions A–D that corresponds to the label.

```
You are an anesthesiologist. Based on the preoperative information below, assign the ASA Physical
Status class.

[patient input]

Options:
A. ASA I
B. ASA II
C. ASA III
D. ASA IV-V
Answer:
```

#### S6.2 Generative zero-shot

The chat template of Qwen3.5-9B was used with thinking mode off, greedy decoding and at most 140 new tokens; the trailing line "Final answer: ASA ..." is parsed from the output.

```
You are an anesthesiologist. Based on the preoperative information below, assign the ASA Physical
Status class (I, II, III, IV-V).

[patient input]

Give a brief rationale (at most 3 sentences), then end with exactly one line: 'Final answer: ASA
<I|II|III|IV-V>'.
```

#### S6.3 Natural-language guidance (readout arm)

The guidance text is inserted before the patient input and concatenated with the readout template for the same model; the wording was fixed before the validation results were seen.

**G0 (no guidance)**

No additional guidance text.

**GOFF (ASA 2020 official definitions)**

```
Definitions (ASA Physical Status, 2020):
I: a normal healthy patient (healthy, non-smoking, no or minimal alcohol use).
II: mild systemic disease without substantive functional limitation (current smoker,
social drinker, pregnancy, obesity 30<BMI<40, well-controlled DM/HTN, mild lung disease).
III: severe systemic disease with substantive functional limitation (poorly controlled DM
or HTN, COPD, morbid obesity BMI>=40, active hepatitis, alcohol dependence, implanted
pacemaker, moderate reduction of ejection fraction, ESRD on regular dialysis, history >3
months of MI, CVA, TIA or coronary stents, malignancy).
```

```
IV: severe systemic disease that is a constant threat to life (recent <3 months MI, CVA,
TIA or stents, ongoing cardiac ischemia, severe valve dysfunction, severe reduction of
ejection fraction, shock, sepsis, DIC, ARDS, ESRD not on regular dialysis).
V: moribund patient not expected to survive without the operation.
```

**GLOC (local recording habit)**

```
Guidance: classify the way anesthesiologists at this hospital routinely record ASA for
abdominal tumor surgery. Their habits: ASA I is rarely recorded; a patient who looks
healthy but is undergoing tumor or major abdominal surgery is usually recorded as II.
Age-related organic changes in elderly patients (atherosclerotic plaques, lacunar
infarcts, mild valvular regurgitation, diastolic dysfunction, mild obstructive or
interstitial lung changes) are usually recorded as III. Well-controlled hypertension or
non-insulin diabetes alone is II; uncontrolled disease, several comorbid systems,
insulin-treated diabetes, coronary artery disease or moderate lung disease is III; ASA IV
is reserved for recent MI or stroke, shock, sepsis, severe cardiac, renal or hepatic
failure.
```

**GSTR (strict adherence to the guideline)**

```
Guidance: follow the ASA Physical Status definitions strictly and literally. A patient
without any systemic disease is ASA I even if the operation is major or the patient is an
adult of any age. Do not upgrade a class because of age, cancer status or the size of the
surgery alone. Mild or well-controlled single-system disease is ASA II; only disease with
real functional limitation is ASA III; ASA IV requires a constant threat to life.
```

#### S6.4 OpenJev (frozen configuration)

The system prompt is the anesthesia version:

```
You are an experienced anesthesiologist acting as a System One decision model. You are shown a STATE
(preoperative information) and a QUESTION. Select exactly one option by its label only. Be
calibrated: judge the ASA Physical Status from documented systemic disease and its functional
impact, not from the type of surgery alone.
```

The other system prompt (generic, the OpenJev default) is:

```
You are a System One decision model. You are shown a STATE and a QUESTION about it. Answer by
selecting exactly one of the listed options. Reply with the option label only. Be calibrated: when
the state does not settle the question, do not pretend it does.
```

The frozen configuration is the score question type with short option descriptions. The rendered user message is:

```
## State
[patient input]

## Question
What is the ASA Physical Status class of this patient? (0 = ASA I ... 3 = ASA IV-V)

## Options
```

```
0. ASA I: A normal healthy patient
1. ASA II: A patient with mild systemic disease
2. ASA III: A patient with severe systemic disease
3. ASA IV-V: A patient with severe systemic disease that is a constant threat to life, or a moribund
patient

Levels are ordered from 0 (lowest) to 3 (highest). Reply with the level number only.
```

The full texts of the two option descriptions follow (official gives ASA's official examples; short is used by the frozen configuration):

**short**

```
0. ASA I: A normal healthy patient
1. ASA II: A patient with mild systemic disease
2. ASA III: A patient with severe systemic disease
3. ASA IV-V: A patient with severe systemic disease that is a constant threat to life, or
a moribund patient
```

**official**

```
0. ASA I: A normal healthy patient. Healthy, non-smoking, no or minimal alcohol use.
1. ASA II: Mild systemic disease without substantive functional limitation. Examples:
current smoker, social alcohol drinker, pregnancy, obesity (30<BMI<40), well-controlled
DM/HTN, mild lung disease.
2. ASA III: Severe systemic disease with substantive functional limitation. Examples:
poorly controlled DM or HTN, COPD, morbid obesity (BMI>=40), active hepatitis, alcohol
dependence, implanted pacemaker, moderately reduced EF, ESRD on regular dialysis, history
(>3 months) of MI, CVA, TIA or CAD/stents, malignancy.
3. ASA IV-V: Severe systemic disease that is a constant threat to life, or a moribund
patient. Examples: recent (<3 months) MI, CVA, TIA or CAD/stents, ongoing cardiac
ischemia, severe valve dysfunction, severely reduced EF, shock, sepsis, DIC, ARDS, ESRD
not on regular dialysis, ruptured aneurysm, massive trauma.
```

The other question type, choice, uses the same system prompt and option descriptions, with option labels ASA I–ASA IV-V, candidates numbered 1–4 and the closed-set ending "Reply with the option number only."

All configurations use a single forward pass that scores fixed candidate strings and generate no text; the scores of the eight configurations are in S3.

#### S6.5 Prompt variants and option-order/format sensitivity

**Three prompt variants of generative zero-shot** (kappa, accuracy, macro AUC, predicted distribution and true distribution on 1,500 MOVER development operations; run time in parentheses)

```
llm P0: kappa 0.454 accuracy 0.471 macroAUC 0.755 predicted [220, 704, 425, 151] true [80,
473, 772, 175] (174s)
llm P1: kappa 0.415 accuracy 0.455 macroAUC 0.757 predicted [414, 409, 545, 132] true [80,
473, 772, 175] (376s)
llm P2: kappa 0.325 accuracy 0.363 macroAUC 0.734 predicted [519, 539, 373, 69] true [80,
473, 772, 175] (493s)
```

**Option-order permutation robustness (zero-shot; orders: orig = I, II, III, IV-V; rev = reversed; shift2 = cyclic shift by 2)**

| Arm | Data | n | κ orig | κ rev | κ shift2 | Flip rate rev | Flip rate shift2 | κ of averaged probability (debiased) | Mean max probability orig |
|---|---|---|---|---|---|---|---|---|---|
| Readout zero-shot 9B | MOVER sub | 2000 | 0.476 | 0.257 | 0.228 | 0.518 | 0.361 | 0.340 | 0.551 |
| Readout zero-shot 9B | Local val | 2000 | 0.434 | 0.252 | 0.143 | 0.438 | 0.334 | 0.286 | 0.544 |

**Option label format (exploratory; OpenJev backend, frozen prompt wording)**

##### S6.6 First 1,500 operations of the MOVER subset

| Format | κ | Macro AUC | Predicted distribution |
|---|---|---|---|
| digits | 0.384 | 0.745 | [106, 1025, 240, 129] |
| letters | 0.355 | 0.735 | [153, 1055, 163, 129] |
| roman | 0.369 | 0.733 | [216, 912, 264, 108] |

Flip rate between formats: digits-letters 0.095; digits-roman 0.105; letters-roman 0.111

##### S6.7 1,000 random operations of the local validation set

| Format | κ | Macro AUC | Predicted distribution |
|---|---|---|---|
| digits | 0.276 | 0.749 | [71, 798, 113, 18] |
| letters | 0.259 | 0.729 | [131, 775, 76, 18] |
| roman | 0.275 | 0.724 | [207, 670, 109, 14] |

Flip rate between formats: digits-letters 0.097; digits-roman 0.162; letters-roman 0.109

### S7 Method of the screen for contradictory content and the 40 manually checked samples

##### Rule-based screen for "hallucination" in generative zero-shot rationales

A rule-based screen gives only a lower bound or rough estimate, and "unsupported" means that the input template contains no information supporting the statement; it is not necessarily clinically wrong. The manual check on a random sample is described at the end.

| Data | Rationales | Unparsable output | With contradicting statement | With unsupported statement | Either |
|---|---|---|---|---|---|
| MOVER subset | 2000 | 22 (1.1%) | 62 (3.1%) | 115 (5.8%) | 171 (8.6%) |
| Local val subset (T1 English template) | 1000 | 1 (0.1%) | 41 (4.1%) | 39 (3.9%) | 80 (8.0%) |

#### S7.1 Manual check (40 flagged rationales drawn at random and read one by one)

- Many rule-based flags were false alarms, for example a statement that goes beyond "no history of hypertension, CAD or diabetes", "BMI 29.2 is overweight, not severely obese", and procedure names that themselves contain "neoplasm" all being counted as unsupported. **The proportions in the table above cannot be read directly as a hallucination rate.**
- Of the 40, **10 (25%) truly contradicted the input**, all being cases where the input said `hypertension no` and the rationale said "controlled hypertension" or "history of hypertension". A further 4 described a BMI of 27.9–29.2 as "mild obesity/obese" (a minor factual error, 10%). The remaining 26 were false alarms or reasonable inferences from values such as weight, creatinine and blood pressure.
- From this, the proportion of rationales containing **a structured fact that contradicts the input** is estimated at about **2%** ($0.25 \times 251/3000 \approx 2.1\%$; sample $n = 40$, 95% CI about 1–3%); counting the BMI misjudgments it is about 3%. In addition, **unparsable output was 1.1% (MOVER) and 0.1% (local)**.
- Limitations: only structured fields were audited (hypertension, coronary disease, diabetes, BMI, etc.); other fabricated content in the narrative is not covered; the recall of the rules is unknown, so 2% is a lower bound. The non-generative readout outputs no text, its output is limited to a closed set, its unparsable rate is 0 and there is no narrative that could be fabricated; this is guaranteed by construction.

#### S7.2 The 40 manually checked samples

| Data | op_id | Flag | Rationale (truncated) | Input history line |
|---|---|---|---|---|
| MOVER subset | 0e232c7ebce00c7f | unsupported:obesity | This 60-year-old male presents with significant obesity (195 kg) and uncontrolled hypertension (170/81 mmHg), but has no history of diabetes, coronary artery disease, or documented organ dysfunction. Although the blood pressure is elevated, the absence of know | History: hypertension yes, coronary artery disease no, diabetes no |
| Local val subset (T1 English template) | L-d78a8521 | unsupported:obesity | The patient is a 55-year-old female with well-controlled hypertension and no history of diabetes, coronary artery disease, or significant organ dysfunction, as evidenced by normal renal function and stable preoperative vitals. Her BMI of 28.7 indicates mild ob | History: hypertension yes, coronary artery disease no, diabetes no |

| Data | op_id | Flag | Rationale (truncated) | Input history line |
|---|---|---|---|---|
| Local val subset (T1 English template) | L-e3805b26 | contradicted:hypertension | The patient is an 80-year-old male with no history of systemic disease such as hypertension, coronary artery disease, or diabetes, and his preoperative labs show only mild, non-chronic renal elevation without other comorbidities. Although his blood pressure is | History: hypertension no, coronary artery disease no, diabetes no |
| MOVER subset | 8fb52b0ef79a184f | unsupported:cancer | The patient is a 69-year-old female with no significant history of systemic disease such as hypertension, coronary artery disease, or diabetes, despite elevated preoperative blood pressure. Although the planned craniotomy for neoplasm excision is a major proce | History: hypertension no, coronary artery disease no, diabetes no |
| MOVER subset | 1b2b171d7a050edd | unsupported:renal disease | The patient is a 48-year-old male with a history of hypertension but no other significant systemic disease, presenting with stable vital signs for an outpatient procedure. Although the planned insertion of a peritoneal dialysis catheter suggests underlying ren | History: hypertension yes, coronary artery disease no, diabetes no |
| Local val subset (T1 English template) | L-61ed1a6b | contradicted:hypertension | The patient is a 41-year-old female with controlled hypertension and no history of systemic disease, which typically suggests ASA I or II status. However, the preoperative labs reveal significant abnormalities, specifically anemia (hemoglobin 9.0 g/dL) and hyp | History: hypertension no, coronary artery disease no, diabetes no |
| MOVER subset | d326154c5bb4f9dc | unsupported:obesity | The patient is a 37-year-old male with no significant systemic disease, normal preoperative vital signs, and a BMI of 29.2, which represents overweight status rather than severe obesity or a comorbid condition. Although the planned procedure (liposuction) is e | History: hypertension no, coronary artery disease no, diabetes no |
| Local val subset (T1 English template) | L-a756c0b7 | contradicted:diabetes; contradicted:hypertension; contradicted:coronary | The patient is a 62-year-old male with a history of hypertension, coronary artery disease, and diabetes explicitly denied, presenting with stable vital signs and normal renal function. Although he has a BMI of 33.3 indicating obesity and mild hypoalbuminemia, | History: hypertension no, coronary artery disease no, diabetes no |
| Local val subset (T1 English template) | L-bc8da5a8 | unsupported:obesity | The patient is a 53-year-old female with well-controlled hypertension and no history of diabetes, coronary artery disease, or significant organ dysfunction, as evidenced by normal renal function and stable preoperative vitals. Her BMI of 28.4 indicates mild ob | History: hypertension yes, coronary artery disease no, diabetes no |
| MOVER subset | b10b56aa06e1620b | unsupported:obesity | The patient is a 42-year-old female with significant obesity (111 kg) and a history of both hypertension and diabetes, indicating systemic disease that limits her functional capacity. Although she has no history of coronary artery disease, the presence of mult | History: hypertension yes, coronary artery disease no, diabetes yes |
| Local val subset (T1 English template) | L-71a52495 | contradicted:hypertension | The patient is a 69-year-old male with controlled hypertension but no history of significant systemic disease such as coronary artery disease, diabetes, or renal insufficiency. His preoperative vital signs and laboratory results are within normal limits, indic | History: hypertension no, coronary artery disease no, diabetes no |
| Local val subset (T1 English template) | L-93652f35 | unsupported:renal disease | The patient is a 63-year-old male with well-controlled hypertension and mild renal insufficiency (creatinine 1.39 mg/dL) but no history of diabetes, coronary artery disease, or other severe systemic disease. His preoperative vital signs are stable, and his lab | History: hypertension yes, coronary artery disease no, diabetes no |

| Data | op_id | Flag | Rationale (truncated) | Input history line |
|---|---|---|---|---|
| MOVER subset | 7e796e2a6f50ac40 | contradicted:hypertension; unsupported:renal disease | The patient is a 62-year-old male with controlled hypertension and mild renal insufficiency (creatinine 1.30 mg/dL) but no history of systemic disease requiring treatment. His preoperative vitals are stable, and he has no significant comorbidities that would l | History: hypertension no, coronary artery disease no, diabetes no |
| Local val subset (T1 English template) | L-59cc2147 | contradicted:hypertension | The patient is a 41-year-old female with no significant systemic disease, normal vital signs, and well-controlled preoperative laboratory values. Although she has a history of hypertension, the prompt explicitly states "hypertension no," indicating she is curr | History: hypertension no, coronary artery disease no, diabetes no |
| MOVER subset | 08c05648f3e7ea9f | unsupported:cancer | The patient is a 44-year-old female with no significant systemic disease, normal vital signs, and unremarkable preoperative laboratory results. Although the planned craniotomy for neoplasm excision is a major surgical procedure, the ASA Physical Status classif | History: hypertension no, coronary artery disease no, diabetes no |
| MOVER subset | ddce5f177dee81ca | unsupported:cancer | The patient is a 39-year-old male with no significant systemic disease, normal preoperative vital signs, and no history of hypertension, coronary artery disease, or diabetes. Although the planned craniectomy for neoplasm excision is a major surgery, the ASA Ph | History: hypertension no, coronary artery disease no, diabetes no |
| Local val subset (T1 English template) | L-3fcf5ae5 | contradicted:hypertension | The patient is a 63-year-old male with controlled hypertension but no history of systemic disease such as coronary artery disease, diabetes, or renal insufficiency, as evidenced by normal creatinine and hemoglobin levels. His preoperative vital signs are stabl | History: hypertension no, coronary artery disease no, diabetes no |
| MOVER subset | 81685e30a98a4fca | unsupported:cancer | The patient is a 42-year-old female with no significant systemic disease such as hypertension, coronary artery disease, or diabetes, and her preoperative vitals are stable. Although the planned craniotomy for neoplasm excision is a major surgery, the ASA Physi | History: hypertension no, coronary artery disease no, diabetes no |
| MOVER subset | 9d0ab528cf6e6751 | unsupported:cancer | The patient is a 28-year-old male with no significant systemic disease, normal vital signs, and unremarkable preoperative laboratory results, indicating a fully compensated healthy individual. Although he has a low BMI, this does not constitute a systemic dise | History: hypertension no, coronary artery disease no, diabetes no |
| MOVER subset | 8214c280a144b378 | unsupported:cancer | The patient is a 50-year-old female with a history of hypertension but no other significant systemic disease, placing her in a generally good health status despite the elevated preoperative blood pressure. The planned procedure is a craniotomy with neoplasm ex | History: hypertension yes, coronary artery disease no, diabetes no |
| Local val subset (T1 English template) | L-e0159fc8 | contradicted:hypertension | The patient is a 54-year-old female with controlled hypertension and no history of systemic disease such as coronary artery disease or diabetes. Her preoperative laboratory values, including normal creatinine and hemoglobin, indicate no significant organ dysfu | History: hypertension no, coronary artery disease no, diabetes no |

| Data | op_id | Flag | Rationale (truncated) | Input history line |
|---|---|---|---|---|
| MOVER subset | b394fa5722fd8f30 | unsupported:renal disease | The patient is a 38-year-old male with no history of systemic disease, but preoperative labs reveal significant abnormalities including anemia (Hb 8.1 g/dL), hypoalbuminemia (1.9 g/dL), and elevated creatinine (1.30 mg/dL) suggestive of renal impairment. These | History: hypertension no, coronary artery disease no, diabetes no |
| MOVER subset | 06a3d02f3e435f47 | unsupported:renal disease | The patient is a 20-year-old male with no significant systemic disease such as hypertension, coronary artery disease, or diabetes, which would otherwise elevate his status. Although he has a BMI of 32.9 (obesity) and mild renal insufficiency indicated by a cre | History: hypertension no, coronary artery disease no, diabetes no |
| Local val subset (T1 English template) | L-e30aad9d | contradicted:hypertension | The patient is a 47-year-old female with a BMI of 29.2 and no significant systemic disease, as evidenced by normal blood pressure, heart rate, and laboratory values including renal function and hemoglobin. Although she has a history of hypertension, the preope | History: hypertension no, coronary artery disease no, diabetes no |
| MOVER subset | 349fc0e9b4ed770e | unsupported:cancer | The patient is a 59-year-old female with no significant history of systemic disease such as hypertension, coronary artery disease, or diabetes, and her preoperative vital signs are stable. Although the planned procedure involves a spine laminectomy with neopla | History: hypertension no, coronary artery disease no, diabetes no |
| MOVER subset | 2ad6ee0e7ea9ee10 | unsupported:obesity | The patient is a 46-year-old male with no significant systemic disease such as hypertension, coronary artery disease, or diabetes, and his preoperative vitals are stable. Although he is obese (BMI 27.9) and undergoing surgery for a fracture, these factors do n | History: hypertension no, coronary artery disease no, diabetes no |
| Local val subset (T1 English template) | L-875a894f | contradicted:hypertension | The patient is a 63-year-old female with a history of coronary artery disease but no other significant systemic diseases, which aligns with the definition of ASA Physical Status II. Her preoperative labs are within normal limits, and her vital signs show only | History: hypertension no, coronary artery disease yes, diabetes no |
| Local val subset (T1 English template) | L-91cea2db | unsupported:cancer | The patient is a 45-year-old female with a BMI of 32.5 (obesity) but no significant systemic disease such as hypertension, diabetes, or coronary artery disease. Her preoperative vital signs and laboratory results are within normal limits, indicating good physi | History: hypertension no, coronary artery disease no, diabetes no |
| MOVER subset | 3e07baf8fab4fbd2 | contradicted:hypertension | The patient is a 74-year-old male with controlled hypertension and no history of significant systemic disease, placing him in a generally good functional state. Although he presents with mild anemia (hemoglobin 8.9 g/dL) and hypoalbuminemia (2.7 g/dL), these f | History: hypertension no, coronary artery disease no, diabetes no |
| Local val subset (T1 English template) | L-728761a1 | unsupported:obesity | The patient is a 69-year-old female with no significant systemic disease, normal vital signs, and unremarkable preoperative laboratory results, including normal renal function and hemoglobin levels. Although she has a BMI of 29.0, this represents mild obesity | History: hypertension no, coronary artery disease no, diabetes no |

| Data | op_id | Flag | Rationale (truncated) | Input history line |
|---|---|---|---|---|
| Local val subset (T1 English template) | L-c35bfb5a | unsupported:obesity | The patient is a 44-year-old male with no significant systemic disease, normal vital signs, and unremarkable preoperative laboratory results. Although he has a BMI of 27.1, this falls within the overweight range rather than obesity, and there is no evidence of | History: hypertension no, coronary artery disease no, diabetes no |
| MOVER subset | 8461e6a7c87a3ace | contradicted:diabetes; contradicted:hypertension; contradicted:coronary | The patient is a 57-year-old male with no significant systemic disease, normal vital signs, and unremarkable preoperative laboratory results. His history explicitly excludes hypertension, coronary artery disease, and diabetes, and his BMI is within the normal | History: hypertension no, coronary artery disease no, diabetes no |
| MOVER subset | 74497e11a81e0395 | contradicted:hypertension | The patient is a 59-year-old female with controlled hypertension and no history of systemic disease, placing her in a good functional status. Although she has mild hypoalbuminemia (2.8 g/dL), her renal function and hemoglobin are stable, and her vital signs ar | History: hypertension no, coronary artery disease no, diabetes no |
| Local val subset (T1 English template) | L-ee5e50b0 | unsupported:cancer | The patient is a 75-year-old female with no significant history of systemic disease such as hypertension, coronary artery disease, or diabetes, and her preoperative labs show normal renal function and hemoglobin levels. Although her albumin is slightly low at | History: hypertension no, coronary artery disease no, diabetes no |
| MOVER subset | b037a0b23c2f8336 | unsupported:obesity | The patient is a 65-year-old female with no significant history of systemic disease such as hypertension, coronary artery disease, or diabetes, and her preoperative labs are unavailable but do not indicate known organ dysfunction. Although she has a BMI of 29. | History: hypertension no, coronary artery disease no, diabetes no |
| Local val subset (T1 English template) | L-e488b09d | contradicted:hypertension | The patient is a 66-year-old male with a history of hypertension but no other significant systemic diseases such as coronary artery disease or diabetes. His preoperative laboratory values, including normal creatinine and hemoglobin, indicate good organ functio | History: hypertension no, coronary artery disease no, diabetes no |
| MOVER subset | e2adc29107645c69 | unsupported:obesity | The patient is a 78-year-old female with no significant systemic disease such as hypertension, coronary artery disease, or diabetes, and her preoperative vital signs are stable. Although her age and obesity (109 kg) present increased perioperative risks, these | History: hypertension no, coronary artery disease no, diabetes no |
| MOVER subset | 91c9386445422815 | contradicted:hypertension | The patient is a 38-year-old male with no significant systemic disease, but he presents with significant physiological derangements including symptomatic tachycardia (119 bpm), hypertension, anemia (Hb 8.2 g/dL), and hypoalbuminemia (2.7 g/dL). These findings | History: hypertension no, coronary artery disease no, diabetes no |
| MOVER subset | efa36c3db536e0fa | contradicted:hypertension | The patient presents with severe anemia (hemoglobin 5.6 g/dL) and uncontrolled hypertension (BP 168/84 mmHg), indicating significant systemic disease that limits her functional capacity and poses a substantial risk for major surgery. Although the planned proce | History: hypertension no, coronary artery disease no, diabetes no |

| Data | op_id | Flag | Rationale (truncated) | Input history line |
|---|---|---|---|---|
| Local val subset (T1 English template) | L-98bbc9a6 | unsupported:renal disease | The patient is a 53-year-old female with no history of systemic disease such as hypertension, coronary artery disease, or diabetes, which would otherwise elevate her status. However, her preoperative labs reveal significant abnormalities including anemia (hemo | History: hypertension no, coronary artery disease no, diabetes no |

### S8 Retrieval-based few-shot kNN and XGBoost supplementary arms (MOVER evaluation set)

**v3 evaluation (MOVER patient-level subset; common sample n = 4000, 2606 patients; cluster bootstrap B = 1000)**

#### S8.1 A. Arms (seed0 predictions; AUC reported where available)

| Arm | κ (95% CI) | Accuracy | Macro AUC |
|---|---|---|---|
| Readout zero-shot 9B (T1) | 0.473 (0.444–0.500) | 0.464 | 0.762 |
| Readout zero-shot 9B (T2, with diagnosis history) | 0.567 (0.542–0.592) | 0.526 | 0.813 |
| Readout zero-shot Qwen3.5-27B (T1) | 0.489 (0.461–0.514) | 0.423 | 0.786 |
| Readout zero-shot Qwen3.8-27B (T1) | 0.115 (0.096–0.134) | 0.162 | 0.664 |
| Retrieval-based few-shot kNN-8 | 0.302 (0.275–0.328) | 0.345 | 0.759 |
| Readout 9B (T1) + 1000-label cutpoint calibration | 0.524 (0.498–0.549) | 0.515 | — |
| Readout 9B (T2) + 1000-label cutpoint calibration | 0.605 (0.583–0.628) | 0.526 | — |
| Readout Qwen3.5-27B + cutpoint calibration | 0.559 (0.533–0.584) | 0.545 | — |
| Readout Qwen3.8-27B + cutpoint calibration | 0.418 (0.387–0.445) | 0.484 | — |
| XGBoost (full training set) | 0.502 (0.474–0.532) | 0.606 | 0.813 |
| LoRA 12k (T1) | 0.582 (0.557–0.606) | 0.652 | 0.849 |
| LoRA (T1) n = 1000 | 0.527 (0.501–0.550) | 0.613 | 0.810 |
| LoRA (T1) n = 3000 | 0.563 (0.537–0.585) | 0.640 | 0.837 |
| LoRA (T2) n = 1000 | 0.586 (0.562–0.610) | 0.631 | 0.842 |
| LoRA (T2) n = 3000 | 0.626 (0.601–0.648) | 0.670 | 0.867 |
| LoRA Qwen3.5-27B (T1) n = 1000 | 0.556 (0.531–0.579) | 0.617 | 0.818 |
| LoRA Qwen3.5-27B (T1) n = 3000 | 0.583 (0.557–0.604) | 0.639 | 0.841 |
| LoRA Qwen3.8-27B (T1) n = 1000 | 0.522 (0.496–0.545) | 0.601 | 0.810 |
| LoRA Qwen3.8-27B (T1) n = 3000 | 0.590 (0.567–0.612) | 0.638 | 0.839 |

#### S8.2 B. Learning curve: mean κ [range] (number of seeds)

| Training labels | LoRA (T1) | XGBoost |
|---|---|---|
| 100 | 0.410 [0.394–0.438] (3) | 0.234 |
| 300 | 0.411 [0.256–0.500] (3) | 0.305 |
| 1000 | 0.505 [0.462–0.527] (3) | 0.395 |
| 3000 | 0.563 [0.551–0.575] (3) | 0.469 |
| 12000 | 0.583 [0.580–0.587] (3) | 0.504 |

#### S8.3 C. Paired contrasts (κ difference, cluster-bootstrap 95% CI; seed0)

- LoRA 12k (T1) – readout zero-shot 9B (T1): +0.109 (+0.085 ~ +0.132)
- Readout 9B (T1) + 1000-label cutpoint calibration – LoRA (T1) n = 1000: -0.003 (-0.023 ~ +0.019) (non-inferiority margin $\delta = 0.05$: lower bound of the difference must be > –0.05)
- Readout 9B (T1) + 1000-label cutpoint calibration – LoRA (T1) n = 3000: -0.038 (-0.059 ~ -0.017)
- LoRA Qwen3.5-27B (T1) n = 1000 – LoRA (T1) n = 1000: +0.029 (+0.014 ~ +0.045)
- LoRA Qwen3.5-27B (T1) n = 3000 – LoRA (T1) n = 3000: +0.021 (+0.009 ~ +0.033)

- Readout zero-shot 9B (T2, with diagnosis history) – readout zero-shot 9B (T1): +0.095 (+0.069 ~ +0.121)
- LoRA (T2) n = 3000 − LoRA (T1) n = 3000: +0.064 (+0.047 ~ +0.080)
- Readout 9B (T2) + 1000-label cutpoint calibration − LoRA (T2) n = 1000: +0.019 (+0.002 ~ +0.036) (T2, non-inferiority $\delta = 0.05$)
- LoRA (T2) n = 3000 − readout 9B (T2) + 1000-label cutpoint calibration: +0.020 (+0.002 ~ +0.039)
- Readout zero-shot Qwen3.8-27B (T1) – readout zero-shot 9B (T1): -0.357 (-0.382 ~ -0.332)
- Readout zero-shot Qwen3.5-27B (T1) – readout zero-shot 9B (T1): +0.015 (-0.006 ~ +0.038)

### S9 Per-seed paired differences

#### Main paired differences (quadratic-weighted κ; patient-level cluster bootstrap B = 1000; A−B)

| Data | Hyp. | Contrast | n | A−B | 95%CI | κ(A) | κ(B) |
|---|---|---|---|---|---|---|---|
| Local val | H2 | Readout T1 calibrated (1,000) − LoRA@1000 seed 20261017 | 2753 | -0.019 | -0.042 ~ +0.005 | 0.534 | 0.553 |
| Local val | H2 | Readout T1 calibrated (1,000) − LoRA@1000 seed 11 | 2753 | -0.028 | -0.051 ~ -0.003 | 0.534 | 0.562 |
| Local val | H2 | Readout T1 calibrated (1,000) − LoRA@1000 seed 12 | 2753 | -0.029 | -0.051 ~ -0.006 | 0.534 | 0.563 |
| Local val | H2 | Readout T1 calibrated (1,000) − LoRA@1000 three-seed mean | 2753 | -0.025 | -0.046 ~ -0.003 | 0.534 | 0.559 |
| Local val | H3 | LoRA@full seed 20261017 − Readout T1 calibrated (1,000) | 2753 | +0.024 | +0.001 ~ +0.047 | 0.558 | 0.534 |
| Local val | H3 | LoRA@full seed 11 − Readout T1 calibrated (1,000) | 2753 | +0.036 | +0.013 ~ +0.059 | 0.570 | 0.534 |
| Local val | H3 | LoRA@full seed 12 − Readout T1 calibrated (1,000) | 2753 | +0.022 | -0.001 ~ +0.045 | 0.556 | 0.534 |
| Local val | H3 | LoRA@full three-seed mean − Readout T1 calibrated (1,000) | 2753 | +0.027 | +0.006 ~ +0.048 | 0.561 | 0.534 |
| Local val | H2 | OpenJev T1 calibrated (1,000) − LoRA@1000 seed 20261017 | 2753 | -0.051 | -0.080 ~ -0.022 | 0.502 | 0.553 |
| Local val | H2 | OpenJev T1 calibrated (1,000) − LoRA@1000 seed 11 | 2753 | -0.060 | -0.087 ~ -0.031 | 0.502 | 0.562 |
| Local val | H2 | OpenJev T1 calibrated (1,000) − LoRA@1000 seed 12 | 2753 | -0.061 | -0.087 ~ -0.033 | 0.502 | 0.563 |
| Local val | H2 | OpenJev T1 calibrated (1,000) − LoRA@1000 three-seed mean | 2753 | -0.057 | -0.084 ~ -0.030 | 0.502 | 0.559 |
| Local val | H3 | LoRA@full seed 20261017 − OpenJev T1 calibrated (1,000) | 2753 | +0.056 | +0.030 ~ +0.082 | 0.558 | 0.502 |
| Local val | H3 | LoRA@full seed 11 − OpenJev T1 calibrated (1,000) | 2753 | +0.068 | +0.042 ~ +0.094 | 0.570 | 0.502 |
| Local val | H3 | LoRA@full seed 12 − OpenJev T1 calibrated (1,000) | 2753 | +0.054 | +0.028 ~ +0.078 | 0.556 | 0.502 |
| Local val | H3 | LoRA@full three-seed mean − OpenJev T1 calibrated (1,000) | 2753 | +0.059 | +0.035 ~ +0.082 | 0.561 | 0.502 |
| Local val | H2 | Readout T2 calibrated (1,000) − LoRA T2@1000(single seed) | 2753 | +0.009 | -0.015 ~ +0.032 | 0.545 | 0.536 |
| Local val | H3 | LoRA T2@full(single seed)− Readout T2 calibrated (1,000) | 2753 | +0.061 | +0.036 ~ +0.088 | 0.606 | 0.545 |
| Local val | H2 | OpenJev T2 calibrated (1,000) − LoRA T2@1000(single seed) | 2753 | +0.019 | -0.006 ~ +0.046 | 0.555 | 0.536 |
| Local val | H3 | LoRA T2@full(single seed)− OpenJev T2 calibrated (1,000) | 2753 | +0.051 | +0.025 ~ +0.077 | 0.606 | 0.555 |
| Local val | Input | Readout T2 calibrated − Readout T1 calibrated | 2753 | +0.011 | -0.016 ~ +0.039 | 0.545 | 0.534 |

| Data | Hyp. | Contrast | n | A−B | 95%CI | κ(A) | κ(B) |
|---|---|---|---|---|---|---|---|
| Local val | Input | LoRA T2@full − LoRA T1@full (seed 20261017) | 2753 | +0.048 | +0.021 ~ +0.075 | 0.606 | 0.558 |
| Local val (generative subset) | H1 | Readout T1 uncalibrated − generative zero-shot | 999 | +0.234 | +0.201 ~ +0.264 | 0.460 | 0.227 |
| Local val (generative subset) | H1 | OpenJev T1 calibrated − generative zero-shot | 999 | +0.298 | +0.263 ~ +0.330 | 0.524 | 0.227 |
| Local val (generative subset) | H1 | Readout T1 calibrated − generative zero-shot | 999 | +0.331 | +0.295 ~ +0.363 | 0.557 | 0.227 |
| MOVER subset | H2 | Readout T1 calibrated (1,000) − LoRA@1000 three-seed mean | 4000 | +0.019 | +0.003 ~ +0.037 | 0.524 | 0.505 |
| MOVER subset | H2 | Readout T1 calibrated (1,000) − LoRA@1000 seed 20261017 | 4000 | -0.003 | -0.025 ~ +0.018 | 0.524 | 0.527 |
| MOVER subset | H2 | Readout T1 calibrated (1,000) − LoRA@1000 seed 11 | 4000 | +0.062 | +0.040 ~ +0.084 | 0.524 | 0.462 |
| MOVER subset | H2 | Readout T1 calibrated (1,000) − LoRA@1000 seed 12 | 4000 | -0.001 | -0.020 ~ +0.019 | 0.524 | 0.525 |
| MOVER subset | H3 | LoRA@3000 three-seed mean − Readout T1 calibrated (1,000) | 4000 | +0.039 | +0.019 ~ +0.058 | 0.563 | 0.524 |
| MOVER subset | H3 | LoRA@12000 three-seed mean − Readout T1 calibrated (1,000) | 4000 | +0.059 | +0.040 ~ +0.080 | 0.583 | 0.524 |
| MOVER subset | H2 | OpenJev T1 calibrated (1,000) − LoRA@1000 three-seed mean | 4000 | -0.014 | -0.032 ~ +0.005 | 0.491 | 0.505 |
| MOVER subset | H2 | OpenJev T1 calibrated (1,000) − LoRA@1000 seed 20261017 | 4000 | -0.036 | -0.060 ~ -0.014 | 0.491 | 0.527 |
| MOVER subset | H2 | OpenJev T1 calibrated (1,000) − LoRA@1000 seed 11 | 4000 | +0.029 | +0.007 ~ +0.049 | 0.491 | 0.462 |
| MOVER subset | H2 | OpenJev T1 calibrated (1,000) − LoRA@1000 seed 12 | 4000 | -0.034 | -0.055 ~ -0.012 | 0.491 | 0.525 |
| MOVER subset | H3 | LoRA@3000 three-seed mean − OpenJev T1 calibrated (1,000) | 4000 | +0.072 | +0.050 ~ +0.094 | 0.563 | 0.491 |
| MOVER subset | H3 | LoRA@12000 three-seed mean − OpenJev T1 calibrated (1,000) | 4000 | +0.093 | +0.069 ~ +0.115 | 0.583 | 0.491 |
| MOVER subset (generative subset) | H1 | Readout T1 uncalibrated − generative zero-shot | 1978 | +0.224 | +0.199 ~ +0.247 | 0.473 | 0.249 |
| MOVER subset (generative subset) | H1 | OpenJev T1 calibrated − generative zero-shot | 1978 | +0.250 | +0.220 ~ +0.277 | 0.499 | 0.249 |
| VitalDB | H2 | Readout T1 calibrated (1,000) − target-site LoRA T1@1000(seed mean) | 1885 | +0.007 | -0.024 ~ +0.039 | 0.507 | 0.500 |
| VitalDB | H2 | Readout T1 calibrated (1,000) − target-site LoRA T1@1000 seed 20261017 | 1885 | +0.007 | -0.024 ~ +0.039 | 0.507 | 0.500 |
| VitalDB | H3 | target-site LoRA T1@3000(seed mean)− Readout T1 calibrated (1,000) | 1885 | +0.041 | +0.010 ~ +0.072 | 0.548 | 0.507 |
| VitalDB | H2 | OpenJev T1 calibrated (1,000) − target-site LoRA T1@1000(seed mean) | 1885 | +0.006 | -0.024 ~ +0.039 | 0.505 | 0.500 |
| VitalDB | H2 | OpenJev T1 calibrated (1,000) − target-site LoRA T1@1000 seed 20261017 | 1885 | +0.006 | -0.024 ~ +0.039 | 0.505 | 0.500 |

| Data | Hyp. | Contrast | n | A−B | 95%CI | κ(A) | κ(B) |
|---|---|---|---|---|---|---|---|
| VitalDB | H3 | target-site LoRA T1@3000(seed mean)− OpenJev T1 calibrated (1,000) | 1885 | +0.042 | +0.012 ~ +0.073 | 0.548 | 0.505 |
| VitalDB | H2 | Readout T2 calibrated (1,000) − target-site LoRA T2@1000(seed mean) | 1885 | -0.000 | -0.023 ~ +0.025 | 0.567 | 0.567 |
| VitalDB | H2 | Readout T2 calibrated (1,000) − target-site LoRA T2@1000 seed 20261017 | 1885 | +0.035 | +0.004 ~ +0.070 | 0.567 | 0.532 |
| VitalDB | H2 | Readout T2 calibrated (1,000) − target-site LoRA T2@1000 seed 11 | 1885 | -0.001 | -0.027 ~ +0.028 | 0.567 | 0.568 |
| VitalDB | H2 | Readout T2 calibrated (1,000) − target-site LoRA T2@1000 seed 12 | 1885 | -0.035 | -0.063 ~ -0.008 | 0.567 | 0.602 |
| VitalDB | H3 | target-site LoRA T2@3000(seed mean)− Readout T2 calibrated (1,000) | 1885 | +0.018 | -0.010 ~ +0.043 | 0.584 | 0.567 |
| VitalDB | H2 | OpenJev T2 calibrated (1,000) − target-site LoRA T2@1000(seed mean) | 1885 | +0.015 | -0.008 ~ +0.037 | 0.582 | 0.567 |
| VitalDB | H2 | OpenJev T2 calibrated (1,000) − target-site LoRA T2@1000 seed 20261017 | 1885 | +0.050 | +0.018 ~ +0.083 | 0.582 | 0.532 |
| VitalDB | H2 | OpenJev T2 calibrated (1,000) − target-site LoRA T2@1000 seed 11 | 1885 | +0.015 | -0.012 ~ +0.041 | 0.582 | 0.568 |
| VitalDB | H2 | OpenJev T2 calibrated (1,000) − target-site LoRA T2@1000 seed 12 | 1885 | -0.020 | -0.041 ~ +0.003 | 0.582 | 0.602 |
| VitalDB | H3 | target-site LoRA T2@3000(seed mean)− OpenJev T2 calibrated (1,000) | 1885 | +0.002 | -0.021 ~ +0.026 | 0.584 | 0.582 |

### S10 VitalDB T1 results

**VitalDB target-site replication (exploratory; test n = 1885, 1788 patients)**

#### S10.1 κ of each arm (against the ASA recorded in VitalDB anesthesia records)

| Arm | κ |
|---|---|
| In-house readout T1 uncalibrated | 0.357 |
| In-house readout T1 + 1000-label cutpoint calibration | 0.507 |
| In-house readout T1 + 100-label cutpoint calibration (mean of 60 runs) | 0.490 |
| Frozen OpenJev T1 uncalibrated | 0.366 |
| Frozen OpenJev T1 + 1000-label cutpoint calibration | 0.505 |
| Frozen OpenJev T1 + 100-label cutpoint calibration (mean of 60 runs) | 0.495 |
| In-house readout T2 uncalibrated | 0.380 |
| In-house readout T2 + 1000-label cutpoint calibration | 0.567 |
| In-house readout T2 + 100-label cutpoint calibration (mean of 60 runs) | 0.545 |
| Frozen OpenJev T2 uncalibrated | 0.370 |
| Frozen OpenJev T2 + 1000-label cutpoint calibration | 0.582 |
| Frozen OpenJev T2 + 100-label cutpoint calibration (mean of 60 runs) | 0.549 |

#### S10.2 Target-site LoRA (κ, seed mean [range])

| Input | n | κ | Seeds |
|---|---|---|---|
| T1 | 1000 | 0.500 [0.500–0.500] | 1 |
| T1 | 3000 | 0.548 [0.548–0.548] | 1 |
| T2 | 100 | 0.282 [0.061–0.444] | 3 |
| T2 | 300 | 0.470 [0.365–0.570] | 3 |
| T2 | 1000 | 0.567 [0.532–0.602] | 3 |
| T2 | 3000 | 0.584 [0.561–0.599] | 3 |

#### S10.3 Paired contrasts (seed 20261017; patient-level bootstrap)

- In-house readout T1 calibrated (1000) − target-site LoRA@1000 (non-inferiority δ = −0.05): +0.007 (-0.023 ~ +0.038)
- In-house readout T1 calibrated (1000) − target-site LoRA@3000: -0.041 (-0.072 ~ -0.011)
- Frozen OpenJev T1 calibrated (1000) − target-site LoRA@1000 (non-inferiority δ = −0.05): +0.006 (-0.025 ~ +0.039)
- Frozen OpenJev T1 calibrated (1000) − target-site LoRA@3000: -0.042 (-0.074 ~ -0.011)
- In-house readout T2 calibrated (1000) − target-site LoRA@1000 (non-inferiority δ = −0.05): +0.035 (+0.003 ~ +0.069)
- In-house readout T2 calibrated (1000) − target-site LoRA@3000: -0.032 (-0.060 ~ -0.005)
- Frozen OpenJev T2 calibrated (1000) − target-site LoRA@1000 (non-inferiority δ = −0.05): +0.050 (+0.017 ~ +0.085)
- Frozen OpenJev T2 calibrated (1000) − target-site LoRA@3000: -0.017 (-0.042 ~ +0.008)

**VitalDB target-site calibration (readout arm, n = 2000)**

Zero-shot κ = 0.334. If k local annotations were available in VitalDB, three cutpoints were fitted on them and evaluated on the remaining operations; 100 repetitions (seed 5).

| **k** | **Mean κ** | **2.5%** | **97.5%** |
|---|---|---|---|
| 50 | 0.469 | 0.406 | 0.505 |
| 100 | 0.485 | 0.444 | 0.510 |
| 200 | 0.490 | 0.455 | 0.511 |
| 500 | 0.497 | 0.479 | 0.513 |
| 1000 | 0.496 | 0.461 | 0.525 |

### S11 Reference verification record

**Item-by-item verification of references (night of 2026-10-20; Semantic Scholar API + PubMed E-utilities + Crossref + OpenAlex + Europe PMC full texts)**

Note: the "#" numbers below are those of the working reference file used at the time of verification, not the numbering of the final reference list; entries not retained in the final list are marked as such where known.

#### S11.1 Identifier level (DOI/PMID/arXiv)

- All 32 entries of the working reference file were resolved to records in Semantic Scholar (20 exact hits, 12 weak matches; all weak matches were due to abbreviated or short titles in the working file and were manually confirmed to be the same paper).
- 20 PMIDs were rechecked with PubMed efetch: title and first author agreed.
- Publication years follow Crossref/PubMed; **several years in the working file needed correction**: #5 is 2026 (J Med Internet Res 2026;28:e89540; Semantic Scholar records 2025 because of a difference in the online date), #6 is 2025;27:e75052, #27 is 2026 (BMC Med Inform Decis Mak, online 2026-09-25, no volume or issue yet), #28 is Appl Intell 2025;55(12):847, #32 is J Med Syst 2026;50(1):105. The canonical format is in the final reference list of the manuscript.

#### S11.2 Content level (numbers/claims in the working file versus the original)

| # | Source of verification | Conclusion |
|---|---|---|
| 1, 2, 4, 5, 7, 9–11, 13, 14, 16–19, 21–26, 30, 31 | API/PubMed abstracts | All numbers in the working file were found in the abstracts; no discrepancy |
| 3 | API (all three resolved) | The three earlier conventional-ML studies exist; the claim of a "registered medical-device software" came from a news report and is **neither in the reference list nor cited in the text** |
| 6 | Europe PMC open-access full text (PMC12174870) | Fully verified: mortality F1 0.4663; without RAG 0.0879; XGBoost 0.4459; ASA micro-F1 0.8409, without RAG 0.6340; 24,491 cases. **Addition**: the XGBoost ASA micro-F1 of that paper is 0.8273, only about 1.4 percentage points below LLaMA-RAG, so it must not be cited as "LLMs clearly outperform conventional methods" |
| 8 | PubMed abstract | Verified: knowledge graph 212,313 entities, 529,845 relations; average accuracy on the attending-physician qualification examination 7.3% higher than ChatGPT-4o (and 2.4% over ChatGPT-3.5-turbo, 3.7% over DeepSeek-V3). The "7.3%" is a **relative** improvement |
| 15 | PubMed abstract | Verified: LLaMA-3.1-8B; MIMIC-IV, UCSF adult/pediatric ICU; up to 16.6 percentage points above GPT-5; stable convergence over 10 virtual centers |

| # | Source of verification | Conclusion |
|---|---|---|
| 27 | OpenAlex abstract | Verified: AUC 0.8118 (PhysioNet/CinC 2019), 0.7780 (MIMIC-IV); 7B LLM + LoRA |
| 29 | PubMed abstract | Verified: MOVER 21,049 cases; GPT-4o few-shot + features accuracy 66%, F1 0.65, AUROC 0.79; LightGBM 54% |
| 12 | No abstract available | Title and journal verified; content claims not checked item by item, cited in the text only as an existence reference for anesthesia-specific LLMs |
| 28 | Crossref title only, no abstract/full text | **Content not verified.** Its methods or results are not cited in the text; it is cited, if at all, only at title level, or deleted |
| 32 | Crossref title only, no abstract/full text | **Content not verified** (commentary article). It may only be described as ”a commentary discussing validity problems of LLM-based ASA grading”, without restating its specific arguments |

#### S11.3 Handling rule

References in the text are taken from the canonical reference list; entries marked ”not verified” above are not restated in detail.

### S12 Characteristics of development versus evaluation splits

##### Characteristics of development and evaluation splits (ASA I / II / III / IV–V, %)

| Data source | Split | Operations | Age, median (IQR) | Female | BMI, median (IQR) | BMI missing | Preop creatinine available | ASA mix |
|---|---|---|---|---|---|---|---|---|
| MOVER | Development split (train80) | 45994 | 57 (41–68) | 47.9% | 27 (23–31) | 20.7% | 40% | 5.1 / 31.5 / 51.2 / 12.1 |
| MOVER | Test set (test20) | 11479 | 57 (40–68) | 47.3% | 27 (23–31) | 20.6% | 41% | 5.1 / 31.0 / 51.3 / 12.6 |
| MOVER | Evaluation subset (4,000) | 4000 | 56 (40–68) | 47.2% | 26 (23–31) | 20.2% | 42% | 4.9 / 30.7 / 51.8 / 12.7 |
| Local hospital | Development set 2021–24 | 4326 | 54 (46–65) | 66.8% | 25 (22–28) | 0.6% | 99% | 1.4 / 45.4 / 50.7 / 2.5 |
| Local hospital | Validation set 2025–26 | 2753 | 53 (45–63) | 71.4% | 25 (23–28) | 0.2% | 97% | 1.3 / 47.0 / 48.8 / 2.9 |
| VitalDB | Training set 70% | 4357 | 59 (49–68) | 48.4% | 23 (21–25) | 0.0% | 96% | 28.9 / 59.4 / 11.1 / 0.6 |
| VitalDB | Test set 30% | 1885 | 59 (47–68) | 51.6% | 23 (21–25) | 0.0% | 96% | 28.3 / 58.8 / 11.7 / 1.2 |

### S13 Per-condition results of natural-language guidance

**Natural-language guidance experiment (readout arm, no training, no labels)**

Local validation set n = 2753; original-record distribution I/II/III/IV–V = [1.3, 47.0, 48.8, 2.9]%. The guidance wording was fixed before results were seen and not modified.

| Input | Guidance | κ (against local records) | Predicted distribution I/II/III/IV–V (%) | κ after +100-label cutpoint calibration | dev800 κ (direction check) |
|---|---|---|---|---|---|
| T1 | No guidance | 0.443 | [11.1, 59.1, 27.4, 2.4] | 0.516 | 0.436 |
| T1 | ASA 2020 official definitions | 0.312 | [43.1, 35.7, 20.2, 1.1] | 0.487 | 0.306 |
| T1 | Local recording habit | 0.272 | [0.0, 85.2, 14.6, 0.2] | 0.522 | 0.262 |
| T1 | Strict guideline | 0.187 | [64.8, 24.2, 10.5, 0.6] | 0.469 | 0.162 |
| T2 | No guidance | 0.505 | [10.4, 20.1, 59.5, 10.0] | 0.531 | 0.571 |
| T2 | ASA 2020 official definitions | 0.435 | [8.7, 13.0, 70.4, 7.9] | 0.531 | 0.516 |
| T2 | Local recording habit | 0.557 | [0.0, 45.3, 50.6, 4.1] | 0.532 | 0.591 |
| T2 | Strict guideline | 0.528 | [18.4, 35.2, 38.9, 7.4] | 0.550 | 0.550 |

#### S13.1 Paired differences against the comparison condition (κ, patient-level bootstrap; against local records)

- T1: ASA 2020 official definitions – no guidance = -0.131 (-0.150 ~ -0.113)
- T1: local recording habit – no guidance = -0.171 (-0.195 ~ -0.146)
- T1: strict guideline – no guidance = -0.256 (-0.275 ~ -0.237)
- T1: local recording habit – strict guideline = +0.085 (+0.064 ~ +0.105) (between the two guidance conditions of opposite direction)
- T2: ASA 2020 official definitions – no guidance = -0.070 (-0.087 ~ -0.052)
- T2: local recording habit – no guidance = +0.052 (+0.028 ~ +0.077)
- T2: strict guideline – no guidance = +0.023 (+0.002 ~ +0.044)
- T2: local recording habit – strict guideline = +0.030 (+0.006 ~ +0.051) (between the two guidance conditions of opposite direction)

For comparison (local validation set, against the original record): readout zero-shot T1 0.443 / T2 0.505; readout + 1000-label calibration T1 0.534; local full-set LoRA T1 0.561 / T2 0.606. Interpretation rule (fixed in advance): if GLOC moves the predicted distribution toward the original record (fewer I, more III) and κ rises, natural language can replace part of the labels; if the distribution shifts but κ does not rise, steerability holds but does not mean greater accuracy; if the distribution does not move either, the claim that natural-language guidance is usable is not supported.

**Natural-language guidance: additional paired differences (local validation set, n = 2753)**

- T1: GLOC (local recording habit, no labels) – unguided readout + 1000-label calibration = -0.262 (-0.292 ~ -0.235)
- T1: GLOC – local full-set LoRA = -0.286 (-0.319 ~ -0.256)
- T2: GLOC (local recording habit, no labels) – unguided readout + 1000-label calibration = +0.012 (-0.011 ~ +0.036)
- T2: GLOC – local full-set LoRA = -0.049 (-0.075 ~ -0.022)
- T2: GLOC – local LoRA T2@1000 (single seed) = +0.021 (-0.005 ~ +0.047)
- T2: GSTR – unguided readout + 1000-label calibration = -0.017 (-0.037 ~ +0.004)

#### S14 Token consumption details

**Token consumption of the same base model (Qwen3.5-9B tokenizer)**

| Data | Cases | Input tokens per case, readout | Input tokens per case, generative |
|---|---|---|---|
| Local val (T1) | 1000 | 171 | 187 |
| MOVER subset (T1) | 1000 | 161 | 177 |

Generative output tokens (measured on the first 96 operations of the local generative subset, thinking mode off, cap 300; MOVER output tokens not re-measured):

| Mode | Operations | Mean | Median | 10th–90th percentile | Reached cap | Contains ”Final answer” |
|---|---|---|---|---|---|---|
| Thinking off (used in this study) | 96 | 89 | 87 | 73–104 | 0.0% | 100.0% |

Local validation set as an example (readout 171 input tokens; generative 187 input tokens + 89 output tokens on average):

| Item | Readout | Generative zero-shot | LoRA (inference) |
|---|---|---|---|
| Tokens processed per case (input + output) | 171 + 0 | 187 + 89 | 171 + 0 |
| Sequential decoding steps per case | 0 (one forward pass, logits of the last position for 4 options) | about 89 | 0 |
| Tokens processed per 10,000 cases | 1.71 M | 2.76 M (0.89 M of them token-by-token generation) | 1.71 M |
| Sequential decoding steps per 10,000 cases | 0 | 0.89 M | 0 |

One-time label-related token consumption (local T1, 171 tokens per case; training for 1 epoch needs forward and backward passes):

| Item | Tokens processed | Note |
|---|---|---|
| Readout + 100-label calibration | 0.017 M (forward only) | Scores 100 cases; three cutpoints fitted on a CPU; no model parameter updated |
| Readout + 1000-label calibration | 0.171 M (forward only) | Scores 1000 cases; three cutpoints fitted on a CPU; no model parameter updated |
| LoRA training n = 300 | 0.051 M (forward + backward + gradient checkpointing) | Updates adapter parameters; trained once per site and the adapter stored |
| LoRA training n = 1000 | 0.171 M (forward + backward + gradient checkpointing) | Updates adapter parameters; trained once per site and the adapter stored |
| LoRA training n = 4326 | 0.738 M (forward + backward + gradient checkpointing) | Updates adapter parameters; trained once per site and the adapter stored |

### S15 Adjacent-class error details

##### Adjacent-class errors (accuracy / adjacent accuracy |difference| ≤ 1 / proportion of errors of 2 or more classes)

| Data | Arm | n | Accuracy | Adjacent accuracy | Error ≥ 2 classes |
|---|---|---|---|---|---|
| Local val | Readout, zero-shot | 2,753 | 0.563 | 0.984 | 0.016 |
| Local val | Readout + 1,000-label calibration | 2,753 | 0.666 | 0.991 | 0.009 |
| Local val | LoRA n=1000 (seed mean) | 2,753 | 0.746 | 0.999 | 0.001 |
| Local val | LoRA n=4326 (seed mean) | 2,753 | 0.747 | 0.998 | 0.002 |
| MOVER subset | Readout, zero-shot | 4,000 | 0.464 | 0.945 | 0.055 |
| MOVER subset | Readout + 1,000-label calibration | 4,000 | 0.515 | 0.967 | 0.034 |
| MOVER subset | LoRA n=1000 (seed mean) | 4,000 | 0.600 | 0.987 | 0.013 |
| MOVER subset | LoRA n=3000 (seed mean) | 4,000 | 0.636 | 0.991 | 0.009 |
| MOVER subset | LoRA n=12000 (seed 20261017) | 4,000 | 0.652 | 0.992 | 0.008 |

#### S16 SHA-256 checksums of the Qwen3.5-9B weight files

Below are the SHA-256 checksums of the files of the Hugging Face weight repository Qwen/Qwen3.5-9B (post-trained version, Apache-2.0) after download on 2026-10-16. Each entry gives the file name first and then the checksum.

```
.gitattributes
f5448ec7bb0a52de6c6fdba5ba48fe01bf216d546515a4ef9d5053e252b55359

LICENSE
50cbab8a892c5f2993b8c7351a99182507472def3b1374558308605d99b86b32

README.md
c5f5a8c2dddab69cfbf05279235aa5fddb137939a06539c4c7637aa900fef6d0

chat_template.jinja
a4aee8afcf2e0711942cf848899be66016f8d14a889ff9ede07bca099c28f715

config.json
d0883072e01861ed0b2d47be3c16c36a8e81c224c7ffaa310c6558fb3f932b05

configuration.json
2d4464e2ead06bc9bc718c781309ad1e7baded626d66e8dcdc8b469ba185faf0

merges.txt
a9d356d7bdf1ef4949e3e748e95b8e10ad9d4e2e838eddc38a0a7b6b94d1db8d

model.safetensors-00001-of-00004.safetensors
db6f444b43d318c92f360a13a25561a6a65b10c0631b8ed305a426dbaa6c380e

model.safetensors-00002-of-00004.safetensors
31c7d7e2dd5d207840b31cc59083c8f4c4718959149e0358c0364052bb9a0330

model.safetensors-00003-of-00004.safetensors
7ec36ba3a4176a44c3c0876ad80c56a2f70c84bf008d82e9501df642f17dadec

model.safetensors-00004-of-00004.safetensors
b62b0c4cd7e44edee103ee8f4fe225f246d5e768e07bfd5f25b63a8aa1fdd0c6

model.safetensors.index.json
26d3539b516be613f39563617cb9d33b3f83d401298125be392c80cefb8f7fe5

preprocessor_config.json
27225450ac9c6529872ee1924fcb0962ff5634834f817040f444118116f4e516

tokenizer.json
5f9e4d4901a92b997e463c1f46055088b6cca5ca61a6522d1b9f64c4bb81cb42

tokenizer_config.json
316230d6a809701f4db5ea8f8fc862bc3a6f3229c937c174e674ff3ca0a64ac8

video_preprocessor_config.json
7768af27c1fafa9cc9011c1dc20067e03f8915e03b63504550e11d5066986d13

vocab.json
ce99b4cb2983d118806ce0a8b777a35b093e2000a503ebde25853284c9dfa003
```

### S17 Local validation subgroups (exploratory)

##### Local validation set subgroups (exploratory; κ and accuracy, 95% CI by patient-level bootstrap)

| Subgroup | n | ASA I/II/III/IV–V (%) | Readout zero-shot κ | Calibrated readout κ | LoRA@1,000 κ (three-seed mean) | Calibrated readout accuracy | LoRA accuracy |
|---|---|---|---|---|---|---|---|
| All | 2,753 | 1.3 / 47.0 / 48.8 / 2.9 | 0.443 (0.417–0.467) | 0.534 (0.505–0.559) | 0.559 (0.533–0.584) | 0.666 (0.648–0.683) | 0.746 (0.729–0.761) |
| Female | 1,966 | 1.8 / 55.6 / 40.8 / 1.8 | 0.424 (0.391–0.453) | 0.526 (0.493–0.559) | 0.542 (0.509–0.574) | 0.676 (0.656–0.696) | 0.751 (0.733–0.769) |
| Male | 787 | 0.1 / 25.5 / 68.7 / 5.6 | 0.350 (0.297–0.401) | 0.413 (0.353–0.468) | 0.410 (0.349–0.465) | 0.640 (0.605–0.675) | 0.733 (0.704–0.761) |
| Age <65 | 2,129 | 1.7 / 57.2 / 40.2 / 0.9 | 0.366 (0.337–0.396) | 0.484 (0.453–0.515) | 0.480 (0.445–0.512) | 0.678 (0.659–0.697) | 0.734 (0.717–0.751) |
| Age ≥65 | 624 | 0.0 / 12.3 / 78.2 / 9.5 | 0.265 (0.200–0.327) | 0.219 (0.141–0.291) | 0.065 (0.032–0.100) | 0.625 (0.588–0.662) | 0.786 (0.753–0.817) |

Note: in the group aged 65 or older, ASA III accounts for 78.2% and there is no class I; the within-group κ is limited by the range of values and should not be interpreted, and accuracy should be used instead.

### S18 Label-pool matching, label-sampling sensitivity and non-inferiority margin sensitivity

**Supplementary analyses after review (CPU part)**

#### S18.1 Label-pool matching (local hospital)

- The 1,000 training labels of LoRA seed 11 coincide with the calibration pool in 1000/1000 cases
- The 1,000 training labels of LoRA seed 12 coincide with the calibration pool in 238/1000 cases
- The 1,000 training labels of LoRA seed 20261017 coincide with the calibration pool in 223/1000 cases

#### S18.2 Label-sampling sensitivity (local hospital; 50 independent 1,000-label pools drawn from the 4,326 development operations, three cutpoints fitted in each and evaluated on the 2,753 validation operations)

- Calibrated readout κ: mean 0.537, label-sampling SD 0.006, range 0.514–0.546; mean accuracy 0.693
- κ of LoRA@1,000, three seeds: 0.553, 0.562, 0.563 (mean 0.559, SD 0.005); mean accuracy 0.746
- Note: the development set has only 4,326 operations and the 50 pools overlap, so the SD underestimates the true between-pool variation; LoRA has only 3 training pools.
- **H2 difference including label sampling and seed variation** (each resample: patient clusters + one random calibration pool + one random LoRA seed, B = 2000): mean -0.022, 95% interval -0.052 ~ +0.006; lower bound > −0.05: no

#### S18.3 Sensitivity to the non-inferiority margin δ (H2, lower bound of the 95% interval of the difference > −δ)

| Data | Contrast | Lower bound | δ = 0.03 | δ = 0.05 | δ = 0.10 |
|---|---|---|---|---|---|
| Local val | Readout T1 calibrated (1,000) – LoRA@1000 seed 20261017 | -0.042 | no | yes | yes |
| Local val | Readout T1 calibrated (1,000) – LoRA@1000 seed 11 | -0.051 | no | no | yes |
| Local val | Readout T1 calibrated (1,000) – LoRA@1000 seed 12 | -0.051 | no | no | yes |
| Local val | Readout T1 calibrated (1,000) – LoRA@1000 three-seed mean | -0.046 | no | yes | yes |
| Local val | OpenJev T1 calibrated (1,000) – LoRA@1000 seed 20261017 | -0.080 | no | no | yes |
| Local val | OpenJev T1 calibrated (1,000) – LoRA@1000 seed 11 | -0.087 | no | no | yes |
| Local val | OpenJev T1 calibrated (1,000) – LoRA@1000 seed 12 | -0.087 | no | no | yes |
| Local val | OpenJev T1 calibrated (1,000) – LoRA@1000 three-seed mean | -0.084 | no | no | yes |
| Local val | Readout T2 calibrated (1,000) – LoRA T2@1000(single seed) | -0.015 | yes | yes | yes |
| Local val | OpenJev T2 calibrated (1,000) – LoRA T2@1000(single seed) | -0.006 | yes | yes | yes |
| MOVER subset | Readout T1 calibrated (1,000) – LoRA@1000 three-seed mean | +0.003 | yes | yes | yes |
| MOVER subset | Readout T1 calibrated (1,000) – LoRA@1000 seed 20261017 | -0.025 | yes | yes | yes |
| MOVER subset | Readout T1 calibrated (1,000) – LoRA@1000 seed 11 | +0.040 | yes | yes | yes |
| MOVER subset | Readout T1 calibrated (1,000) – LoRA@1000 seed 12 | -0.020 | yes | yes | yes |
| MOVER subset | OpenJev T1 calibrated (1,000) – LoRA@1000 three-seed mean | -0.032 | no | yes | yes |

| Data | Contrast | Lower bound | δ = 0.03 | δ = 0.05 | δ = 0.10 |
|---|---|---|---|---|---|
| MOVER subset | OpenJev T1 calibrated (1,000) – LoRA@1000 seed 20261017 | -0.060 | no | no | yes |
| MOVER subset | OpenJev T1 calibrated (1,000) – LoRA@1000 seed 11 | +0.007 | yes | yes | yes |
| MOVER subset | OpenJev T1 calibrated (1,000) – LoRA@1000 seed 12 | -0.055 | no | no | yes |
| VitalDB | Readout T1 calibrated (1,000) – target-site LoRA T1@1000(seed mean) | -0.024 | yes | yes | yes |
| VitalDB | Readout T1 calibrated (1,000) – target-site LoRA T1@1000 seed 20261017 | -0.024 | yes | yes | yes |
| VitalDB | OpenJev T1 calibrated (1,000) – target-site LoRA T1@1000(seed mean) | -0.024 | yes | yes | yes |
| VitalDB | OpenJev T1 calibrated (1,000) – target-site LoRA T1@1000 seed 20261017 | -0.024 | yes | yes | yes |
| VitalDB | Readout T2 calibrated (1,000) – target-site LoRA T2@1000(seed mean) | -0.023 | yes | yes | yes |
| VitalDB | Readout T2 calibrated (1,000) – target-site LoRA T2@1000 seed 20261017 | +0.004 | yes | yes | yes |
| VitalDB | Readout T2 calibrated (1,000) – target-site LoRA T2@1000 seed 11 | -0.027 | yes | yes | yes |
| VitalDB | Readout T2 calibrated (1,000) – target-site LoRA T2@1000 seed 12 | -0.063 | no | no | yes |
| VitalDB | OpenJev T2 calibrated (1,000) – target-site LoRA T2@1000(seed mean) | -0.008 | yes | yes | yes |
| VitalDB | OpenJev T2 calibrated (1,000) – target-site LoRA T2@1000 seed 20261017 | +0.018 | yes | yes | yes |
| VitalDB | OpenJev T2 calibrated (1,000) – target-site LoRA T2@1000 seed 11 | -0.012 | yes | yes | yes |
| VitalDB | OpenJev T2 calibrated (1,000) – target-site LoRA T2@1000 seed 12 | -0.041 | no | yes | yes |

#### S18.4 Crossover of calibrated readout and LoRA (linear interpolation in log n; the calibration curve uses subsets of the same 1,000-label pool)

- Local κ: per-seed crossovers [259, 264, 318] (calibration curve extrapolated as a plateau for $k > 1{,}000$); crossover of the mean curve 269
- Local accuracy: calibrated readout (1,000) 0.666; LoRA n = 100/300/1000/4326 (three-seed mean): 0.677, 0.737, 0.746, 0.747. LoRA accuracy is already higher than that of the calibrated readout at n = 100, so the accuracy crossover is below 100 labels.

#### S18.5 MOVER label-sampling sensitivity (a further 5,000 operations from the training split, 50 independent 1,000-label pools; evaluation subset n = 4,000)

- Calibrated readout κ: mean 0.525, between-pool SD 0.004, range 0.513–0.530; mean accuracy 0.514
- κ of LoRA@1,000, three seeds: 0.527, 0.462, 0.525 (mean 0.505)
- H2 difference including label sampling and seed variation: +0.020 (-0.020 ~ +0.080); lower bound > −0.05: yes

#### S19 Crossover of calibrated readout and LoRA (break-even)

##### Break-even (calibrated readout vs LoRA)

The calibration curve is taken from subsets of the same 1,000-label pool (mean of 100 resamples per k); for k > 1,000 it is extrapolated at the level of k = 1,000; LoRA is taken per seed. Crossovers are by linear interpolation in log n.

- **Local hospital ($\kappa$)**: per-seed crossover seed 20261017: 259; seed 11: 264; seed 12: 318; mean curve 269
- Local LoRA accuracy (three-seed mean) at n = [100, 300, 1000, 4326]: [0.677, 0.737, 0.746, 0.747]; calibrated readout (pool 11) 0.666, (mean of 50 pools) 0.693.
- **MOVER ($\kappa$)**: per-seed crossover seed 20261017: 986; seed 11: 1832; seed 12: 958 (calibration curve extrapolated beyond k > 1,000)
- MOVER LoRA accuracy (three-seed mean) at n = [100, 300, 1000, 3000, 12000]: [0.577, 0.562, 0.6, 0.636, 0.65]; calibrated readout (pool 11) 0.515.
- **VitalDB T2 ($\kappa$)**: per-seed crossover seed 12: 268; seed 20261017: 1775; seed 11: 996
- VitalDB T2 LoRA accuracy (three-seed mean) at n = [100, 300, 1000, 3000]: [0.632, 0.664, 0.683, 0.699]

### S20 Option-order permutation averaging and the Chinese T1 template

**Option-order permutation averaging and the Chinese template (review revision)**

#### S20.1 Local validation set (T1 English) (evaluation n = 2,753, calibration pool 1,000)

| Order | Uncalibrated κ | Calibrated κ | Macro AUC | Agreement with original-order predictions |
|---|---|---|---|---|
| orig | 0.443 | 0.534 | 0.794 | 1.000 |
| rev | 0.258 | 0.529 | 0.735 | 0.557 |
| s1 | 0.331 | 0.530 | 0.746 | 0.641 |
| s2 | 0.142 | 0.518 | 0.751 | 0.661 |
| s3 | 0.493 | 0.528 | 0.785 | 0.602 |
| r5 | 0.505 | 0.528 | 0.808 | 0.745 |
| **Average of probabilities over 6 orders** | **0.396** | **0.531** | 0.785 | — |

- Uncalibrated κ of single orders ranges over 0.142–0.505 (mean 0.362); after calibration 0.518–0.534 (mean 0.528).
- ECE of the averaged probability: 0.024; original order: 0.030.
- **H2 (order-averaged calibrated readout – LoRA@1,000 three-seed mean)**: -0.028 (-0.051 ~ -0.006); reference (original order): -0.025 (-0.048 ~ -0.004)

#### S20.2 MOVER evaluation subset (T1 English) (evaluation n = 4,000, calibration pool 1,000)

| Order | Uncalibrated κ | Calibrated κ | Macro AUC | Agreement with original-order predictions |
|---|---|---|---|---|
| orig | 0.473 | 0.524 | 0.762 | 1.000 |
| rev | 0.245 | 0.488 | 0.694 | 0.484 |
| s1 | 0.315 | 0.517 | 0.719 | 0.580 |
| s2 | 0.222 | 0.481 | 0.727 | 0.640 |
| s3 | 0.449 | 0.511 | 0.750 | 0.564 |
| r5 | 0.435 | 0.517 | 0.760 | 0.531 |
| **Average of probabilities over 6 orders** | **0.414** | **0.523** | 0.745 | — |

- Uncalibrated κ of single orders ranges over 0.222–0.473 (mean 0.356); after calibration 0.481–0.524 (mean 0.506).
- ECE of the averaged probability: 0.091; original order: 0.092.
- **H2 (order-averaged calibrated readout – LoRA@1,000 three-seed mean)**: +0.018 (-0.000 ~ +0.035); reference (original order): +0.019 (+0.002 ~ +0.037)

#### S20.3 Chinese T1 template (local validation set, n = 2,753; calibration pool 1,000, same pool 11)

| Condition | Uncalibrated κ | Calibrated κ | Macro AUC |
|---|---|---|---|
| T1 English template + English prompt shell | 0.443 | 0.534 | 0.794 |
| T2 raw Chinese visit note + English prompt shell | 0.505 | 0.545 | 0.833 |
| T1 Chinese template + English prompt shell | 0.462 | 0.516 | 0.788 |

| Condition | Uncalibrated κ | Calibrated κ | Macro AUC |
|---|---|---|---|
| T1 Chinese template + Chinese prompt shell | 0.512 | 0.530 | 0.821 |

#### S21 Latency decomposition into quantization and adapter

The readout ran in fp16; readout4 is the 4-bit quantized readout (no adapter); lora is the 4-bit model plus adapter. The difference between the last two reflects the adapter overhead, and the difference from fp16 reflects the quantization overhead. gen is generative zero-shot.

| **Arm** | **Description** | **p50** | **p95** | **Batch-8 throughput** | **Peak memory** |
|---|---|---|---|---|---|
| readout | — | 111ms | 118ms | 11.9 cases/s | 18.8GB |
| lora | curve3000 | 179ms | 195ms | 9.7 cases/s | 8.7GB |
| gen | — | 7067ms | 9105ms | 0.8 cases/s | 18.9GB |
| readout4 | — | 156ms | 158ms | 12.0 cases/s | 8.6GB |
| lora | curve1000 | 179ms | 182ms | 10.3 cases/s | 8.7GB |
| readout | — | 114ms | 117ms | 12.8 cases/s | 18.8GB |

### S22 fp16 LoRA run across two cards

**fp16 LoRA run across two cards (local validation set, n = 2,753)**

| Arm | κ (95% CI) | Accuracy | Macro AUC |
|---|---|---|---|
| fp16 LoRA (two cards, n = 1,000, seed 11) | 0.553 (0.525–0.580) | 0.741 | 0.845 |
| 4-bit LoRA (one card, same 1,000 labels) | 0.562 (0.534–0.588) | 0.747 | 0.846 |
| fp16 readout + 1,000-label calibration (same pool) | 0.534 (0.508–0.559) | 0.666 | — |

- fp16 LoRA − 4-bit LoRA (same labels): -0.008 (-0.021 ~ +0.006)
- **H2 (label-matched, fp16 LoRA)**: fp16 readout calibrated − fp16 LoRA = -0.019 (-0.045 ~ +0.006)
- Training 324 s (62 steps, layers split across the two cards); inference on the full validation set 246 s; batch-1 latency p50 160 ms, p95 163 ms (including inter-card transfer, adapter not merged); peak training memory 10.4/9.5 GB (two cards).

### S23 Accuracy of the 4-bit quantized readout

##### 4-bit (NF4) quantized readout versus fp16 readout (zero-shot, natural order, English T1)

| Data | Precision | Uncalibrated κ | κ calibrated on 1,000 labels | Macro AUC | Agreement with fp16 predictions |
|---|---|---|---|---|---|
| Local val | fp16 | 0.443 | 0.534 | 0.794 | — |
| Local val | 4-bit | 0.346 | 0.514 | 0.739 | 0.588 |
| | 4-bit − fp16 | -0.097 (-0.114 ~ -0.081) | -0.020 (-0.033 ~ -0.005) | | |
| MOVER subset | fp16 | 0.473 | 0.524 | 0.762 | — |
| MOVER subset | 4-bit | 0.378 | 0.521 | 0.733 | 0.622 |
| | 4-bit − fp16 | -0.095 (-0.112 ~ -0.080) | -0.003 (-0.015 ~ +0.009) | | |

#### S23.1 H2 with matched precision (readout and LoRA both in 4-bit)

| Data | Contrast (calibrated readout − LoRA@1,000, both 4-bit) | Difference (95% CI) |
|---|---|---|
| Local val | Seed 11 (label-matched) | -0.047 (-0.073 ~ -0.023) |
| Local val | Three-seed mean | -0.045 (-0.069 ~ -0.022) |
| MOVER subset | Three-seed mean | +0.016 (-0.002 ~ +0.034) |

For comparison: the label-matched contrast with both in fp16 (fp16 readout calibrated − fp16 LoRA, local hospital, seed 11) is −0.019 (−0.045 ~ +0.006); see the fp16 LoRA run across two cards.

### S24 Published agreement ($\kappa$) for ASA-PS grading

##### Published agreement (κ) for ASA-PS grading: between physicians, physician versus reference, AI versus physicians

Search and verification: PubMed E-utilities (2026-10-20); values were taken one by one from the original PubMed abstracts. Studies differ in population, rating method, κ weighting and prevalence; the table serves as background and cannot be converted directly.

| Study (PMID) | Design and population | Type of κ | Reported value |
|---|---|---|---|
| Di Maio 2026 (42649825) | 101 orthopedic patients, two clinicians rating independently | Quadratic-weighted κ, between physicians | 0.713 |
| Minerva Anestesiol 2026 meta-analysis (42189086) | 13 studies, 402,336 cases, AI versus clinicians | Quadratic-weighted κ, pooled | 0.69 (95% CI 0.62–0.76) |
| Kuza 2017 (28891913) | 8 fictitious polytrauma cases, 349 anesthesiologists and trauma surgeons; subset of 201 | Weighted κ: versus reference; between raters | Versus reference 0.63 (0.594–0.666); between raters 0.43 (0.360–0.491) |
| Abouleish 2020 (33292640) | Agreement of ASA between preoperative screening and day of surgery, before and after an intervention | Weighted κ | Before 0.62 (adults), 0.48 (children); after 0.85 (adults 0.86), 0.78 (children) |
| Liu 2026 (41503664) | Pediatric-specific ASA-PS, two independent anesthesiologists and the case anesthesiologist | Weighted κ | Between the two independent raters 0.76; case anesthesiologist versus independent raters 0.5 |
| Shichino 2017 (29123855) | ASA rating in emergency gastrointestinal surgery | Fleiss κ (and ICC) | κ 0.55 (0.54–0.56); ICC 0.79 |
| El youzouri 2026 (41634571) | Prospective observational, 684 cases, anesthesiologists and surgeons, reference a multidisciplinary panel | Cohen κ | Anesthesiologist versus reference 0.36; surgeon versus reference 0.21; between the two professions 0.25 |
| Clinical agreement study 2018 (29946447) | Retrospective, 101 arthroplasty patients, raters in the medical clinic, the preanesthesia clinic and on the day of surgery | κ | 0.170 and 0.156 (between providers of different departments) |
| Pediatric oncology 2018 (29441411) | 1,177 pediatric oncology anesthesia records | κ | Between different anesthesiologists −0.042 (−0.17 to 0.09); same physician before and after 0.48 |

**How to read it**: κ between physicians or between physicians and a reference ranges from about 0 to 0.85. Weighted κ between trained anesthesiologists or under favorable conditions is 0.62–0.85, κ across specialties, time points or against a reference panel is −0.04–0.43, and Fleiss κ is 0.55. The pooled quadratic-weighted κ of AI against clinicians is 0.69.

### S25 Comparability of the H1 subset and paired accuracy differences

**Comparability of the H1 subset and paired accuracy differences**

#### S25.1 Local validation set

| Group | n | Median age | Female | ASA I/II/III/IV–V (%) | Readout κ (uncalibrated) |
|---|---|---|---|---|---|
| H1 subset (random 1,000) | 1000 | 53 | 72.3% | 0.8 / 49.0 / 46.8 / 3.4 | 0.461 |
| Not included | 1753 | 54 | 70.9% | 1.6 / 45.9 / 49.9 / 2.6 | 0.432 |
| Whole validation set | 2753 | 53 | 71.4% | 1.3 / 47.0 / 48.8 / 2.9 | 0.443 |

#### S25.2 MOVER evaluation subset

| Group | n | Median age | Female | ASA I/II/III/IV–V (%) | Readout κ (uncalibrated) |
|---|---|---|---|---|---|
| H1 subset (first 2,000) | 2000 | 57 | 48.2% | 4.2 / 30.9 / 51.3 / 13.6 | 0.476 |
| Not included (last 2,000) | 2000 | 56 | 46.2% | 5.6 / 30.4 / 52.2 / 11.8 | 0.470 |
| Evaluation subset 4,000 | 4000 | 56 | 47.2% | 4.9 / 30.7 / 51.8 / 12.7 | 0.473 |

#### S25.3 Accuracy: LoRA (three-seed mean) – calibrated readout with 1,000 labels (pool 11); 95% CI by patient-level bootstrap

| Data | LoRA n | LoRA accuracy | Calibrated readout accuracy | Difference (95% CI) |
|---|---|---|---|---|
| Local hospital | 100 | 0.677 | 0.666 | +0.011 (-0.006 ~ +0.029) |
| Local hospital | 300 | 0.737 | 0.666 | +0.071 (+0.056 ~ +0.086) |
| Local hospital | 1000 | 0.746 | 0.666 | +0.080 (+0.065 ~ +0.095) |
| MOVER subset | 100 | 0.577 | 0.515 | +0.062 (+0.045 ~ +0.083) |
| MOVER subset | 300 | 0.562 | 0.515 | +0.047 (+0.033 ~ +0.064) |
| MOVER subset | 1000 | 0.600 | 0.515 | +0.085 (+0.068 ~ +0.102) |
