## Supplementary material for "Non-generative readout with few-label calibration versus LoRA fine-tuning of a large language model for ASA physical status classification: accuracy, speed, cost and label efficiency in three data sources": Analysis code and aggregate results: 00_smoke.py

```
"""Smoke test on SYNTHETIC Chinese notes (no patient data): (1) logit readout of ASA options,
(2) one QLoRA forward/backward step on SM75. Usage: 00_smoke.py <model_dir> [--train]"""
import sys, time, torch
from transformers import AutoTokenizer, AutoModelForCausalLM, BitsAndBytesConfig
path = sys.argv[1]; train = "--train" in sys.argv
CASES = [  # synthetic, label = ASA class
 ("女，34岁，体健，无既往史，拟行腹腔镜胆囊切除术。", 1),
 ("男，26岁，既往体健，吸烟0.5包/日，拟行阑尾切除术。", 1),
 ("男，62岁，高血压10年，口服氨氯地平控制良好，拟行结肠癌根治术。", 2),
 ("女，55岁，2型糖尿病5年，血糖控制尚可，BMI 31，拟行子宫全切术。", 2),
 ("男，71岁，冠心病支架术后2年，高血压，糖尿病，活动耐量尚可，拟行胃癌根治术。", 3),
 ("女，68岁，COPD，FEV1占预计值55%，长期吸入支气管扩张剂，拟行肺叶切除术。", 3),
 ("男，78岁，冠心病，3个月内发生心肌梗死，心功能NYHA III级，肾功能不全，拟行肠梗阻手术。", 4),
 ("女，80岁，重度主动脉瓣狭窄伴心衰，持续吸氧，拟行髋部骨折手术。", 4),
]
OPT = "ABCDE"
def prompt(t):
    return ("你是麻醉科医师。请根据术前信息判断ASA分级。\n病历：%s\n选项：A. ASA I  B. ASA II  C. ASA III  D. ASA IV  E. ASA V\n答案：" % t)
tok = AutoTokenizer.from_pretrained(path)
kw = dict(dtype=torch.float16, device_map={"": 0})
if train: kw["quantization_config"] = BitsAndBytesConfig(load_in_4bit=True, bnb_4bit_compute_dtype=torch.float16, bnb_4bit_quant_type="nf4")
t0 = time.time(); m = AutoModelForCausalLM.from_pretrained(path, **kw).eval(); print("load s", round(time.time()-t0,1), "mem GB", round(torch.cuda.memory_allocated()/1e9,1))
ids = [tok.encode(" "+c, add_special_tokens=False)[-1] for c in OPT]; ids2 = [tok.encode(c, add_special_tokens=False)[-1] for c in OPT]
ok = 0
for t, y in CASES:
    x = tok(prompt(t), return_tensors="pt").to(0)
    with torch.no_grad(): lg = m(**x).logits[0, -1].float()
    p = torch.softmax(torch.maximum(lg[ids], lg[ids2]), 0)
    pred = int(p.argmax()) + 1; ok += pred == y
    print(y, pred, [round(v, 2) for v in p.tolist()])
print("readout acc", ok, "/", len(CASES))
if train:
    from peft import LoraConfig, get_peft_model, prepare_model_for_kbit_training
    m = prepare_model_for_kbit_training(m); m = get_peft_model(m, LoraConfig(r=16, lora_alpha=32, target_modules="all-linear", task_type="CAUSAL_LM"))
    m.print_trainable_parameters(); m.train()
    x = tok(prompt(CASES[0][0]) + "A", return_tensors="pt").to(0)
    out = m(**x, labels=x["input_ids"]); out.loss.backward(); print("loss", float(out.loss), "peak GB", round(torch.cuda.max_memory_allocated()/1e9,1), "TRAIN STEP OK")
```
