## Supplementary material for "Non-generative readout with few-label calibration versus LoRA fine-tuning of a large language model for ASA physical status classification: accuracy, speed, cost and label efficiency in three data sources": Analysis code and aggregate results: 01_build_asa_dataset.py

```
"""构建 ASA 分级数据集（只读 NE_ERAS_JEV 原始导出；输出患者级数据仍写回 NE_ERAS_JEV/data/derived，不复制到本项目）。
单位：手术（operation）。标签：手术麻醉概览 asa_level_norm（麻醉记录中的 ASA；麻醉前访视表 ASA 字段 0% 填写，不可用）。
输入：手术前最近一次麻醉前访视（≤ 麻醉开始，且 ≤30 天）。不使用任何术中或术后信息。"""
import sys, os, re, numpy as np, pandas as pd
sys.path.insert(0, os.path.expanduser("~/research/NE_ERAS_JEV/analysis/scripts/lib"))
from v2io import load, ROOT
num = lambda s: pd.to_numeric(s, errors="coerce")
OUT = f"{ROOT}/data/derived/asa_dataset_v1.csv"; L = []
def p(s=""): print(s); L.append(s)

ov = load("手术麻醉概览").dropna(subset=["operation_id"]).drop_duplicates("operation_id")
ov["t_anes"] = pd.to_datetime(ov.anesthesia_start_time, errors="coerce")
p("# ASA 数据集构建报告（v1）\n")
p(f"- 手术麻醉概览 {len(ov):,} 台；有麻醉开始时间 {ov.t_anes.notna().sum():,}；有 ASA {num(ov.asa_level_norm).notna().sum():,}")
ops = ov[ov.t_anes.notna() & num(ov.asa_level_norm).between(1, 5)].copy()
ops["asa"] = num(ops.asa_level_norm).astype(int)
p(f"- 同时有麻醉开始时间与 ASA：{len(ops):,} 台，{ops.patient_sn.nunique():,} 名患者")

# 基线
vis = load("全部就诊").drop_duplicates("patient_sn")
base = load("", usecols=["patient_sn", "gender"])
ops = ops.merge(vis[["patient_sn", "age_double"]], on="patient_sn", how="left").merge(base, on="patient_sn", how="left")
h, w = num(ops.patient_height), num(ops.patient_weight)
ops["weight_kg"] = w.where(w.between(25, 200)); ops["age"] = num(ops.age_double); ops["female"] = (ops.gender == "女").astype(int)
ops["bmi"] = (w.where(w.between(25, 200)) / (h.where(h.between(120, 210)) / 100) ** 2)
ops["emergency"] = (ops.prioritys == "急诊").astype(int)
# 身高体重缺失者：用「生命体征」表（体温单）补。身高取任一时间最近一次；体重取手术前 30 天内最近一次
_vs = load("生命体征", usecols=["patient_sn", "exam_time", "height", "weight"])
_vs["t"] = pd.to_datetime(_vs.exam_time, errors="coerce"); _vs["h"] = num(_vs.height).where(lambda x: x.between(120, 210)); _vs["w"] = num(_vs.weight).where(lambda x: x.between(25, 200))
_m = ops[["operation_id", "patient_sn", "t_anes"]].merge(_vs, on="patient_sn"); _m["dt"] = (_m.t - _m.t_anes).dt.total_seconds().abs()
_h = _m.dropna(subset=["h"]).sort_values("dt").drop_duplicates("operation_id").set_index("operation_id").h
_w = _m[(_m.t <= _m.t_anes) & (_m.t >= _m.t_anes - pd.Timedelta(days=30))].dropna(subset=["w"]).sort_values("t").drop_duplicates("operation_id", keep="last").set_index("operation_id").w
_h0, _w0 = h.isna().sum(), w.isna().sum()
h = h.where(h.notna(), ops.operation_id.map(_h)); w = w.where(w.notna(), ops.operation_id.map(_w))
p(f"- 体温单补充：身高缺失 {_h0}→{h.isna().sum()}，体重缺失 {_w0}→{w.isna().sum()}（均为手术台数）")
ops["weight_kg"] = w.where(w.between(25, 200)); ops["bmi"] = ops.weight_kg / (h.where(h.between(120, 210)) / 100) ** 2

ops = ops[ops.age >= 18]; p(f"- 年龄 ≥18 岁：{len(ops):,} 台")

# 麻醉前访视：手术前最近一次，且 30 天内
pv = load("麻醉前访视"); pv["t_pv"] = pd.to_datetime(pv.anesthetic_time, errors="coerce")
m = ops[["operation_id", "patient_sn", "t_anes"]].merge(pv, on="patient_sn")
m = m[(m.t_pv <= m.t_anes) & (m.t_pv >= m.t_anes - pd.Timedelta(days=30))].sort_values("t_pv").drop_duplicates("operation_id", keep="last")
ops = ops.merge(m.drop(columns=["patient_sn", "t_anes"]), on="operation_id", how="inner")
p(f"- 有手术前 30 天内的麻醉前访视：{len(ops):,} 台")

# 结构化特征（SBP/DBP 英文名互换，按中文标签：diastolic_pressure 列实为收缩压）
ops["sbp"] = num(ops.diastolic_pressure); ops["dbp"] = num(ops.systolic_pressure)
bad = ~ops.sbp.between(60, 260) | ~ops.dbp.between(30, 160) | (ops.sbp <= ops.dbp); ops.loc[bad, ["sbp", "dbp"]] = np.nan
ops["hr"] = num(ops.p_cf).where(lambda s: s.between(30, 200)); ops["rr"] = num(ops.r_cf).where(lambda s: s.between(6, 50))
# 口径统一（与 MOVER 一致）：BP、心率均取体温单中麻醉开始前最近一次，不设时间间隔上限；覆盖访视表中的值
_v = load("生命体征", usecols=["patient_sn", "exam_time", "systolic_pressure", "diastolic_pressure", "heart_rate", "pulse"])
_v["t"] = pd.to_datetime(_v.exam_time, errors="coerce"); _v["s"], _v["d"] = num(_v.systolic_pressure), num(_v.diastolic_pressure)
_v["h"] = num(_v.heart_rate).where(lambda x: x.between(30, 200)); _v["h"] = _v.h.where(_v.h.notna(), num(_v.pulse).where(lambda x: x.between(30, 200)))
_v.loc[~(_v.s.between(60, 260) & _v.d.between(30, 160) & (_v.s > _v.d)), ["s", "d"]] = np.nan
_q = ops[["operation_id", "patient_sn", "t_anes"]].merge(_v[["patient_sn", "t", "s", "d", "h"]], on="patient_sn"); _q = _q[_q.t <= _q.t_anes].sort_values("t")
_bp = _q.dropna(subset=["s"]).drop_duplicates("operation_id", keep="last").set_index("operation_id"); _hr = _q.dropna(subset=["h"]).drop_duplicates("operation_id", keep="last").set_index("operation_id").h
ops["sbp"], ops["dbp"], ops["hr"] = ops.operation_id.map(_bp.s), ops.operation_id.map(_bp.d), ops.operation_id.map(_hr)
p(f"- 体温单麻醉开始前最近一次：BP {ops.sbp.notna().mean():.3f}，心率 {ops.hr.notna().mean():.3f}（无时间间隔上限）")
ops["temp"] = num(ops.temperature).where(lambda s: s.between(34, 41))
for k, c in [("htn", "is_hypertension"), ("chd", "is_coronary_heart_disease"), ("dm", "is_diabetes")]: ops[k] = (ops[c] == "是").astype(int)
G = {"Ⅰ": 1, "1": 1, "I": 1, "Ⅱ": 2, "2": 2, "II": 2, "Ⅲ": 3, "3": 3, "III": 3, "Ⅳ": 4, "4": 4, "IV": 4}
ops["cardiac_grade"] = ops.cardiac_function_grading.map(G)
lab = load("全部检验", usecols=["patient_sn", "test_time", "item_name", "test_result"])
lab = lab[lab.item_name.isin(["肌酐(Crea)-静脉血", "血红蛋白(Hb)-静脉血", "白蛋白(ALB)-静脉血"])]
lab = lab.assign(t=pd.to_datetime(lab.test_time, errors="coerce"), v=num(lab.test_result)).dropna(subset=["t", "v"])
lab = lab.merge(ops[["operation_id", "patient_sn", "t_anes"]], on="patient_sn")
lab = lab[(lab.t <= lab.t_anes) & (lab.t >= lab.t_anes - pd.Timedelta(days=30))].sort_values("t").drop_duplicates(["operation_id", "item_name"], keep="last")
for k, it in [("cr", "肌酐(Crea)-静脉血"), ("hb", "血红蛋白(Hb)-静脉血"), ("alb", "白蛋白(ALB)-静脉血")]:
    ops = ops.merge(lab[lab.item_name == it].set_index("operation_id").v.rename(k), on="operation_id", how="left")

# 文本：只用术前访视字段；ASA 字样屏蔽
ASA_PAT = re.compile(r"(?i)ASA\s*(?:分级|级|评分|physical status)?\s*[:：]?\s*(?:class\s*)?(?:[IVⅠⅡⅢⅣⅤ]+|[1-5])\s*(?:级|[-~/＋+](?:[IVⅠⅡⅢⅣⅤ]+|[1-5])\s*级?)?|ASA|美国麻醉医师协会(?:分级|评分)?\s*[:：]?\s*(?:[IVⅠⅡⅢⅣⅤ]+|[1-5])?\s*级?")
FIELDS = [("术前诊断", "pre_operative_diagnosis"), ("拟行手术", "planned_procedure_name_cn"), ("其他疾病", "other_diseases"),
          ("肺功能", "pulmonary_function"), ("肺部疾病", "pulmonary_disease_name"), ("术前特殊情况", "special_circumstances"),
          ("其他说明", "preoperative_visit_structure")]
def clean(v): return "" if pd.isna(v) or str(v).strip() in ("", "无", "nan") else str(v).strip()
raw_leak = ops.apply(lambda r: any(ASA_PAT.search(clean(r[c])) for _, c in FIELDS), axis=1)
def build(r):
    parts = [f"{int(r.age)}岁{'女' if r.female else '男'}"] if pd.notna(r.age) else []
    if pd.notna(r.bmi): parts.append(f"BMI {r.bmi:.1f}")
    if pd.notna(r.sbp): parts.append(f"血压 {r.sbp:.0f}/{r.dbp:.0f} mmHg")
    if pd.notna(r.hr): parts.append(f"心率 {r.hr:.0f}")
    parts.append("高血压" if r.htn else "无高血压"); parts.append("冠心病" if r.chd else "无冠心病"); parts.append("糖尿病" if r.dm else "无糖尿病")
    if pd.notna(r.cardiac_grade): parts.append(f"心功能 {int(r.cardiac_grade)} 级")
    if r.emergency: parts.append("急诊")
    out = ["；".join(parts)]
    for lab_, c in FIELDS:
        v = ASA_PAT.sub("[已屏蔽]", clean(r[c]))
        if v: out.append(f"{lab_}：{v}")
    return "\n".join(out)
ops["text"] = ops.apply(build, axis=1)
ops["text_len"] = ops.text.str.len()
p(f"- 原始访视文本含 ASA 字样（已屏蔽）：{raw_leak.sum():,} 台（{raw_leak.mean():.1%}）；屏蔽后残留 'ASA'：{ops.text.str.contains('(?i)ASA').sum()}")

# 时间切分：按患者首台手术年份；验证期患者若在开发期有手术则剔除（避免跨期泄漏）
ops["year"] = ops.t_anes.dt.year
first = ops.groupby("patient_sn").year.min().rename("first_year")
ops = ops.merge(first, on="patient_sn")
ops["split"] = np.where(ops.first_year <= 2024, "dev", "val")
ops = ops[~((ops.split == "val") & (ops.year <= 2024))]
ops.loc[(ops.split == "dev") & (ops.year >= 2025), "split"] = "dev_drop"   # 开发期患者在验证期的再次手术：不进入任何集合
ops = ops[ops.split != "dev_drop"]
p("\n## 年份 × 集合\n"); p(pd.crosstab(ops.year, ops.split).to_markdown())
p("\n## ASA 分布\n"); p(pd.crosstab(ops.asa, ops.split, margins=True).to_markdown())
p(f"\n- 患者数：{ops.patient_sn.nunique():,}；每名患者最多手术台数 {ops.groupby('patient_sn').size().max()}（同一患者不跨 dev/val）")
p("\n## 文本长度（字符）\n"); p(ops.text_len.describe().round(0).to_frame().T.to_markdown())
p("\n## 结构化特征缺失率\n"); p(ops[["age", "bmi", "sbp", "hr", "temp", "rr", "cardiac_grade", "cr", "hb", "alb"]].isna().mean().round(3).to_frame("缺失率").T.to_markdown())
keep = ["operation_id", "patient_sn", "year", "split", "asa", "age", "female", "bmi", "emergency", "weight_kg", "sbp", "dbp", "hr", "rr", "temp", "htn", "chd", "dm", "cardiac_grade", "cr", "hb", "alb", "surgical_name", "text"]
ops[keep].to_csv(OUT, index=False, encoding="utf-8-sig")
try: os.chmod(OUT, 0o600)
except PermissionError: pass
p(f"\n输出：{OUT}（{len(ops):,} 行；权限 600；患者级数据不复制到本项目）")
open(os.path.expanduser("~/research/Anes_JEV_vs_LoRA/analysis/reports/asa_dataset_v1.md"), "w").write("\n".join(L))
```
