## Supplementary material for "Non-generative readout with few-label calibration versus LoRA fine-tuning of a large language model for ASA physical status classification: accuracy, speed, cost and label efficiency in three data sources": Analysis code and aggregate results: 02_build_mover_dataset.py

```
"""MOVER（Epic）ASA 数据集：与本院共有的变量。输出 NE_ERAS_JEV/data/derived/asa_mover_v1.csv。
注：patient_history 无日期，慢性病标志（HTN/CHD/DM）可能含术后录入的诊断，作为已知局限；术前生命体征待 flowsheets。"""
import os, re, numpy as np, pandas as pd
R = os.path.expanduser("~/research/NE_ERAS_JEV"); E = f"{R}/data/raw/mover/extracted/epic/EPIC_EMR/EMR"; OUT = f"{R}/data/derived/asa_mover_v1.csv"
L = []
def p(s=""): print(s); L.append(s)
num = lambda s: pd.to_numeric(s, errors="coerce")
d = pd.read_csv(f"{E}/patient_information.csv", low_memory=False)
p("# MOVER ASA 数据集构建报告（v1）\n"); p(f"- patient_information {len(d):,} 行；LOG_ID {d.LOG_ID.nunique():,}；患者 {d.MRN.nunique():,}")
d = d.drop_duplicates("LOG_ID"); d["asa"] = num(d.ASA_RATING_C); d = d[d.asa.between(1, 5)].copy(); d["asa"] = d.asa.astype(int)
p(f"- 去重后有 ASA I–V：{len(d):,} 台，{d.MRN.nunique():,} 名患者")
d["age"] = num(d.BIRTH_DATE)  # 该列实为年龄（17–90）
d = d[d.age >= 18]; p(f"- 年龄 ≥18：{len(d):,} 台")
d["female"] = (d.SEX == "Female").astype(int); d["weight_kg"] = (num(d.WEIGHT) / 35.274).where(lambda s: s.between(25, 250))
_h = d.HEIGHT.astype(str).str.extract(r"^\s*(\d+)'\s*(\d+)?")   # 身高为英尺英寸字符串，如 5' 4
d["height_cm"] = ((num(_h[0]) * 12 + num(_h[1]).fillna(0)) * 2.54).where(lambda s: s.between(120, 220)); d["bmi"] = d.weight_kg / (d.height_cm / 100) ** 2
d["inpatient"] = (d.PATIENT_CLASS_GROUP == "Inpatient").astype(int)
d["t_or"] = pd.to_datetime(d.IN_OR_DTTM, errors="coerce"); d["procedure"] = d.PRIMARY_PROCEDURE_NM.fillna("").str.strip().str.lower()
d["year"] = pd.to_datetime(d.SURGERY_DATE, errors="coerce").dt.year
h = pd.read_csv(f"{E}/patient_history.csv", dtype=str)
for k, pat in [("htn", r"^(401|402|403|404|405|I1[0-6])"), ("chd", r"^(410|411|412|413|414|I2[0-5])"), ("dm", r"^(250|E1[0-3])")]:
    d[k] = d.MRN.isin(set(h[h.diagnosis_code.fillna("").str.match(pat)].mrn)).astype(int)
# 术前化验（30 天内最近一次）
want = {"2160-0": "cr", "718-7": "hb", "30350-3": "hb", "1751-7": "alb"}; parts = []
for ch in pd.read_csv(f"{E}/patient_labs.csv", usecols=["LOG_ID", "Lab Code", "Observation Value", "Collection Datetime"], dtype=str, chunksize=2_000_000):
    ch = ch[ch["Lab Code"].isin(want) & ch.LOG_ID.isin(set(d.LOG_ID))]; parts.append(ch)
lab = pd.concat(parts); lab["k"] = lab["Lab Code"].map(want); lab["v"] = num(lab["Observation Value"]); lab["t"] = pd.to_datetime(lab["Collection Datetime"], errors="coerce")
lab = lab.dropna(subset=["v", "t"]).merge(d[["LOG_ID", "t_or"]], on="LOG_ID")
lab = lab[(lab.t <= lab.t_or) & (lab.t >= lab.t_or - pd.Timedelta(days=30))].sort_values("t").drop_duplicates(["LOG_ID", "k"], keep="last")
for k in ["cr", "hb", "alb"]: d = d.merge(lab[lab.k == k].set_index("LOG_ID").v.rename(k), on="LOG_ID", how="left")
# 按患者随机 80/20（固定种子）
rng = np.random.default_rng(20261017); pats = d.MRN.unique(); test = set(rng.choice(pats, int(len(pats) * 0.2), replace=False))
d["split"] = np.where(d.MRN.isin(test), "test20", "train80")
p("\n## 集合 × ASA\n"); p(pd.crosstab(d.asa, d.split, margins=True).to_markdown())
p("\n## 年份\n"); p(d.year.value_counts().sort_index().to_frame("n").T.to_markdown())
p("\n## 缺失率\n"); p(d[["age", "bmi", "weight_kg", "cr", "hb", "alb"]].isna().mean().round(3).to_frame("缺失率").T.to_markdown())
p(f"\n- 慢性病标志比例：HTN {d.htn.mean():.1%}，CHD {d.chd.mean():.1%}，DM {d.dm.mean():.1%}")
keep = ["LOG_ID", "MRN", "year", "split", "asa", "age", "female", "weight_kg", "height_cm", "bmi", "inpatient", "htn", "chd", "dm", "cr", "hb", "alb", "procedure"]
d[keep].to_csv(OUT, index=False); os.chmod(OUT, 0o600); p(f"\n输出：{OUT}（{len(d):,} 行，权限 600）")
open(os.path.expanduser("~/research/Anes_JEV_vs_LoRA/analysis/reports/asa_mover_v1.md"), "w").write("\n".join(L))
```
