## Supplementary material for "Non-generative readout with few-label calibration versus LoRA fine-tuning of a large language model for ASA physical status classification: accuracy, speed, cost and label efficiency in three data sources": Analysis code and aggregate results: 03_render_common.py

```
"""两库共有变量 → 统一英文模板文本 + 统一结构化特征。输出 NE_ERAS_JEV/data/derived/asa_common_{mover,local}.csv。
换算：本院肌酐 μmol/L→mg/dL（/88.4），血红蛋白、白蛋白 g/L→g/dL（/10）。本院手术名保留中文原文（模型为双语）。
共有变量：年龄、性别、体重、BMI、住院/门诊、高血压/冠心病/糖尿病、肌酐、血红蛋白、白蛋白、术式。"""
import os, numpy as np, pandas as pd
R = os.path.expanduser("~/research/NE_ERAS_JEV/data/derived")
def render(r):
    f = lambda v, fmt: "unknown" if pd.isna(v) else fmt % v
    return ("Age: %s years; Sex: %s; Weight: %s kg; BMI: %s; Setting: %s\n"
            "Preoperative vitals (last before OR): BP %s/%s mmHg, heart rate %s bpm\n"
            "History: hypertension %s, coronary artery disease %s, diabetes %s\n"
            "Preoperative labs: creatinine %s mg/dL, hemoglobin %s g/dL, albumin %s g/dL\n"
            "Planned procedure: %s") % (f(r.age, "%.0f"), "female" if r.female else "male", f(r.weight_kg, "%.0f"), f(r.bmi, "%.1f"),
            "inpatient" if r.inpatient else "outpatient", f(r.sbp, "%.0f"), f(r.dbp, "%.0f"), f(r.hr, "%.0f"), *["yes" if r[k] else "no" for k in ("htn", "chd", "dm")],
            f(r.cr_mgdl, "%.2f"), f(r.hb_gdl, "%.1f"), f(r.alb_gdl, "%.1f"), (r.procedure or "unknown"))
m = pd.read_csv(f"{R}/asa_mover_v1.csv"); m = m.rename(columns={"LOG_ID": "op_id"}); m["cr_mgdl"], m["hb_gdl"], m["alb_gdl"] = m.cr, m.hb, m.alb
l = pd.read_csv(f"{R}/asa_dataset_v1.csv"); l = l.rename(columns={"operation_id": "op_id"}); l["inpatient"] = 1
l["cr_mgdl"], l["hb_gdl"], l["alb_gdl"] = l.cr / 88.4, l.hb / 10, l.alb / 10; l["procedure"] = l.surgical_name.fillna("").str.strip(" ,，")
m["procedure"] = m["procedure"].fillna("")
cols = ["op_id", "split", "asa", "age", "female", "weight_kg", "bmi", "inpatient", "sbp", "dbp", "hr", "htn", "chd", "dm", "cr_mgdl", "hb_gdl", "alb_gdl", "procedure"]
for name, d in (("mover", m), ("local", l)):
    d = d[cols].copy(); d["asa4"] = d.asa.clip(upper=4); d["text"] = d.apply(render, axis=1)
    out = f"{R}/asa_common_{name}.csv"; d.to_csv(out, index=False, encoding="utf-8-sig")
    try: os.chmod(out, 0o600)
    except PermissionError: pass
    print(name, len(d), "ASA4", (d.asa4.value_counts(normalize=True).sort_index() * 100).round(1).to_dict(), "| labs available", d.cr_mgdl.notna().mean().round(2))
print(d.text.iloc[0]); print("---"); print(m.assign(asa4=1).pipe(lambda x: x).iloc[0:0].shape)
```
