## Supplementary material for "Non-generative readout with few-label calibration versus LoRA fine-tuning of a large language model for ASA physical status classification: accuracy, speed, cost and label efficiency in three data sources": Analysis code and aggregate results: 04_extract_flowsheet_vitals.py

```
"""流式读取 MOVER Epic flowsheets 压缩包（不解压到磁盘），仅保留 Vital Signs 类的 BP/Pulse/Temp/Resp/SpO2/MAP/体重/身高，且 RECORD_TYPE=PRE-OP、记录时间不晚于入手术室的行（不设时间间隔上限），每台手术每个指标只保留入室前最近一次（分块内；若同一 LOG_ID 跨分块，后处理再取最近），
并统计各 (FLO_NAME, FLO_DISPLAY_NAME, RECORD_TYPE) 的行数。输出：data/raw/mover/extracted/flowsheet_preop_vitals.csv 与统计表。"""
import tarfile, pandas as pd, os, collections, time, io
class R(io.RawIOBase):
    def __init__(s, f): s.f = f
    def readable(s): return True
    def seekable(s): return False
    def readinto(s, b):
        d = s.f.read(len(b)); b[:len(d)] = d; return len(d)
SRC = "/mnt/c/backup/mover_raw/Epic_flowsheets_cleaned.tar.gz"
OUT = os.path.expanduser("~/research/NE_ERAS_JEV/data/raw/mover/extracted")
E = os.path.expanduser("~/research/NE_ERAS_JEV/data/raw/mover/extracted/epic/EPIC_EMR/EMR/patient_information.csv")
_pi = pd.read_csv(E, usecols=["LOG_ID", "IN_OR_DTTM"], dtype=str).drop_duplicates("LOG_ID"); _pi["t_or"] = pd.to_datetime(_pi.IN_OR_DTTM, errors="coerce")
T_OR = _pi.dropna(subset=["t_or"]).set_index("LOG_ID").t_or
ITEMS = {"BP", "Pulse", "Temp", "Resp", "SpO2", "MAP (mmHg)", "Weight", "Height"}
cnt = collections.Counter(); first = True; t0 = time.time(); rows = 0
with tarfile.open(SRC, "r|gz") as tf, open(f"{OUT}/flowsheet_preop_vitals.csv", "w", encoding="utf-8") as o:
    for m in tf:
        if not m.isfile(): continue
        f = io.TextIOWrapper(io.BufferedReader(R(tf.extractfile(m)), 1 << 20), encoding='utf-8')
        for ch in pd.read_csv(f, usecols=["LOG_ID", "FLO_NAME", "FLO_DISPLAY_NAME", "RECORD_TYPE", "RECORDED_TIME", "MEAS_VALUE"], dtype=str, chunksize=2_000_000):
            ch["FLO_NAME"] = ch.FLO_NAME.str.strip(); ch["FLO_DISPLAY_NAME"] = ch.FLO_DISPLAY_NAME.str.strip()
            for k, v in ch.groupby(["FLO_NAME", "FLO_DISPLAY_NAME", "RECORD_TYPE"], dropna=False).size().items(): cnt[k] += v
            keep = ch[ch.FLO_NAME.str.contains("Vital Signs", na=False) & (ch.RECORD_TYPE == "PRE-OP") & ch.FLO_DISPLAY_NAME.isin(ITEMS)].copy()
            keep["t"] = pd.to_datetime(keep.RECORDED_TIME, errors="coerce"); keep["t_or"] = keep.LOG_ID.map(T_OR)
            keep = keep[(keep.t <= keep.t_or)].drop(columns=["t_or"]).drop_duplicates()
            keep = keep.sort_values("t").groupby(["LOG_ID", "FLO_DISPLAY_NAME"], as_index=False).tail(1)  # 每台手术每个指标只留入室前最近一次（跨分片的合并在后处理）
            keep.to_csv(o, header=first, index=False); first = False; rows += len(keep)
        print(m.name, "kept", rows, f"{time.time() - t0:.0f}s", flush=True)
pd.Series(cnt, name="n").rename_axis(["FLO_NAME", "FLO_DISPLAY_NAME", "RECORD_TYPE"]).sort_values(ascending=False).head(400).to_csv(f"{OUT}/flowsheet_name_counts.csv")
os.chmod(f"{OUT}/flowsheet_preop_vitals.csv", 0o600); print("DONE", rows)
```
