## Supplementary material for "Non-generative readout with few-label calibration versus LoRA fine-tuning of a large language model for ASA physical status classification: accuracy, speed, cost and label efficiency in three data sources": Analysis code and aggregate results: 05_merge_mover_vitals.py

```
"""把 flowsheets 术前最近一次 BP/脉搏并入 asa_mover_v1.csv（新增 sbp, dbp, hr, vit_min_before_or 之外的列仅 sbp/dbp/hr）。
跨分片同一 LOG_ID 取记录时间最近者；合理范围外置缺失（SBP 50-260，DBP 20-150，HR 20-220）。"""
import os, pandas as pd
R = os.path.expanduser("~/research/NE_ERAS_JEV/data")
d = pd.read_csv(f"{R}/raw/mover/extracted/flowsheet_preop_vitals.csv", dtype=str); d["t"] = pd.to_datetime(d.t)
d = d.sort_values("t").groupby(["LOG_ID", "FLO_DISPLAY_NAME"]).tail(1)
bp = d[d.FLO_DISPLAY_NAME == "BP"].set_index("LOG_ID").MEAS_VALUE.str.extract(r"^\s*(\d+)\s*/\s*(\d+)\s*$").astype(float); bp.columns = ["sbp", "dbp"]
bp = bp.where(bp.sbp.between(50, 260) & bp.dbp.between(20, 150))
bp["sbp"], bp["dbp"] = bp.sbp.where(bp.dbp.notna()), bp.dbp
hr = pd.to_numeric(d[d.FLO_DISPLAY_NAME == "Pulse"].set_index("LOG_ID").MEAS_VALUE, errors="coerce"); hr = hr.where(hr.between(20, 220)).rename("hr")
m = pd.read_csv(f"{R}/derived/asa_mover_v1.csv").drop(columns=["sbp", "dbp", "hr"], errors="ignore")
m = m.merge(bp, left_on="LOG_ID", right_index=True, how="left").merge(hr, left_on="LOG_ID", right_index=True, how="left")
out = f"{R}/derived/asa_mover_v1.csv"; m.to_csv(out, index=False)
try: os.chmod(out, 0o600)
except PermissionError: pass
print(len(m), m[["sbp", "dbp", "hr"]].notna().mean().round(3).to_dict())
```
