## Supplementary material for "Non-generative readout with few-label calibration versus LoRA fine-tuning of a large language model for ASA physical status classification: accuracy, speed, cost and label efficiency in three data sources": Analysis code and aggregate results: 06_zeroshot_dev200.py

```
"""本院开发集（dev，2021–24）随机 200 台，Qwen3.5-9B 零样本选项字母 logits 读出：测速度、概率输出与粗略性能。
只读 dev，不碰封存验证集。逐例概率写入 NE_ERAS_JEV/data/derived/（患者级，不放本项目）。用法：06_zeroshot_dev200.py <model_dir> [n]"""
import os, sys, time, numpy as np, pandas as pd, torch
from sklearn.metrics import cohen_kappa_score
from transformers import AutoTokenizer, AutoModelForCausalLM
path = sys.argv[1]; n = int(sys.argv[2]) if len(sys.argv) > 2 else 200
D = os.path.expanduser("~/research/NE_ERAS_JEV/data/derived")
d = pd.read_csv(f"{D}/asa_common_local.csv"); d = d[d.split == "dev"].sample(n, random_state=20261017).reset_index(drop=True)
OPT = "ABCD"
def prompt(t):
    return ("You are an anesthesiologist. Based on the preoperative information below, assign the ASA Physical Status class.\n\n%s\n\n"
            "Options:\nA. ASA I\nB. ASA II\nC. ASA III\nD. ASA IV-V\nAnswer:") % t
tok = AutoTokenizer.from_pretrained(path); m = AutoModelForCausalLM.from_pretrained(path, dtype=torch.float16, device_map={"": 0}).eval()
ids = [tok.encode(" " + c, add_special_tokens=False)[-1] for c in OPT]; ids2 = [tok.encode(c, add_special_tokens=False)[-1] for c in OPT]
P = []; t0 = time.time()
for t in d.text:
    x = tok(prompt(t), return_tensors="pt").to(0)
    with torch.no_grad(): lg = m(**x).logits[0, -1].float()
    P.append(torch.softmax(torch.maximum(lg[ids], lg[ids2]), 0).cpu().numpy())
el = time.time() - t0; P = np.array(P); pred = P.argmax(1) + 1; y = d.asa4.values
out = pd.DataFrame(P, columns=[f"p{i}" for i in range(1, 5)]); out.insert(0, "op_id", d.op_id); out["asa4"] = y; out["pred"] = pred
out.to_csv(f"{D}/zeroshot_dev{n}_qwen35_9b.csv", index=False)
print(f"n={n} 总用时 {el:.0f}s，{el/n:.2f}s/例，峰值显存 {torch.cuda.max_memory_allocated()/1e9:.1f} GB")
print("准确率", round((pred == y).mean(), 3), "| 加权κ(二次)", round(cohen_kappa_score(y, pred, weights="quadratic"), 3), "| 多数类准确率", round((y == 2).mean(), 3))
print("预测分布", np.bincount(pred, minlength=5)[1:].tolist(), "真实分布", np.bincount(y, minlength=5)[1:].tolist(), "| 平均最大概率", round(P.max(1).mean(), 3))
```
