## Supplementary material for "Non-generative readout with few-label calibration versus LoRA fine-tuning of a large language model for ASA physical status classification: accuracy, speed, cost and label efficiency in three data sources": Analysis code and aggregate results: 07_infer_zeroshot.py

```
"""零样本读出：llm = Qwen3.5-9B 选项字母 logits（生成式零样本）。
用法：07_infer_zeroshot.py <llm> <local|mover_test> [gpu]
输出 NE_ERAS_JEV/data/derived/pred_<arm>_<set>.csv（op_id, asa4, p1..p4）；断点续跑。"""
import os, sys, time, numpy as np, pandas as pd, torch
arm, ds = sys.argv[1], sys.argv[2]; gpu = int(sys.argv[3]) if len(sys.argv) > 3 else 0
D = os.path.expanduser("~/research/NE_ERAS_JEV/data/derived")
d = (lambda x: x[x.op_id.isin(pd.read_csv(f"{D}/vitaldb_sub2000.csv").op_id)])(pd.read_csv(f"{D}/asa_common_vitaldb.csv")) if ds == "vitaldb" else pd.read_csv(f"{D}/asa_common_{'local' if ds.startswith('local') else 'mover'}.csv")
if ds == "mover_test": d = d[d.split == "test20"]
if ds == "mover_cal": d = d[d.split == "train80"].sample(1000, random_state=11)   # 校准集：训练划分内 1000 台
if ds == "mover_sub": d = d[d.op_id.isin(pd.read_csv(f"{D}/mover_test_sub4000.csv").op_id)]
SUF = ""
if ds == "local_t2":
    raw = pd.read_csv(f"{D}/asa_dataset_v1.csv", usecols=["operation_id", "text"]).rename(columns={"operation_id": "op_id", "text": "raw"}); d = d.merge(raw, on="op_id", how="left"); d["text"] = d.raw
if os.environ.get("TEXTV") == "t2":
    t2 = pd.read_csv(f"{D}/asa_t2_mover.csv").set_index("op_id").text2; d["text"] = d.op_id.map(t2); SUF = "_t2"
d = d.reset_index(drop=True); out = f"{D}/pred_{arm}_{ds}{SUF}{os.environ.get('MTAG', '')}.csv"
done = pd.read_csv(out) if os.path.exists(out) else pd.DataFrame(columns=["op_id"])
todo = d[~d.op_id.isin(done.op_id)]; print(arm, ds, "总", len(d), "待做", len(todo), flush=True)
dev = f"cuda:{gpu}"; rows = []; t0 = time.time()
def flush():
    global done, rows
    if rows: done = pd.concat([done, pd.DataFrame(rows)]); done.to_csv(out, index=False); rows = []
if arm == "llm":
    from transformers import AutoTokenizer, AutoModelForCausalLM
    path = os.environ.get("MODEL", "/mnt/c/backup/Qwen3.5-9B"); tok = AutoTokenizer.from_pretrained(path)
    if os.environ.get("QUANT") == "4":
        from transformers import BitsAndBytesConfig
        bnb = BitsAndBytesConfig(load_in_4bit=True, bnb_4bit_quant_type="nf4", bnb_4bit_compute_dtype=torch.float16, bnb_4bit_use_double_quant=True)
        m = AutoModelForCausalLM.from_pretrained(path, quantization_config=bnb, dtype=torch.float16, device_map={"": gpu}).eval()
    else: m = AutoModelForCausalLM.from_pretrained(path, dtype=torch.float16, device_map={"": gpu}).eval()
    ids = [tok.encode(" " + c, add_special_tokens=False)[-1] for c in "ABCD"]; ids2 = [tok.encode(c, add_special_tokens=False)[-1] for c in "ABCD"]
    P = ("You are an anesthesiologist. Based on the preoperative information below, assign the ASA Physical Status class.\n\n%s\n\n"
         "Options:\nA. ASA I\nB. ASA II\nC. ASA III\nD. ASA IV-V\nAnswer:")
    def infer(t):
        x = tok(P % t, return_tensors="pt").to(dev)
        with torch.no_grad(): lg = m(**x).logits[0, -1].float()
        return torch.softmax(torch.maximum(lg[ids], lg[ids2]), 0).cpu().numpy()
else:
    sys.exit(f"unsupported arm: {arm}")
for i, r in enumerate(todo.itertuples()):
    p = infer(r.text); rows.append(dict(op_id=r.op_id, asa4=r.asa4, p1=p[0], p2=p[1], p3=p[2], p4=p[3]))
    if (i + 1) % 500 == 0: flush(); print(i + 1, f"{(time.time()-t0)/(i+1):.2f}s/例", flush=True)
flush(); print("完成", len(done), f"{time.time()-t0:.0f}s")
```
