## Supplementary material for "Non-generative readout with few-label calibration versus LoRA fine-tuning of a large language model for ASA physical status classification: accuracy, speed, cost and label efficiency in three data sources": Analysis code and aggregate results: 08_lora.py

```
"""任务 QLoRA（Qwen3.5-9B，4-bit，r=16）：训练 MOVER train80 的随机子集，损失为末位 A–D 四个选项 logits 上的交叉熵（与零样本同一读出）。
用法：08_lora.py train <n_train> <gpu> <tag>   |   08_lora.py infer <tag> <local|mover_test> <gpu>
适配器存 ~/research/Anes_JEV_vs_LoRA/models/<tag>；预测写 NE_ERAS_JEV/data/derived/pred_lora_<tag>_<set>.csv。"""
import os, sys, time, random, numpy as np, pandas as pd, torch
from transformers import AutoTokenizer, AutoModelForCausalLM, BitsAndBytesConfig
from peft import LoraConfig, get_peft_model, PeftModel, prepare_model_for_kbit_training
SEED = int(os.environ.get("SEED", 20261017)); torch.manual_seed(SEED); mode = sys.argv[1]; path = os.environ.get("MODEL", "/mnt/c/backup/Qwen3.5-9B")
D = os.path.expanduser("~/research/NE_ERAS_JEV/data/derived"); M = os.path.expanduser("~/research/Anes_JEV_vs_LoRA/models")
P = ("You are an anesthesiologist. Based on the preoperative information below, assign the ASA Physical Status class.\n\n%s\n\n"
     "Options:\nA. ASA I\nB. ASA II\nC. ASA III\nD. ASA IV-V\nAnswer:")
tok = AutoTokenizer.from_pretrained(path); tok.padding_side = "left"
ids = torch.tensor([tok.encode(" " + c, add_special_tokens=False)[-1] for c in "ABCD"])
bnb = BitsAndBytesConfig(load_in_4bit=True, bnb_4bit_quant_type="nf4", bnb_4bit_compute_dtype=torch.float16, bnb_4bit_use_double_quant=True)
T2 = pd.read_csv(f"{D}/asa_t2_mover.csv").set_index("op_id").text2 if os.environ.get("TEXTV") == "t2" else None
def local_df():
    """本院：T1=共有结构化模板文本；T2=原始术前访视文本（asa_dataset_v1.text，ASA 字样已屏蔽）"""
    c = pd.read_csv(f"{D}/asa_common_local.csv")
    if os.environ.get("TEXTV") == "t2":
        raw = pd.read_csv(f"{D}/asa_dataset_v1.csv", usecols=["operation_id", "text"]).rename(columns={"operation_id": "op_id", "text": "raw"})
        c = c.merge(raw, on="op_id", how="left"); c["text"] = c.raw
    return c
SRC = os.environ.get("SRC", "mover")
def load(gpu): return AutoModelForCausalLM.from_pretrained(path, quantization_config=bnb, dtype=torch.float16, device_map=("auto" if gpu < 0 else {"": gpu}), **({"max_memory": {0: "19GiB", 1: "19GiB"}} if gpu < 0 else {}))
def fwd(m, texts, dev):
    x = tok([P % t for t in texts], return_tensors="pt", padding=True).to(dev)
    return m(**x, logits_to_keep=1).logits[:, -1].float()[:, ids.to(dev)]
if mode == "train":
    n, gpu, tag = int(sys.argv[2]), int(sys.argv[3]), sys.argv[4]; dev = f"cuda:{max(gpu,0)}"
    if SRC == "local":
        d = local_df(); d = d[d.split == "dev"]; d = (d.sample(n, random_state=SEED) if n < len(d) else d.sample(frac=1, random_state=SEED)).reset_index(drop=True)
    else:
        d = pd.read_csv(f"{D}/asa_common_mover.csv"); d = d[d.split == "train80"].sample(n, random_state=SEED).reset_index(drop=True)
        if T2 is not None: d["text"] = d.op_id.map(T2)
    n = len(d)
    if os.environ.get("MODEL"):   # 27B：避免 prepare_model_for_kbit_training 把 24 万词表的 embedding/lm_head 升成 fp32（各约 5GB）
        m = load(gpu); m.gradient_checkpointing_enable(gradient_checkpointing_kwargs={"use_reentrant": False}); m.enable_input_require_grads()
    else: m = prepare_model_for_kbit_training(load(gpu), use_gradient_checkpointing=True)
    m = get_peft_model(m, LoraConfig(r=16, lora_alpha=32, lora_dropout=0.05, target_modules=["q_proj", "k_proj", "v_proj", "o_proj", "gate_proj", "up_proj", "down_proj"], task_type="CAUSAL_LM"))
    m.print_trainable_parameters()
    opt = torch.optim.AdamW([p for p in m.parameters() if p.requires_grad], lr=1e-4, weight_decay=0.0)
    bs, acc = (2, 8) if os.environ.get("MODEL") else (4, 4); steps = n // (bs * acc); sched = torch.optim.lr_scheduler.OneCycleLR(opt, max_lr=1e-4, total_steps=steps, pct_start=0.05, anneal_strategy="cos")
    m.train(); t0 = time.time(); la = []
    for s in range(steps):
        for a in range(acc):
            b = d.iloc[(s * acc + a) * bs:(s * acc + a + 1) * bs]
            lg = fwd(m, list(b.text), dev); loss = torch.nn.functional.cross_entropy(lg, torch.tensor(b.asa4.values - 1, device=dev)) / acc
            loss.backward(); la.append(loss.item() * acc)
        torch.nn.utils.clip_grad_norm_(m.parameters(), 1.0); opt.step(); sched.step(); opt.zero_grad()
        if (s + 1) % 10 == 0: print(f"step {s+1}/{steps} loss {np.mean(la[-40:]):.4f} {(time.time()-t0)/(s+1):.1f}s/step", flush=True)
        if (s + 1) % 100 == 0: m.save_pretrained(f"{M}/{tag}")
    m.save_pretrained(f"{M}/{tag}"); print("训练完成", f"{time.time()-t0:.0f}s")
else:
    tag, ds, gpu = sys.argv[2], sys.argv[3], int(sys.argv[4]); dev = f"cuda:{max(gpu,0)}"
    d = (lambda x: x[x.op_id.isin(pd.read_csv(f"{D}/vitaldb_sub2000.csv").op_id)])(pd.read_csv(f"{D}/asa_common_vitaldb.csv")) if ds == "vitaldb" else (local_df() if ds.startswith("local") else pd.read_csv(f"{D}/asa_common_mover.csv"))
    if ds == "local_val": d = d[d.split == "val"]
    if ds == "mover_test": d = d[d.split == "test20"]
    if ds == "mover_sub": d = d[d.op_id.isin(pd.read_csv(f"{D}/mover_test_sub4000.csv").op_id)]
    if T2 is not None and not ds.startswith("local") and ds != "vitaldb": d = d.assign(text=d.op_id.map(T2))
    d = d.reset_index(drop=True); out = f"{D}/pred_lora_{tag}_{ds}.csv"
    m = PeftModel.from_pretrained(load(gpu), f"{M}/{tag}").eval(); rows = []; t0 = time.time(); bs = 8
    for i in range(0, len(d), bs):
        b = d.iloc[i:i + bs]
        with torch.no_grad(): p = torch.softmax(fwd(m, list(b.text), dev), -1).cpu().numpy()
        for r, q in zip(b.itertuples(), p): rows.append(dict(op_id=r.op_id, asa4=r.asa4, p1=q[0], p2=q[1], p3=q[2], p4=q[3]))
        if (i // bs + 1) % 100 == 0: print(i + bs, f"{(time.time()-t0)/(i+bs):.2f}s/例", flush=True)
    pd.DataFrame(rows).to_csv(out, index=False); print("完成", len(rows), f"{time.time()-t0:.0f}s")
```
