## Supplementary material for "Non-generative readout with few-label calibration versus LoRA fine-tuning of a large language model for ASA physical status classification: accuracy, speed, cost and label efficiency in three data sources": Analysis code and aggregate results: 09_xgb.py

```
"""XGBoost 结构化基线：MOVER train80 拟合，预测 MOVER test20 与本院全部（不含术式文本，跨库可迁移）。输出 pred_xgb_{mover_test,local}.csv"""
import os, numpy as np, pandas as pd, xgboost as xgb
D = os.path.expanduser("~/research/NE_ERAS_JEV/data/derived")
F = ["age", "female", "weight_kg", "bmi", "inpatient", "sbp", "dbp", "hr", "htn", "chd", "dm", "cr_mgdl", "hb_gdl", "alb_gdl"]
m = pd.read_csv(f"{D}/asa_common_mover.csv"); l = pd.read_csv(f"{D}/asa_common_local.csv")
tr = m[m.split == "train80"]; te = m[m.split == "test20"]
clf = xgb.XGBClassifier(n_estimators=400, max_depth=5, learning_rate=0.05, subsample=0.8, colsample_bytree=0.8, n_jobs=16, tree_method="hist", objective="multi:softprob")
clf.fit(tr[F], tr.asa4 - 1)
for name, d in (("mover_test", te), ("local", l)):
    p = clf.predict_proba(d[F]); o = pd.DataFrame(p, columns=["p1", "p2", "p3", "p4"]); o.insert(0, "asa4", d.asa4.values); o.insert(0, "op_id", d.op_id.values)
    o.to_csv(f"{D}/pred_xgb_{name}.csv", index=False); print(name, len(o), "acc", round((p.argmax(1) + 1 == d.asa4.values).mean(), 3))
```
