## Supplementary material for "Non-generative readout with few-label calibration versus LoRA fine-tuning of a large language model for ASA physical status classification: accuracy, speed, cost and label efficiency in three data sources": Analysis code and aggregate results: 10_eval.py

```
"""汇总各臂在 MOVER 测试集与本院的表现：准确率、二次加权κ、宏AUC、Brier、ECE，κ 的患者外 bootstrap 95%CI；
并给出臂间 κ 差值的配对 bootstrap CI。输出 analysis/reports/arms_eval.md"""
import os, sys, glob, numpy as np, pandas as pd
from sklearn.metrics import cohen_kappa_score, roc_auc_score
D = os.path.expanduser("~/research/NE_ERAS_JEV/data/derived"); R = os.path.expanduser("~/research/Anes_JEV_vs_LoRA/analysis/reports")
def load(arm, ds):
    f = f"{D}/pred_{arm}_{ds}.csv"; return pd.read_csv(f).set_index("op_id") if os.path.exists(f) else None
def kap(y, p): return cohen_kappa_score(y, p, weights="quadratic")
def metrics(d):
    y = d.asa4.values.astype(int); P = d[["p1", "p2", "p3", "p4"]].values; pr = P.argmax(1) + 1; Y = np.eye(4)[y - 1]
    auc = np.nanmean([roc_auc_score(Y[:, k], P[:, k]) for k in range(4) if 0 < Y[:, k].sum() < len(y)])
    conf = P.max(1); ok = (pr == y); ece = 0
    for lo in np.linspace(0, 1, 11)[:-1]:
        s = (conf > lo) & (conf <= lo + 0.1)
        if s.any(): ece += s.mean() * abs(ok[s].mean() - conf[s].mean())
    return dict(n=len(y), acc=ok.mean(), kappa=kap(y, pr), auc=auc, brier=((P - Y) ** 2).sum(1).mean(), ece=ece)
ARMS = [("llm", "生成式零样本 Qwen3.5-9B"), ("lora_mover12k", "任务 QLoRA（MOVER 12k 训练）"), ("xgb", "XGBoost（MOVER 训练）")]
rng = np.random.default_rng(20261018); out = ["# 各臂比较（MOVER 训练 → MOVER 测试 / 本院外部检验）\n"]
for ds, title in (("mover_test", "MOVER 20% 样本外"),):
    out.append(f"\n## {title}\n\n| 臂 | n | 准确率 | 加权κ (95%CI) | 宏AUC | Brier | ECE |\n|---|---|---|---|---|---|---|")
    P = {a: load(a, ds) for a, _ in ARMS}; P = {a: v for a, v in P.items() if v is not None}
    if not P: continue
    ids = sorted(set.intersection(*[set(v.index) for v in P.values()])); B = 1000; idx = rng.integers(0, len(ids), (B, len(ids)))
    kb = {}
    for a, nm in ARMS:
        if a not in P: continue
        d = P[a].loc[ids]; m = metrics(d); y = d.asa4.values.astype(int); pr = d[["p1", "p2", "p3", "p4"]].values.argmax(1) + 1
        kb[a] = np.array([kap(y[i], pr[i]) for i in idx]); lo, hi = np.percentile(kb[a], [2.5, 97.5])
        out.append(f"| {nm} | {m['n']} | {m['acc']:.3f} | {m['kappa']:.3f} ({lo:.3f}–{hi:.3f}) | {m['auc']:.3f} | {m['brier']:.3f} | {m['ece']:.3f} |")
    out.append("\n预测分布 I/II/III/IV–V：" + "；".join(f"{a}: {np.bincount(P[a].loc[ids][['p1','p2','p3','p4']].values.argmax(1), minlength=4).tolist()}" for a in P) + f"；真实: {np.bincount(P[list(P)[0]].loc[ids].asa4.values.astype(int) - 1, minlength=4).tolist()}")
    out.append("\n臂间 κ 差值（配对 bootstrap，95%CI）：")
    for a, b in (("lora_mover12k", "llm"),):
        if a in kb and b in kb: df = kb[a] - kb[b]; out.append(f"- {a} − {b}：{df.mean():+.3f}（{np.percentile(df,2.5):+.3f} ~ {np.percentile(df,97.5):+.3f}）")
open(f"{R}/arms_eval.md", "w").write("\n".join(out)); print("\n".join(out))
```
