## Supplementary material for "Non-generative readout with few-label calibration versus LoRA fine-tuning of a large language model for ASA physical status classification: accuracy, speed, cost and label efficiency in three data sources": Analysis code and aggregate results: 11_diff.py

````
"""仅 MOVER 测试集：同一底模（Qwen3.5-9B）下各臂差异在哪。输出混淆矩阵、各类召回、AUC（排序能力）、κ（阈值相关）、
以及用 4 个概率做先验/线性校正后的 κ（折半交叉：测试集随机分两半互相拟合，4 类 logit 偏置+线性，参数极少）。"""
import os, numpy as np, pandas as pd
from sklearn.metrics import cohen_kappa_score, roc_auc_score, confusion_matrix
from sklearn.linear_model import LogisticRegression
D = os.path.expanduser("~/research/NE_ERAS_JEV/data/derived"); R = os.path.expanduser("~/research/Anes_JEV_vs_LoRA/analysis/reports")
ARMS = [("llm", "生成式零样本"), ("lora_mover12k", "任务QLoRA"), ("xgb", "XGBoost")]
out = ["# MOVER 测试集：各臂差异分析（底模均为 Qwen3.5-9B，XGBoost 除外）\n"]
P = {a: pd.read_csv(f"{D}/pred_{a}_mover_test.csv").set_index("op_id") for a, _ in ARMS if os.path.exists(f"{D}/pred_{a}_mover_test.csv")}
ids = sorted(set.intersection(*[set(v.index) for v in P.values()])); rng = np.random.default_rng(20261018); half = rng.random(len(ids)) < 0.5
y = next(iter(P.values())).loc[ids].asa4.values.astype(int); out.append(f"n={len(ids)}；真实分布 I/II/III/IV–V = {np.bincount(y-1,minlength=4).tolist()}\n")
for a, nm in ARMS:
    if a not in P: continue
    X = P[a].loc[ids][["p1", "p2", "p3", "p4"]].values; pr = X.argmax(1) + 1; Y = np.eye(4)[y - 1]
    auc = [roc_auc_score(Y[:, k], X[:, k]) for k in range(4)]; cm = confusion_matrix(y, pr, labels=[1, 2, 3, 4])
    L = np.log(np.clip(X, 1e-6, 1)); cal = np.zeros(len(ids), int)
    for h in (True, False):
        lr = LogisticRegression(C=10, max_iter=2000).fit(L[half == h], y[half == h]); cal[half != h] = lr.predict(L[half != h])
    out.append(f"## {nm}\n- κ {cohen_kappa_score(y,pr,weights='quadratic'):.3f}；准确率 {(pr==y).mean():.3f}；各类 AUC(一对其余) {[round(v,3) for v in auc]}；平均最大概率 {X.max(1).mean():.3f}")
    out.append(f"- 线性校正后 κ {cohen_kappa_score(y,cal,weights='quadratic'):.3f}（折半交叉；差距=阈值/先验偏移可修复的部分）")
    out.append("- 混淆矩阵（行=真实，列=预测，I/II/III/IV–V）：\n```\n" + "\n".join(" ".join(f"{v:6d}" for v in r) for r in cm) + "\n```")
open(f"{R}/arms_diff_mover.md", "w").write("\n".join(out)); print("\n".join(out))
````
