## Supplementary material for "Non-generative readout with few-label calibration versus LoRA fine-tuning of a large language model for ASA physical status classification: accuracy, speed, cost and label efficiency in three data sources": Analysis code and aggregate results: 12_prompt_variants.py

```
"""提示词变体试验（只用 MOVER train80 随机 1500 台，不碰测试集与本院）。P0 现用；P1 ASA-PS 2020 官方表（含举例）；P2 = P1 + 操作规则。
用法：12_prompt_variants.py <llm> <gpu> [n]"""
import os, sys, time, numpy as np, pandas as pd, torch
from sklearn.metrics import cohen_kappa_score, roc_auc_score
arm, gpu = sys.argv[1], int(sys.argv[2]); n = int(sys.argv[3]) if len(sys.argv) > 3 else 1500
D = os.path.expanduser("~/research/NE_ERAS_JEV/data/derived")
d = pd.read_csv(f"{D}/asa_common_mover.csv"); d = d[d.split == "train80"].sample(n, random_state=7).reset_index(drop=True)
OFF = {"I": "A normal healthy patient. Example: healthy, non-smoking, no or minimal alcohol use.",
 "II": "A patient with mild systemic disease. Examples: current smoker, social alcohol drinker, pregnancy, obesity (30<BMI<40), well-controlled DM/HTN, mild lung disease.",
 "III": "A patient with severe systemic disease. Examples: poorly controlled DM or HTN, COPD, morbid obesity (BMI>=40), active hepatitis, alcohol dependence or abuse, implanted pacemaker, moderate reduction of ejection fraction, ESRD undergoing regularly scheduled dialysis, history (>3 months) of MI, CVA, TIA, or CAD/stents, malignancy.",
 "IV": "A patient with severe systemic disease that is a constant threat to life. Examples: recent (<3 months) MI, CVA, TIA, or CAD/stents, ongoing cardiac ischemia or severe valve dysfunction, severe reduction of ejection fraction, shock, sepsis, DIC, ARDS, ESRD not undergoing regularly scheduled dialysis.",
 "V": "A moribund patient who is not expected to survive without the operation. Examples: ruptured abdominal/thoracic aneurysm, massive trauma, intracranial bleed with mass effect, ischemic bowel with significant cardiac pathology or multiple organ dysfunction."}
RULES = ("Rules: judge only from the information given; do not infer unlisted diseases. Hypertension controlled on treatment = II, uncontrolled (>=180/110 or >=3 drugs) = III. "
         "Diabetes: non-insulin and well controlled = II, insulin-treated or poorly controlled = III. BMI>=40 = III; BMI 30-39.9 = II. Coronary artery disease or heart failure with documented limitation = III. "
         "Anemia/hypoalbuminemia/electrolyte abnormalities alone do not raise the class. Take the highest class supported by any single condition.")
def defs(v):
    if v == 0: return ""
    s = "ASA Physical Status definitions (ASA 2020):\n" + "\n".join(f"{k}. {t}" for k, t in OFF.items()) + "\n\n"
    return s + (RULES + "\n\n" if v == 2 else "")
def lab(y, p): return cohen_kappa_score(y, p, weights="quadratic")
y = d.asa4.values; res = []
if arm == "llm":
    from transformers import AutoTokenizer, AutoModelForCausalLM
    path = "/mnt/c/backup/Qwen3.5-9B"; tok = AutoTokenizer.from_pretrained(path)
    m = AutoModelForCausalLM.from_pretrained(path, dtype=torch.float16, device_map={"": gpu}).eval()
    a1 = [tok.encode(" " + c, add_special_tokens=False)[-1] for c in "ABCD"]; a2 = [tok.encode(c, add_special_tokens=False)[-1] for c in "ABCD"]
    def run(v):
        out = []
        for t in d.text:
            q = ("You are an anesthesiologist. Based on the preoperative information below, assign the ASA Physical Status class.\n\n" + defs(v) +
                 "Patient information:\n%s\n\nOptions:\nA. ASA I\nB. ASA II\nC. ASA III\nD. ASA IV-V\nAnswer:") % t
            x = tok(q, return_tensors="pt").to(f"cuda:{gpu}")
            with torch.no_grad(): lg = m(**x).logits[0, -1].float()
            out.append(torch.softmax(torch.maximum(lg[a1], lg[a2]), 0).cpu().numpy())
        return np.array(out)
else:
    sys.exit(f"unsupported arm: {arm}")

for v in (0, 1, 2):
    t0 = time.time(); P = run(v); pr = P.argmax(1) + 1; Y = np.eye(4)[y - 1]
    auc = np.mean([roc_auc_score(Y[:, k], P[:, k]) for k in range(4)])
    line = f"{arm} P{v}: κ {lab(y, pr):.3f} 准确率 {(pr==y).mean():.3f} 宏AUC {auc:.3f} 预测分布 {np.bincount(pr-1,minlength=4).tolist()} 真实 {np.bincount(y-1,minlength=4).tolist()} ({time.time()-t0:.0f}s)"
    print(line, flush=True); res.append(line)
open(os.path.expanduser(f"~/research/Anes_JEV_vs_LoRA/analysis/reports/prompt_variants_{arm}.txt"), "w").write("\n".join(res))
```
