## Supplementary material for "Non-generative readout with few-label calibration versus LoRA fine-tuning of a large language model for ASA physical status classification: accuracy, speed, cost and label efficiency in three data sources": Analysis code and aggregate results: 13_generative_zeroshot.py

```
"""生成式零样本（臂3）：Qwen3.5-9B 对话模板、关闭思考模式，先给 ≤3 句理由再给 'Final answer'，贪心解码，解析类别。
在 mover_test_sub4000 的前 N 台上运行。用法：13_generative_zeroshot.py <gpu> <N>。输出 pred_gen_mover_sub.csv（硬标签，无概率）。"""
import os, re, sys, time, numpy as np, pandas as pd, torch
from transformers import AutoTokenizer, AutoModelForCausalLM
gpu, N = int(sys.argv[1]), int(sys.argv[2]); D = os.path.expanduser("~/research/NE_ERAS_JEV/data/derived")
d = pd.read_csv(f"{D}/asa_common_mover.csv"); ids = pd.read_csv(f"{D}/mover_test_sub4000.csv").op_id.tolist()[:N]
d = d[d.op_id.isin(ids)].reset_index(drop=True); out = f"{D}/pred_gen_mover_sub.csv"
done = pd.read_csv(out) if os.path.exists(out) else pd.DataFrame(columns=["op_id"]); d = d[~d.op_id.isin(done.op_id)].reset_index(drop=True)
path = "/mnt/c/backup/Qwen3.5-9B"; tok = AutoTokenizer.from_pretrained(path); tok.padding_side = "left"
m = AutoModelForCausalLM.from_pretrained(path, dtype=torch.float16, device_map={"": gpu}).eval()
Q = ("You are an anesthesiologist. Based on the preoperative information below, assign the ASA Physical Status class (I, II, III, IV-V).\n\n%s\n\n"
     "Give a brief rationale (at most 3 sentences), then end with exactly one line: 'Final answer: ASA <I|II|III|IV-V>'.")
def prompt(t):
    msgs = [{"role": "user", "content": Q % t}]
    try: return tok.apply_chat_template(msgs, tokenize=False, add_generation_prompt=True, enable_thinking=False)
    except TypeError: return tok.apply_chat_template(msgs, tokenize=False, add_generation_prompt=True)
def parse(s):
    mm = re.findall(r"Final answer:\s*ASA\s*(IV-V|IV|III|II|I|V)\b", s)
    return {"I": 1, "II": 2, "III": 3, "IV": 4, "IV-V": 4, "V": 4}.get(mm[-1]) if mm else None
rows = []; t0 = time.time(); bs = 8
for i in range(0, len(d), bs):
    b = d.iloc[i:i + bs]; x = tok([prompt(t) for t in b.text], return_tensors="pt", padding=True).to(f"cuda:{gpu}")
    with torch.no_grad(): g = m.generate(**x, max_new_tokens=140, do_sample=False)
    txt = tok.batch_decode(g[:, x.input_ids.shape[1]:], skip_special_tokens=True)
    for r, s in zip(b.itertuples(), txt): rows.append(dict(op_id=r.op_id, asa4=r.asa4, pred=parse(s), raw=s.replace("\n", " ")[:400]))
    if (i // bs + 1) % 10 == 0:
        pd.concat([done, pd.DataFrame(rows)]).to_csv(out, index=False); print(i + bs, f"{(time.time()-t0)/(i+bs):.2f}s/例", flush=True)
pd.concat([done, pd.DataFrame(rows)]).to_csv(out, index=False); print("完成", len(rows))
```
