## Supplementary material for "Non-generative readout with few-label calibration versus LoRA fine-tuning of a large language model for ASA physical status classification: accuracy, speed, cost and label efficiency in three data sources": Analysis code and aggregate results: 14_eval_v2.py

```
"""v2 评估（仅 MOVER）：固定患者级测试子集 sub4000（生成式臂用其前 2000 台的交集）；患者为单位的 cluster bootstrap；
学习曲线（LoRA / XGBoost 随训练标签数）；校准零样本（用训练划分内 1000 台标签拟合 4 类对数概率的多项逻辑回归）。输出 reports/arms_eval_v2.md"""
import os, numpy as np, pandas as pd, xgboost as xgb
from sklearn.metrics import cohen_kappa_score, roc_auc_score
from sklearn.linear_model import LogisticRegression
D = os.path.expanduser("~/research/NE_ERAS_JEV/data/derived"); R = os.path.expanduser("~/research/Anes_JEV_vs_LoRA/analysis/reports")
sub = set(pd.read_csv(f"{D}/mover_test_sub4000.csv").op_id); pid = pd.read_csv(f"{D}/mover_pid_map.csv").set_index("op_id").pid
def rd(f): return pd.read_csv(f"{D}/{f}.csv").set_index("op_id") if os.path.exists(f"{D}/{f}.csv") else None
def sel(d): return d[d.index.isin(sub)] if d is not None else None
PC = ["p1", "p2", "p3", "p4"]
def kap(y, p): return cohen_kappa_score(y, p, weights="quadratic")
arms = {"非生成式读出零样本（OpenJEV式，Qwen3.5-9B）": sel(rd("pred_llm_mover_test")), "XGBoost（全训练集）": sel(rd("pred_xgb_mover_test")), "任务LoRA 12k": sel(rd("pred_lora_mover12k_mover_test"))}
for n in (100, 300, 1000, 3000): arms[f"任务LoRA {n}"] = rd(f"pred_lora_curve{n}_mover_sub")
arms["检索式少样本 kNN-8（无训练）"] = rd("pred_knn8_mover_sub")
# 校准零样本
for key, nm in (("llm", "非生成式读出零样本"),):
    c = rd(f"pred_{key}_mover_cal"); t = sel(rd(f"pred_{key}_mover_test"))
    if c is not None and t is not None and len(c) >= 900:
        lr = LogisticRegression(C=10, max_iter=3000).fit(np.log(np.clip(c[PC].values, 1e-6, 1)), c.asa4.astype(int))
        P = lr.predict_proba(np.log(np.clip(t[PC].values, 1e-6, 1))); q = t.copy(); q[PC] = P; arms[f"{nm} + 1000标签校准（多项逻辑回归，取众数）"] = q
        # 序数感知校准：期望分 E=Σk·p_k，3 个切点在 1000 标签上最大化加权κ（坐标搜索）
        ec = (c[PC].values * np.arange(1, 5)).sum(1); yc = c.asa4.astype(int).values; cuts = np.array([1.5, 2.5, 3.5]); grid = np.linspace(1.0, 4.0, 121)
        for _ in range(3):
            for j in range(3):
                best = max(grid, key=lambda g: kap(yc, 1 + (ec[:, None] > np.where(np.arange(3) == j, g, cuts)).sum(1))); cuts[j] = best
        et = (t[PC].values * np.arange(1, 5)).sum(1); pred = 1 + (et[:, None] > cuts).sum(1); q2 = t.copy(); q2[PC] = np.eye(4)[pred - 1] * 0.97 + 0.01; arms[f"{nm} + 1000标签序数切点校准"] = q2
g = rd("pred_gen_mover_sub")
rng = np.random.default_rng(20261018); B = 1000; out = ["# v2 评估（MOVER 患者级测试子集；cluster bootstrap 以患者为单位）\n"]
common = sorted(set.intersection(*[set(v.index) for v in arms.values() if v is not None]))
if g is not None: gc = g.dropna(subset=["pred"]); gids = sorted(set(gc.index) & set(common))
def cboot(ids, fn):
    pats = pid.loc[ids].values; up = np.unique(pats); idx = {p: np.where(pats == p)[0] for p in up}; res = []
    for _ in range(B):
        s = np.concatenate([idx[p] for p in rng.choice(up, len(up))]); res.append(fn(s))
    return np.array(res)
def table(ids, arms_, extra=None):
    rows = []; boots = {}
    for nm, d in arms_.items():
        d = d.loc[ids]; y = d.asa4.values.astype(int); pr = d[PC].values.argmax(1) + 1 if "p1" in d else d.pred.values.astype(int)
        b = cboot(ids, lambda s: kap(y[s], pr[s])); boots[nm] = b
        auc = np.mean([roc_auc_score((y == k + 1).astype(int), d[PC[k]].values) for k in range(4)]) if "p1" in d else np.nan
        rows.append(f"| {nm} | {kap(y,pr):.3f} ({np.percentile(b,2.5):.3f}–{np.percentile(b,97.5):.3f}) | {(pr==y).mean():.3f} | {auc:.3f} |")
    return rows, boots
out.append(f"## A. 全臂（共同样本 n={len(common)}，患者数 {pid.loc[common].nunique()}）\n\n| 臂 | 加权κ (95%CI) | 准确率 | 宏AUC |\n|---|---|---|---|"); r, bt = table(common, {k: v for k, v in arms.items() if v is not None}); out += r
if g is not None:
    a2 = {k: v for k, v in arms.items() if v is not None and ("零样本" in k or "任务LoRA 12k" in k or "XGBoost" in k)}; a2["生成式零样本（Qwen3.5-9B，生成理由后作答）"] = gc
    out.append(f"\n## B. 含生成式零样本（可解析 {len(gc)}/{len(g)}，共同样本 n={len(gids)}）\n\n| 臂 | 加权κ (95%CI) | 准确率 | 宏AUC |\n|---|---|---|---|"); r2, bt2 = table(gids, a2); out += r2
    zs = "非生成式读出零样本（OpenJEV式，Qwen3.5-9B）"; gg = "生成式零样本（Qwen3.5-9B，生成理由后作答）"
    for a, b in ((zs, gg), ("任务LoRA 12k", zs), ("任务LoRA 12k", gg)):
        df = bt2[a] - bt2[b]; out.append(f"- κ({a}) − κ({b}) = {df.mean():+.3f}（{np.percentile(df,2.5):+.3f} ~ {np.percentile(df,97.5):+.3f}）")
# 学习曲线
Fc = ["age", "female", "weight_kg", "bmi", "inpatient", "sbp", "dbp", "hr", "htn", "chd", "dm", "cr_mgdl", "hb_gdl", "alb_gdl"]
m = pd.read_csv(f"{D}/asa_common_mover.csv"); tr = m[m.split == "train80"]; te = m[m.op_id.isin(common)].set_index("op_id").loc[common]; y = te.asa4.values.astype(int)
out.append("\n## C. 学习曲线（κ，共同样本；训练标签数 n）\n\n| n | 任务LoRA | XGBoost |\n|---|---|---|")
lk = {100: "任务LoRA 100", 300: "任务LoRA 300", 1000: "任务LoRA 1000", 3000: "任务LoRA 3000", 12000: "任务LoRA 12k"}
for n in (100, 300, 1000, 3000, 12000, len(tr)):
    s = tr.sample(n, random_state=20261017) if n < len(tr) else tr
    c = xgb.XGBClassifier(n_estimators=300, max_depth=4, learning_rate=0.05, subsample=0.8, n_jobs=16, tree_method="hist").fit(s[Fc], s.asa4 - 1)
    kx = kap(y, c.predict_proba(te[Fc]).argmax(1) + 1)
    kl = "—"; ks = []
    for tg in ("", "s11", "s12"):
        d = rd(f"pred_lora_curve{n}{tg}_mover_sub") if n < 12000 else (arms["任务LoRA 12k"] if tg == "" else None)
        if d is not None: ks.append(kap(y, d.loc[common][PC].values.argmax(1) + 1))
    if ks: kl = f"{np.mean(ks):.3f}（{len(ks)}次: " + ", ".join(f"{v:.3f}" for v in ks) + "）"
    out.append(f"| {n if n<len(tr) else str(n)+'（全部）'} | {kl} | {kx:.3f} |")
open(f"{R}/arms_eval_v2.md", "w").write("\n".join(out)); print("\n".join(out))
```
