## Supplementary material for "Non-generative readout with few-label calibration versus LoRA fine-tuning of a large language model for ASA physical status classification: accuracy, speed, cost and label efficiency in three data sources": Analysis code and aggregate results: 15_fewshot_knn.py

```
"""检索式少样本（训练无关）：对每个测试病例，在 MOVER train80 中按结构化特征 + 术式 TF-IDF 取 k 个最近的带标签病例作示例，
放入提示后做同一选项字母 logits 读出（不更新参数）。用法：15_fewshot_knn.py <k> <gpu> [mover_sub|mover_cal]"""
import os, sys, time, numpy as np, pandas as pd, torch
from sklearn.feature_extraction.text import TfidfVectorizer
from sklearn.preprocessing import StandardScaler
from sklearn.neighbors import NearestNeighbors
from scipy.sparse import hstack, csr_matrix
from transformers import AutoTokenizer, AutoModelForCausalLM
k, gpu = int(sys.argv[1]), int(sys.argv[2]); ds = sys.argv[3] if len(sys.argv) > 3 else "mover_sub"
D = os.path.expanduser("~/research/NE_ERAS_JEV/data/derived")
m = pd.read_csv(f"{D}/asa_common_mover.csv"); tr = m[m.split == "train80"].reset_index(drop=True)
if ds == "mover_sub": te = m[m.op_id.isin(pd.read_csv(f"{D}/mover_test_sub4000.csv").op_id)].reset_index(drop=True)
else: te = tr.sample(500, random_state=13).reset_index(drop=True); tr = tr[~tr.op_id.isin(te.op_id)].reset_index(drop=True)
F = ["age", "female", "weight_kg", "bmi", "inpatient", "sbp", "dbp", "hr", "htn", "chd", "dm", "cr_mgdl", "hb_gdl", "alb_gdl"]
med = tr[F].median(); sc = StandardScaler().fit(tr[F].fillna(med)); tf = TfidfVectorizer(min_df=3, ngram_range=(1, 2)).fit(tr.procedure.fillna(""))
def feat(d): return hstack([csr_matrix(np.nan_to_num(sc.transform(d[F].fillna(med)), nan=0) * 0.5), tf.transform(d.procedure.fillna("")) * 2.0]).tocsr()
nn = NearestNeighbors(n_neighbors=k, metric="cosine", algorithm="brute").fit(feat(tr)); _, nb = nn.kneighbors(feat(te))
path = "/mnt/c/backup/Qwen3.5-9B"; tok = AutoTokenizer.from_pretrained(path)
mod = AutoModelForCausalLM.from_pretrained(path, dtype=torch.float16, device_map={"": gpu}).eval()
a1 = [tok.encode(" " + c, add_special_tokens=False)[-1] for c in "ABCD"]; a2 = [tok.encode(c, add_special_tokens=False)[-1] for c in "ABCD"]; LAB = "ABCD"
H = ("You are an anesthesiologist. Based on the preoperative information, assign the ASA Physical Status class.\nOptions: A. ASA I  B. ASA II  C. ASA III  D. ASA IV-V\n"
     "Below are labelled examples from the same institution, followed by the case to classify.\n\n")
rows = []; t0 = time.time()
for i, r in enumerate(te.itertuples()):
    demos = "".join(f"Example {j+1}:\n{tr.text[q]}\nAnswer: {LAB[int(tr.asa4[q])-1]}\n\n" for j, q in enumerate(nb[i][::-1]))
    x = tok(H + demos + f"Case to classify:\n{r.text}\nAnswer:", return_tensors="pt").to(f"cuda:{gpu}")
    with torch.no_grad(): lg = mod(**x).logits[0, -1].float()
    p = torch.softmax(torch.maximum(lg[a1], lg[a2]), 0).cpu().numpy(); rows.append(dict(op_id=r.op_id, asa4=r.asa4, p1=p[0], p2=p[1], p3=p[2], p4=p[3]))
    if (i + 1) % 200 == 0: print(i + 1, f"{(time.time()-t0)/(i+1):.2f}s/例", flush=True)
pd.DataFrame(rows).to_csv(f"{D}/pred_knn{k}_{ds}.csv", index=False); print("完成", len(rows), f"{time.time()-t0:.0f}s")
```
