## Supplementary material for "Non-generative readout with few-label calibration versus LoRA fine-tuning of a large language model for ASA physical status classification: accuracy, speed, cost and label efficiency in three data sources": Analysis code and aggregate results: 17_build_t2_mover.py

```
"""MOVER 的 T2 输入：在 T1 共有结构化文本后追加该患者“诊断史”（patient_history 的 dx_name，去重，最多 30 条/700 字符）。
注意：patient_history 无日期，可能含术后录入的诊断（已知局限）。输出 NE_ERAS_JEV/data/derived/asa_t2_mover.csv（op_id, text2）。"""
import os, pandas as pd
D = os.path.expanduser("~/research/NE_ERAS_JEV/data/derived"); E = os.path.expanduser("~/research/NE_ERAS_JEV/data/raw/mover/extracted/epic/EPIC_EMR/EMR")
m = pd.read_csv(f"{D}/asa_common_mover.csv", usecols=["op_id", "text"]); pid = pd.read_csv(f"{D}/mover_pid_map.csv")
h = pd.read_csv(f"{E}/patient_history.csv", dtype=str).dropna(subset=["dx_name"]); h["dx_name"] = h.dx_name.str.replace(r"\s*\(CMS-HCC\)|\(\d+\.\d+\)", "", regex=True).str.strip()
g = h.drop_duplicates(["mrn", "dx_name"]).groupby("mrn").dx_name.apply(lambda s: "; ".join(list(s)[:30])[:700])
m = m.merge(pid, on="op_id", how="left"); m["dx"] = m.pid.map(g).fillna("none recorded")
m["text2"] = m.text + "\nDiagnoses on record (undated history): " + m.dx
m[["op_id", "text2"]].to_csv(f"{D}/asa_t2_mover.csv", index=False); print(len(m), (m.dx != "none recorded").mean().round(3)); print(m.text2.iloc[5])
```
