## Supplementary material for "Non-generative readout with few-label calibration versus LoRA fine-tuning of a large language model for ASA physical status classification: accuracy, speed, cost and label efficiency in three data sources": Analysis code and aggregate results: 18_latency.py

```
"""延迟/吞吐/显存基准（须在空闲 GPU 上单独运行）。每臂：批大小 1 的 p50/p95 延迟（100 例，预热 5 例）与批大小 8 的吞吐；峰值显存。
用法：18_latency.py <readout|lora|gen> <gpu> [lora_tag]。输出追加到 reports/latency.tsv"""
import os, sys, time, numpy as np, pandas as pd, torch
arm, gpu = sys.argv[1], int(sys.argv[2]); tag = sys.argv[3] if len(sys.argv) > 3 else None
D = os.path.expanduser("~/research/NE_ERAS_JEV/data/derived"); M = os.path.expanduser("~/research/Anes_JEV_vs_LoRA/models")
d = pd.read_csv(f"{D}/asa_common_mover.csv"); d = d[d.op_id.isin(pd.read_csv(f"{D}/mover_test_sub4000.csv").op_id)].sample(120, random_state=3).reset_index(drop=True)
texts = list(d.text); dev = f"cuda:{gpu}"; torch.cuda.set_device(gpu); torch.cuda.reset_peak_memory_stats()
P = ("You are an anesthesiologist. Based on the preoperative information below, assign the ASA Physical Status class.\n\n%s\n\nOptions:\nA. ASA I\nB. ASA II\nC. ASA III\nD. ASA IV-V\nAnswer:")
t_load = time.time()
if arm in ("readout", "readout4", "lora", "gen"):
    from transformers import AutoTokenizer, AutoModelForCausalLM, BitsAndBytesConfig
    path = "/mnt/c/backup/Qwen3.5-9B"; tok = AutoTokenizer.from_pretrained(path); tok.padding_side = "left"
    if arm == "lora":
        from peft import PeftModel
        bnb = BitsAndBytesConfig(load_in_4bit=True, bnb_4bit_quant_type="nf4", bnb_4bit_compute_dtype=torch.float16, bnb_4bit_use_double_quant=True)
        m = PeftModel.from_pretrained(AutoModelForCausalLM.from_pretrained(path, quantization_config=bnb, dtype=torch.float16, device_map={"": gpu}), f"{M}/{tag}").eval()
    elif arm == "readout4":   # 审稿修改：4-bit 量化读出（无适配器），与 4-bit LoRA 对照
        bnb = BitsAndBytesConfig(load_in_4bit=True, bnb_4bit_quant_type="nf4", bnb_4bit_compute_dtype=torch.float16, bnb_4bit_use_double_quant=True)
        m = AutoModelForCausalLM.from_pretrained(path, quantization_config=bnb, dtype=torch.float16, device_map={"": gpu}).eval()
    else: m = AutoModelForCausalLM.from_pretrained(path, dtype=torch.float16, device_map={"": gpu}).eval()
    def run(tt):
        if arm == "gen":
            msgs = [tok.apply_chat_template([{"role": "user", "content": P % t + "\nGive a brief rationale (at most 3 sentences), then 'Final answer: ASA <class>'."}], tokenize=False, add_generation_prompt=True, enable_thinking=False) for t in tt]
            x = tok(msgs, return_tensors="pt", padding=True).to(dev)
            with torch.no_grad(): m.generate(**x, max_new_tokens=140, do_sample=False)
        else:
            x = tok([P % t for t in tt], return_tensors="pt", padding=True).to(dev)
            with torch.no_grad(): m(**x, logits_to_keep=1)
else:
    sys.exit(f"unsupported arm: {arm}")

for t in texts[:5]: run([t])
lat = []
for t in texts[:100]:
    torch.cuda.synchronize(); t0 = time.time(); run([t]); torch.cuda.synchronize(); lat.append(time.time() - t0)
torch.cuda.synchronize(); t0 = time.time()
for i in range(0, 96, 8): run(texts[i:i + 8])
torch.cuda.synchronize(); thr = 96 / (time.time() - t0)
line = f"{arm}\t{tag or ''}\tp50={np.percentile(lat,50)*1000:.0f}ms\tp95={np.percentile(lat,95)*1000:.0f}ms\tbatch8={thr:.1f}例/s\t峰值显存={torch.cuda.max_memory_allocated()/1e9:.1f}GB\t加载={load_s:.0f}s"
print(line); open(os.path.expanduser("~/research/Anes_JEV_vs_LoRA/analysis/reports/latency.tsv"), "a").write(line + "\n")
```
