## Supplementary material for "Non-generative readout with few-label calibration versus LoRA fine-tuning of a large language model for ASA physical status classification: accuracy, speed, cost and label efficiency in three data sources": Analysis code and aggregate results: 20_eval_v3.py

```
"""v3 评估（仅 MOVER，患者级测试子集 sub4000）。输出 reports/arms_eval_v3.md
表 A：主要臂；表 B：学习曲线（均值[范围]，种子数）；表 C：配对对比（cluster bootstrap by patient，seed0 预测）。"""
import os, glob, numpy as np, pandas as pd, xgboost as xgb
from sklearn.metrics import cohen_kappa_score, roc_auc_score
from sklearn.linear_model import LogisticRegression
D = os.path.expanduser("~/research/NE_ERAS_JEV/data/derived"); R = os.path.expanduser("~/research/Anes_JEV_vs_LoRA/analysis/reports")
sub = set(pd.read_csv(f"{D}/mover_test_sub4000.csv").op_id); pid = pd.read_csv(f"{D}/mover_pid_map.csv").set_index("op_id").pid; PC = ["p1", "p2", "p3", "p4"]
def rd(f):
    p = f"{D}/{f}.csv"; return pd.read_csv(p).set_index("op_id") if os.path.exists(p) else None
def kap(y, p): return cohen_kappa_score(y, p, weights="quadratic")
def cut_cal(c, t):
    ec = (c[PC].values * np.arange(1, 5)).sum(1); yc = c.asa4.astype(int).values; cuts = np.array([1.5, 2.5, 3.5]); grid = np.linspace(1.0, 4.0, 121)
    for _ in range(3):
        for j in range(3): cuts[j] = max(grid, key=lambda g: kap(yc, 1 + (ec[:, None] > np.where(np.arange(3) == j, g, cuts)).sum(1)))
    et = (t[PC].values * np.arange(1, 5)).sum(1); pred = 1 + (et[:, None] > cuts).sum(1); q = t.copy(); q[PC] = np.eye(4)[pred - 1] * 0.97 + 0.01; return q
A = {}   # 主表臂（seed0）
A["读出零样本 9B（T1）"] = rd("pred_llm_mover_test")
for nm, key, suf in (("读出零样本 9B（T2 含诊断史）", "llm", "_t2"), ("读出零样本 Qwen3.5-27B（T1）", "llm", ""), ("读出零样本 Qwen3.8-27B（T1）", "llm", "")):
    pass
A["读出零样本 9B（T2 含诊断史）"] = rd("pred_llm_mover_sub_t2")
A["读出零样本 Qwen3.5-27B（T1）"] = rd("pred_llm_mover_sub_q35_27b"); A["读出零样本 Qwen3.8-27B（T1）"] = rd("pred_llm_mover_sub_q38_27b")
A["检索式少样本 kNN-8"] = rd("pred_knn8_mover_sub")
for nm, tst, cal in (("读出 9B（T1）+1000标签切点校准", "pred_llm_mover_test", "pred_llm_mover_cal"), ("读出 9B（T2）+1000标签切点校准", "pred_llm_mover_sub_t2", "pred_llm_mover_cal_t2"),
                     ("读出 Qwen3.5-27B + 切点校准", "pred_llm_mover_sub_q35_27b", "pred_llm_mover_cal_q35_27b"), ("读出 Qwen3.8-27B + 切点校准", "pred_llm_mover_sub_q38_27b", "pred_llm_mover_cal_q38_27b")):
    t, c = rd(tst), rd(cal)
    if t is not None and c is not None and len(c) >= 900: A[nm] = cut_cal(c, t.loc[t.index.isin(sub)])
A["XGBoost（全训练集）"] = rd("pred_xgb_mover_test")
def seeds(pattern_fn, tags): return [rd(pattern_fn(t)) for t in tags if rd(pattern_fn(t)) is not None]
CURVE = {"读出+LoRA（T1）": lambda n, s: f"pred_lora_curve{n}{s}_mover_sub"}
SD = {"": "", "s11": "11", "s12": "12"}
def curve_preds(name, n):
    out = []
    for sfx in ("", "s11", "s12"):
        f = CURVE[name](n, sfx) if name.startswith("读出") else CURVE[name](n, SD[sfx] if sfx else "")
        d = rd(f)
        if d is not None: out.append(d)
    return out
for n in (12000,):
    pass
A["读出+LoRA 12k（T1）"] = rd("pred_lora_mover12k_mover_test")
for nm in ("读出+LoRA（T1）",):
    for n in (1000, 3000):
        p = curve_preds(nm, n)
        if p: A[f"{nm} n={n}"] = p[0]
for n in (1000, 3000):
    d = rd(f"pred_lora_t2lora{n}_mover_sub")
    if d is not None: A[f"读出+LoRA（T2）n={n}"] = d
for tg, nm in (("q35", "Qwen3.5-27B"), ("q38", "Qwen3.8-27B")):
    for n in (1000, 3000):
        d = rd(f"pred_lora_L27{tg}_{n}_mover_sub")
        if d is not None: A[f"读出+LoRA {nm}（T1）n={n}"] = d
A = {k: v for k, v in A.items() if v is not None}
common = sorted(set.intersection(*[set(v.index) & sub for v in A.values()]))
rng = np.random.default_rng(20261018); B = 1000; pats = pid.loc[common].values; up = np.unique(pats); idx = {p: np.where(pats == p)[0] for p in up}
boots = [np.concatenate([idx[p] for p in rng.choice(up, len(up))]) for _ in range(B)]
y = A[next(iter(A))].loc[common].asa4.values.astype(int)
out = [f"# v3 评估（MOVER 患者级子集；共同样本 n={len(common)}，患者 {len(up)}；cluster bootstrap B={B}）\n", "## A. 各臂（seed0 预测；有 AUC 的才报）\n", "| 臂 | κ (95%CI) | 准确率 | 宏AUC |", "|---|---|---|---|"]
KB = {}
for nm, d in A.items():
    d = d.loc[common]; pr = d[PC].values.argmax(1) + 1; kb = np.array([kap(y[s], pr[s]) for s in boots]); KB[nm] = kb
    isH = ((d[PC].values > 0.9).sum(1) == 1).all()   # 切点校准臂概率为伪独热
    auc = "—" if isH else f"{np.mean([roc_auc_score((y == k + 1).astype(int), d[PC[k]].values) for k in range(4)]):.3f}"
    out.append(f"| {nm} | {kap(y,pr):.3f} ({np.percentile(kb,2.5):.3f}–{np.percentile(kb,97.5):.3f}) | {(pr==y).mean():.3f} | {auc} |")
# B 学习曲线
Fc = ["age", "female", "weight_kg", "bmi", "inpatient", "sbp", "dbp", "hr", "htn", "chd", "dm", "cr_mgdl", "hb_gdl", "alb_gdl"]
m = pd.read_csv(f"{D}/asa_common_mover.csv"); tr = m[m.split == "train80"]; te = m[m.op_id.isin(common)].set_index("op_id").loc[common]
out += ["\n## B. 学习曲线：κ 均值 [范围]（种子数）\n", "| 训练标签数 | 读出+LoRA（T1） | XGBoost |", "|---|---|---|"]
def fmt(ps):
    ks = [kap(y, p.loc[common][PC].values.argmax(1) + 1) for p in ps]
    return "—" if not ks else f"{np.mean(ks):.3f} [{min(ks):.3f}–{max(ks):.3f}]（{len(ks)}）"
for n in (100, 300, 1000, 3000, 12000):
    lo = curve_preds("读出+LoRA（T1）", n) if n < 12000 else ([rd("pred_lora_mover12k_mover_test").loc[lambda x: x.index.isin(sub)]] + [d for d in [rd("pred_lora_curve12000s11_mover_sub"), rd("pred_lora_curve12000s12_mover_sub")] if d is not None])
    s = tr.sample(n, random_state=20261017); c = xgb.XGBClassifier(n_estimators=300, max_depth=4, learning_rate=0.05, subsample=0.8, n_jobs=16, tree_method="hist").fit(s[Fc], s.asa4 - 1)
    out.append(f"| {n} | {fmt(lo)} | {kap(y, c.predict_proba(te[Fc]).argmax(1) + 1):.3f} |")
# C 配对对比
out += ["\n## C. 配对对比（κ 差，cluster bootstrap 95%CI；seed0）\n"]
def contrast(a, b, note=""):
    if a in KB and b in KB: df = KB[a] - KB[b]; out.append(f"- {a} − {b}：{df.mean():+.3f}（{np.percentile(df,2.5):+.3f} ~ {np.percentile(df,97.5):+.3f}）{note}")
contrast("读出+LoRA 12k（T1）", "读出零样本 9B（T1）"); contrast("读出 9B（T1）+1000标签切点校准", "读出+LoRA（T1） n=1000", "（非劣效界值 δ=0.05：差的下界需 > −0.05）")
contrast("读出 9B（T1）+1000标签切点校准", "读出+LoRA（T1） n=3000"); 
contrast("读出+LoRA Qwen3.5-27B（T1）n=1000", "读出+LoRA（T1） n=1000"); contrast("读出+LoRA Qwen3.5-27B（T1）n=3000", "读出+LoRA（T1） n=3000"); contrast("读出零样本 9B（T2 含诊断史）", "读出零样本 9B（T1）"); contrast("读出+LoRA（T2）n=3000", "读出+LoRA（T1） n=3000"); contrast("读出 9B（T2）+1000标签切点校准", "读出+LoRA（T2）n=1000", "（T2，非劣效 δ=0.05）"); contrast("读出+LoRA（T2）n=3000", "读出 9B（T2）+1000标签切点校准"); contrast("读出零样本 Qwen3.8-27B（T1）", "读出零样本 9B（T1）"); contrast("读出零样本 Qwen3.5-27B（T1）", "读出零样本 9B（T1）")
open(f"{R}/arms_eval_v3.md", "w").write("\n".join(out)); print("\n".join(out))
```
