## Supplementary material for "Non-generative readout with few-label calibration versus LoRA fine-tuning of a large language model for ASA physical status classification: accuracy, speed, cost and label efficiency in three data sources": Analysis code and aggregate results: 22_eval_local.py

```
"""本院评估（按冻结方案）：val（2025–26）为检验集；dev 仅作训练/校准。参考标准=本院麻醉记录 ASA。输出 reports/local_eval.md"""
import os, numpy as np, pandas as pd, xgboost as xgb
from sklearn.metrics import cohen_kappa_score, roc_auc_score
D = os.path.expanduser("~/research/NE_ERAS_JEV/data/derived"); R = os.path.expanduser("~/research/Anes_JEV_vs_LoRA/analysis/reports"); PC = ["p1", "p2", "p3", "p4"]
c = pd.read_csv(f"{D}/asa_common_local.csv"); raw = pd.read_csv(f"{D}/asa_dataset_v1.csv", usecols=["operation_id", "patient_sn"]).rename(columns={"operation_id": "op_id"})
c = c.merge(raw, on="op_id"); val = set(c[c.split == "val"].op_id); pid = c.set_index("op_id").patient_sn
def rd(f):
    p = f"{D}/{f}.csv"; return pd.read_csv(p).set_index("op_id") if os.path.exists(p) else None
def kap(y, p): return cohen_kappa_score(y, p, weights="quadratic")
def v(d): return None if d is None else d[d.index.isin(val)]
def cut_cal(cal, t):
    ec = (cal[PC].values * np.arange(1, 5)).sum(1); yc = cal.asa4.astype(int).values; cuts = np.array([1.5, 2.5, 3.5]); grid = np.linspace(1.0, 4.0, 121)
    for _ in range(3):
        for j in range(3): cuts[j] = max(grid, key=lambda g: kap(yc, 1 + (ec[:, None] > np.where(np.arange(3) == j, g, cuts)).sum(1)))
    et = (t[PC].values * np.arange(1, 5)).sum(1); pr = 1 + (et[:, None] > cuts).sum(1); q = t.copy(); q[PC] = np.eye(4)[pr - 1] * 0.97 + 0.01; return q, cuts
A = {}; notes = []
A["读出零样本 T1"] = v(rd("pred_llm_local")); A["读出零样本 T2"] = v(rd("pred_llm_local_t2"))
dev_ids = c[c.split == "dev"].op_id.sample(1000, random_state=11)
for nm, f in (("T1", "pred_llm_local"), ("T2", "pred_llm_local_t2")):
    full = rd(f)
    if full is not None:
        q, cuts = cut_cal(full.loc[dev_ids], v(full)); A[f"读出零样本 {nm} + 1000本院标签切点校准"] = q; notes.append(f"{nm} 切点 {np.round(cuts,2).tolist()}")
for n, tg in ((100, "100"), (300, "300"), (1000, "1000"), (4326, "4326")):
    d = rd(f"pred_lora_LOC{tg}s20261017_local_val")
    if d is not None: A[f"本院 LoRA T1 n={n}"] = d
for n in (1000, 4326):
    d = rd(f"pred_lora_LOCT2_{n}_local_val")
    if d is not None: A[f"本院 LoRA T2 n={n}"] = d
A["MOVER-LoRA 12k 迁移"] = rd("pred_lora_mover12k_local_val"); A["XGBoost（MOVER 训练）"] = v(rd("pred_xgb_local"))
Fc = ["age", "female", "weight_kg", "bmi", "inpatient", "sbp", "dbp", "hr", "htn", "chd", "dm", "cr_mgdl", "hb_gdl", "alb_gdl"]
dv = c[c.split == "dev"]; te = c[c.split == "val"].set_index("op_id")
xg = xgb.XGBClassifier(n_estimators=300, max_depth=4, learning_rate=0.05, subsample=0.8, n_jobs=16, tree_method="hist").fit(dv[Fc], dv.asa4 - 1)
q = te[["asa4"]].copy(); q[PC] = xg.predict_proba(te[Fc]); A["XGBoost（本院 dev 训练）"] = q
A = {k: x for k, x in A.items() if x is not None}
ids = sorted(set.intersection(*[set(x.index) for x in A.values()])); rng = np.random.default_rng(20261019)
pats = pid.loc[ids].values; up = np.unique(pats); idx = {p: np.where(pats == p)[0] for p in up}; boots = [np.concatenate([idx[p] for p in rng.choice(up, len(up))]) for _ in range(1000)]
y = A[next(iter(A))].loc[ids].asa4.values.astype(int)
out = [f"# 本院评估（val 2025–26；共同样本 n={len(ids)}，患者 {len(up)}；参考=本院麻醉记录 ASA；cluster bootstrap B=1000）\n", "；".join(notes) + "\n", "| 臂 | κ (95%CI) | 准确率 | 宏AUC |", "|---|---|---|---|"]; KB = {}
for nm, d in A.items():
    d = d.loc[ids]; pr = d[PC].values.argmax(1) + 1; kb = np.array([kap(y[s], pr[s]) for s in boots]); KB[nm] = kb
    hard = ((d[PC].values > 0.9).sum(1) == 1).all(); auc = "—" if hard else f"{np.mean([roc_auc_score((y == k + 1).astype(int), d[PC[k]].values) for k in range(4)]):.3f}"
    out.append(f"| {nm} | {kap(y,pr):.3f} ({np.percentile(kb,2.5):.3f}–{np.percentile(kb,97.5):.3f}) | {(pr==y).mean():.3f} | {auc} |")
# 学习曲线（多种子）
out += ["\n## 本院 LoRA T1 学习曲线：κ 均值 [范围]（种子数；val 全部 2,753）\n", "| n | 本院 LoRA | XGBoost（本院） |", "|---|---|---|"]
vall = sorted(val); yv = c.set_index("op_id").loc[vall].asa4.values.astype(int)
for n in (100, 300, 1000, 4326):
    ks = []
    for sd in ("20261017", "11", "12"):
        d = rd(f"pred_lora_LOC{n}s{sd}_local_val")
        if d is not None: ks.append(kap(yv, d.loc[vall][PC].values.argmax(1) + 1))
    s = dv.sample(n, random_state=20261017) if n < len(dv) else dv
    xc = xgb.XGBClassifier(n_estimators=300, max_depth=4, learning_rate=0.05, subsample=0.8, n_jobs=16, tree_method="hist").fit(s[Fc], s.asa4 - 1)
    out.append(f"| {n} | " + ("—" if not ks else f"{np.mean(ks):.3f} [{min(ks):.3f}–{max(ks):.3f}]（{len(ks)}）") + f" | {kap(yv, xc.predict_proba(te.loc[vall][Fc]).argmax(1)+1):.3f} |")
# 生成式（val 1000 子集）
g = rd("pred_gen_local_val")
if g is not None:
    gc = g.dropna(subset=["pred"]); gi = sorted(set(gc.index) & set(ids)); yy = gc.loc[gi].asa4.values.astype(int)
    out.append(f"\n## 生成式零样本（val 子集，可解析 {len(gc)}/{len(g)}，n={len(gi)}）\n")
    out.append(f"- 生成式 κ {kap(yy, gc.loc[gi].pred.values.astype(int)):.3f}；同子集读出零样本 T1 κ {kap(yy, A['读出零样本 T1'].loc[gi][PC].values.argmax(1)+1):.3f}")
out += ["\n## 配对对比（κ 差，cluster bootstrap 95%CI；seed0）\n"]
def contrast(a, b, note=""):
    if a in KB and b in KB: df = KB[a] - KB[b]; out.append(f"- {a} − {b}：{df.mean():+.3f}（{np.percentile(df,2.5):+.3f} ~ {np.percentile(df,97.5):+.3f}）{note}")
contrast("读出零样本 T1 + 1000本院标签切点校准", "本院 LoRA T1 n=1000", "（H2，δ=0.05）"); contrast("本院 LoRA T1 n=4326", "读出零样本 T1 + 1000本院标签切点校准", "（H3）")
contrast("本院 LoRA T1 n=4326", "读出零样本 T1"); contrast("本院 LoRA T1 n=1000", "MOVER-LoRA 12k 迁移")
contrast("读出零样本 T2", "读出零样本 T1"); contrast("本院 LoRA T2 n=4326", "本院 LoRA T1 n=4326")
open(f"{R}/local_eval.md", "w").write("\n".join(out)); print("\n".join(out))
```
