## Supplementary material for "Non-generative readout with few-label calibration versus LoRA fine-tuning of a large language model for ASA physical status classification: accuracy, speed, cost and label efficiency in three data sources": Analysis code and aggregate results: 24_option_order.py

```
"""稳健性：选项顺序置换。零样本读出（llm）在 3 种顺序（原顺序 / 反序 / 循环移 2）下的概率，映射回类别后计算反转率、各顺序 κ 与三顺序平均（去偏）κ。
数据：MOVER sub4000 前 N 台 与 本院 val 随机 N 台（种子 5）。用法：24_option_order.py <llm> <gpu> <N>。输出 pred_perm_<arm>_<ds>.csv（每例 3 组概率）。"""
import os, sys, time, numpy as np, pandas as pd, torch
arm, gpu, N = sys.argv[1], int(sys.argv[2]), int(sys.argv[3]); VAR = sys.argv[4] if len(sys.argv) > 4 else "perm"; D = os.path.expanduser("~/research/NE_ERAS_JEV/data/derived")
PERMS = {"orig": [0, 1, 2, 3], "rev": [3, 2, 1, 0], "shift2": [2, 3, 0, 1]} if VAR == "perm" else {"orig": [0, 1, 2, 3]}   # 位置 i 放第 PERMS[i] 个类别
NAMES = ["ASA I", "ASA II", "ASA III", "ASA IV-V"]
mv = pd.read_csv(f"{D}/asa_common_mover.csv"); mv = mv[mv.op_id.isin(pd.read_csv(f"{D}/mover_test_sub4000.csv").op_id.tolist()[:N])]
lc = pd.read_csv(f"{D}/asa_common_local.csv"); lc = lc[lc.split == "val"].sample(N, random_state=5)
if arm == "llm":
    from transformers import AutoTokenizer, AutoModelForCausalLM
    path = "/mnt/c/backup/Qwen3.5-9B"; tok = AutoTokenizer.from_pretrained(path); m = AutoModelForCausalLM.from_pretrained(path, dtype=torch.float16, device_map={"": gpu}).eval()
    LAB = "1234" if VAR == "num" else "ABCD"
    a1 = [tok.encode(" " + c, add_special_tokens=False)[-1] for c in LAB]; a2 = [tok.encode(c, add_special_tokens=False)[-1] for c in LAB]
    def infer(t, perm):
        opts = "\n".join(f"{LAB[i]}. {NAMES[perm[i]]}" for i in range(4))
        q = f"You are an anesthesiologist. Based on the preoperative information below, assign the ASA Physical Status class.\n\n{t}\n\nOptions:\n{opts}\nAnswer:"
        x = tok(q, return_tensors="pt").to(f"cuda:{gpu}")
        with torch.no_grad(): lg = m(**x).logits[0, -1].float()
        p = torch.softmax(torch.maximum(lg[a1], lg[a2]), 0).cpu().numpy(); out = np.zeros(4); out[perm] = p; return out   # 位置概率 → 类别概率
else:
    sys.exit(f"unsupported arm: {arm}")

for ds, d in (("mover", mv), ("local", lc)):
    rows = []; t0 = time.time()
    for i, r in enumerate(d.itertuples()):
        row = dict(op_id=r.op_id, asa4=r.asa4)
        for nm, perm in PERMS.items():
            p = infer(r.text, perm); row.update({f"{nm}_p{k+1}": p[k] for k in range(4)})
        rows.append(row)
        if (i + 1) % 250 == 0: print(ds, i + 1, f"{(time.time()-t0)/(i+1):.2f}s/例", flush=True)
    pd.DataFrame(rows).to_csv(f"{D}/pred_perm_{arm}{'_num' if VAR == 'num' else ''}_{ds}.csv", index=False)
print("完成")
```
