## Supplementary material for "Non-generative readout with few-label calibration versus LoRA fine-tuning of a large language model for ASA physical status classification: accuracy, speed, cost and label efficiency in three data sources": Analysis code and aggregate results: 26_perm_eval.py

```
"""选项顺序置换稳健性评估：反转率（argmax 类别与原顺序不同的比例）、各顺序 κ、三顺序平均概率（去偏）κ。输出 reports/option_order_robustness.md"""
import os, numpy as np, pandas as pd
from sklearn.metrics import cohen_kappa_score as ck
D = os.path.expanduser("~/research/NE_ERAS_JEV/data/derived"); R = os.path.expanduser("~/research/Anes_JEV_vs_LoRA/analysis/reports"); O = ["orig", "rev", "shift2"]
def P(d, o): return d[[f"{o}_p{k}" for k in range(1, 5)]].values
out = ["# 选项顺序置换稳健性（零样本；顺序：orig=I,II,III,IV-V；rev=反序；shift2=循环移 2）\n", "| 臂 | 数据 | n | κ orig | κ rev | κ shift2 | 反转率 rev | 反转率 shift2 | 平均概率（去偏）κ | 平均最大概率 orig |", "|---|---|---|---|---|---|---|---|---|---|"]
for arm, nm in (("llm", "读出零样本 9B"),):
    for ds, dn in (("mover", "MOVER sub"), ("local", "本院 val")):
        f = f"{D}/pred_perm_{arm}_{ds}.csv"
        if not os.path.exists(f): continue
        d = pd.read_csv(f); y = d.asa4.astype(int).values; pr = {o: P(d, o).argmax(1) + 1 for o in O}; avg = sum(P(d, o) for o in O) / 3
        k = {o: ck(y, pr[o], weights="quadratic") for o in O}
        out.append(f"| {nm} | {dn} | {len(d)} | {k['orig']:.3f} | {k['rev']:.3f} | {k['shift2']:.3f} | {(pr['rev']!=pr['orig']).mean():.3f} | {(pr['shift2']!=pr['orig']).mean():.3f} | {ck(y, avg.argmax(1)+1, weights='quadratic'):.3f} | {P(d,'orig').max(1).mean():.3f} |")
open(f"{R}/option_order_robustness.md", "w").write("\n".join(out)); print("\n".join(out))
```
