## Supplementary material for "Non-generative readout with few-label calibration versus LoRA fine-tuning of a large language model for ASA physical status classification: accuracy, speed, cost and label efficiency in three data sources": Analysis code and aggregate results: 30_openjev.py

```
"""真正的 OpenJev（GitHub30/OpenJev，MIT，commit aae11e1…；与 TypeSafe Jev 的 OpenAPI 线协议兼容）在 Qwen3.5-9B 上运行 ASA 分级。
同一请求含两个问题：choice（四个命名选项，主）与 score（四级有序，次）；提示词与解码完全由 OpenJev 默认配置产生（系统提示、编号选项、聊天模板）。
记录候选对数概率（温度 1，不校准）；校准另做。用法：30_openjev.py <mover_sub|mover_cal|local|local_t2> <gpu>。输出 pred_ojev_<ds>.csv，可断点续跑。"""
import os, sys, time, numpy as np, pandas as pd, torch
from openjev import SystemOneEngine, load_backend
from openjev.schema import SystemOneRequest
from openjev.prompting import compile_request
ds, gpu = sys.argv[1], int(sys.argv[2]); FROZEN = os.environ.get("FROZEN") == "1"; D = os.path.expanduser("~/research/NE_ERAS_JEV/data/derived")
if ds.startswith("local"):
    d = pd.read_csv(f"{D}/asa_common_local.csv")
    if ds == "local_t2":
        raw = pd.read_csv(f"{D}/asa_dataset_v1.csv", usecols=["operation_id", "text"]).rename(columns={"operation_id": "op_id", "text": "raw"}); d = d.merge(raw, on="op_id", how="left"); d["text"] = d.raw
elif ds == "vitaldb": d = pd.read_csv(f"{D}/asa_common_vitaldb.csv"); d = d[d.op_id.isin(pd.read_csv(f"{D}/vitaldb_sub2000.csv").op_id)]
else:
    d = pd.read_csv(f"{D}/asa_common_mover.csv")
    d = d[d.op_id.isin(pd.read_csv(f"{D}/mover_test_sub4000.csv").op_id)] if ds == "mover_sub" else d[d.split == "train80"].sample(1000, random_state=11)
d = d.reset_index(drop=True)
if os.environ.get("NMAX"): d = d.head(int(os.environ["NMAX"]))
out = f"{D}/pred_ojev{'F' if FROZEN else ''}_{ds}{os.environ.get('MTAG', '')}.csv"
done = pd.read_csv(out) if os.path.exists(out) else pd.DataFrame(columns=["op_id"]); todo = d[~d.op_id.isin(done.op_id)]; print(ds, "总", len(d), "待做", len(todo), flush=True)
LV = ["ASA I: A normal healthy patient", "ASA II: A patient with mild systemic disease", "ASA III: A patient with severe systemic disease",
      "ASA IV-V: A patient with severe systemic disease that is a constant threat to life, or a moribund patient"]
Q = {"choice": {"type": "choice", "instructions": "What is the ASA Physical Status class of this patient?", "criteria": {"ASA I": LV[0][7:], "ASA II": LV[1][8:], "ASA III": LV[2][9:], "ASA IV-V": LV[3][10:]}},
     "score": {"type": "score", "instructions": "What is the ASA Physical Status class of this patient? (0 = ASA I ... 3 = ASA IV-V)", "criteria": LV}}
MODEL = os.environ.get("MODEL", "/mnt/c/backup/Qwen3.5-9B"); kw = {}
if os.environ.get("QUANT") == "4":
    from transformers import BitsAndBytesConfig
    kw = dict(quantization_config=BitsAndBytesConfig(load_in_4bit=True, bnb_4bit_quant_type="nf4", bnb_4bit_compute_dtype=torch.float16, bnb_4bit_use_double_quant=True), device_map={"": gpu})
engine = SystemOneEngine(backend=load_backend(MODEL, device=f"cuda:{gpu}", dtype=torch.float16, **kw), model_name="openjev")
if FROZEN:
    import json
    ns = {}; exec("SYS =" + open(os.path.expanduser("~/research/Anes_JEV_vs_LoRA/analysis/scripts/33_openjev_tune.py")).read().split("SYS =", 1)[1].split("sm = lambda")[0], ns)
    CH = json.load(open(os.path.expanduser("~/research/Anes_JEV_vs_LoRA/analysis/reports/openjev_tuning_choice.json")))["selected"]; QT = CH["qtype"]
    from openjev.prompting import PromptConfig
    engine.prompt_config = PromptConfig() if ns["SYS"][CH["system"]] is None else PromptConfig(system_prompt=ns["SYS"][CH["system"]])
    Q = {QT: ns["questions"](ns["SHORT"] if CH["desc"] == "short" else ns["OFFIC"])[QT]}
rows = []; t0 = time.time()
def flush():
    global done, rows
    if rows: done = pd.concat([done, pd.DataFrame(rows)]); done.to_csv(out, index=False); rows = []
for i, r in enumerate(todo.itertuples()):
    raws, _ = engine.evaluate_raw(SystemOneRequest.model_validate({"state": r.text, "questions": Q}))
    by = {x.decision.name: x.logprobs for x in raws}
    if FROZEN: rows.append(dict(op_id=r.op_id, asa4=r.asa4, qtype=QT, **{f"c{k}": by[QT][k] for k in range(4)}))
    else: rows.append(dict(op_id=r.op_id, asa4=r.asa4, **{f"c{k}": by["choice"][k] for k in range(4)}, **{f"s{k}": by["score"][k] for k in range(4)}))
    if i == 0: print("提示词示例：\n", compile_request(r.text, SystemOneRequest.model_validate({"state": r.text, "questions": Q}).questions, engine.prompt_config)[0].prompt.user[:900], flush=True)
    if (i + 1) % 250 == 0: flush(); print(i + 1, f"{(time.time()-t0)/(i+1):.2f}s/例", flush=True)
flush(); print("完成", len(done))
```
