## Supplementary material for "Non-generative readout with few-label calibration versus LoRA fine-tuning of a large language model for ASA physical status classification: accuracy, speed, cost and label efficiency in three data sources": Analysis code and aggregate results: 31_make_openjev_jsonl.py

```
"""OpenJev 训练 JSONL（choice 题型，gold=选项名）。与任务 LoRA 同一训练子集（种子 20261017）。输出 NE_ERAS_JEV/data/derived/ojev_train_<src>_<n>.jsonl"""
import os, sys, json, pandas as pd
D = os.path.expanduser("~/research/NE_ERAS_JEV/data/derived")
LV = {"ASA I": "A normal healthy patient", "ASA II": "A patient with mild systemic disease", "ASA III": "A patient with severe systemic disease",
      "ASA IV-V": "A patient with severe systemic disease that is a constant threat to life, or a moribund patient"}
NM = {1: "ASA I", 2: "ASA II", 3: "ASA III", 4: "ASA IV-V"}
Q = {"choice": {"type": "choice", "instructions": "What is the ASA Physical Status class of this patient?", "criteria": LV}}
for src, n in (("mover", 1000), ("mover", 3000), ("local", 1000), ("local", 4326)):
    if src == "mover": d = pd.read_csv(f"{D}/asa_common_mover.csv"); d = d[d.split == "train80"].sample(n, random_state=20261017)
    else: d = pd.read_csv(f"{D}/asa_common_local.csv"); d = d[d.split == "dev"]; d = d.sample(n, random_state=20261017) if n < len(d) else d.sample(frac=1, random_state=20261017)
    p = f"{D}/ojev_train_{src}_{n}.jsonl"
    with open(p, "w", encoding="utf-8") as f:
        for r in d.itertuples(): f.write(json.dumps({"state": r.text, "questions": Q, "gold": {"choice": NM[int(r.asa4)]}}, ensure_ascii=False) + "\n")
    os.chmod(p, 0o600); print(p, len(d))
```
