## Supplementary material for "Non-generative readout with few-label calibration versus LoRA fine-tuning of a large language model for ASA physical status classification: accuracy, speed, cost and label efficiency in three data sources": Analysis code and aggregate results: 32_ojev_eval_quick.py

```
import os, sys, numpy as np, pandas as pd
from sklearn.metrics import cohen_kappa_score as ck, roc_auc_score
D=os.path.expanduser("~/research/NE_ERAS_JEV/data/derived"); f=sys.argv[1]; ref=sys.argv[2] if len(sys.argv)>2 else None
d=pd.read_csv(f"{D}/{f}.csv"); y=d.asa4.astype(int).values
sm=lambda a:(lambda e:e/e.sum(1,keepdims=True))(np.exp(a-a.max(1,keepdims=True)))
C=sm(d[[f"c{k}" for k in range(4)]].values); S=sm(d[[f"s{k}" for k in range(4)]].values)
def rep(nm,P,pr=None):
    pr=P.argmax(1)+1 if pr is None else pr
    auc=np.mean([roc_auc_score((y==k+1).astype(int),P[:,k]) for k in range(4)])
    print(f"{nm}: n={len(y)} κ={ck(y,pr,weights='quadratic'):.3f} acc={(pr==y).mean():.3f} AUC={auc:.3f} 分布={np.bincount(pr-1,minlength=4).tolist()}")
rep("OpenJev choice",C); rep("OpenJev score(argmax)",S); es=(S*np.arange(1,5)).sum(1); rep("OpenJev score(期望四舍五入)",S,np.clip(np.rint(es),1,4).astype(int))
if ref:
    r=pd.read_csv(f"{D}/{ref}.csv").set_index("op_id").loc[d.op_id]; P=r[["p1","p2","p3","p4"]].values; print("自写读出(同一批):","κ",round(ck(y,P.argmax(1)+1,weights='quadratic'),3))
```
