## Supplementary material for "Non-generative readout with few-label calibration versus LoRA fine-tuning of a large language model for ASA physical status classification: accuracy, speed, cost and label efficiency in three data sources": Analysis code and aggregate results: 33_openjev_tune.py

```
"""OpenJev 配置调参（仅 MOVER 调参集：train80 随机 800 台，种子 17；与校准集 seed11、测试子集、本院均不重叠）。
4 组提示（通用/麻醉场景系统提示 × 一句话/官方举例选项描述），每组同一请求同时产出 choice 与 score，共 8 个配置。
选择准则：2 折交叉的 3 切点校准后二次加权 κ；并报未校准 κ、宏 AUC。用法：33_openjev_tune.py <gpu> [n]。输出 reports/openjev_tuning.md 与 reports/openjev_tuning_choice.json"""
import os, sys, json, itertools, numpy as np, pandas as pd, torch
from sklearn.metrics import cohen_kappa_score as ck, roc_auc_score
from openjev import SystemOneEngine, load_backend
from openjev.prompting import PromptConfig
from openjev.schema import SystemOneRequest
gpu = int(sys.argv[1]); N = int(sys.argv[2]) if len(sys.argv) > 2 else 800; D = os.path.expanduser("~/research/NE_ERAS_JEV/data/derived"); R = os.path.expanduser("~/research/Anes_JEV_vs_LoRA/analysis/reports")
m = pd.read_csv(f"{D}/asa_common_mover.csv"); tr = m[m.split == "train80"]; cal = set(tr.sample(1000, random_state=11).op_id)
d = tr[~tr.op_id.isin(cal)].sample(N, random_state=17).reset_index(drop=True)
SYS = {"generic": None, "anes": "You are an experienced anesthesiologist acting as a System One decision model. You are shown a STATE (preoperative information) and a QUESTION. Select exactly one option by its label only. Be calibrated: judge the ASA Physical Status from documented systemic disease and its functional impact, not from the type of surgery alone."}
SHORT = ["A normal healthy patient", "A patient with mild systemic disease", "A patient with severe systemic disease", "A patient with severe systemic disease that is a constant threat to life, or a moribund patient"]
OFFIC = ["A normal healthy patient. Healthy, non-smoking, no or minimal alcohol use.",
         "Mild systemic disease without substantive functional limitation. Examples: current smoker, social alcohol drinker, pregnancy, obesity (30<BMI<40), well-controlled DM/HTN, mild lung disease.",
         "Severe systemic disease with substantive functional limitation. Examples: poorly controlled DM or HTN, COPD, morbid obesity (BMI>=40), active hepatitis, alcohol dependence, implanted pacemaker, moderately reduced EF, ESRD on regular dialysis, history (>3 months) of MI, CVA, TIA or CAD/stents, malignancy.",
         "Severe systemic disease that is a constant threat to life, or a moribund patient. Examples: recent (<3 months) MI, CVA, TIA or CAD/stents, ongoing cardiac ischemia, severe valve dysfunction, severely reduced EF, shock, sepsis, DIC, ARDS, ESRD not on regular dialysis, ruptured aneurysm, massive trauma."]
NM = ["ASA I", "ASA II", "ASA III", "ASA IV-V"]
def questions(desc):
    return {"choice": {"type": "choice", "instructions": "What is the ASA Physical Status class of this patient?", "criteria": {n: t for n, t in zip(NM, desc)}},
            "score": {"type": "score", "instructions": "What is the ASA Physical Status class of this patient? (0 = ASA I ... 3 = ASA IV-V)", "criteria": [f"{n}: {t}" for n, t in zip(NM, desc)]}}
sm = lambda a: (lambda e: e / e.sum(1, keepdims=True))(np.exp(a - a.max(1, keepdims=True)))
def cuts_fit(E, y):
    c = np.array([1.5, 2.5, 3.5]); g = np.linspace(1.0, 4.0, 121)
    for _ in range(3):
        for j in range(3): c[j] = max(g, key=lambda v: ck(y, 1 + (E[:, None] > np.where(np.arange(3) == j, v, c)).sum(1), weights="quadratic"))
    return c
def cv_kappa(E, y):
    rng = np.random.RandomState(0); f = rng.rand(len(y)) < 0.5; pr = np.zeros(len(y), int)
    for h in (True, False): c = cuts_fit(E[f == h], y[f == h]); pr[f != h] = 1 + (E[f != h][:, None] > c).sum(1)
    return ck(y, pr, weights="quadratic")
y = d.asa4.astype(int).values; rows = []; out = ["# OpenJev 配置调参（MOVER 调参集 n=%d；全部配置如实列出）\n" % len(d), "| 系统提示 | 选项描述 | 题型 | 未校准κ | 3切点校准κ(2折) | 宏AUC | 预测分布 |", "|---|---|---|---|---|---|---|"]
for sk, dk in itertools.product(SYS, ("short", "official")):
    pc = PromptConfig() if SYS[sk] is None else PromptConfig(system_prompt=SYS[sk])
    eng = SystemOneEngine(backend=load_backend("/mnt/c/backup/Qwen3.5-9B", device=f"cuda:{gpu}", dtype=torch.float16), prompt_config=pc) if "eng" not in globals() else eng
    eng.prompt_config = pc; Q = questions(SHORT if dk == "short" else OFFIC); L = {"choice": [], "score": []}
    for r in d.itertuples():
        raws, _ = eng.evaluate_raw(SystemOneRequest.model_validate({"state": r.text, "questions": Q}))
        for x in raws: L[x.decision.name].append(x.logprobs)
    for qt in ("choice", "score"):
        P = sm(np.array(L[qt])); E = (P * np.arange(1, 5)).sum(1); pr = P.argmax(1) + 1 if qt == "choice" else np.clip(np.rint(E), 1, 4).astype(int)
        auc = np.mean([roc_auc_score((y == k + 1).astype(int), P[:, k]) for k in range(4)]); k0 = ck(y, pr, weights="quadratic"); k1 = cv_kappa(E, y)
        rows.append(dict(system=sk, desc=dk, qtype=qt, kappa_raw=k0, kappa_cal=k1, auc=auc))
        out.append(f"| {sk} | {dk} | {qt} | {k0:.3f} | {k1:.3f} | {auc:.3f} | {np.bincount(pr-1,minlength=4).tolist()} |")
    open(f"{R}/openjev_tuning.md", "w").write("\n".join(out)); print(out[-2:], flush=True)
best = sorted(rows, key=lambda r: (-round(r["kappa_cal"], 3), r["system"] != "generic", r["desc"] != "short", r["qtype"] != "choice"))[0]
json.dump({"selected": best, "all": rows, "n_tune": len(d), "tune_seed": 17}, open(f"{R}/openjev_tuning_choice.json", "w"), ensure_ascii=False, indent=1)
open(f"{R}/openjev_tuning.md", "a").write(f"\n\n选定配置（冻结）：{best}\n"); print("选定", best)
```
