## Supplementary material for "Non-generative readout with few-label calibration versus LoRA fine-tuning of a large language model for ASA physical status classification: accuracy, speed, cost and label efficiency in three data sources": Analysis code and aggregate results: 34_ojev_lora.py

```
"""OpenJev + LoRA（研究 B：非生成式决策模型 + 微调）。提示、聊天模板、候选标签、闭集 softmax 与 OpenJev 完全一致（直接调用其 compile_request / HFPromptRenderer），
仅把训练循环改为 4-bit + 批处理 + 梯度检查点（OpenJev 原脚本为 bf16 逐例训练，SM75 显存/速度不可行）。损失变体（事先限定）：
  nll    = OpenJev 默认目标（候选集上的负对数似然）
  brier  = nll + 1.0×Brier（OpenJev 选项 --brier-weight）
  rps    = nll + 1.0×RPS（有序排序概率评分，proper；本项目对有序 ASA 的扩展）
冻结的提示配置读自 reports/openjev_tuning_choice.json（OpenJev 调参结果）。
用法：34_ojev_lora.py train <mover|local> <n> <variant> <gpu> <tag> | infer <tag> <mover_sub|mover_tune|local_val> <gpu>
种子由环境变量 SEED 控制；训练集与调参集（seed18，800 台）不重叠。"""
import os, sys, json, time, numpy as np, pandas as pd, torch
from openjev.backends.hf import HFPromptRenderer
from openjev.prompting import PromptConfig, compile_request
from openjev.schema import SystemOneRequest
from transformers import AutoTokenizer, AutoModelForCausalLM, BitsAndBytesConfig
from peft import LoraConfig, get_peft_model, PeftModel
SEED = int(os.environ.get("SEED", 20261017)); torch.manual_seed(SEED); mode = sys.argv[1]
D = os.path.expanduser("~/research/NE_ERAS_JEV/data/derived"); R = os.path.expanduser("~/research/Anes_JEV_vs_LoRA/analysis/reports"); M = os.path.expanduser("~/research/Anes_JEV_vs_LoRA/models")
src33 = open(os.path.expanduser("~/research/Anes_JEV_vs_LoRA/analysis/scripts/33_openjev_tune.py")).read()
ns = {}; exec("SYS =" + src33.split("SYS =", 1)[1].split("sm = lambda")[0], ns)    # 复用调参脚本里的 SYS / SHORT / OFFIC / NM / questions 定义，保证配置一致
SYS, SHORT, OFFIC, NM, questions = ns["SYS"], ns["SHORT"], ns["OFFIC"], ns["NM"], ns["questions"]
CH = json.load(open(f"{R}/openjev_tuning_choice.json"))["selected"]; QT = CH["qtype"]
pc = PromptConfig() if SYS[CH["system"]] is None else PromptConfig(system_prompt=SYS[CH["system"]]); QS = questions(SHORT if CH["desc"] == "short" else OFFIC)
path = "/mnt/c/backup/Qwen3.5-9B"; tok = AutoTokenizer.from_pretrained(path); tok.padding_side = "left"; rend = HFPromptRenderer(tok)
def prompt_ids(text):
    qs = SystemOneRequest.model_validate({"state": text, "questions": QS}).questions
    dec = [x for x in compile_request(text, qs, pc) if x.name == QT][0]; return rend.render_prompt(dec.prompt), dec.candidates
_, CANDS = prompt_ids("x"); cid = [rend.candidate_ids([c])[0] for c in CANDS]; assert all(len(c) == 1 for c in cid); CID = torch.tensor([c[0] for c in cid])
bnb = BitsAndBytesConfig(load_in_4bit=True, bnb_4bit_quant_type="nf4", bnb_4bit_compute_dtype=torch.float16, bnb_4bit_use_double_quant=True)
def load(gpu): return AutoModelForCausalLM.from_pretrained(path, quantization_config=bnb, dtype=torch.float16, device_map={"": gpu})
def batch_logits(m, texts, dev):
    ids = [prompt_ids(t)[0] for t in texts]; w = max(len(i) for i in ids); pad = tok.pad_token_id
    x = torch.tensor([[pad] * (w - len(i)) + i for i in ids], device=dev); a = torch.tensor([[0] * (w - len(i)) + [1] * len(i) for i in ids], device=dev)
    return m(input_ids=x, attention_mask=a, logits_to_keep=1, use_cache=False).logits[:, -1].float()[:, CID.to(dev)]
def loss_fn(lg, y, variant):
    lp = lg.log_softmax(-1); nll = torch.nn.functional.cross_entropy(lg, y)
    if variant == "nll": return nll
    P = lp.exp(); Y = torch.nn.functional.one_hot(y, lg.shape[1]).float()
    if variant == "brier": return nll + ((P - Y) ** 2).sum(1).mean()
    if variant == "rps": return nll + ((P.cumsum(1) - Y.cumsum(1)) ** 2).sum(1).mean()
    raise ValueError(variant)
def data(src):
    if src == "local":
        c = pd.read_csv(f"{D}/asa_common_local.csv"); return c, c[c.split == "dev"], c[c.split == "val"]
    m = pd.read_csv(f"{D}/asa_common_mover.csv"); return m, m[m.split == "train80"], None
if mode == "train":
    src, n, variant, gpu, tag = sys.argv[2], int(sys.argv[3]), sys.argv[4], int(sys.argv[5]), sys.argv[6]; dev = f"cuda:{gpu}"
    _, tr, _ = data(src); d = (tr.sample(n, random_state=SEED) if n < len(tr) else tr.sample(frac=1, random_state=SEED)).reset_index(drop=True); n = len(d)
    mdl = load(gpu); mdl.gradient_checkpointing_enable(gradient_checkpointing_kwargs={"use_reentrant": False}); mdl.enable_input_require_grads()
    mdl = get_peft_model(mdl, LoraConfig(r=16, lora_alpha=32, lora_dropout=0.05, target_modules=["q_proj", "k_proj", "v_proj", "o_proj", "gate_proj", "up_proj", "down_proj"], task_type="CAUSAL_LM")); mdl.train()
    opt = torch.optim.AdamW([p for p in mdl.parameters() if p.requires_grad], lr=1e-4); bs, acc = 4, 4; steps = n // (bs * acc)
    sch = torch.optim.lr_scheduler.OneCycleLR(opt, max_lr=1e-4, total_steps=steps, pct_start=0.05, anneal_strategy="cos"); t0 = time.time(); la = []
    for s in range(steps):
        for a in range(acc):
            b = d.iloc[(s * acc + a) * bs:(s * acc + a + 1) * bs]; l = loss_fn(batch_logits(mdl, list(b.text), dev), torch.tensor(b.asa4.values.astype(int) - 1, device=dev), variant) / acc; l.backward(); la.append(l.item() * acc)
        torch.nn.utils.clip_grad_norm_(mdl.parameters(), 1.0); opt.step(); sch.step(); opt.zero_grad()
        if (s + 1) % 10 == 0: print(f"step {s+1}/{steps} loss {np.mean(la[-40:]):.4f} {(time.time()-t0)/(s+1):.1f}s/step", flush=True)
    mdl.save_pretrained(f"{M}/{tag}"); print("训练完成", f"{time.time()-t0:.0f}s")
else:
    tag, ds, gpu = sys.argv[2], sys.argv[3], int(sys.argv[4]); dev = f"cuda:{gpu}"
    if ds == "vitaldb": d = pd.read_csv(f"{D}/asa_common_vitaldb.csv"); d = d[d.op_id.isin(pd.read_csv(f"{D}/vitaldb_sub2000.csv").op_id)]
    elif ds == "local_val": _, _, d = data("local")
    else:
        m_, tr, _ = data("mover"); cal = set(tr.sample(1000, random_state=11).op_id)
        if ds == "mover_sub": d = m_[m_.op_id.isin(pd.read_csv(f"{D}/mover_test_sub4000.csv").op_id)]
        else:   # mover_tune：LoRA 变体选择用，train80 内随机 800 台，排除校准集、OpenJev 调参集(seed17)与各训练子集(seed 20261017/11/12 的 n≤3000 抽样)
            ex = set(tr.sample(1000, random_state=11).op_id) | set(tr[~tr.op_id.isin(cal)].sample(800, random_state=17).op_id)
            for sd in (20261017, 11, 12):
                for nn in (100, 300, 1000, 3000): ex |= set(tr.sample(nn, random_state=sd).op_id)
            d = tr[~tr.op_id.isin(ex)].sample(800, random_state=18)
    d = d.reset_index(drop=True); mdl = PeftModel.from_pretrained(load(gpu), f"{M}/{tag}").eval(); rows = []; t0 = time.time()
    for i in range(0, len(d), 8):
        b = d.iloc[i:i + 8]
        with torch.no_grad(): p = batch_logits(mdl, list(b.text), dev).softmax(-1).cpu().numpy()
        for r, q in zip(b.itertuples(), p): rows.append(dict(op_id=r.op_id, asa4=r.asa4, p1=q[0], p2=q[1], p3=q[2], p4=q[3]))
        if (i // 8 + 1) % 100 == 0: print(i + 8, f"{(time.time()-t0)/(i+8):.2f}s/例", flush=True)
    pd.DataFrame(rows).to_csv(f"{D}/pred_ojlora_{tag}_{ds}.csv", index=False); print("完成", len(rows))
```
