## Supplementary material for "Non-generative readout with few-label calibration versus LoRA fine-tuning of a large language model for ASA physical status classification: accuracy, speed, cost and label efficiency in three data sources": Analysis code and aggregate results: 35_calib_curve.py

```
"""校准标签数曲线：零样本读出 + 切点校准，k 个标签（k∈{0,25,50,100,200,500,1000}），每个 k 随机抽 100 次（有放回的重复抽样子集，非自助），报告 κ 均值与 2.5–97.5 分位；
与同标签量的 LoRA 学习曲线并列。MOVER：校准集=train80 内 1000 台（种子 11），评估=sub4000；本院：校准集=dev 内 1000 台（种子 11），评估=val。输出 reports/calibration_curve.md"""
import os, numpy as np, pandas as pd
from sklearn.metrics import cohen_kappa_score as ck
D = os.path.expanduser("~/research/NE_ERAS_JEV/data/derived"); R = os.path.expanduser("~/research/Anes_JEV_vs_LoRA/analysis/reports"); PC = ["p1", "p2", "p3", "p4"]
def kap(y, p): return ck(y, p, weights="quadratic")
def fit(E, y):
    c = np.array([1.5, 2.5, 3.5]); g = np.linspace(1.0, 4.0, 61)
    for _ in range(2):
        for j in range(3): c[j] = max(g, key=lambda v: kap(y, 1 + (E[:, None] > np.where(np.arange(3) == j, v, c)).sum(1)))
    return c
def curve(cal, te, name):
    Ec = (cal[PC].values * np.arange(1, 5)).sum(1); yc = cal.asa4.astype(int).values; Et = (te[PC].values * np.arange(1, 5)).sum(1); yt = te.asa4.astype(int).values; rng = np.random.default_rng(7)
    rows = [f"\n## {name}\n", "| 校准标签数 k | κ 均值 | 2.5% | 97.5% |", "|---|---|---|---|", f"| 0（未校准 argmax） | {kap(yt, te[PC].values.argmax(1)+1):.3f} | — | — |"]
    for k in (25, 50, 100, 200, 500, 1000):
        ks = []
        for _ in range(100 if k < 1000 else 1):
            i = rng.choice(len(yc), k, replace=False); c = fit(Ec[i], yc[i]); ks.append(kap(yt, 1 + (Et[:, None] > c).sum(1)))
        rows.append(f"| {k} | {np.mean(ks):.3f} | {np.percentile(ks,2.5):.3f} | {np.percentile(ks,97.5):.3f} |")
    return rows
out = ["# 零样本读出 + 切点校准：校准标签数曲线（对照 LoRA 同标签量）"]
sub = set(pd.read_csv(f"{D}/mover_test_sub4000.csv").op_id)
cal = pd.read_csv(f"{D}/pred_llm_mover_cal.csv").set_index("op_id"); te = pd.read_csv(f"{D}/pred_llm_mover_test.csv").set_index("op_id"); te = te[te.index.isin(sub)]
out += curve(cal, te, "MOVER（sub4000）；对照任务 LoRA：n=100 0.410、300 0.411、1000 0.505、3000 0.563")
c = pd.read_csv(f"{D}/asa_common_local.csv", usecols=["op_id", "split"]); full = pd.read_csv(f"{D}/pred_llm_local.csv").set_index("op_id")
dev = c[c.split == "dev"].op_id.sample(1000, random_state=11); val = set(c[c.split == "val"].op_id)
out += curve(full.loc[dev], full[full.index.isin(val)], "本院 val；对照本院 LoRA：n=100 0.423、300 0.542、1000 0.559、4326 0.561")
open(f"{R}/calibration_curve.md", "w").write("\n".join(out)); print("\n".join(out))
```
