## Supplementary material for "Non-generative readout with few-label calibration versus LoRA fine-tuning of a large language model for ASA physical status classification: accuracy, speed, cost and label efficiency in three data sources": Analysis code and aggregate results: 36_build_vitaldb.py

```
"""VitalDB（SNUH，公开）作为第二个外部检验集：6,388 例非心脏手术，含 ASA。统一为与 MOVER/本院相同的 T1 模板；缺失项（术前血压心率、冠心病标志）写 unknown。
排除 ASA 缺失与 ASA 6（脑死亡供体）。ASA IV–V 合并（V 极少）。输出 NE_ERAS_JEV/data/derived/asa_common_vitaldb.csv。"""
import os, gzip, io, numpy as np, pandas as pd
R = os.path.expanduser("~/research/NE_ERAS_JEV"); c = pd.read_csv(io.BytesIO(gzip.decompress(open(f"{R}/data/raw/vitaldb/cases.csv.gz", "rb").read())))
c = c[c.asa.isin([1, 2, 3, 4, 5])].copy(); c["asa4"] = c.asa.astype(int).clip(upper=4)
f = lambda v, fmt: "unknown" if pd.isna(v) else fmt % v
def render(r):
    proc = f"{r.opname} ({r.optype})" if pd.notna(r.opname) else "unknown"
    return ("Age: %s years; Sex: %s; Weight: %s kg; BMI: %s; Setting: inpatient\nPreoperative vitals (last before OR): BP unknown/unknown mmHg, heart rate unknown bpm\n"
            "History: hypertension %s, coronary artery disease unknown, diabetes %s\nPreoperative labs: creatinine %s mg/dL, hemoglobin %s g/dL, albumin %s g/dL\nPlanned procedure: %s") % (
            f(r.age, "%.0f"), "female" if r.sex == "F" else "male", f(r.weight, "%.0f"), f(r.bmi, "%.1f"), "yes" if r.preop_htn == 1 else "no", "yes" if r.preop_dm == 1 else "no",
            f(r.preop_cr, "%.2f"), f(r.preop_hb, "%.1f"), f(r.preop_alb, "%.1f"), proc)
c["text"] = [render(r) for r in c.itertuples()]
out = c[["caseid", "subjectid", "asa4", "emop", "text"]].rename(columns={"caseid": "op_id"}); out["op_id"] = "vdb" + out.op_id.astype(str); out["split"] = "ext"
p = f"{R}/data/derived/asa_common_vitaldb.csv"; out.to_csv(p, index=False, encoding="utf-8-sig")
print(len(out), out.asa4.value_counts(normalize=True).sort_index().round(3).to_dict(), "患者数", out.subjectid.nunique()); print(out.text.iloc[0])
```
