## Supplementary material for "Non-generative readout with few-label calibration versus LoRA fine-tuning of a large language model for ASA physical status classification: accuracy, speed, cost and label efficiency in three data sources": Analysis code and aggregate results: 37_pick_variant.py

```
"""在 LoRA 调参集（mover_tune，800 台）上比较 OpenJev+LoRA 的 3 个损失变体，按未校准二次加权 κ 选择（平手取更简单：nll<brier<rps），写 reports/ojlora_variant_choice.json 与 ojlora_variant_tuning.md"""
import os, json, numpy as np, pandas as pd
from sklearn.metrics import cohen_kappa_score as ck, roc_auc_score
D = os.path.expanduser("~/research/NE_ERAS_JEV/data/derived"); R = os.path.expanduser("~/research/Anes_JEV_vs_LoRA/analysis/reports"); PC = ["p1", "p2", "p3", "p4"]; rows = []
for v in ("nll", "brier", "rps"):
    d = pd.read_csv(f"{D}/pred_ojlora_OJV_{v}_mover_tune.csv"); y = d.asa4.astype(int).values; P = d[PC].values; pr = P.argmax(1) + 1; Y = np.eye(4)[y - 1]
    conf = P.max(1); ok = pr == y; ece = sum(((conf > lo) & (conf <= lo + .1)).mean() * abs(ok[(conf > lo) & (conf <= lo + .1)].mean() - conf[(conf > lo) & (conf <= lo + .1)].mean()) for lo in np.linspace(0, .9, 10) if ((conf > lo) & (conf <= lo + .1)).any())
    rows.append(dict(variant=v, kappa=ck(y, pr, weights="quadratic"), acc=ok.mean(), auc=float(np.mean([roc_auc_score(Y[:, k], P[:, k]) for k in range(4)])), brier=float(((P - Y) ** 2).sum(1).mean()), ece=float(ece)))
best = sorted(rows, key=lambda r: (-round(r["kappa"], 3), ["nll", "brier", "rps"].index(r["variant"])))[0]
json.dump({"selected": best["variant"], "all": rows, "n": len(d)}, open(f"{R}/ojlora_variant_choice.json", "w"), indent=1)
open(f"{R}/ojlora_variant_tuning.md", "w").write("# OpenJev+LoRA 损失变体（LoRA 调参集 n=%d，n=1000 训练，种子 20261017）\n\n| 变体 | κ | 准确率 | 宏AUC | Brier | ECE |\n|---|---|---|---|---|---|\n" % len(d) + "\n".join(f"| {r['variant']} | {r['kappa']:.3f} | {r['acc']:.3f} | {r['auc']:.3f} | {r['brier']:.3f} | {r['ece']:.3f} |" for r in rows) + f"\n\n选定：{best['variant']}\n")
print(best)
```
