## Supplementary material for "Non-generative readout with few-label calibration versus LoRA fine-tuning of a large language model for ASA physical status classification: accuracy, speed, cost and label efficiency in three data sources": Analysis code and aggregate results: 38_label_format.py

```
"""探索性臂：选项标签格式（数字 1–5 / 字母 A–E / 罗马数字 I–V），五个选项（ASA I…V），OpenJev 后端打分（含其前缀歧义终止符处理），IV 与 V 概率合并为第 4 类。
提示（系统提示/选项描述）用 OpenJev 调参冻结配置（choice 措辞）。MOVER：sub4000 前 1500 台；本院：val 随机 1000 台（种子 5）。输出 pred_fmt_<fmt>_<ds>.csv"""
import os, sys, json, time, numpy as np, pandas as pd, torch
from openjev.backends import load_backend
from openjev.backends.base import ScoringTask
from openjev.prompting import Prompt, PromptConfig, DEFAULT_SYSTEM_PROMPT
gpu = int(sys.argv[1]); D = os.path.expanduser("~/research/NE_ERAS_JEV/data/derived"); R = os.path.expanduser("~/research/Anes_JEV_vs_LoRA/analysis/reports")
ns = {}; exec("SYS =" + open(os.path.expanduser("~/research/Anes_JEV_vs_LoRA/analysis/scripts/33_openjev_tune.py")).read().split("SYS =", 1)[1].split("sm = lambda")[0], ns)
CH = json.load(open(f"{R}/openjev_tuning_choice.json"))["selected"]; SYSP = ns["SYS"][CH["system"]] or DEFAULT_SYSTEM_PROMPT; DESC = ns["SHORT"] if CH["desc"] == "short" else ns["OFFIC"]
DESC5 = [DESC[0], DESC[1], DESC[2], "Severe systemic disease that is a constant threat to life", "A moribund patient who is not expected to survive without the operation"]
NAMES = ["ASA I", "ASA II", "ASA III", "ASA IV", "ASA V"]
FM = {"digits": ["1", "2", "3", "4", "5"], "letters": ["A", "B", "C", "D", "E"], "roman": ["I", "II", "III", "IV", "V"]}
def user(state, labels):
    opts = "\n".join(f"{l}. {n}: {t}" for l, n, t in zip(labels, NAMES, DESC5))
    return f"## State\n{state}\n\n## Question\nWhat is the ASA Physical Status class of this patient?\n\n## Options\n{opts}\n\nReply with the option label only."
be = load_backend("/mnt/c/backup/Qwen3.5-9B", device=f"cuda:{gpu}", dtype=torch.float16)
mv = pd.read_csv(f"{D}/asa_common_mover.csv"); mv = mv[mv.op_id.isin(pd.read_csv(f"{D}/mover_test_sub4000.csv").op_id.tolist()[:1500])]
lc = pd.read_csv(f"{D}/asa_common_local.csv"); lc = lc[lc.split == "val"].sample(1000, random_state=5)
for ds, d in (("mover", mv), ("local", lc)):
    out = {f: [] for f in FM}; t0 = time.time()
    for i, r in enumerate(d.itertuples()):
        res = be.score([ScoringTask(prompt=Prompt(system=SYSP, user=user(r.text, FM[f])), candidates=FM[f]) for f in FM])
        for f, x in zip(FM, res):
            lp = np.array(x.logprobs); p = np.exp(lp - lp.max()); p /= p.sum(); out[f].append([r.op_id, r.asa4, p[0], p[1], p[2], p[3] + p[4]])
        if (i + 1) % 250 == 0: print(ds, i + 1, f"{(time.time()-t0)/(i+1):.2f}s/例", flush=True)
    for f in FM: pd.DataFrame(out[f], columns=["op_id", "asa4", "p1", "p2", "p3", "p4"]).to_csv(f"{D}/pred_fmt_{f}_{ds}.csv", index=False)
print("完成")
```
