## Supplementary material for "Non-generative readout with few-label calibration versus LoRA fine-tuning of a large language model for ASA physical status classification: accuracy, speed, cost and label efficiency in three data sources": Analysis code and aggregate results: 39_vdb_gate.py

```
"""VitalDB 第一阶段闸门（子集 2000 台，患者级）。事先规定：继续后续 VitalDB 分析（冻结 OpenJev、本院 LoRA 迁移）当且仅当
 校准读出（切点取自 MOVER 校准集 1000 标签） − MOVER-LoRA 12k 迁移 的 κ 差的患者级 bootstrap 95%CI 下界 > −0.05（即在第二外部集上同样非劣效）；
 否则 VitalDB 止于第一阶段并如实报告为未能重复。输出 reports/vitaldb_gate.json 与 vitaldb_stage1.md"""
import os, json, numpy as np, pandas as pd
from sklearn.metrics import cohen_kappa_score as ck, roc_auc_score
D = os.path.expanduser("~/research/NE_ERAS_JEV/data/derived"); R = os.path.expanduser("~/research/Anes_JEV_vs_LoRA/analysis/reports"); PC = ["p1", "p2", "p3", "p4"]
kap = lambda y, p: ck(y, p, weights="quadratic")
v = pd.read_csv(f"{D}/asa_common_vitaldb.csv", usecols=["op_id", "subjectid"]).set_index("op_id")
zs = pd.read_csv(f"{D}/pred_llm_vitaldb.csv").set_index("op_id"); lo = pd.read_csv(f"{D}/pred_lora_mover12k_vitaldb.csv").set_index("op_id"); ids = sorted(set(zs.index) & set(lo.index)); zs, lo = zs.loc[ids], lo.loc[ids]
cal = pd.read_csv(f"{D}/pred_llm_mover_cal.csv").set_index("op_id"); Ec = (cal[PC].values * np.arange(1, 5)).sum(1); yc = cal.asa4.astype(int).values
c = np.array([1.5, 2.5, 3.5]); g = np.linspace(1.0, 4.0, 121)
for _ in range(3):
    for j in range(3): c[j] = max(g, key=lambda t: kap(yc, 1 + (Ec[:, None] > np.where(np.arange(3) == j, t, c)).sum(1)))
y = zs.asa4.astype(int).values; p0 = zs[PC].values.argmax(1) + 1; pc = 1 + ((zs[PC].values * np.arange(1, 5)).sum(1)[:, None] > c).sum(1); pl = lo[PC].values.argmax(1) + 1
pat = v.loc[ids].subjectid.values; up = np.unique(pat); ix = {p: np.where(pat == p)[0] for p in up}; rng = np.random.default_rng(3); B = [np.concatenate([ix[p] for p in rng.choice(up, len(up))]) for _ in range(1000)]
def boot(a, b): d = np.array([kap(y[s], a[s]) - kap(y[s], b[s]) for s in B]); return d.mean(), np.percentile(d, 2.5), np.percentile(d, 97.5)
auc = lambda P: np.mean([roc_auc_score((y == k + 1).astype(int), P[:, k]) for k in range(1, 4) if 0 < (y == k + 1).sum() < len(y)])
res = {"n": len(ids), "kappa_zs": kap(y, p0), "kappa_zs_cal": kap(y, pc), "kappa_lora12k": kap(y, pl), "auc_zs": auc(zs[PC].values), "auc_lora": auc(lo[PC].values), "cuts": c.round(2).tolist(),
       "diff_cal_minus_lora": boot(pc, pl), "diff_lora_minus_zs": boot(pl, p0), "dist_true": np.bincount(y - 1, minlength=4).tolist(), "dist_zs": np.bincount(p0 - 1, minlength=4).tolist(), "dist_cal": np.bincount(pc - 1, minlength=4).tolist(), "dist_lora": np.bincount(pl - 1, minlength=4).tolist()}
res["go"] = bool(res["diff_cal_minus_lora"][1] > -0.05); json.dump(res, open(f"{R}/vitaldb_gate.json", "w"), indent=1, default=float)
open(f"{R}/vitaldb_stage1.md", "w").write("# VitalDB 第一阶段（子集 n=%d）\n\n| 臂 | κ | 宏AUC(II-IV维度) |\n|---|---|---|\n| 读出零样本 | %.3f | %.3f |\n| 读出 + MOVER 标签切点校准 | %.3f | — |\n| MOVER-LoRA 12k 迁移 | %.3f | %.3f |\n\n校准读出 − LoRA 迁移：%+.3f（%+.3f ~ %+.3f）；LoRA 迁移 − 零样本：%+.3f（%+.3f ~ %+.3f）\n\n预测分布 真实 %s｜零样本 %s｜校准 %s｜LoRA %s\n\n闸门（下界 > −0.05）：%s\n" % (
    len(ids), res["kappa_zs"], res["auc_zs"], res["kappa_zs_cal"], res["kappa_lora12k"], res["auc_lora"], *res["diff_cal_minus_lora"], *res["diff_lora_minus_zs"], res["dist_true"], res["dist_zs"], res["dist_cal"], res["dist_lora"], "通过，继续" if res["go"] else "未通过，VitalDB 止于第一阶段"))
print(open(f"{R}/vitaldb_stage1.md").read())
```
