## Supplementary material for "Non-generative readout with few-label calibration versus LoRA fine-tuning of a large language model for ASA physical status classification: accuracy, speed, cost and label efficiency in three data sources": Analysis code and aggregate results: 40_vdb_stage2.py

```
"""VitalDB 第二阶段（探索性外部重复，子集 n=2000）：冻结 OpenJev 与本院 LoRA 的跨院迁移。

口径与 39_vdb_gate.py 一致：
- 读出臂切点校准：切点在 MOVER 校准集（train80，seed 11，1000 台）上拟合，再用于 VitalDB。
- OpenJev（协议修订 1 §5）：先在 MOVER 校准集上按 NLL 最小化拟合温度 T，再在同一校准集上拟合期望分的 3 个切点。
- LoRA 迁移：argmax，不校准。
- 区间：患者级 cluster bootstrap，B=1000，种子 3；配对差为同一重抽样下两臂 κ 之差。

输出：analysis/reports/vitaldb_stage2.md 与 vitaldb_stage2.json。
"""
import os, json, numpy as np, pandas as pd
from sklearn.metrics import cohen_kappa_score as ck, roc_auc_score

D = os.path.expanduser("~/research/NE_ERAS_JEV/data/derived")
R = os.path.expanduser("~/research/Anes_JEV_vs_LoRA/analysis/reports")
PC = ["p1", "p2", "p3", "p4"]
OC = [f"c{k}" for k in range(4)]
kap = lambda y, p: ck(y, p, weights="quadratic")


def softmax(a):
    a = np.asarray(a, float)
    e = np.exp(a - a.max(1, keepdims=True))
    return e / e.sum(1, keepdims=True)


def cuts_fit(E, y):
    """坐标搜索 3 个切点，最大化二次加权 κ（与 39_vdb_gate.py 相同）。"""
    c = np.array([1.5, 2.5, 3.5]); g = np.linspace(1.0, 4.0, 121)
    for _ in range(3):
        for j in range(3):
            c[j] = max(g, key=lambda t: kap(y, 1 + (E[:, None] > np.where(np.arange(3) == j, t, c)).sum(1)))
    return c


def apply_cuts(E, c):
    return 1 + (E[:, None] > c).sum(1)


v = pd.read_csv(f"{D}/asa_common_vitaldb.csv", usecols=["op_id", "subjectid"]).set_index("op_id")
readout = pd.read_csv(f"{D}/pred_llm_vitaldb.csv").set_index("op_id")
m12k = pd.read_csv(f"{D}/pred_lora_mover12k_vitaldb.csv").set_index("op_id")
loc = pd.read_csv(f"{D}/pred_lora_LOC4326s20261017_vitaldb.csv").set_index("op_id")
ids = sorted(set(readout.index) & set(m12k.index) & set(loc.index))

oj = None
p_oj = f"{D}/pred_ojevF_vitaldb.csv"
if os.path.exists(p_oj):
    oj = pd.read_csv(p_oj).set_index("op_id")
    ids = [i for i in ids if i in oj.index]

readout, m12k, loc = readout.loc[ids], m12k.loc[ids], loc.loc[ids]
if oj is not None:
    oj = oj.loc[ids]
y = readout.asa4.astype(int).values
pat = v.loc[ids].subjectid.values
up = np.unique(pat)
ix = {p: np.where(pat == p)[0] for p in up}

# 读出臂：MOVER 校准集切点
cal = pd.read_csv(f"{D}/pred_llm_mover_cal.csv").set_index("op_id")
Ecal = (cal[PC].values * np.arange(1, 5)).sum(1); ycal = cal.asa4.astype(int).values
c_read = cuts_fit(Ecal, ycal)

# OpenJev：NLL 温度 + 期望分切点（均只在校准集上拟合）
T, c_oj = None, None
if oj is not None:
    gcal = pd.read_csv(f"{D}/pred_ojevF_mover_cal.csv").set_index("op_id")
    Lc = gcal[OC].values; yg = gcal.asa4.astype(int).values
    nll = lambda t: float(-np.log(np.clip(softmax(Lc / t)[np.arange(len(yg)), yg - 1], 1e-12, None)).mean())
    T = float(min(np.linspace(0.3, 3.0, 271), key=nll))
    c_oj = cuts_fit((softmax(Lc / T) * np.arange(1, 5)).sum(1), yg)

# 各臂预测与（用于 AUC 的）概率矩阵
arms, probs = {}, {}
probs["读出零样本"] = readout[PC].values
arms["读出零样本"] = probs["读出零样本"].argmax(1) + 1
arms["读出 + MOVER 切点校准"] = apply_cuts((readout[PC].values * np.arange(1, 5)).sum(1), c_read)
probs["读出 + MOVER 切点校准"] = probs["读出零样本"]
probs["MOVER-LoRA 12k 迁移"] = m12k[PC].values
arms["MOVER-LoRA 12k 迁移"] = probs["MOVER-LoRA 12k 迁移"].argmax(1) + 1
probs["本院 LoRA 迁移（LO-C4326 种子 20261017）"] = loc[PC].values
arms["本院 LoRA 迁移（LO-C4326 种子 20261017）"] = probs["本院 LoRA 迁移（LO-C4326 种子 20261017）"].argmax(1) + 1
if oj is not None:
    Po = softmax(oj[OC].values)
    probs["冻结 OpenJev（未校准）"] = Po
    arms["冻结 OpenJev（未校准）"] = Po.argmax(1) + 1
    probs["冻结 OpenJev + 温度/切点校准"] = Po
    arms["冻结 OpenJev + 温度/切点校准"] = apply_cuts((Po * np.arange(1, 5)).sum(1), c_oj)

rng = np.random.default_rng(3)
B = [np.concatenate([ix[p] for p in rng.choice(up, len(up))]) for _ in range(1000)]


def boot(a, b):
    d = np.array([kap(y[s], a[s]) - kap(y[s], b[s]) for s in B])
    return [round(float(x), 4) for x in (d.mean(), np.percentile(d, 2.5), np.percentile(d, 97.5))]


def auc(P):
    return float(np.mean([roc_auc_score((y == k + 1).astype(int), P[:, k])
                          for k in range(1, 4) if 0 < (y == k + 1).sum() < len(y)]))


res = {"n": int(len(ids)), "patients": int(len(up)), "temperature": T,
       "cuts_readout": c_read.round(2).tolist(), "cuts_ojev": None if c_oj is None else c_oj.round(2).tolist(),
       "arms": {}, "diffs": {}}
rows = []
for nm, pr in arms.items():
    k = kap(y, pr)
    res["arms"][nm] = {"kappa": round(float(k), 4), "acc": round(float((pr == y).mean()), 4),
                       "auc": round(auc(probs[nm]), 4), "dist": np.bincount(pr - 1, minlength=4).tolist()}
    rows.append((nm, k, (pr == y).mean(), auc(probs[nm]), np.bincount(pr - 1, minlength=4).tolist()))

pairs = [("本院 LoRA 迁移（LO-C4326 种子 20261017）", "MOVER-LoRA 12k 迁移"),
         ("读出 + MOVER 切点校准", "本院 LoRA 迁移（LO-C4326 种子 20261017）")]
if oj is not None:
    pairs += [("冻结 OpenJev + 温度/切点校准", "本院 LoRA 迁移（LO-C4326 种子 20261017）"),
              ("冻结 OpenJev + 温度/切点校准", "读出 + MOVER 切点校准")]
for a, b in pairs:
    res["diffs"][f"{a} − {b}"] = boot(arms[a], arms[b])

hdr = "| 臂 | κ | 准确率 | 宏AUC | 预测分布 |\n|---|---|---|---|---|"
body = "\n".join(f"| {nm} | {k:.3f} | {ac:.3f} | {au:.3f} | {d} |" for nm, k, ac, au, d in rows)
dtxt = "\n".join(f"- {k}：{v[0]:+.3f}（{v[1]:+.3f} ~ {v[2]:+.3f}）" for k, v in res["diffs"].items())
txt = f"""# VitalDB 第二阶段（探索性外部重复，子集 n={len(ids)}，患者 {len(up)}）

阶段一闸门通过（校准读出 − MOVER-LoRA 迁移，患者级 95%CI 下界 +0.058 > −0.05），按事前规定继续第二阶段：冻结 OpenJev 与本院 LoRA 的跨院迁移（不训练，不重校准）。

| 项 | 值 |
|---|---|
| OpenJev 温度 T（MOVER 校准集 NLL 最小化） | {T} |
| 读出切点（MOVER 校准集） | {c_read.round(2).tolist()} |
| OpenJev 切点（MOVER 校准集） | {None if c_oj is None else c_oj.round(2).tolist()} |

{hdr}
{body}

患者级配对 bootstrap（B=1000，种子 3）：

{dtxt}

真值分布：{np.bincount(y - 1, minlength=4).tolist()}
"""
open(f"{R}/vitaldb_stage2.md", "w").write(txt)
json.dump(res, open(f"{R}/vitaldb_stage2.json", "w"), ensure_ascii=False, indent=1)
print(txt)
```
