## Supplementary material for "Non-generative readout with few-label calibration versus LoRA fine-tuning of a large language model for ASA physical status classification: accuracy, speed, cost and label efficiency in three data sources": Analysis code and aggregate results: 41_vdb_target_cal.py

```
"""VitalDB 目标站点校准曲线（探索性）：若在 VitalDB 本地有 k 个标注，切点校准能让读出臂达到多少 κ。

做法：随机抽 k 台拟合 3 个切点（在期望分上最大化二次加权 κ），在其余台上评估；重复 100 次，
报告均值与 2.5/97.5 百分位。k=50,100,200,500,1000。不使用任何其他站点标签。
输出：analysis/reports/vitaldb_target_cal.md 与 .json
"""
import os, json, numpy as np, pandas as pd
from sklearn.metrics import cohen_kappa_score as ck

D = os.path.expanduser("~/research/NE_ERAS_JEV/data/derived")
R = os.path.expanduser("~/research/Anes_JEV_vs_LoRA/analysis/reports")
PC = ["p1", "p2", "p3", "p4"]
kap = lambda y, p: ck(y, p, weights="quadratic")


def cuts_fit(E, y):
    c = np.array([1.5, 2.5, 3.5]); g = np.linspace(1.0, 4.0, 121)
    for _ in range(3):
        for j in range(3):
            c[j] = max(g, key=lambda t: kap(y, 1 + (E[:, None] > np.where(np.arange(3) == j, t, c)).sum(1)))
    return c


d = pd.read_csv(f"{D}/pred_llm_vitaldb.csv")
y = d.asa4.astype(int).values
E = (d[PC].values * np.arange(1, 5)).sum(1)
p0 = d[PC].values.argmax(1) + 1
res = {"n": int(len(y)), "zero_shot_kappa": float(kap(y, p0)), "curve": []}
lines = ["# VitalDB 目标站点校准（读出臂，n=%d）\n" % len(y),
         "零样本 κ=%.3f。若 VitalDB 本地有 k 个标注，用其拟合 3 个切点、在其余台上评估；重复 100 次（种子 5）。\n" % res["zero_shot_kappa"],
         "| k | κ 均值 | 2.5% | 97.5% |", "|---|---|---|---|"]
rng = np.random.default_rng(5)
for k in (50, 100, 200, 500, 1000):
    vals = []
    for _ in range(100):
        idx = rng.permutation(len(y)); tr, te = idx[:k], idx[k:]
        c = cuts_fit(E[tr], y[tr])
        vals.append(kap(y[te], 1 + (E[te][:, None] > c).sum(1)))
    m, lo, hi = float(np.mean(vals)), float(np.percentile(vals, 2.5)), float(np.percentile(vals, 97.5))
    res["curve"].append({"k": k, "mean": m, "lo": lo, "hi": hi})
    lines.append(f"| {k} | {m:.3f} | {lo:.3f} | {hi:.3f} |")
open(f"{R}/vitaldb_target_cal.md", "w").write("\n".join(lines) + "\n")
json.dump(res, open(f"{R}/vitaldb_target_cal.json", "w"), indent=1)
print("\n".join(lines))
```
