## Supplementary material for "Non-generative readout with few-label calibration versus LoRA fine-tuning of a large language model for ASA physical status classification: accuracy, speed, cost and label efficiency in three data sources": Analysis code and aggregate results: 42_ojlora_confirm.py

```
"""OpenJev+LoRA 与任务 LoRA 的同集配对比较（MOVER 测试子集 mover_sub，患者级）。

只在同一批病例上比较：同一 n、同一种子，OpenJev+LoRA 对任务 LoRA（字母 logits 读出 + CE）。
调参集（mover_tune, seed18）的数字不进入本表。
区间：患者级 cluster bootstrap，B=1000，种子 3；配对差为同一重抽样下两臂 κ 之差。

输出：analysis/reports/ojlora_confirm_mover.md / .json
"""
import os, json, glob, re, numpy as np, pandas as pd
from sklearn.metrics import cohen_kappa_score as ck, roc_auc_score

D = os.path.expanduser("~/research/NE_ERAS_JEV/data/derived")
R = os.path.expanduser("~/research/Anes_JEV_vs_LoRA/analysis/reports")
PC = ["p1", "p2", "p3", "p4"]
kap = lambda y, p: ck(y, p, weights="quadratic")

m = pd.read_csv(f"{D}/mover_pid_map.csv").set_index("op_id")


def task_file(n, seed):
    if seed == 20261017:
        return f"{D}/pred_lora_curve{n}_mover_sub.csv"
    return f"{D}/pred_lora_curve{n}s{seed}_mover_sub.csv"


rows, res = [], {"n_test": None, "patients": None, "pairs": []}
oj_files = sorted(glob.glob(f"{D}/pred_ojlora_OJmover*s*_mover_sub.csv"))
loaded = {}
for f in oj_files:
    mm = re.search(r"OJmover(\d+)s(\d+)_mover_sub", f)
    if not mm:
        continue
    n, seed = int(mm.group(1)), int(mm.group(2))
    tf = task_file(n, seed)
    if not os.path.exists(tf):
        res["pairs"].append({"n": n, "seed": seed, "note": "任务 LoRA 预测缺失"})
        continue
    oj = pd.read_csv(f).set_index("op_id"); lo = pd.read_csv(tf).set_index("op_id")
    ids = sorted(set(oj.index) & set(lo.index))
    oj, lo = oj.loc[ids], lo.loc[ids]
    y = oj.asa4.astype(int).values
    po, pl = oj[PC].values.argmax(1) + 1, lo[PC].values.argmax(1) + 1
    pat = m.loc[ids].pid.values; up = np.unique(pat); ix = {p: np.where(pat == p)[0] for p in up}
    rng = np.random.default_rng(3)
    B = [np.concatenate([ix[p] for p in rng.choice(up, len(up))]) for _ in range(1000)]
    d = np.array([kap(y[s], po[s]) - kap(y[s], pl[s]) for s in B])
    diff = [round(float(x), 4) for x in (d.mean(), np.percentile(d, 2.5), np.percentile(d, 97.5))]
    auc = lambda P: float(np.mean([roc_auc_score((y == k + 1).astype(int), P[:, k])
                                   for k in range(1, 4) if 0 < (y == k + 1).sum() < len(y)]))
    rec = {"n": n, "seed": seed, "n_test": int(len(ids)), "patients": int(len(up)),
           "kappa_oj": round(float(kap(y, po)), 4), "kappa_task": round(float(kap(y, pl)), 4),
           "acc_oj": round(float((po == y).mean()), 4), "acc_task": round(float((pl == y).mean()), 4),
           "auc_oj": round(auc(oj[PC].values), 4), "auc_task": round(auc(lo[PC].values), 4),
           "diff_oj_minus_task": diff}
    res["pairs"].append(rec); res["n_test"] = int(len(ids)); res["patients"] = int(len(up))

lines = ["# OpenJev+LoRA 与任务 LoRA 的同集配对比较（MOVER 测试子集）\n",
         "同一批 %s 例（患者 %s），同一 n、同一种子；调参集数字不在此表。\n" % (res["n_test"], res["patients"]),
         "| n | 种子 | OpenJev+LoRA κ | 任务 LoRA κ | 差值（95%CI） | OJ 准确率/AUC | 任务准确率/AUC |",
         "|---|---|---|---|---|---|---|"]
for r in res["pairs"]:
    if "note" in r:
        continue
    lines.append("| %d | %d | %.3f | %.3f | %+.3f（%+.3f ~ %+.3f） | %.3f / %.3f | %.3f / %.3f |" % (
        r["n"], r["seed"], r["kappa_oj"], r["kappa_task"], *r["diff_oj_minus_task"],
        r["acc_oj"], r["auc_oj"], r["acc_task"], r["auc_task"]))
ok = [r for r in res["pairs"] if "note" not in r]
if ok:
    ds = [r["diff_oj_minus_task"][0] for r in ok]
    lines.append("\n差值均值 %+.4f（%d 个种子）" % (np.mean(ds), len(ok)))
    lines.append("\n注：差值为 OpenJev+LoRA − 任务 LoRA；正值表示 OpenJev+LoRA 更高。")
open(f"{R}/ojlora_confirm_mover.md", "w").write("\n".join(lines) + "\n")
json.dump(res, open(f"{R}/ojlora_confirm_mover.json", "w"), indent=1, ensure_ascii=False)
print("\n".join(lines))
```
