## Supplementary material for "Non-generative readout with few-label calibration versus LoRA fine-tuning of a large language model for ASA physical status classification: accuracy, speed, cost and label efficiency in three data sources": Analysis code and aggregate results: 50_ojev_frozen_mover.py

```
"""冻结 OpenJev（anes 提示 + 一句话描述 + score 题型）在 MOVER 测试子集的结果：未校准 / 3 切点校准（1000 标签），与自写读出同集配对（患者级 bootstrap）。输出 reports/ojev_frozen_mover.md"""
import os, numpy as np, pandas as pd
from sklearn.metrics import cohen_kappa_score as ck, roc_auc_score
D = os.path.expanduser("~/research/NE_ERAS_JEV/data/derived"); R = os.path.expanduser("~/research/Anes_JEV_vs_LoRA/analysis/reports")
sm = lambda a: (lambda e: e / e.sum(1, keepdims=True))(np.exp(a - a.max(1, keepdims=True)))
def load(f):
    d = pd.read_csv(f"{D}/{f}.csv").set_index("op_id"); d = d[d.qtype == "score"] if "qtype" in d else d
    return d
t = load("pred_ojevF_mover_sub"); c = load("pred_ojevF_mover_cal"); print(t.shape, c.shape, t.columns.tolist())
cols = [x for x in t.columns if x.startswith("c")]; P = lambda d: sm(d[cols].values)
def cuts_fit(E, y):
    cu = np.array([1.5, 2.5, 3.5]); g = np.linspace(1.0, 4.0, 121)
    for _ in range(3):
        for j in range(3): cu[j] = max(g, key=lambda v: ck(y, 1 + (E[:, None] > np.where(np.arange(3) == j, v, cu)).sum(1), weights="quadratic"))
    return cu
Et = (P(t) * np.arange(1, 5)).sum(1); Ec = (P(c) * np.arange(1, 5)).sum(1); yt = t.asa4.astype(int).values; yc = c.asa4.astype(int).values
cu = cuts_fit(Ec, yc); pr_cal = 1 + (Et[:, None] > cu).sum(1); pr_raw = P(t).argmax(1) + 1; pr_exp = np.clip(np.rint(Et), 1, 4).astype(int)
auc = np.mean([roc_auc_score((yt == k + 1).astype(int), P(t)[:, k]) for k in range(4)])
own = pd.read_csv(f"{D}/pred_llm_mover_test.csv").set_index("op_id").loc[t.index]; pown = own[["p1", "p2", "p3", "p4"]].values.argmax(1) + 1
pid = pd.read_csv(f"{D}/mover_pid_map.csv").set_index("op_id").pid.loc[t.index].values; up = np.unique(pid); idx = {p: np.where(pid == p)[0] for p in up}
rng = np.random.default_rng(3); B = [np.concatenate([idx[p] for p in rng.choice(up, len(up))]) for _ in range(1000)]
def ci(f): v = np.array([f(s) for s in B]); return f"{np.percentile(v,2.5):+.3f} ~ {np.percentile(v,97.5):+.3f}"
k = lambda pr: ck(yt, pr, weights="quadratic")
out = [f"# 冻结 OpenJev（Qwen3.5-9B）MOVER 测试子集 n={len(t)}，患者 {len(up)}\n", f"切点（1000 标签）：{np.round(cu,2).tolist()}；宏 AUC {auc:.3f}\n", "| 臂 | κ |", "|---|---|",
       f"| OpenJev score 未校准（argmax） | {k(pr_raw):.3f} |", f"| OpenJev score 期望四舍五入 | {k(pr_exp):.3f} |", f"| OpenJev + 3 切点校准 | {k(pr_cal):.3f} |", f"| 自写读出（未校准，同集） | {k(pown):.3f} |", ""]
out.append(f"- OpenJev 切点校准 − 自写读出：{k(pr_cal)-k(pown):+.3f}（{ci(lambda s: ck(yt[s], pr_cal[s], weights='quadratic') - ck(yt[s], pown[s], weights='quadratic'))}）")
open(f"{R}/ojev_frozen_mover.md", "w").write("\n".join(out)); print("\n".join(out))
```
