## Supplementary material for "Non-generative readout with few-label calibration versus LoRA fine-tuning of a large language model for ASA physical status classification: accuracy, speed, cost and label efficiency in three data sources": Analysis code and aggregate results: 51_ojev_frozen_local.py

```
"""冻结 OpenJev 在本院 val 上的 H1–H3 重复（扩展臂）：参考=本院麻醉记录 ASA；校准=本院 dev 随机 1000 标签（种子 11，同自写读出）。输出 reports/ojev_frozen_local.md"""
import os, numpy as np, pandas as pd
from sklearn.metrics import cohen_kappa_score as ck, roc_auc_score
D = os.path.expanduser("~/research/NE_ERAS_JEV/data/derived"); R = os.path.expanduser("~/research/Anes_JEV_vs_LoRA/analysis/reports"); PC = ["p1", "p2", "p3", "p4"]
sm = lambda a: (lambda e: e / e.sum(1, keepdims=True))(np.exp(a - a.max(1, keepdims=True)))
c = pd.read_csv(f"{D}/asa_common_local.csv", usecols=["op_id", "split", "asa4"]).merge(pd.read_csv(f"{D}/asa_dataset_v1.csv", usecols=["operation_id", "patient_sn"]).rename(columns={"operation_id": "op_id"}), on="op_id").set_index("op_id")
oj = pd.read_csv(f"{D}/pred_ojevF_local.csv"); oj = oj[oj.qtype == "score"].set_index("op_id") if "qtype" in oj else oj.set_index("op_id")
cols = [x for x in oj.columns if x.startswith("c")]; E = pd.Series((sm(oj[cols].values) * np.arange(1, 5)).sum(1), index=oj.index); Pj = pd.DataFrame(sm(oj[cols].values), index=oj.index)
dev = c[c.split == "dev"].sample(1000, random_state=11).index; val = c[c.split == "val"].index.intersection(oj.index)
def cuts_fit(Ev, y):
    cu = np.array([1.5, 2.5, 3.5]); g = np.linspace(1.0, 4.0, 121)
    for _ in range(3):
        for j in range(3): cu[j] = max(g, key=lambda v: ck(y, 1 + (Ev[:, None] > np.where(np.arange(3) == j, v, cu)).sum(1), weights="quadratic"))
    return cu
cu = cuts_fit(E.loc[dev].values, c.loc[dev].asa4.astype(int).values); y = c.loc[val].asa4.astype(int).values
pr_cal = 1 + (E.loc[val].values[:, None] > cu).sum(1); pr_raw = Pj.loc[val].values.argmax(1) + 1
rd = lambda f: pd.read_csv(f"{D}/{f}.csv").set_index("op_id")
lora1k = rd("pred_lora_LOC1000s20261017_local_val").loc[val][PC].values.argmax(1) + 1; lora_all = rd("pred_lora_LOC4326s20261017_local_val").loc[val][PC].values.argmax(1) + 1
own = rd("pred_llm_local").loc[val][PC].values.argmax(1) + 1
pid = c.loc[val].patient_sn.values; up = np.unique(pid); idx = {p: np.where(pid == p)[0] for p in up}; rng = np.random.default_rng(5); B = [np.concatenate([idx[p] for p in rng.choice(up, len(up))]) for _ in range(1000)]
K = lambda s, pr: ck(y[s], pr[s], weights="quadratic")
def d(a, b): v = np.array([K(s, a) - K(s, b) for s in B]); return f"{ck(y,a,weights='quadratic')-ck(y,b,weights='quadratic'):+.3f}（{np.percentile(v,2.5):+.3f} ~ {np.percentile(v,97.5):+.3f}）"
auc = np.mean([roc_auc_score((y == k + 1).astype(int), Pj.loc[val].values[:, k]) for k in range(4)])
out = [f"# 冻结 OpenJev 在本院 val（n={len(val)}，患者 {len(up)}）\n", f"切点（dev 1000 标签）{np.round(cu,2).tolist()}；宏 AUC {auc:.3f}\n", "| 臂 | κ |", "|---|---|"]
for nm, pr in (("OpenJev score 未校准（argmax）", pr_raw), ("OpenJev + 1000 本院标签切点校准", pr_cal), ("自写读出（未校准）", own), ("本院 LoRA n=1000", lora1k), ("本院 LoRA 全量", lora_all)): out.append(f"| {nm} | {ck(y,pr,weights='quadratic'):.3f} |")
out += ["", "配对差（95%CI，患者级 bootstrap）：", f"- H2：OpenJev 校准 − 本院 LoRA@1000 = {d(pr_cal, lora1k)}（非劣效界值 −0.05）", f"- H3：本院 LoRA 全量 − OpenJev 校准 = {d(lora_all, pr_cal)}", f"- OpenJev 校准 − 自写读出 = {d(pr_cal, own)}"]
g = rd("pred_gen_local_val").dropna(subset=["pred"]); gi = g.index.intersection(val)
if len(gi): ys = c.loc[gi].asa4.astype(int).values; prs = pd.Series(pr_cal, index=val).loc[gi].values; out.append(f"- H1（生成式 val 子集 n={len(gi)}）：生成式 κ {ck(ys, g.loc[gi].pred.astype(int).values, weights='quadratic'):.3f}；OpenJev 校准 κ {ck(ys, prs, weights='quadratic'):.3f}")
open(f"{R}/ojev_frozen_local.md", "w").write("\n".join(out)); print("\n".join(out))
```
