## Supplementary material for "Non-generative readout with few-label calibration versus LoRA fine-tuning of a large language model for ASA physical status classification: accuracy, speed, cost and label efficiency in three data sources": Analysis code and aggregate results: 52_ojev_scale.py

```
"""规模敏感性（预先指定）：冻结 OpenJev，Qwen3.5-9B 对 Qwen3.5-27B，均 4-bit NF4，MOVER 测试子集前 2,000 台，score 题型。
描述性对比：未校准 κ（argmax 与期望四舍五入）、宏 AUC、配对 κ 差（患者级 bootstrap），不设界值。输出 reports/ojev_scale.md"""
import os, numpy as np, pandas as pd
from sklearn.metrics import cohen_kappa_score as ck, roc_auc_score
D = os.path.expanduser("~/research/NE_ERAS_JEV/data/derived"); R = os.path.expanduser("~/research/Anes_JEV_vs_LoRA/analysis/reports")
sm = lambda a: (lambda e: e / e.sum(1, keepdims=True))(np.exp(a - a.max(1, keepdims=True)))
def ld(f): d = pd.read_csv(f"{D}/{f}.csv"); d = d[d.qtype == "score"] if "qtype" in d else d; return d.set_index("op_id")
a, b = ld("pred_ojevF_mover_sub_q4_9b"), ld("pred_ojevF_mover_sub_q4_27b"); ids = a.index.intersection(b.index); a, b = a.loc[ids], b.loc[ids]; y = a.asa4.astype(int).values
cols = [x for x in a.columns if x.startswith("c")]; Pa, Pb = sm(a[cols].values), sm(b[cols].values)
ex = lambda P: np.clip(np.rint((P * np.arange(1, 5)).sum(1)), 1, 4).astype(int); am = lambda P: P.argmax(1) + 1
pid = pd.read_csv(f"{D}/mover_pid_map.csv").set_index("op_id").pid.loc[ids].values; up = np.unique(pid); idx = {p: np.where(pid == p)[0] for p in up}
rng = np.random.default_rng(7); B = [np.concatenate([idx[p] for p in rng.choice(up, len(up))]) for _ in range(1000)]
K = lambda pr, s=slice(None): ck(y[s], pr[s], weights="quadratic"); auc = lambda P: np.mean([roc_auc_score((y == k + 1).astype(int), P[:, k]) for k in range(4)])
out = [f"# 冻结 OpenJev 规模敏感性（4-bit NF4，MOVER 子集 n={len(ids)}，患者 {len(up)}）\n", "| 模型 | κ（argmax） | κ（期望四舍五入） | 宏AUC | 预测分布(argmax) |", "|---|---|---|---|---|"]
for nm, P in (("Qwen3.5-9B", Pa), ("Qwen3.5-27B", Pb)): out.append(f"| {nm} | {K(am(P)):.3f} | {K(ex(P)):.3f} | {auc(P):.3f} | {np.bincount(am(P)-1,minlength=4).tolist()} |")
for nm, f in (("κ(argmax)", am), ("κ(期望四舍五入)", ex)):
    v = np.array([K(f(Pb), s) - K(f(Pa), s) for s in B]); out.append(f"\n- 27B − 9B，{nm}：{K(f(Pb))-K(f(Pa)):+.3f}（{np.percentile(v,2.5):+.3f} ~ {np.percentile(v,97.5):+.3f}）")
va = np.array([np.mean([roc_auc_score((y[s] == k + 1).astype(int), Pb[s, k]) for k in range(4)]) - np.mean([roc_auc_score((y[s] == k + 1).astype(int), Pa[s, k]) for k in range(4)]) for s in B[:300]])
out.append(f"- 27B − 9B，宏 AUC：{auc(Pb)-auc(Pa):+.3f}（{np.percentile(va,2.5):+.3f} ~ {np.percentile(va,97.5):+.3f}，300 次）")
open(f"{R}/ojev_scale.md", "w").write("\n".join(out)); print("\n".join(out))
```
