## Supplementary material for "Non-generative readout with few-label calibration versus LoRA fine-tuning of a large language model for ASA physical status classification: accuracy, speed, cost and label efficiency in three data sources": Analysis code and aggregate results: 54_label_format_eval.py

```
"""选项标签格式（数字/字母/罗马）探索性比较（OpenJev 后端，choice 五选项，IV 与 V 概率合并）。未校准 κ、宏 AUC、预测分布；格式间两两反转率。输出 reports/label_format.md"""
import os, numpy as np, pandas as pd
from sklearn.metrics import cohen_kappa_score as ck, roc_auc_score
D = os.path.expanduser("~/research/NE_ERAS_JEV/data/derived"); R = os.path.expanduser("~/research/Anes_JEV_vs_LoRA/analysis/reports"); out = ["# 选项标签格式（探索性；OpenJev 后端，冻结提示措辞）\n"]
for ds, dn in (("mover", "MOVER 子集前 1,500 台"), ("local", "本院 val 随机 1,000 台")):
    P = {f: pd.read_csv(f"{D}/pred_fmt_{f}_{ds}.csv") for f in ("digits", "letters", "roman")}; y = P["digits"].asa4.astype(int).values; pr = {f: d[["p1", "p2", "p3", "p4"]].values.argmax(1) + 1 for f, d in P.items()}
    out += [f"## {dn}\n", "| 格式 | κ | 宏AUC | 预测分布 |", "|---|---|---|---|"]
    for f, d in P.items(): out.append(f"| {f} | {ck(y, pr[f], weights='quadratic'):.3f} | {np.mean([roc_auc_score((y==k+1).astype(int), d[f'p{k+1}'].values) for k in range(4)]):.3f} | {np.bincount(pr[f]-1, minlength=4).tolist()} |")
    out.append("\n格式间反转率：" + "；".join(f"{a}-{b} {(pr[a]!=pr[b]).mean():.3f}" for a, b in (("digits", "letters"), ("digits", "roman"), ("letters", "roman"))) + "\n")
open(f"{R}/label_format.md", "w").write("\n".join(out)); print("\n".join(out))
```
