## Supplementary material for "Non-generative readout with few-label calibration versus LoRA fine-tuning of a large language model for ASA physical status classification: accuracy, speed, cost and label efficiency in three data sources": Analysis code and aggregate results: 55_ojlora_confirm_local.py

```
"""OpenJev+LoRA 与任务 LoRA 的同集配对比较（本院 val，患者级 cluster bootstrap，B=1000）。同一 n、同一种子。输出 reports/ojlora_confirm_local.md"""
import os, re, glob, numpy as np, pandas as pd
from sklearn.metrics import cohen_kappa_score as ck, roc_auc_score
D = os.path.expanduser("~/research/NE_ERAS_JEV/data/derived"); R = os.path.expanduser("~/research/Anes_JEV_vs_LoRA/analysis/reports"); PC = ["p1", "p2", "p3", "p4"]
pid = pd.read_csv(f"{D}/asa_dataset_v1.csv", usecols=["operation_id", "patient_sn"]).rename(columns={"operation_id": "op_id"}).set_index("op_id").patient_sn
K = lambda y, p: ck(y, p, weights="quadratic"); rows = []
for f in sorted(glob.glob(f"{D}/pred_ojlora_OJlocal*s*_local_val.csv")):
    mm = re.search(r"OJlocal(\d+)s(\d+)_local_val", f); n, sd = int(mm.group(1)), int(mm.group(2)); tf = f"{D}/pred_lora_LOC{n}s{sd}_local_val.csv"
    if not os.path.exists(tf): continue
    oj, lo = pd.read_csv(f).set_index("op_id"), pd.read_csv(tf).set_index("op_id"); ids = oj.index.intersection(lo.index); oj, lo = oj.loc[ids], lo.loc[ids]; y = oj.asa4.astype(int).values
    po, pl = oj[PC].values.argmax(1) + 1, lo[PC].values.argmax(1) + 1; p = pid.loc[ids].values; up = np.unique(p); ix = {q: np.where(p == q)[0] for q in up}
    rng = np.random.default_rng(3); B = [np.concatenate([ix[q] for q in rng.choice(up, len(up))]) for _ in range(1000)]; d = np.array([K(y[s], po[s]) - K(y[s], pl[s]) for s in B])
    au = lambda P: np.mean([roc_auc_score((y == k + 1).astype(int), P[:, k]) for k in range(4)])
    rows.append(f"| {n} | {sd} | {K(y,po):.3f} | {K(y,pl):.3f} | {d.mean():+.3f}（{np.percentile(d,2.5):+.3f} ~ {np.percentile(d,97.5):+.3f}） | {au(oj[PC].values):.3f} / {au(lo[PC].values):.3f} |")
out = ["# OpenJev+LoRA 对任务 LoRA（本院 val，同集配对）\n", "| n | 种子 | OpenJev+LoRA κ | 任务 LoRA κ | 差值（95%CI） | AUC（OJ / 任务） |", "|---|---|---|---|---|---|"] + rows
open(f"{R}/ojlora_confirm_local.md", "w").write("\n".join(out)); print("\n".join(out))
```
