## Supplementary material for "Non-generative readout with few-label calibration versus LoRA fine-tuning of a large language model for ASA physical status classification: accuracy, speed, cost and label efficiency in three data sources": Analysis code and aggregate results: 60_build_vitaldb_t2.py

```
"""VitalDB 的 T1/T2 文本与目标站点划分。T1=与 MOVER/本院一致的结构化模板（已有）；T2=T1 + 术前诊断(dx)、手术名、科室/入路/麻醉方式/是否急诊、术前心电图、肺功能、更多术前化验（均为术前信息；不含任何术中/术后变量）。
划分按患者：test 30%，train 70%（种子 20261020）。输出 NE_ERAS_JEV/data/derived/asa_common_vitaldb_t2.csv（op_id, subjectid, asa4, split, text1, text2）。"""
import os, gzip, io, numpy as np, pandas as pd
R = os.path.expanduser("~/research/NE_ERAS_JEV"); c = pd.read_csv(io.BytesIO(gzip.decompress(open(f"{R}/data/raw/vitaldb/cases.csv.gz", "rb").read())))
c = c[c.asa.isin([1, 2, 3, 4, 5])].copy(); c["asa4"] = c.asa.astype(int).clip(upper=4); c["op_id"] = "vdb" + c.caseid.astype(str)
t1 = pd.read_csv(f"{R}/data/derived/asa_common_vitaldb.csv", usecols=["op_id", "text"]).rename(columns={"text": "text1"})
f = lambda v, fmt: "unknown" if pd.isna(v) else fmt % v
def extra(r):
    return ("\nDiagnosis: %s\nDepartment: %s; surgical approach: %s; anesthesia type: %s; %s\nPreoperative ECG: %s\nPreoperative pulmonary function: %s\n"
            "Additional preoperative labs: platelets %s, prothrombin time (activity %%) %s, aPTT (sec) %s, Na %s, K %s, glucose %s, AST %s, ALT %s, BUN %s") % (
            str(r.dx).strip() if pd.notna(r.dx) else "unknown", r.department, r.approach, r.ane_type, "emergency operation" if r.emop == 1 else "elective operation",
            r.preop_ecg, r.preop_pft, f(r.preop_plt, "%.0f"), f(r.preop_pt, "%.2f"), f(r.preop_aptt, "%.1f"), f(r.preop_na, "%.0f"), f(r.preop_k, "%.1f"), f(r.preop_gluc, "%.0f"), f(r.preop_ast, "%.0f"), f(r.preop_alt, "%.0f"), f(r.preop_bun, "%.1f"))
c["ex"] = [extra(r) for r in c.itertuples()]; c = c.merge(t1, on="op_id"); c["text2"] = c.text1 + c.ex
subj = np.sort(c.subjectid.unique()); rng = np.random.RandomState(20261020); test = set(rng.choice(subj, int(len(subj) * 0.3), replace=False)); c["split"] = np.where(c.subjectid.isin(test), "test", "train")
out = c[["op_id", "subjectid", "asa4", "split", "text1", "text2"]]; p = f"{R}/data/derived/asa_common_vitaldb_t2.csv"; out.to_csv(p, index=False, encoding="utf-8-sig"); os.chmod(p, 0o600)
print(out.groupby("split").agg(n=("op_id", "size"), pat=("subjectid", "nunique")), out.groupby("split").asa4.value_counts(normalize=True).unstack().round(3)); print(out.text2.iloc[3])
```
