## Supplementary material for "Non-generative readout with few-label calibration versus LoRA fine-tuning of a large language model for ASA physical status classification: accuracy, speed, cost and label efficiency in three data sources": Analysis code and aggregate results: 61_vdb_run.py

```
"""VitalDB 目标站点实验（T1/T2 两种输入；划分与文本见 60_build_vitaldb_t2.py）。
 zs  <t1|t2> <gpu>               自写读出零样本（提示 P0，字母 A–D）→ pred_vdb_zs_<t>_{test,cal}.csv（cal=训练划分内 1000 台，种子 11）
 oj  <t1|t2> <gpu>               冻结 OpenJev（读自 openjev_tuning_choice.json）→ pred_vdb_oj_<t>_{test,cal}.csv
 train <t1|t2> <n> <seed> <gpu> <tag>   目标站点任务 LoRA（与 08_lora.py 同配方：4-bit QLoRA r=16, lr 1e-4, 1 epoch, 批 16, 末位 A–D logits 交叉熵）
 infer <tag> <t1|t2> <gpu>       → pred_vdb_lora_<tag>_test.csv"""
import os, sys, json, time, numpy as np, pandas as pd, torch
mode = sys.argv[1]; D = os.path.expanduser("~/research/NE_ERAS_JEV/data/derived"); M = os.path.expanduser("~/research/Anes_JEV_vs_LoRA/models"); R = os.path.expanduser("~/research/Anes_JEV_vs_LoRA/analysis/reports")
V = pd.read_csv(f"{D}/asa_common_vitaldb_t2.csv"); path = "/mnt/c/backup/Qwen3.5-9B"
def part(tv, which):
    d = V.assign(text=V["text1" if tv == "t1" else "text2"]); tr = d[d.split == "train"]
    return d[d.split == "test"] if which == "test" else tr.sample(1000, random_state=11)
P = ("You are an anesthesiologist. Based on the preoperative information below, assign the ASA Physical Status class.\n\n%s\n\nOptions:\nA. ASA I\nB. ASA II\nC. ASA III\nD. ASA IV-V\nAnswer:")
def flush(rows, name): pd.DataFrame(rows).to_csv(f"{D}/{name}.csv", index=False); os.chmod(f"{D}/{name}.csv", 0o600)
if mode == "zs":
    tv, gpu = sys.argv[2], int(sys.argv[3]); from transformers import AutoTokenizer, AutoModelForCausalLM
    tok = AutoTokenizer.from_pretrained(path); m = AutoModelForCausalLM.from_pretrained(path, dtype=torch.float16, device_map={"": gpu}).eval()
    a1 = [tok.encode(" " + c, add_special_tokens=False)[-1] for c in "ABCD"]; a2 = [tok.encode(c, add_special_tokens=False)[-1] for c in "ABCD"]
    for which in ("test", "cal"):
        rows = []
        for r in part(tv, which).itertuples():
            x = tok(P % r.text, return_tensors="pt").to(f"cuda:{gpu}")
            with torch.no_grad(): lg = m(**x).logits[0, -1].float()
            p = torch.softmax(torch.maximum(lg[a1], lg[a2]), 0).cpu().numpy(); rows.append(dict(op_id=r.op_id, asa4=r.asa4, p1=p[0], p2=p[1], p3=p[2], p4=p[3]))
        flush(rows, f"pred_vdb_zs_{tv}_{which}"); print("zs", tv, which, len(rows), flush=True)
elif mode == "oj":
    tv, gpu = sys.argv[2], int(sys.argv[3]); from openjev import SystemOneEngine, load_backend; from openjev.schema import SystemOneRequest; from openjev.prompting import PromptConfig
    ns = {}; exec("SYS =" + open(os.path.expanduser("~/research/Anes_JEV_vs_LoRA/analysis/scripts/33_openjev_tune.py")).read().split("SYS =", 1)[1].split("sm = lambda")[0], ns)
    CH = json.load(open(f"{R}/openjev_tuning_choice.json"))["selected"]; QT = CH["qtype"]
    eng = SystemOneEngine(backend=load_backend(path, device=f"cuda:{gpu}", dtype=torch.float16), model_name="openjev")
    eng.prompt_config = PromptConfig() if ns["SYS"][CH["system"]] is None else PromptConfig(system_prompt=ns["SYS"][CH["system"]])
    Q = {QT: ns["questions"](ns["SHORT"] if CH["desc"] == "short" else ns["OFFIC"])[QT]}
    for which in ("test", "cal"):
        rows = []; t0 = time.time()
        for i, r in enumerate(part(tv, which).itertuples()):
            raws, _ = eng.evaluate_raw(SystemOneRequest.model_validate({"state": r.text, "questions": Q})); lp = raws[0].logprobs
            rows.append(dict(op_id=r.op_id, asa4=r.asa4, qtype=QT, **{f"c{k}": lp[k] for k in range(4)}))
            if (i + 1) % 250 == 0: print("oj", tv, which, i + 1, f"{(time.time()-t0)/(i+1):.2f}s/例", flush=True)
        flush(rows, f"pred_vdb_oj_{tv}_{which}")
else:
    from transformers import AutoTokenizer, AutoModelForCausalLM, BitsAndBytesConfig; from peft import LoraConfig, get_peft_model, PeftModel, prepare_model_for_kbit_training
    tok = AutoTokenizer.from_pretrained(path); tok.padding_side = "left"; ids = torch.tensor([tok.encode(" " + c, add_special_tokens=False)[-1] for c in "ABCD"])
    bnb = BitsAndBytesConfig(load_in_4bit=True, bnb_4bit_quant_type="nf4", bnb_4bit_compute_dtype=torch.float16, bnb_4bit_use_double_quant=True)
    def fwd(m, texts, dev): x = tok([P % t for t in texts], return_tensors="pt", padding=True).to(dev); return m(**x, logits_to_keep=1).logits[:, -1].float()[:, ids.to(dev)]
    if mode == "train":
        tv, n, seed, gpu, tag = sys.argv[2], int(sys.argv[3]), int(sys.argv[4]), int(sys.argv[5]), sys.argv[6]; dev = f"cuda:{gpu}"; torch.manual_seed(seed)
        d = V.assign(text=V["text1" if tv == "t1" else "text2"]); d = d[d.split == "train"]; d = (d.sample(n, random_state=seed) if n < len(d) else d.sample(frac=1, random_state=seed)).reset_index(drop=True); n = len(d)
        m = prepare_model_for_kbit_training(AutoModelForCausalLM.from_pretrained(path, quantization_config=bnb, dtype=torch.float16, device_map={"": gpu}), use_gradient_checkpointing=True)
        m = get_peft_model(m, LoraConfig(r=16, lora_alpha=32, lora_dropout=0.05, target_modules=["q_proj", "k_proj", "v_proj", "o_proj", "gate_proj", "up_proj", "down_proj"], task_type="CAUSAL_LM"))
        opt = torch.optim.AdamW([p for p in m.parameters() if p.requires_grad], lr=1e-4); bs, acc = 4, 4; steps = max(n // 16, 1)
        sch = torch.optim.lr_scheduler.OneCycleLR(opt, max_lr=1e-4, total_steps=steps, pct_start=0.05, anneal_strategy="cos"); m.train(); t0 = time.time(); la = []
        for s in range(steps):
            for a in range(acc):
                b = d.iloc[(s * acc + a) * bs:(s * acc + a + 1) * bs]
                loss = torch.nn.functional.cross_entropy(fwd(m, list(b.text), dev), torch.tensor(b.asa4.values - 1, device=dev)) / acc; loss.backward(); la.append(loss.item() * acc)
            torch.nn.utils.clip_grad_norm_(m.parameters(), 1.0); opt.step(); sch.step(); opt.zero_grad()
            if (s + 1) % 10 == 0: print(f"step {s+1}/{steps} loss {np.mean(la[-40:]):.4f} {(time.time()-t0)/(s+1):.1f}s/step", flush=True)
        m.save_pretrained(f"{M}/{tag}"); print("训练完成", f"{time.time()-t0:.0f}s")
    else:
        tag, tv, gpu = sys.argv[2], sys.argv[3], int(sys.argv[4]); dev = f"cuda:{gpu}"
        m = PeftModel.from_pretrained(AutoModelForCausalLM.from_pretrained(path, quantization_config=bnb, dtype=torch.float16, device_map={"": gpu}), f"{M}/{tag}").eval(); d = part(tv, "test").reset_index(drop=True); rows = []
        for i in range(0, len(d), 8):
            b = d.iloc[i:i + 8]
            with torch.no_grad(): p = torch.softmax(fwd(m, list(b.text), dev), -1).cpu().numpy()
            rows += [dict(op_id=r.op_id, asa4=r.asa4, p1=q[0], p2=q[1], p3=q[2], p4=q[3]) for r, q in zip(b.itertuples(), p)]
        flush(rows, f"pred_vdb_lora_{tag}_test"); print("完成", len(rows))
```
