## Supplementary material for "Non-generative readout with few-label calibration versus LoRA fine-tuning of a large language model for ASA physical status classification: accuracy, speed, cost and label efficiency in three data sources": Analysis code and aggregate results: 80_fig_cost.py

```
"""成本-精度图：左=每例延迟(对数)与 κ（本院 val；生成式 999 台子集）；右=κ 随标签数（校准读出 vs LoRA vs XGBoost），本院与 MOVER。数据来自 reports/ 与 latency.tsv 的已有数字。"""
import matplotlib; matplotlib.use("Agg")
import matplotlib.pyplot as plt
from matplotlib import font_manager as fm
for f in ("/usr/share/fonts/opentype/noto/NotoSansCJK-Regular.ttc", "/usr/share/fonts/opentype/noto/NotoSerifCJK-Regular.ttc", "/usr/share/fonts/opentype/noto/NotoSerifCJK-Bold.ttc"):
    if __import__("os").path.exists(f): fm.fontManager.addfont(f)
plt.rcParams["font.sans-serif"] = ["Noto Sans CJK SC", "Noto Serif CJK SC", "DejaVu Sans"]; plt.rcParams["axes.unicode_minus"] = False
fig, ax = plt.subplots(1, 3, figsize=(15, 4.4))
pts = [("生成式零样本", 7067, 0.227, "#b04a4a"), ("读出零样本", 111, 0.443, "#4a78b0"), ("读出+100标签校准", 111, 0.526, "#2c5aa0"), ("读出+1000标签校准", 111, 0.534, "#1d3f73"), ("LoRA n=1000", 179, 0.559, "#6a8f3c"), ("LoRA 全量", 179, 0.561, "#3f6a1e")]
for n, x, y, c in pts: ax[0].scatter(x * (1.0 + 0.0), y, s=70, color=c, zorder=3, label=f"{n}  κ={y:.3f}")
ax[0].legend(fontsize=7.5, loc="center right"); ax[0].set_xscale("log"); ax[0].set_xlabel("单例推理延迟 p50 (ms，批=1，对数轴)"); ax[0].set_ylabel("加权 κ（本院 val）"); ax[0].set_title("速度 vs 精度"); ax[0].grid(alpha=.3)
def curve(a, title, kc, kk, lc, ln, xg, xv):
    a.plot([1] + kc, [kk[0]] + kk[1:], "o-", color="#2c5aa0", label="读出+k标签校准（无训练）")
    a.plot(lc, ln, "s-", color="#3f6a1e", label="任务 LoRA（n 个标签训练）")
    a.set_xscale("log"); a.set_xlabel("本地标签数（对数轴；k=0 画在 1）"); a.set_ylabel("加权 κ"); a.set_title(title); a.grid(alpha=.3); a.legend(fontsize=8, loc="lower right")
curve(ax[1], "本院 val", [25, 50, 100, 200, 500, 1000], [0.443, 0.491, 0.516, 0.526, 0.528, 0.535, 0.540], [100, 300, 1000, 4326], [0.423, 0.542, 0.559, 0.561], None, None)
curve(ax[2], "MOVER 测试子集", [25, 50, 100, 200, 500, 1000], [0.473, 0.467, 0.495, 0.508, 0.517, 0.521, 0.524], [100, 300, 1000, 3000, 12000], [0.410, 0.411, 0.505, 0.563, 0.583], None, None)
plt.tight_layout(); plt.savefig("figures/fig_cost_efficiency.png", dpi=160); print("ok")
```
