## Supplementary material for "Non-generative readout with few-label calibration versus LoRA fine-tuning of a large language model for ASA physical status classification: accuracy, speed, cost and label efficiency in three data sources": Analysis code and aggregate results: 81_hallucination_audit.py

```
"""生成式零样本的“幻觉”粗查（规则法）：对输出的理由文本，检查是否肯定性地提到输入中没有或与输入矛盾的合并症。
分类：contradicted=输入明确写 no（高血压/冠心病/糖尿病）而理由肯定提到；unsupported=输入模板里没有该信息（心衰/COPD/肾病/吸烟/癌症/贫血/肥胖等，按可用数值判断支持度）而理由肯定提到。
否定语境（no/without/denies/absence/not/free of 等出现在前 40 字符）不计。仅为规则法粗估，另抽样人工核对。输出 reports/hallucination_audit.md 与抽样表。
同时统计输出不可解析率（非生成式读出为 0，因输出限定在闭集内）。"""
import os, re, numpy as np, pandas as pd
D = os.path.expanduser("~/research/NE_ERAS_JEV/data/derived"); R = os.path.expanduser("~/research/Anes_JEV_vs_LoRA/analysis/reports")
NEG = re.compile(r"(\bno\b|\bwithout\b|\bdenies\b|\babsence\b|\bnot\b|free of|negative for|\bnor\b|\bnone\b|unremarkable|no history|lack)", re.I)
CL = re.compile(r"[.;]|\b(but|although|however|while|whereas|yet|though|except)\b", re.I)
def mentions(txt, pat):
    """子句级否定：以句号/分号/转折词切分子句；子句内提及之前出现否定词则视为否定。返回每次提及是否为肯定性。"""
    out = []; pos = 0
    for part in CL.split(txt):
        if part is None: continue
        pass
    # 重新按位置切分
    bounds = [0] + [m.end() for m in CL.finditer(txt)] + [len(txt)]
    for a, b in zip(bounds[:-1], bounds[1:]):
        clause = txt[a:b]
        for m in re.finditer(pat, clause, re.I):
            out.append(not NEG.search(clause[:m.start()]))
    return out

def flags(rat, inp):
    g = lambda k: (re.search(rf"{k} (yes|no)", inp) or [None, None])[1]
    cr = re.search(r"creatinine ([\d\.]+)", inp); hb = re.search(r"hemoglobin ([\d\.]+)", inp); bmi = re.search(r"BMI: ([\d\.]+)", inp)
    cr = float(cr.group(1)) if cr else None; hb = float(hb.group(1)) if hb else None; bmi = float(bmi.group(1)) if bmi else None
    proc = (re.search(r"Planned procedure: (.*)", inp) or [None, ""])[1] if True else ""
    res = []
    for nm, pat, key in (("diabetes", r"diabet", "diabetes"), ("hypertension", r"hypertens", "hypertension"), ("coronary", r"coronary|\bCAD\b|angina|ischemic heart|myocardial infarct", "coronary artery disease")):
        v = g(key)
        if any(mentions(rat, pat)) and v == "no": res.append(("contradicted", nm))
    sup = {"heart failure": (r"heart failure|cardiac failure|\bCHF\b", False), "COPD/lung disease": (r"COPD|chronic obstructive|asthma|pulmonary disease|lung disease", False),
           "renal disease": (r"renal (disease|failure|impairment|insufficiency)|kidney (disease|failure)|\bCKD\b", cr is not None and cr > 1.5), "smoking": (r"smok", False),
           "anemia": (r"anemi", hb is not None and hb < 12), "obesity": (r"obes", bmi is not None and bmi >= 30), "cancer": (r"cancer|malignan|carcinoma|neoplasm|tumou?r", bool(re.search(r"(?i)cancer|carcinoma|tumou?r|malig|resection|gastrectomy|colectomy|mastectomy|prostatectomy|nephrectomy|hepatectomy|Whipple|radical", proc))),
           "sepsis/shock": (r"sepsis|septic|shock", False)}
    for nm, (pat, supported) in sup.items():
        if any(mentions(rat, pat)) and not supported: res.append(("unsupported", nm))
    return res
out = ["# 生成式零样本理由文本的“幻觉”粗查（规则法）\n", "规则法只能给出下限/粗估，且“unsupported”含义是“输入模板里没有支持该说法的信息”，不等同于临床上一定错误；抽样人工核对见文末。\n", "| 数据 | 理由条数 | 输出不可解析 | 含矛盾说法 | 含无依据说法 | 二者任一 |", "|---|---|---|---|---|---|"]
samples = []
for nm, gf, cf, key in (("MOVER 子集", "pred_gen_mover_sub", "asa_common_mover", "op_id"), ("本院 val 子集（T1 英文模板）", "pred_gen_local_val", "asa_common_local", "op_id")):
    g = pd.read_csv(f"{D}/{gf}.csv"); c = pd.read_csv(f"{D}/{cf}.csv", usecols=["op_id", "text"]); x = g.merge(c, on="op_id"); n = len(x); bad = x.pred.isna().sum(); con = unsup = anyc = 0
    for r in x.itertuples():
        fl = flags(str(r.raw), r.text); k1 = any(a == "contradicted" for a, _ in fl); k2 = any(a == "unsupported" for a, _ in fl); con += k1; unsup += k2; anyc += (k1 or k2)
        if (k1 or k2) and len(samples) < 400: samples.append((nm, r.op_id, "; ".join(f"{a}:{b}" for a, b in fl), str(r.raw)[:260], r.text.split("\n")[2] if "\n" in r.text else ""))
    out.append(f"| {nm} | {n} | {bad}（{bad/n:.1%}） | {con}（{con/n:.1%}） | {unsup}（{unsup/n:.1%}） | {anyc}（{anyc/n:.1%}） |")
s = pd.DataFrame(samples, columns=["数据", "op_id", "标记", "理由(截断)", "输入History行"]); s.sample(min(40, len(s)), random_state=1).to_csv(f"{R}/hallucination_audit_sample.csv", index=False, encoding="utf-8-sig")
open(f"{R}/hallucination_audit.md", "w").write("\n".join(out)); print("\n".join(out))
```
