## Supplementary material for "Non-generative readout with few-label calibration versus LoRA fine-tuning of a large language model for ASA physical status classification: accuracy, speed, cost and label efficiency in three data sources": Analysis code and aggregate results: 82_calibration_check.py

```
"""校准度检查（“已校准概率”这一卖点）：ECE（10 等宽箱，取最大概率）、多类 Brier；原始概率 vs 单参数温度缩放（温度在 MOVER 校准集 / 本院 dev 1000 标签上拟合）。
对象：自写读出、冻结 OpenJev（score 题）、任务 LoRA。输出 reports/calibration_check.md"""
import os, numpy as np, pandas as pd
from scipy.optimize import minimize_scalar
D = os.path.expanduser("~/research/NE_ERAS_JEV/data/derived"); R = os.path.expanduser("~/research/Anes_JEV_vs_LoRA/analysis/reports"); PC = ["p1", "p2", "p3", "p4"]
sm = lambda a, T=1.0: (lambda z: np.exp(z) / np.exp(z).sum(1, keepdims=True))((a - a.max(1, keepdims=True)) / T)
def ece(P, y):
    conf, ok = P.max(1), P.argmax(1) + 1 == y; e = 0
    for lo in np.linspace(0, 1, 11)[:-1]:
        s = (conf > lo) & (conf <= lo + 0.1)
        if s.any(): e += s.mean() * abs(ok[s].mean() - conf[s].mean())
    return e
brier = lambda P, y: ((P - np.eye(4)[y - 1]) ** 2).sum(1).mean()
nll = lambda P, y: -np.log(np.clip(P[np.arange(len(y)), y - 1], 1e-12, 1)).mean()
def fitT(L, y): return minimize_scalar(lambda T: nll(sm(L, T), y), bounds=(0.05, 20), method="bounded").x
rd = lambda f: pd.read_csv(f"{D}/{f}.csv").set_index("op_id")
def logits_from_p(d): return np.log(np.clip(d[PC].values, 1e-9, 1))
def oj(f):
    d = pd.read_csv(f"{D}/{f}.csv"); d = d[d.qtype == "score"] if "qtype" in d else d; d = d.set_index("op_id"); return d[[c for c in d.columns if c.startswith("c")]].values, d
out = ["# 概率校准度（ECE↓、Brier↓；原始 vs 温度缩放）\n", "| 数据 | 臂 | ECE 原始（95% CI） | ECE 温度缩放 | Brier 原始 | Brier 温度缩放 | 温度 T |", "|---|---|---|---|---|---|---|"]
sub = set(pd.read_csv(f"{D}/mover_test_sub4000.csv").op_id); loc = pd.read_csv(f"{D}/asa_common_local.csv", usecols=["op_id", "split", "asa4"]).set_index("op_id")
# MOVER：校准集 = pred_*_mover_cal；测试 = sub
PM = pd.read_csv(f"{D}/mover_pid_map.csv").set_index("op_id").pid; PL = pd.read_csv(f"{D}/asa_dataset_v1.csv", usecols=["operation_id", "patient_sn"]).rename(columns={"operation_id": "op_id"}).set_index("op_id").patient_sn
def row(ds, nm, Lc, yc, Lt, yt, ids):
    T = fitT(Lc, yc); P0, P1 = sm(Lt), sm(Lt, T); pid = (PM if ds.startswith("MOVER") else PL).loc[list(ids)].values; up = np.unique(pid); ix = {q: np.where(pid == q)[0] for q in up}; rng = np.random.default_rng(5); bs = [np.concatenate([ix[q] for q in rng.choice(up, len(up))]) for _ in range(300)]
    def ci(f, P): v = np.array([f(P[b], yt[b]) for b in bs]); return f"{f(P, yt):.3f}（{np.percentile(v, 2.5):.3f}–{np.percentile(v, 97.5):.3f}）"
    out.append(f"| {ds} | {nm} | {ci(ece, P0)} | {ci(ece, P1)} | {ci(brier, P0)} | {ci(brier, P1)} | {T:.2f} |")
c, t = rd("pred_llm_mover_cal"), rd("pred_llm_mover_test"); t = t[t.index.isin(sub)]; row("MOVER 子集", "自写读出", logits_from_p(c), c.asa4.astype(int).values, logits_from_p(t), t.asa4.astype(int).values, t.index)
Lc, dc = oj("pred_ojevF_mover_cal"); Lt, dt = oj("pred_ojevF_mover_sub"); row("MOVER 子集", "冻结 OpenJev", Lc, dc.asa4.astype(int).values, Lt, dt.asa4.astype(int).values, dt.index)
l = rd("pred_lora_curve1000_mover_sub"); row("MOVER 子集", "任务 LoRA n=1000（自身校准集=其测试集的一半折半）", logits_from_p(l.iloc[: len(l) // 2]), l.asa4.astype(int).values[: len(l) // 2], logits_from_p(l.iloc[len(l) // 2:]), l.asa4.astype(int).values[len(l) // 2:], l.index[len(l) // 2:])
dev = loc[loc.split == "dev"].sample(1000, random_state=11).index; val = loc[loc.split == "val"].index
d = rd("pred_llm_local"); row("本院 val", "自写读出", logits_from_p(d.loc[dev]), loc.loc[dev].asa4.astype(int).values, logits_from_p(d.loc[val]), loc.loc[val].asa4.astype(int).values, val)
Lall, dall = oj("pred_ojevF_local"); ix = pd.Series(range(len(dall)), index=dall.index); row("本院 val", "冻结 OpenJev", Lall[ix.loc[dev].values], loc.loc[dev].asa4.astype(int).values, Lall[ix.loc[val].values], loc.loc[val].asa4.astype(int).values, val)
l = rd("pred_lora_LOC1000s20261017_local_val"); h = len(l) // 2; row("本院 val", "任务 LoRA n=1000（折半）", logits_from_p(l.iloc[:h]), l.asa4.astype(int).values[:h], logits_from_p(l.iloc[h:]), l.asa4.astype(int).values[h:], l.index[h:])
open(f"{R}/calibration_check.md", "w").write("\n".join(out)); print("\n".join(out))
```
