## Supplementary material for "Non-generative readout with few-label calibration versus LoRA fine-tuning of a large language model for ASA physical status classification: accuracy, speed, cost and label efficiency in three data sources": Analysis code and aggregate results: 83_guidance.py

```
"""自然语言指导实验（开题卖点“可用自然语言指导”）：同一批本院病例，在读出臂提示前加入不同的自然语言指导，看输出分布与 κ 的变化；无训练、无标签。
变体（事先写定，不看 val 结果修改措辞）：G0 无指导；GOFF ASA 2020 官方定义（含举例）；GLOC 本院记录习惯（依据课题负责人陈述的偏倚：少评 I 级、老年器质性改变倾向 III，其余为一般临床常识）；GSTR 严格按指南（与 GLOC 方向相反）。
数据：本院 dev 随机 800 台（种子 41，仅作方向核查）与 val 全部 2,753 台；输入 T1（结构化模板）与 T2（原始访视文本）。输出 pred_guid_<G>_<t>_<ds>.csv。用法：83_guidance.py <gpu>"""
import os, sys, time, numpy as np, pandas as pd, torch
from transformers import AutoTokenizer, AutoModelForCausalLM
gpu = int(sys.argv[1]); D = os.path.expanduser("~/research/NE_ERAS_JEV/data/derived"); path = "/mnt/c/backup/Qwen3.5-9B"
G = {"G0": "",
     "GOFF": ("Definitions (ASA Physical Status, 2020):\nI: a normal healthy patient (healthy, non-smoking, no or minimal alcohol use).\nII: mild systemic disease without substantive functional limitation (current smoker, social drinker, pregnancy, obesity 30<BMI<40, well-controlled DM/HTN, mild lung disease).\n"
              "III: severe systemic disease with substantive functional limitation (poorly controlled DM or HTN, COPD, morbid obesity BMI>=40, active hepatitis, alcohol dependence, implanted pacemaker, moderate reduction of ejection fraction, ESRD on regular dialysis, history >3 months of MI, CVA, TIA or coronary stents, malignancy).\n"
              "IV: severe systemic disease that is a constant threat to life (recent <3 months MI, CVA, TIA or stents, ongoing cardiac ischemia, severe valve dysfunction, severe reduction of ejection fraction, shock, sepsis, DIC, ARDS, ESRD not on regular dialysis).\nV: moribund patient not expected to survive without the operation.\n"),
     "GLOC": ("Guidance: classify the way anesthesiologists at this hospital routinely record ASA for abdominal tumor surgery. Their habits: ASA I is rarely recorded; a patient who looks healthy but is undergoing tumor or major abdominal surgery is usually recorded as II. "
              "Age-related organic changes in elderly patients (atherosclerotic plaques, lacunar infarcts, mild valvular regurgitation, diastolic dysfunction, mild obstructive or interstitial lung changes) are usually recorded as III. Well-controlled hypertension or non-insulin diabetes alone is II; "
              "uncontrolled disease, several comorbid systems, insulin-treated diabetes, coronary artery disease or moderate lung disease is III; ASA IV is reserved for recent MI or stroke, shock, sepsis, severe cardiac, renal or hepatic failure.\n"),
     "GSTR": ("Guidance: follow the ASA Physical Status definitions strictly and literally. A patient without any systemic disease is ASA I even if the operation is major or the patient is an adult of any age. Do not upgrade a class because of age, cancer status or the size of the surgery alone. "
              "Mild or well-controlled single-system disease is ASA II; only disease with real functional limitation is ASA III; ASA IV requires a constant threat to life.\n")}
P = "You are an anesthesiologist. Based on the preoperative information below, assign the ASA Physical Status class.\n\n%s%s\n\nOptions:\nA. ASA I\nB. ASA II\nC. ASA III\nD. ASA IV-V\nAnswer:"
c = pd.read_csv(f"{D}/asa_common_local.csv"); raw = pd.read_csv(f"{D}/asa_dataset_v1.csv", usecols=["operation_id", "text"]).rename(columns={"operation_id": "op_id", "text": "t2"}); c = c.merge(raw, on="op_id")
dev = c[c.split == "dev"].sample(800, random_state=41); val = c[c.split == "val"]
tok = AutoTokenizer.from_pretrained(path); m = AutoModelForCausalLM.from_pretrained(path, dtype=torch.float16, device_map={"": gpu}).eval()
a1 = [tok.encode(" " + x, add_special_tokens=False)[-1] for x in "ABCD"]; a2 = [tok.encode(x, add_special_tokens=False)[-1] for x in "ABCD"]
for ds, d in (("dev800", dev), ("val", val)):
    for tv in ("t1", "t2"):
        for g, gt in G.items():
            out = f"{D}/pred_guid_{g}_{tv}_{ds}.csv"
            if os.path.exists(out): continue
            rows = []; t0 = time.time()
            for r in d.itertuples():
                txt = r.text if tv == "t1" else r.t2; x = tok(P % (gt + ("\n" if gt else ""), txt), return_tensors="pt").to(f"cuda:{gpu}")
                with torch.no_grad(): lg = m(**x).logits[0, -1].float()
                p = torch.softmax(torch.maximum(lg[a1], lg[a2]), 0).cpu().numpy(); rows.append(dict(op_id=r.op_id, asa4=r.asa4, p1=p[0], p2=p[1], p3=p[2], p4=p[3]))
            pd.DataFrame(rows).to_csv(out, index=False); print(ds, tv, g, len(rows), f"{time.time()-t0:.0f}s", flush=True)
```
