## Supplementary material for "Non-generative readout with few-label calibration versus LoRA fine-tuning of a large language model for ASA physical status classification: accuracy, speed, cost and label efficiency in three data sources": Analysis code and aggregate results: 84_vdb_report.py

```
"""VitalDB 目标站点重复验证（探索性；测试集 1,885 台，患者为 cluster）：零样本读出/冻结 OpenJev（T1/T2）、k 标签切点校准（k=100 与 1000，来自训练划分内标签）、目标站点任务 LoRA（T2：n=100/300/1000/3000；T1：n=1000/3000）。
主对比：同标签量下 校准读出 − 目标站点 LoRA（δ=−0.05 非劣效）；LoRA 3000 − 校准读出(k=1000)。输出 reports/vitaldb_target_site.md"""
import os, glob, re, numpy as np, pandas as pd
from sklearn.metrics import cohen_kappa_score as ck, roc_auc_score
D = os.path.expanduser("~/research/NE_ERAS_JEV/data/derived"); R = os.path.expanduser("~/research/Anes_JEV_vs_LoRA/analysis/reports"); PC = ["p1", "p2", "p3", "p4"]
V = pd.read_csv(f"{D}/asa_common_vitaldb_t2.csv", usecols=["op_id", "subjectid", "split"]).set_index("op_id"); test_ids = V[V.split == "test"].index
sm = lambda a: (lambda e: e / e.sum(1, keepdims=True))(np.exp(a - a.max(1, keepdims=True)))
K = lambda y, p: ck(y, p, weights="quadratic"); rd = lambda f: pd.read_csv(f"{D}/{f}.csv").set_index("op_id") if os.path.exists(f"{D}/{f}.csv") else None
def expscore_zs(d): return (d[PC].values * np.arange(1, 5)).sum(1)
def expscore_oj(d): c = [x for x in d.columns if x.startswith("c")]; return (sm(d[c].values) * np.arange(1, 5)).sum(1)
def cuts_fit(E, y):
    cu = np.array([1.5, 2.5, 3.5]); g = np.linspace(1.0, 4.0, 121)
    for _ in range(3):
        for j in range(3): cu[j] = max(g, key=lambda v: K(y, 1 + (E[:, None] > np.where(np.arange(3) == j, v, cu)).sum(1)))
    return cu
out = ["# VitalDB 目标站点重复验证（探索性；test n=%d，患者 %d）\n" % (len(test_ids), V.loc[test_ids].subjectid.nunique())]
ytest = None; arms = {}   # 名称 -> 预测类别（对 test_ids 顺序）
rng = np.random.default_rng(5)
for tv in ("t1", "t2"):
    for key, nm, ef in (("zs", "自写读出", expscore_zs), ("oj", "冻结 OpenJev", expscore_oj)):
        t, c = rd(f"pred_vdb_{key}_{tv}_test"), rd(f"pred_vdb_{key}_{tv}_cal")
        if t is None or c is None: continue
        t = t.loc[test_ids] if t.index.isin(test_ids).all() else t[t.index.isin(test_ids)]; ytest = t.asa4.astype(int).values; ids = t.index
        raw = (t[PC].values.argmax(1) + 1) if key == "zs" else (sm(t[[x for x in t.columns if x.startswith("c")]].values).argmax(1) + 1)
        arms[f"{nm} {tv.upper()} 未校准"] = (ids, raw); Ec, yc, Et = ef(c), c.asa4.astype(int).values, ef(t)
        arms[f"{nm} {tv.upper()} +1000标签切点校准"] = (ids, 1 + (Et[:, None] > cuts_fit(Ec, yc)).sum(1))
        ks = []
        for _ in range(60):
            s = rng.choice(len(yc), 100, replace=False); ks.append(K(ytest, 1 + (Et[:, None] > cuts_fit(Ec[s], yc[s])).sum(1)))
        arms[f"{nm} {tv.upper()} +100标签切点校准(60次均值)"] = (ids, None, float(np.mean(ks)))
curve = {}
for f in sorted(glob.glob(f"{D}/pred_vdb_lora_VDB_t*_test.csv")):
    mm = re.search(r"VDB_(t\d)_(\d+)_(\d+)_test", f); tv, n, sd = mm.group(1), int(mm.group(2)), int(mm.group(3)); d = rd(os.path.basename(f)[:-4]); curve.setdefault((tv, n), {})[sd] = d
pid = V.subjectid
def boot(ids, y, fn, B=1000):
    p = pid.loc[ids].values; up = np.unique(p); ix = {q: np.where(p == q)[0] for q in up}; g = np.random.default_rng(9); return [fn(np.concatenate([ix[q] for q in g.choice(up, len(up))])) for _ in range(B)]
out += ["## 各臂 κ（对 VitalDB 麻醉记录 ASA）\n", "| 臂 | κ |", "|---|---|"]
for nm, v in arms.items():
    if len(v) == 3: out.append(f"| {nm} | {v[2]:.3f} |")
    else: out.append(f"| {nm} | {K(ytest, v[1]):.3f} |")
out += ["", "## 目标站点 LoRA（κ，种子均值 [范围]）\n", "| 输入 | n | κ | 种子数 |", "|---|---|---|---|"]
for (tv, n), dd in sorted(curve.items()):
    ks = [K(d.loc[test_ids].asa4.astype(int).values, d.loc[test_ids][PC].values.argmax(1) + 1) for d in dd.values()]; out.append(f"| {tv.upper()} | {n} | {np.mean(ks):.3f} [{min(ks):.3f}–{max(ks):.3f}] | {len(ks)} |")
def contrast(a, tv, n, label, sign=1):
    key = [k for k in arms if k == a]
    if not key or (tv, n) not in curve: return
    d0 = curve[(tv, n)].get(20261017) if 20261017 in curve[(tv, n)] else list(curve[(tv, n)].values())[0]; ids, pr = arms[key[0]][0], arms[key[0]][1]
    if pr is None: return
    l = d0.loc[ids]; y = l.asa4.astype(int).values; pl = l[PC].values.argmax(1) + 1; b = boot(ids, y, lambda s: sign * (K(y[s], pr[s]) - K(y[s], pl[s])))
    out.append(f"- {label}：{sign*(K(y,pr)-K(y,pl)):+.3f}（{np.percentile(b,2.5):+.3f} ~ {np.percentile(b,97.5):+.3f}）")
out += ["", "## 配对对比（种子 20261017；患者级 bootstrap）"]
for tv in ("T1", "T2"):
    for nm in ("自写读出", "冻结 OpenJev"):
        contrast(f"{nm} {tv} +1000标签切点校准", tv.lower(), 1000, f"{nm} {tv} 校准(1000) − 目标站点 LoRA@1000（非劣效 δ=−0.05）")
        contrast(f"{nm} {tv} +1000标签切点校准", tv.lower(), 3000, f"{nm} {tv} 校准(1000) − 目标站点 LoRA@3000", 1)
open(f"{R}/vitaldb_target_site.md", "w").write("\n".join(out)); print("\n".join(out))
```
