## Supplementary material for "Non-generative readout with few-label calibration versus LoRA fine-tuning of a large language model for ASA physical status classification: accuracy, speed, cost and label efficiency in three data sources": Analysis code and aggregate results: 85_guidance_report.py

```
"""自然语言指导实验报告：G0/GOFF/GLOC/GSTR × T1/T2，本院 val（n=2753）。指标：κ(对本院记录)、预测分布（I/II/III/IV–V 占比，与原记录对照）、指导相对 G0 的分布位移、
指导后再用 100 个 dev 标签做切点校准（dev800 内抽 100，60 次均值）的 κ；对照本院 LoRA 全量与读出+校准。患者级 cluster bootstrap 给 κ 差。输出 reports/guidance_report.md"""
import os, numpy as np, pandas as pd
from sklearn.metrics import cohen_kappa_score as ck
D = os.path.expanduser("~/research/NE_ERAS_JEV/data/derived"); R = os.path.expanduser("~/research/Anes_JEV_vs_LoRA/analysis/reports"); PC = ["p1", "p2", "p3", "p4"]
K = lambda y, p: ck(y, p, weights="quadratic"); rd = lambda f: pd.read_csv(f"{D}/{f}.csv").set_index("op_id") if os.path.exists(f"{D}/{f}.csv") else None
pid = pd.read_csv(f"{D}/asa_dataset_v1.csv", usecols=["operation_id", "patient_sn"]).rename(columns={"operation_id": "op_id"}).set_index("op_id").patient_sn
def cuts_fit(E, y):
    cu = np.array([1.5, 2.5, 3.5]); g = np.linspace(1.0, 4.0, 121)
    for _ in range(3):
        for j in range(3): cu[j] = max(g, key=lambda v: K(y, 1 + (E[:, None] > np.where(np.arange(3) == j, v, cu)).sum(1)))
    return cu
G = ["G0", "GOFF", "GLOC", "GSTR"]; NM = {"G0": "无指导", "GOFF": "ASA 2020 官方定义", "GLOC": "本院记录习惯", "GSTR": "严格按指南"}
orig = rd("pred_guid_G0_t1_val").asa4.astype(int); od = np.bincount(orig.values - 1, minlength=4) / len(orig)
out = ["# 自然语言指导实验（读出臂，无训练、无标签）\n", f"本院 val n={len(orig)}；原记录分布 I/II/III/IV–V = {np.round(od*100,1).tolist()}%。指导措辞在看到结果前写定，未修改。\n",
       "| 输入 | 指导 | κ(对本院记录) | 预测分布 I/II/III/IV–V (%) | +100标签切点校准 κ | dev800 κ（方向核查） |", "|---|---|---|---|---|---|"]
res = {}
for tv in ("t1", "t2"):
    for g in G:
        v, dv = rd(f"pred_guid_{g}_{tv}_val"), rd(f"pred_guid_{g}_{tv}_dev800")
        if v is None: continue
        y = v.asa4.astype(int).values; pr = v[PC].values.argmax(1) + 1; res[(tv, g)] = (v.index, y, pr)
        dist = np.round(np.bincount(pr - 1, minlength=4) / len(pr) * 100, 1).tolist(); kc = "—"
        if dv is not None:
            E, Ev = (dv[PC].values * np.arange(1, 5)).sum(1), (v[PC].values * np.arange(1, 5)).sum(1); yd = dv.asa4.astype(int).values; rng = np.random.default_rng(3); ks = []
            for _ in range(60): s = rng.choice(len(yd), 100, replace=False); ks.append(K(y, 1 + (Ev[:, None] > cuts_fit(E[s], yd[s])).sum(1)))
            kc = f"{np.mean(ks):.3f}"
        kd = "—" if dv is None else f"{K(dv.asa4.astype(int).values, dv[PC].values.argmax(1) + 1):.3f}"
        out.append(f"| {tv.upper()} | {NM[g]} | {K(y,pr):.3f} | {dist} | {kc} | {kd} |")
out += ["", "## 与对照的配对差（κ，患者级 bootstrap；对本院记录）"]
def d(tv, a, b, note=""):
    if (tv, a) not in res or (tv, b) not in res: return
    ids, y, pa = res[(tv, a)]; pb = res[(tv, b)][2]; p = pid.loc[ids].values; up = np.unique(p); ix = {q: np.where(p == q)[0] for q in up}; gg = np.random.default_rng(7)
    bs = [np.concatenate([ix[q] for q in gg.choice(up, len(up))]) for _ in range(1000)]; v = np.array([K(y[s], pa[s]) - K(y[s], pb[s]) for s in bs])
    out.append(f"- {tv.upper()}：{NM[a]} − {NM[b]} = {K(y,pa)-K(y,pb):+.3f}（{np.percentile(v,2.5):+.3f} ~ {np.percentile(v,97.5):+.3f}） {note}")
for tv in ("t1", "t2"):
    for g in ("GOFF", "GLOC", "GSTR"): d(tv, g, "G0")
    d(tv, "GLOC", "GSTR", "（两个方向相反的指导之间）")
out += ["", "对照（本院 val，对原记录）：读出零样本 T1 0.443 / T2 0.505；读出 + 1000 标签校准 T1 0.534；本院 LoRA 全量 T1 0.561 / T2 0.606。",
        "解读规则（事先写定）：若 GLOC 把预测分布推向原记录分布（I 少、III 多）且 κ 上升，说明自然语言可替代部分标签；若分布位移但 κ 不升，说明可操控性成立但不等于更准；若分布也不动，则“可用自然语言指导”卖点未被支持。"]
open(f"{R}/guidance_report.md", "w").write("\n".join(out)); print("\n".join(out))
```
