## Supplementary material for "Non-generative readout with few-label calibration versus LoRA fine-tuning of a large language model for ASA physical status classification: accuracy, speed, cost and label efficiency in three data sources": Analysis code and aggregate results: 86_table1.py

```
"""Table 1：三个数据源的队列特征。输出 reports/table1.md。MOVER 与本院用 asa_common_*；VitalDB 读 cases.csv.gz（ASA 1–5，排除 ASA 6 与缺失）。"""
import os, io, gzip, numpy as np, pandas as pd
D = os.path.expanduser("~/research/NE_ERAS_JEV/data/derived"); R = os.path.expanduser("~/research/Anes_JEV_vs_LoRA/analysis/reports"); Rw = os.path.expanduser("~/research/NE_ERAS_JEV/data/raw")
def q(x): x = x.dropna(); return f"{x.median():.0f} ({x.quantile(.25):.0f}–{x.quantile(.75):.0f})"
def pct(b): return f"{100*np.mean(b):.1f}%"
def dist(a): v = np.bincount(a.astype(int) - 1, minlength=4)[:4] / len(a) * 100; return " / ".join(f"{x:.1f}" for x in v)
rows = []
m = pd.read_csv(f"{D}/asa_common_mover.csv"); pid = pd.read_csv(f"{D}/mover_pid_map.csv").set_index("op_id").pid
rows.append(("MOVER（UCI 手术室，美国）", len(m), pid.loc[m.op_id].nunique(), q(m.age), pct(m.female == 1), q(m.bmi), pct(m.inpatient == 1), "—", dist(m.asa4)))
l = pd.read_csv(f"{D}/asa_common_local.csv"); ld = pd.read_csv(f"{D}/asa_dataset_v1.csv", usecols=["operation_id", "patient_sn", "emergency"])
rows.append(("本院（腹部及盆腔手术，2021–26）", len(l), ld.patient_sn.nunique(), q(l.age), pct(l.female == 1), q(l.bmi), "100%", pct(ld.emergency == 1), dist(l.asa4)))
v = pd.read_csv(io.BytesIO(gzip.decompress(open(f"{Rw}/vitaldb/cases.csv.gz", "rb").read()))); v = v[v.asa.isin([1, 2, 3, 4, 5])]
rows.append(("VitalDB（首尔大学医院，非心脏手术）", len(v), v.subjectid.nunique(), q(v.age), pct(v.sex == "F"), q(v.bmi), "—", pct(v.emop == 1), dist(v.asa.clip(upper=4))))
out = ["# Table 1 队列特征（ASA 分布为 I / II / III / IV–V，%）\n", "| 数据源 | 手术台数 | 患者数 | 年龄 中位(IQR) | 女性 | BMI 中位(IQR) | 住院 | 急诊 | ASA I/II/III/IV–V (%) |", "|---|---|---|---|---|---|---|---|---|"]
out += [f"| {a} | {b:,} | {c:,} | {d} | {e} | {f} | {g} | {h} | {i} |" for a, b, c, d, e, f, g, h, i in rows]
open(f"{R}/table1.md", "w").write("\n".join(out)); print("\n".join(out))
```
