## Supplementary material for "Non-generative readout with few-label calibration versus LoRA fine-tuning of a large language model for ASA physical status classification: accuracy, speed, cost and label efficiency in three data sources": Analysis code and aggregate results: 87_paired_differences.py

```
"""主要配对差汇总（二次加权 κ；患者级 cluster bootstrap B=1000；H1–H3；LoRA 逐种子 + 种子均值）。输出 reports/paired_differences.csv/.md，供正文表 2 与图 3 使用。
自写读出与 OpenJev 的 3 切点校准用与既有脚本相同的坐标搜索；校准标签：本院 dev 随机 1000（种子 11）、MOVER train80 随机 1000（种子 11，pred_*_mover_cal）。"""
import os, re, numpy as np, pandas as pd
from sklearn.metrics import cohen_kappa_score as ck
D = os.path.expanduser("~/research/NE_ERAS_JEV/data/derived"); R = os.path.expanduser("~/research/Anes_JEV_vs_LoRA/analysis/reports"); PC = ["p1", "p2", "p3", "p4"]
K = lambda y, p: ck(y, p, weights="quadratic"); sm = lambda a: (lambda e: e / e.sum(1, keepdims=True))(np.exp(a - a.max(1, keepdims=True)))
def rd(f): p = f"{D}/{f}.csv"; return pd.read_csv(p).set_index("op_id") if os.path.exists(p) else None
def cuts_fit(E, y):
    cu = np.array([1.5, 2.5, 3.5]); g = np.linspace(1.0, 4.0, 121)
    for _ in range(3):
        for j in range(3): cu[j] = max(g, key=lambda v: K(y, 1 + (E[:, None] > np.where(np.arange(3) == j, v, cu)).sum(1)))
    return cu
def ojP(d): d = d[d.qtype == "score"] if "qtype" in d else d; cols = [x for x in d.columns if x.startswith("c") and x[1:].isdigit()]; return pd.DataFrame(sm(d[cols].values), index=d.index)
Ex = lambda P: (np.asarray(P) * np.arange(1, 5)).sum(1)
rows = []
def run(dsname, ids, y, pid, arms, contrasts, rngseed):
    """arms: name -> pred array aligned to ids；contrasts: (hyp, label, [armA...], [armB...])，多个 arm 取种子均值。"""
    up = np.unique(pid); idx = {p: np.where(pid == p)[0] for p in up}; rng = np.random.default_rng(rngseed)
    B = [np.concatenate([idx[p] for p in rng.choice(up, len(up))]) for _ in range(1000)]
    KB = {n: np.array([K(y[s], p[s]) for s in B]) for n, p in arms.items()}; K0 = {n: K(y, p) for n, p in arms.items()}
    for hyp, label, A, Bn in contrasts:
        if not all(a in arms for a in A + Bn): continue
        d = np.mean([KB[a] for a in A], 0) - np.mean([KB[b] for b in Bn], 0); pt = np.mean([K0[a] for a in A]) - np.mean([K0[b] for b in Bn])
        rows.append(dict(dataset=dsname, hyp=hyp, contrast=label, n=len(y), patients=len(up), diff=pt, lo=np.percentile(d, 2.5), hi=np.percentile(d, 97.5),
                         kA=np.mean([K0[a] for a in A]), kB=np.mean([K0[b] for b in Bn])))
# ---------------- 本院 ----------------
c = pd.read_csv(f"{D}/asa_common_local.csv", usecols=["op_id", "split", "asa4"]).merge(pd.read_csv(f"{D}/asa_dataset_v1.csv", usecols=["operation_id", "patient_sn"]).rename(columns={"operation_id": "op_id"}), on="op_id").set_index("op_id")
val = c[c.split == "val"].index; dev1k = c[c.split == "dev"].sample(1000, random_state=11).index
def local_arms(ids):
    A = {}
    for tv, f, fo in (("T1", "pred_llm_local", "pred_ojevF_local"), ("T2", "pred_llm_local_t2", "pred_ojevF_local_t2")):
        r = rd(f); A[f"读出 {tv} 未校准"] = r.loc[ids][PC].values.argmax(1) + 1
        cu = cuts_fit(Ex(r.loc[dev1k][PC].values), c.loc[dev1k].asa4.astype(int).values); A[f"读出 {tv} 校准"] = 1 + (Ex(r.loc[ids][PC].values)[:, None] > cu).sum(1)
        o = ojP(pd.read_csv(f"{D}/{fo}.csv").set_index("op_id")) if True else None
        cuo = cuts_fit(Ex(o.loc[dev1k].values), c.loc[dev1k].asa4.astype(int).values); A[f"OpenJev {tv} 校准"] = 1 + (Ex(o.loc[ids].values)[:, None] > cuo).sum(1); A[f"OpenJev {tv} 未校准"] = o.loc[ids].values.argmax(1) + 1
    for n in (100, 300, 1000, 4326):
        for sd in ("20261017", "11", "12"):
            d = rd(f"pred_lora_LOC{n}s{sd}_local_val")
            if d is not None: A[f"LoRA T1 n={n} s{sd}"] = d.loc[ids][PC].values.argmax(1) + 1
    for n in (1000, 4326): A[f"LoRA T2 n={n}"] = rd(f"pred_lora_LOCT2_{n}_local_val").loc[ids][PC].values.argmax(1) + 1
    return A
SD = ["20261017", "11", "12"]; L1 = lambda n: [f"LoRA T1 n={n} s{s}" for s in SD]
def h23(tv, arm, lab):
    out = []
    if tv == "T1":
        for s in SD: out.append(("H2", f"{lab} T1 校准(1000) − LoRA@1000 种子{s}", [f"{arm} T1 校准"], [f"LoRA T1 n=1000 s{s}"]))
        out.append(("H2", f"{lab} T1 校准(1000) − LoRA@1000 三种子均值", [f"{arm} T1 校准"], L1(1000)))
        for s in SD: out.append(("H3", f"LoRA@全量 种子{s} − {lab} T1 校准(1000)", [f"LoRA T1 n=4326 s{s}"], [f"{arm} T1 校准"]))
        out.append(("H3", f"LoRA@全量 三种子均值 − {lab} T1 校准(1000)", L1(4326), [f"{arm} T1 校准"]))
    else:
        out += [("H2", f"{lab} T2 校准(1000) − LoRA T2@1000（单种子）", [f"{arm} T2 校准"], ["LoRA T2 n=1000"]), ("H3", f"LoRA T2@全量（单种子）− {lab} T2 校准(1000)", ["LoRA T2 n=4326"], [f"{arm} T2 校准"])]
    return out
ids = list(val); A = local_arms(ids); y = c.loc[ids].asa4.astype(int).values; pid = c.loc[ids].patient_sn.values
ctr = h23("T1", "读出", "读出") + h23("T1", "OpenJev", "OpenJev") + h23("T2", "读出", "读出") + h23("T2", "OpenJev", "OpenJev")
ctr += [("输入", "读出 T2 校准 − 读出 T1 校准", ["读出 T2 校准"], ["读出 T1 校准"]), ("输入", "LoRA T2@全量 − LoRA T1@全量(种子20261017)", ["LoRA T2 n=4326"], ["LoRA T1 n=4326 s20261017"])]
run("本院 val", ids, y, pid, A, ctr, 20261019)
g = rd("pred_gen_local_val").dropna(subset=["pred"]); gi = sorted(set(g.index) & set(val)); Ag = local_arms(gi); Ag["生成式零样本"] = g.loc[gi].pred.astype(int).values
run("本院 val（生成式子集）", gi, c.loc[gi].asa4.astype(int).values, c.loc[gi].patient_sn.values, Ag,
    [("H1", "读出 T1 未校准 − 生成式零样本", ["读出 T1 未校准"], ["生成式零样本"]), ("H1", "OpenJev T1 校准 − 生成式零样本", ["OpenJev T1 校准"], ["生成式零样本"]), ("H1", "读出 T1 校准 − 生成式零样本", ["读出 T1 校准"], ["生成式零样本"])], 7)
# ---------------- MOVER ----------------
sub = pd.read_csv(f"{D}/mover_test_sub4000.csv").op_id.tolist(); pm = pd.read_csv(f"{D}/mover_pid_map.csv").set_index("op_id").pid
rt = rd("pred_llm_mover_test"); rc = rd("pred_llm_mover_cal"); ot = ojP(pd.read_csv(f"{D}/pred_ojevF_mover_sub.csv").set_index("op_id")); oc = ojP(pd.read_csv(f"{D}/pred_ojevF_mover_cal.csv").set_index("op_id"))
ycal = rc.asa4.astype(int).values; ocal = pd.read_csv(f"{D}/pred_ojevF_mover_cal.csv").set_index("op_id"); ocal = ocal[ocal.qtype == "score"] if "qtype" in ocal else ocal; yco = ocal.asa4.astype(int).values
def mover_arms(ids):
    A = {}; A["读出 T1 未校准"] = rt.loc[ids][PC].values.argmax(1) + 1
    cu = cuts_fit(Ex(rc[PC].values), ycal); A["读出 T1 校准"] = 1 + (Ex(rt.loc[ids][PC].values)[:, None] > cu).sum(1)
    cuo = cuts_fit(Ex(oc.values), yco); A["OpenJev T1 校准"] = 1 + (Ex(ot.loc[ids].values)[:, None] > cuo).sum(1)
    for n in (100, 300, 1000, 3000):
        for sfx, s in (("", "20261017"), ("s11", "11"), ("s12", "12")):
            d = rd(f"pred_lora_curve{n}{sfx}_mover_sub")
            if d is not None: A[f"LoRA T1 n={n} s{s}"] = d.loc[ids][PC].values.argmax(1) + 1
    for s, f in (("20261017", "pred_lora_mover12k_mover_test"), ("11", "pred_lora_curve12000s11_mover_sub"), ("12", "pred_lora_curve12000s12_mover_sub")):
        d = rd(f)
        if d is not None: A[f"LoRA T1 n=12000 s{s}"] = d.loc[ids][PC].values.argmax(1) + 1
    return A
ids = [i for i in sub if i in rt.index]; A = mover_arms(ids); y = rt.loc[ids].asa4.astype(int).values
L = lambda n: [f"LoRA T1 n={n} s{s}" for s in SD]
ctr = []
for arm in ("读出", "OpenJev"):
    ctr += [("H2", f"{arm} T1 校准(1000) − LoRA@1000 三种子均值", [f"{arm} T1 校准"], L(1000))] + [("H2", f"{arm} T1 校准(1000) − LoRA@1000 种子{s}", [f"{arm} T1 校准"], [f"LoRA T1 n=1000 s{s}"]) for s in SD]
    for n in (3000, 12000): ctr.append(("H3", f"LoRA@{n} 三种子均值 − {arm} T1 校准(1000)", L(n), [f"{arm} T1 校准"]))
run("MOVER 子集", ids, y, pm.loc[ids].values, A, ctr, 3)
gm = rd("pred_gen_mover_sub").dropna(subset=["pred"]); gi = [i for i in gm.index if i in rt.index]; Ag = mover_arms(gi); Ag["生成式零样本"] = gm.loc[gi].pred.astype(int).values
run("MOVER 子集（生成式子集）", gi, rt.loc[gi].asa4.astype(int).values, pm.loc[gi].values, Ag, [("H1", "读出 T1 未校准 − 生成式零样本", ["读出 T1 未校准"], ["生成式零样本"]), ("H1", "OpenJev T1 校准 − 生成式零样本", ["OpenJev T1 校准"], ["生成式零样本"])], 9)
# ---------------- VitalDB（目标站点，T2；文件齐备时） ----------------
try:
    exec(open(os.path.expanduser("~/research/Anes_JEV_vs_LoRA/analysis/scripts/87b_vdb_pairs.py")).read())
except FileNotFoundError: pass
df = pd.DataFrame(rows); df.to_csv(f"{R}/paired_differences.csv", index=False, encoding="utf-8-sig")
md = ["# 主要配对差（二次加权 κ；患者级 cluster bootstrap B=1000；A−B）\n", "| 数据 | 假设 | 对比 | n | A−B | 95%CI | κ(A) | κ(B) |", "|---|---|---|---|---|---|---|---|"]
for r in rows: md.append(f"| {r['dataset']} | {r['hyp']} | {r['contrast']} | {r['n']} | {r['diff']:+.3f} | {r['lo']:+.3f} ~ {r['hi']:+.3f} | {r['kA']:.3f} | {r['kB']:.3f} |")
open(f"{R}/paired_differences.md", "w").write("\n".join(md)); print("\n".join(md))
```
