## Supplementary material for "Non-generative readout with few-label calibration versus LoRA fine-tuning of a large language model for ASA physical status classification: accuracy, speed, cost and label efficiency in three data sources": Analysis code and aggregate results: 87b_vdb_pairs.py

```
# 由 87_paired_differences.py 通过 exec 载入（共享 rows、run、rd、cuts_fit、Ex、ojP 等）。VitalDB 目标站点：校准读出/OpenJev(1000 标签) 对 目标站点 LoRA。
import glob
Vd = pd.read_csv(f"{D}/asa_common_vitaldb_t2.csv", usecols=["op_id", "subjectid", "split", "asa4"]).set_index("op_id"); tid = list(Vd[Vd.split == "test"].index)
A = {}; 
for tv in ("t1", "t2"):
    for key, nm in (("zs", "读出"), ("oj", "OpenJev")):
        t, c = rd(f"pred_vdb_{key}_{tv}_test"), rd(f"pred_vdb_{key}_{tv}_cal")
        if t is None or c is None: continue
        if key == "zs": Et, Ec, raw = Ex(t.loc[tid][PC].values), Ex(c[PC].values), t.loc[tid][PC].values.argmax(1) + 1
        else: Et, Ec, raw = Ex(ojP(t).loc[tid].values), Ex(ojP(c).values), ojP(t).loc[tid].values.argmax(1) + 1
        A[f"{nm} {tv.upper()} 未校准"] = raw; A[f"{nm} {tv.upper()} 校准"] = 1 + (Et[:, None] > cuts_fit(Ec, c.asa4.astype(int).values)).sum(1)
for f in sorted(glob.glob(f"{D}/pred_vdb_lora_VDB_t*_test.csv")):
    mm = re.search(r"VDB_(t\d)_(\d+)_(\d+)_test", f); d = rd(os.path.basename(f)[:-4]); A[f"VLoRA {mm.group(1).upper()} n={mm.group(2)} s{mm.group(3)}"] = d.loc[tid][PC].values.argmax(1) + 1
SDV = ["20261017", "11", "12"]; ctr = []
for tv in ("T1", "T2"):
    for arm in ("读出", "OpenJev"):
        seeds = [f"VLoRA {tv} n=1000 s{s}" for s in SDV if f"VLoRA {tv} n=1000 s{s}" in A]
        if seeds: ctr.append(("H2", f"{arm} {tv} 校准(1000) − 目标站点 LoRA {tv}@1000（种子均值）" if arm == "读出" else f"{arm} {tv} 校准(1000) − 目标站点 LoRA {tv}@1000（种子均值）", [f"{arm} {tv} 校准"], seeds))
        for s in SDV:
            if f"VLoRA {tv} n=1000 s{s}" in A: ctr.append(("H2", f"{arm} {tv} 校准(1000) − 目标站点 LoRA {tv}@1000 种子{s}", [f"{arm} {tv} 校准"], [f"VLoRA {tv} n=1000 s{s}"]))
        s3 = [f"VLoRA {tv} n=3000 s{s}" for s in SDV if f"VLoRA {tv} n=3000 s{s}" in A]
        if s3: ctr.append(("H3", f"目标站点 LoRA {tv}@3000（种子均值）− {arm} {tv} 校准(1000)", s3, [f"{arm} {tv} 校准"]))
run("VitalDB", tid, Vd.loc[tid].asa4.astype(int).values, Vd.loc[tid].subjectid.values, A, ctr, 11)
```
