## Supplementary material for "Non-generative readout with few-label calibration versus LoRA fine-tuning of a large language model for ASA physical status classification: accuracy, speed, cost and label efficiency in three data sources": Analysis code and aggregate results: 93_figures.py

```
"""论文图（matplotlib，开源）：图1 研究设计；图2 速度-精度与标签效率；图3 配对差森林图（H1–H3）；图4 VitalDB 目标站点（文件齐备时）。中英文各一版，输出 figures/fig{N}_{zh,en}.{png,pdf,svg}。
数据全部来自 analysis/reports/（paired_differences.csv、calibration_curve.md 与 arms_eval_v3.md/local_eval.md 中的数字）。"""
import os, sys, numpy as np, pandas as pd
import matplotlib; matplotlib.use("Agg")
import matplotlib.pyplot as plt
from matplotlib import font_manager as fm
from matplotlib.patches import FancyBboxPatch
for f in ("/usr/share/fonts/opentype/noto/NotoSansCJK-Regular.ttc", "/usr/share/fonts/opentype/noto/NotoSansCJK-Bold.ttc"):
    if os.path.exists(f): fm.fontManager.addfont(f)
plt.rcParams.update({"font.sans-serif": ["Noto Sans CJK SC", "DejaVu Sans"], "axes.unicode_minus": False, "axes.spines.top": False, "axes.spines.right": False, "font.size": 9, "pdf.fonttype": 42, "svg.fonttype": "none"})
ROOT = os.path.expanduser("~/research/Anes_JEV_vs_LoRA"); R = f"{ROOT}/analysis/reports"; OUT = f"{ROOT}/figures"; os.makedirs(OUT, exist_ok=True)
PD = pd.read_csv(f"{R}/paired_differences.csv")
C = dict(gen="#b04a4a", read="#2c5aa0", oj="#d08a2c", lora="#3f6a1e", xgb="#7a7a7a")
T = {  # 双语文本
 "zh": dict(f2a="速度与精度（本院验证集）", f2x="单例推理延迟 p50（ms，批量=1，对数轴）", f2y="二次加权 κ", labs="本地标签数（对数轴；最左点为 k = 0，未校准）",
            gen="生成式零样本", rd0="读出零样本", rd100="读出+100 标签校准", rd1k="读出+1,000 标签校准", lo1k="LoRA n=1,000", lofull="LoRA 全量", rdk="读出 + k 标签校准（无训练）", lok="任务 LoRA（n 个标签训练）", xg="XGBoost",
            local="本院验证集", mover="MOVER 测试子集", ci="种子范围/校准 2.5–97.5 分位"),
 "en": dict(f2a="Speed versus accuracy (local validation set)", f2x="Per-case latency p50 (ms, batch = 1, log scale)", f2y="Quadratic weighted κ", labs="Local labels (log scale; leftmost point is k = 0, uncalibrated)",
            gen="Generative zero-shot", rd0="Readout, zero-shot", rd100="Readout + 100-label calibration", rd1k="Readout + 1,000-label calibration", lo1k="LoRA, n = 1,000", lofull="LoRA, full", rdk="Readout + k-label calibration (no training)", lok="Task LoRA (trained on n labels)", xg="XGBoost",
            local="Local validation set", mover="MOVER test subset", ci="Seed range / calibration 2.5–97.5th percentile")}
def save(fig, name, lang):
    for ext in ("png", "pdf", "svg"): fig.savefig(f"{OUT}/{name}_{lang}.{ext}", dpi=300, bbox_inches="tight")
    plt.close(fig)
# ---------------- 图 2 ----------------
def fig2(lang):
    t = T[lang]; fig, ax = plt.subplots(1, 3, figsize=(13.5, 4.2))
    pts = [(t["gen"], 7067, 0.227, C["gen"]), (t["rd0"], 111, 0.443, C["read"]), (t["rd100"], 111, 0.526, "#3f77c0"), (t["rd1k"], 111, 0.534, "#1d3f73"), (t["lo1k"], 179, 0.559, C["lora"]), (t["lofull"], 179, 0.561, "#2a4a14")]
    for n, x, y, c in pts: ax[0].scatter(x, y, s=60, color=c, zorder=3, label=f"{n}  κ = {y:.3f}")
    ax[0].set_xscale("log"); ax[0].set_xlabel(t["f2x"]); ax[0].set_ylabel(t["f2y"]); ax[0].set_title(t["f2a"], fontsize=10); ax[0].legend(fontsize=7, loc="center right", frameon=False); ax[0].set_ylim(0.15, 0.62); ax[0].grid(alpha=.25)
    def curve(a, title, ks, km, klo, khi, ln, lm, llo, lhi, xn, xm):
        a.plot([1] + ks, km, "o-", color=C["read"], label=t["rdk"], ms=4); a.fill_between([1] + ks, klo, khi, color=C["read"], alpha=.15, lw=0)
        a.errorbar(ln, lm, yerr=[np.array(lm) - np.array(llo), np.array(lhi) - np.array(lm)], fmt="s-", color=C["lora"], capsize=2.5, ms=4, label=t["lok"])
        a.plot(xn, xm, "^--", color=C["xgb"], ms=4, label=t["xg"])
        a.set_xscale("log"); a.set_xlabel(t["labs"]); a.set_ylabel(t["f2y"]); a.set_title(title, fontsize=10); a.grid(alpha=.25); a.set_ylim(0.2, 0.62)
    curve(ax[1], t["local"], [25, 50, 100, 200, 500, 1000], [0.443, 0.491, 0.516, 0.526, 0.528, 0.535, 0.540], [0.443, 0.384, 0.471, 0.474, 0.501, 0.520, 0.540], [0.443, 0.546, 0.547, 0.547, 0.544, 0.540, 0.540],
          [100, 300, 1000, 4326], [0.423, 0.542, 0.559, 0.561], [0.338, 0.530, 0.553, 0.556], [0.469, 0.556, 0.563, 0.570], [100, 300, 1000, 4326], [0.431, 0.467, 0.521, 0.549])
    curve(ax[2], t["mover"], [25, 50, 100, 200, 500, 1000], [0.473, 0.467, 0.495, 0.508, 0.517, 0.521, 0.524], [0.473, 0.372, 0.422, 0.476, 0.495, 0.516, 0.524], [0.473, 0.519, 0.530, 0.528, 0.529, 0.529, 0.524],
          [100, 300, 1000, 3000, 12000], [0.410, 0.411, 0.505, 0.563, 0.583], [0.394, 0.256, 0.462, 0.551, 0.580], [0.438, 0.500, 0.527, 0.575, 0.587], [100, 300, 1000, 3000, 12000], [0.234, 0.305, 0.395, 0.469, 0.504])
    h, l = ax[2].get_legend_handles_labels(); fig.legend(h, l, loc="lower center", ncol=3, fontsize=8, frameon=False, bbox_to_anchor=(0.66, -0.06)); ax[1].text(0.02, 0.02, t["ci"], transform=ax[1].transAxes, fontsize=6.5, color="#555")
    for a, l in zip(ax, "ABC"): a.text(-0.12, 1.06, l, transform=a.transAxes, fontsize=12, fontweight="bold")
    plt.tight_layout(); save(fig, "fig2_cost_efficiency", lang)
# ---------------- 图 1 ----------------
def fig1(lang):
    zh = lang == "zh"; fig, ax = plt.subplots(figsize=(12.5, 6.2)); ax.axis("off"); ax.set_xlim(0, 101); ax.set_ylim(0, 60)
    def box(x, y, w, h, txt, fc="#eef3fa", ec="#2c5aa0", fs=8.2, bold=False):
        ax.add_patch(FancyBboxPatch((x, y), w, h, boxstyle="round,pad=0.25,rounding_size=1.2", fc=fc, ec=ec, lw=1.1)); ax.text(x + w / 2, y + h / 2, txt, ha="center", va="center", fontsize=fs, fontweight="bold" if bold else "normal", linespacing=1.35)
    arrow = lambda x1, y1, x2, y2: ax.annotate("", xy=(x2, y2), xytext=(x1, y1), arrowprops=dict(arrowstyle="-|>", color="#555", lw=1.1))
    hd = dict(zh=["数据源", "输入", "比较臂（同一底模 Qwen3.5-9B）", "评价"], en=["Data sources", "Inputs", "Arms (same base model, Qwen3.5-9B)", "Evaluation"])[lang]
    for x, h in zip((1, 27, 49, 82), hd): ax.text(x, 57.5, h, fontsize=10, fontweight="bold")
    if zh:
        box(1, 41, 23, 13, "MOVER（美国）\n57,473 台 / 38,245 人\n开发：80%；测试子集 4,000 台", fc="#f4f4f4", ec="#777")
        box(1, 24, 23, 13, "本院（腹部及盆腔手术）\n7,079 台 / 6,804 人\n开发 2021–24：4,326\n时间外验证 2025–26：2,753", fc="#f4f4f4", ec="#777")
        box(1, 7, 23, 13, "VitalDB（首尔）\n6,242 台 / 5,961 人\n训练 70%；测试 1,885 台", fc="#f4f4f4", ec="#777")
        box(27, 41, 18, 13, "T1：共有结构化变量\n（年龄、性别、BMI、病史、\n化验、手术名）", fc="#fff6e6", ec="#d08a2c"); box(27, 24, 18, 13, "T2：T1 + 术前诊断文本\n（本院访视文本；\nVitalDB 诊断与检查）", fc="#fff6e6", ec="#d08a2c")
        arms = ["生成式零样本（理由 + 答案）", "非生成式读出（选项 logits）", "OpenJev（冻结配置）", "读出 / OpenJev + k 标签切点校准", "任务 LoRA（n = 100…全量，3 种子）"]
        ev = "二次加权 κ\n（患者级 cluster\nbootstrap）\n\nH1 读出 > 生成式\nH2 校准读出非劣效于\nLoRA@1,000（δ = 0.05）\nH3 LoRA 全量 >\n校准读出\n\n速度 · 训练成本\n标签数 · ECE\n矛盾内容粗查\n规模敏感性"
    else:
        box(1, 41, 23, 13, "MOVER (USA)\n57,473 ops / 38,245 patients\nDevelopment 80%; test subset 4,000", fc="#f4f4f4", ec="#777")
        box(1, 24, 23, 13, "Local hospital (abdominal/pelvic)\n7,079 ops / 6,804 patients\nDevelopment 2021–24: 4,326\nTemporal validation 2025–26: 2,753", fc="#f4f4f4", ec="#777")
        box(1, 7, 23, 13, "VitalDB (Seoul)\n6,242 ops / 5,961 patients\nTrain 70%; test 1,885", fc="#f4f4f4", ec="#777")
        box(27, 41, 18, 13, "T1: shared structured\nvariables (age, sex, BMI,\nhistory, labs, procedure)", fc="#fff6e6", ec="#d08a2c"); box(27, 24, 18, 13, "T2: T1 + preoperative\ndiagnosis text (local visit\nnote; VitalDB diagnoses/tests)", fc="#fff6e6", ec="#d08a2c")
        arms = ["Generative zero-shot (rationale + answer)", "Non-generative readout (option logits)", "OpenJev (frozen configuration)", "Readout / OpenJev + k-label cutpoint calibration", "Task LoRA (n = 100…full, 3 seeds)"]
        ev = "Quadratic weighted κ\n(patient-level cluster\nbootstrap)\n\nH1 readout > generative\nH2 calibrated readout\nnon-inferior to LoRA@1,000\n(δ = 0.05)\nH3 full LoRA >\ncalibrated readout\n\nSpeed · training cost\nlabels needed · ECE\ncontradiction audit\nscale sensitivity"
    cols = ["#b04a4a", "#2c5aa0", "#d08a2c", "#1d3f73", "#3f6a1e"]
    for i, (a, c) in enumerate(zip(arms, cols)): box(49, 47 - i * 9.6, 30, 7.6, a, fc="white", ec=c, fs=8)
    box(82, 8, 17.5, 43, ev, fc="#eef7ee", ec="#3f6a1e", fs=6.6)
    arrow(24.5, 47.5, 26.8, 47.5); arrow(24.5, 30.5, 26.8, 30.5); arrow(24.5, 13.5, 26.8, 26.5)
    arrow(45.2, 47.5, 48.8, 47.5); arrow(45.2, 30.5, 48.8, 30.5); arrow(79.3, 32, 81.7, 32)
    plt.savefig  # noqa
    save(fig, "fig1_design", lang)
# ---------------- 图 3 森林图 ----------------
def fig3(lang):
    zh = lang == "zh"; P = PD.copy()
    def g(ds, hyp, contrast): 
        r = P[(P.dataset == ds) & (P.hyp == hyp) & (P.contrast == contrast)]
        return None if r.empty else r.iloc[0]
    L = "本院 val" if True else ""
    rows = [  # (分组, 标签zh, 标签en, ds, hyp, contrast, 颜色)
      ("H1", "读出 − 生成式（本院）", "Readout − generative (local)", "本院 val（生成式子集）", "H1", "读出 T1 未校准 − 生成式零样本", C["read"]),
      ("H1", "OpenJev 校准 − 生成式（本院）", "OpenJev cal. − generative (local)", "本院 val（生成式子集）", "H1", "OpenJev T1 校准 − 生成式零样本", C["oj"]),
      ("H1", "读出 − 生成式（MOVER）", "Readout − generative (MOVER)", "MOVER 子集（生成式子集）", "H1", "读出 T1 未校准 − 生成式零样本", C["read"]),
      ("H1", "OpenJev 校准 − 生成式（MOVER）", "OpenJev cal. − generative (MOVER)", "MOVER 子集（生成式子集）", "H1", "OpenJev T1 校准 − 生成式零样本", C["oj"]),
      ("H2", "读出 T1，本院（3 种子均值）", "Readout T1, local (3-seed mean)", "本院 val", "H2", "读出 T1 校准(1000) − LoRA@1000 三种子均值", C["read"]),
      ("H2", "读出 T1，本院（预先指定种子）", "Readout T1, local (prespecified seed)", "本院 val", "H2", "读出 T1 校准(1000) − LoRA@1000 种子20261017", C["read"]),
      ("H2", "读出 T1，本院（种子 11，标签匹配）", "Readout T1, local (seed 11, label-matched)", "本院 val", "H2", "读出 T1 校准(1000) − LoRA@1000 种子11", C["read"]),
      ("H2", "OpenJev T1，本院（3 种子均值）", "OpenJev T1, local (3-seed mean)", "本院 val", "H2", "OpenJev T1 校准(1000) − LoRA@1000 三种子均值", C["oj"]),
      ("H2", "读出 T2，本院（单种子）", "Readout T2, local (single seed)", "本院 val", "H2", "读出 T2 校准(1000) − LoRA T2@1000（单种子）", C["read"]),
      ("H2", "OpenJev T2，本院（单种子）", "OpenJev T2, local (single seed)", "本院 val", "H2", "OpenJev T2 校准(1000) − LoRA T2@1000（单种子）", C["oj"]),
      ("H2", "读出 T1，MOVER（3 种子均值）", "Readout T1, MOVER (3-seed mean)", "MOVER 子集", "H2", "读出 T1 校准(1000) − LoRA@1000 三种子均值", C["read"]),
      ("H2", "OpenJev T1，MOVER（3 种子均值）", "OpenJev T1, MOVER (3-seed mean)", "MOVER 子集", "H2", "OpenJev T1 校准(1000) − LoRA@1000 三种子均值", C["oj"]),
      ("H2", "读出 T2，VitalDB", "Readout T2, VitalDB", "VitalDB", "H2", "读出 T2 校准(1000) − 目标站点 LoRA T2@1000（种子均值）", C["read"]),
      ("H3", "LoRA 全量 − 读出 T1，本院（3 种子均值）", "Full LoRA − readout T1, local (3-seed mean)", "本院 val", "H3", "LoRA@全量 三种子均值 − 读出 T1 校准(1000)", C["read"]),
      ("H3", "LoRA 全量 − OpenJev T1，本院", "Full LoRA − OpenJev T1, local", "本院 val", "H3", "LoRA@全量 三种子均值 − OpenJev T1 校准(1000)", C["oj"]),
      ("H3", "LoRA T2 全量 − 读出 T2，本院", "Full LoRA − readout T2, local", "本院 val", "H3", "LoRA T2@全量（单种子）− 读出 T2 校准(1000)", C["read"]),
      ("H3", "LoRA T2 全量 − OpenJev T2，本院", "Full LoRA − OpenJev T2, local", "本院 val", "H3", "LoRA T2@全量（单种子）− OpenJev T2 校准(1000)", C["oj"]),
      ("H3", "LoRA@3,000 − 读出 T1，MOVER", "LoRA@3,000 − readout T1, MOVER", "MOVER 子集", "H3", "LoRA@3000 三种子均值 − 读出 T1 校准(1000)", C["read"]),
      ("H3", "LoRA@12,000 − 读出 T1，MOVER", "LoRA@12,000 − readout T1, MOVER", "MOVER 子集", "H3", "LoRA@12000 三种子均值 − 读出 T1 校准(1000)", C["read"]),
      ("H3", "LoRA@3,000 − 目标站点读出 T2，VitalDB", "LoRA@3,000 − readout T2, VitalDB", "VitalDB", "H3", "目标站点 LoRA T2@3000（种子均值）− 读出 T2 校准(1000)", C["read"])]
    rows = [(a, bz, be, g(d, h, c), col) for a, bz, be, d, h, c, col in rows]; rows = [r for r in rows if r[3] is not None]
    import json as _j; _N = _j.load(open(f"{R}/review_numbers_105.json"))
    class _R:  # 含标签抽样与种子变异的 H2（50 个标签池 × 3 个 LoRA 种子）
        def __getitem__(self, k): return getattr(self, k)
    _r = _R(); _r.__dict__.update({"diff": _N["comb_diff"], "lo": _N["comb_lo"], "hi": _N["comb_hi"]})
    _k = [i for i, x in enumerate(rows) if x[0] == "H2" and "标签匹配" in x[1]]
    rows.insert((_k[0] + 1) if _k else len(rows), ("H2", "读出 T1，本院（50 个标签池，含标签抽样）", "Readout T1, local (50 label pools, label-draw incl.)", _r, C["read"]))
    fig, axs = plt.subplots(3, 1, figsize=(8.2, 0.34 * len(rows) + 2.6), gridspec_kw=dict(height_ratios=[sum(r[0] == k for r in rows) for k in ("H1", "H2", "H3")], hspace=0.55), sharex=False)
    ttl = dict(H1=("H1  读出 − 生成式零样本（优效）", "H1  Readout − generative zero-shot (superiority)"), H2=("H2  校准读出(1,000) − LoRA@1,000（非劣效，界值 −0.05）", "H2  Calibrated readout (1,000) − LoRA@1,000 (non-inferiority, margin −0.05)"), H3=("H3  LoRA 全量 − 校准读出(1,000)（优效）", "H3  Full LoRA − calibrated readout (1,000) (superiority)"))
    for ax, k in zip(axs, ("H1", "H2", "H3")):
        rr = [r for r in rows if r[0] == k]
        for i, (a, bz, be, r, col) in enumerate(rr):
            y = len(rr) - 1 - i; ax.plot([r.lo, r.hi], [y, y], color=col, lw=1.8); ax.scatter(r["diff"], y, color=col, s=28, zorder=3)
            ax.text(1.02, y, f"{r['diff']:+.3f} ({r.lo:+.3f}, {r.hi:+.3f})", transform=ax.get_yaxis_transform(), va="center", fontsize=7.5)
        ax.set_yticks(range(len(rr))); ax.set_yticklabels([(bz if zh else be) for a, bz, be, r, col in rr][::-1], fontsize=7.8); ax.axvline(0, color="#333", lw=0.8)
        if k == "H2": ax.axvline(-0.05, color="#b04a4a", ls="--", lw=1)
        ax.set_title(ttl[k][0 if zh else 1], fontsize=9, loc="left", fontweight="bold"); ax.set_ylim(-0.7, len(rr) - 0.3); ax.grid(axis="x", alpha=.25)
        ax.set_xlabel("κ 差值（95% CI）" if zh else "κ difference (95% CI)", fontsize=8)
    fig.suptitle("")
    save(fig, "fig3_forest", lang)
for lang in ("zh", "en"):
    fig1(lang); fig2(lang); fig3(lang)
print("done")
```
