## Supplementary material for "Non-generative readout with few-label calibration versus LoRA fine-tuning of a large language model for ASA physical status classification: accuracy, speed, cost and label efficiency in three data sources": Analysis code and aggregate results: 94_split_table.py

```
"""开发/评价划分之间的特征比较（TRIPOD-LLM 16c）。输出 reports/table_splits.md。"""
import os, io, gzip, numpy as np, pandas as pd
D = os.path.expanduser("~/research/NE_ERAS_JEV/data/derived"); R = os.path.expanduser("~/research/Anes_JEV_vs_LoRA/analysis/reports"); Rw = os.path.expanduser("~/research/NE_ERAS_JEV/data/raw")
q = lambda x: (lambda x: f"{x.median():.0f} ({x.quantile(.25):.0f}–{x.quantile(.75):.0f})")(x.dropna()); dist = lambda a: " / ".join(f"{x:.1f}" for x in np.bincount(a.astype(int) - 1, minlength=4)[:4] / len(a) * 100)
rows = []
def add(ds, sp, d, lab): rows.append((ds, sp, len(d), q(d.age), f"{100*(d.female==1).mean():.1f}%", q(d.bmi), f"{100*d.bmi.isna().mean():.1f}%", f"{100*d[lab].notna().mean():.0f}%", dist(d.asa4)))
m = pd.read_csv(f"{D}/asa_common_mover.csv"); sub = set(pd.read_csv(f"{D}/mover_test_sub4000.csv").op_id)
add("MOVER", "开发划分 (train80)", m[m.split == "train80"], "cr_mgdl"); add("MOVER", "测试集 (test20)", m[m.split == "test20"], "cr_mgdl"); add("MOVER", "评价子集 (4,000)", m[m.op_id.isin(sub)], "cr_mgdl")
l = pd.read_csv(f"{D}/asa_common_local.csv"); add("本院", "开发集 2021–24", l[l.split == "dev"], "cr_mgdl"); add("本院", "验证集 2025–26", l[l.split == "val"], "cr_mgdl")
v = pd.read_csv(io.BytesIO(gzip.decompress(open(f"{Rw}/vitaldb/cases.csv.gz", "rb").read()))); v["op_id"] = "vdb" + v.caseid.astype(str); t = pd.read_csv(f"{D}/asa_common_vitaldb_t2.csv", usecols=["op_id", "split", "asa4"])
v = v.merge(t, on="op_id"); v["female"] = (v.sex == "F").astype(int); v["cr_mgdl"] = v.preop_cr
add("VitalDB", "训练集 70%", v[v.split == "train"], "cr_mgdl"); add("VitalDB", "测试集 30%", v[v.split == "test"], "cr_mgdl")
out = ["# 开发与评价划分的特征比较（ASA 为 I / II / III / IV–V，%）\n", "| 数据源 | 划分 | 手术台数 | 年龄 中位(IQR) | 女性 | BMI 中位(IQR) | BMI 缺失 | 术前肌酐可用 | ASA 分布 |", "|---|---|---|---|---|---|---|---|---|"]
out += ["| " + " | ".join(map(str, r)) + " |" for r in rows]; open(f"{R}/table_splits.md", "w").write("\n".join(out)); print("\n".join(out))
```
