## Supplementary material for "Non-generative readout with few-label calibration versus LoRA fine-tuning of a large language model for ASA physical status classification: accuracy, speed, cost and label efficiency in three data sources": Analysis code and aggregate results: 95_fig4_vdb.py

```
"""图 4：VitalDB 目标站点标签效率（T2，测试集 1,885 台）：读出 + k 标签校准（k 取自训练划分的 1,000 标签池，每个 k 重复 100 次）、OpenJev 同法、目标站点 LoRA（种子均值与范围）。输出 figures/fig4_vitaldb_{zh,en}.* 与 reports/vitaldb_curve.md"""
import os, re, glob, numpy as np, pandas as pd
import matplotlib; matplotlib.use("Agg")
import matplotlib.pyplot as plt
from matplotlib import font_manager as fm
from sklearn.metrics import cohen_kappa_score as ck
for f in ("/usr/share/fonts/opentype/noto/NotoSansCJK-Regular.ttc",):
    if os.path.exists(f): fm.fontManager.addfont(f)
plt.rcParams.update({"font.sans-serif": ["Noto Sans CJK SC", "DejaVu Sans"], "axes.unicode_minus": False, "axes.spines.top": False, "axes.spines.right": False, "font.size": 9, "pdf.fonttype": 42, "svg.fonttype": "none"})
D = os.path.expanduser("~/research/NE_ERAS_JEV/data/derived"); ROOT = os.path.expanduser("~/research/Anes_JEV_vs_LoRA"); PC = ["p1", "p2", "p3", "p4"]
K = lambda y, p: ck(y, p, weights="quadratic"); sm = lambda a: (lambda e: e / e.sum(1, keepdims=True))(np.exp(a - a.max(1, keepdims=True)))
rd = lambda f: pd.read_csv(f"{D}/{f}.csv").set_index("op_id")
V = pd.read_csv(f"{D}/asa_common_vitaldb_t2.csv", usecols=["op_id", "split"]).set_index("op_id"); tid = V[V.split == "test"].index
def cuts_fit(E, y):
    cu = np.array([1.5, 2.5, 3.5]); g = np.linspace(1.0, 4.0, 121)
    for _ in range(3):
        for j in range(3): cu[j] = max(g, key=lambda v: K(y, 1 + (E[:, None] > np.where(np.arange(3) == j, v, cu)).sum(1)))
    return cu
def E_of(d, key): return (d[PC].values * np.arange(1, 5)).sum(1) if key == "zs" else (sm(d[[x for x in d.columns if x.startswith("c")]].values) * np.arange(1, 5)).sum(1)
res = {}; rng = np.random.default_rng(7); ks_ = [25, 50, 100, 200, 500, 1000]
for key, nm in (("zs", "readout"), ("oj", "openjev")):
    t, c = rd(f"pred_vdb_{key}_t2_test").loc[tid], rd(f"pred_vdb_{key}_t2_cal"); y = t.asa4.astype(int).values; Et, Ec, yc = E_of(t, key), E_of(c, key), c.asa4.astype(int).values
    raw = (t[PC].values.argmax(1) if key == "zs" else sm(t[[x for x in t.columns if x.startswith("c")]].values).argmax(1)) + 1; rows = [(0, K(y, raw), K(y, raw), K(y, raw))]
    for k in ks_:
        v = []
        for _ in range(100 if k < 1000 else 1):
            i = rng.choice(len(yc), k, replace=False); v.append(K(y, 1 + (Et[:, None] > cuts_fit(Ec[i], yc[i])).sum(1)))
        rows.append((k, np.mean(v), np.percentile(v, 2.5), np.percentile(v, 97.5)))
    res[nm] = rows
lo = {}
for f in glob.glob(f"{D}/pred_vdb_lora_VDB_t2_*_test.csv"):
    m = re.search(r"VDB_t2_(\d+)_(\d+)_test", f); d = rd(os.path.basename(f)[:-4]).loc[tid]; lo.setdefault(int(m.group(1)), []).append(K(d.asa4.astype(int).values, d[PC].values.argmax(1) + 1))
lon = sorted(lo); lm = [np.mean(lo[n]) for n in lon]; ll = [min(lo[n]) for n in lon]; lh = [max(lo[n]) for n in lon]
md = ["# VitalDB 目标站点标签效率曲线（T2，测试集 n=%d）\n" % len(tid), "| k | 读出 κ 均值 [2.5–97.5%] | OpenJev κ 均值 [2.5–97.5%] |", "|---|---|---|"]
for a, b in zip(res["readout"], res["openjev"]): md.append(f"| {a[0]} | {a[1]:.3f} [{a[2]:.3f}–{a[3]:.3f}] | {b[1]:.3f} [{b[2]:.3f}–{b[3]:.3f}] |")
md += ["", "| LoRA n | 种子数 | κ 均值 [范围] |", "|---|---|---|"] + [f"| {n} | {len(lo[n])} | {np.mean(lo[n]):.3f} [{min(lo[n]):.3f}–{max(lo[n]):.3f}] |" for n in lon]
open(f"{ROOT}/analysis/reports/vitaldb_curve.md", "w").write("\n".join(md)); print("\n".join(md))
T = {"zh": ("VitalDB 测试集（T2）", "目标站点标签数（对数轴；最左点为 k = 0，未校准）", "二次加权 κ", "读出 + k 标签校准", "OpenJev + k 标签校准", "目标站点 LoRA（种子均值，误差线为范围）"),
     "en": ("VitalDB test set (T2)", "Target-site labels (log scale; leftmost point is k = 0, uncalibrated)", "Quadratic weighted κ", "Readout + k-label calibration", "OpenJev + k-label calibration", "Target-site LoRA (seed mean; bars = range)")}
for lang, t in T.items():
    fig, ax = plt.subplots(figsize=(5.4, 4.0))
    for nm, col, lab in (("readout", "#2c5aa0", t[3]), ("openjev", "#d08a2c", t[4])):
        r = res[nm]; x = [1 if a[0] == 0 else a[0] for a in r]; ax.plot(x, [a[1] for a in r], "o-", color=col, ms=4, label=lab); ax.fill_between(x, [a[2] for a in r], [a[3] for a in r], color=col, alpha=.13, lw=0)
    ax.errorbar(lon, lm, yerr=[np.array(lm) - np.array(ll), np.array(lh) - np.array(lm)], fmt="s-", color="#3f6a1e", capsize=2.5, ms=4, label=t[5])
    ax.set_xscale("log"); ax.set_xlabel(t[1]); ax.set_ylabel(t[2]); ax.set_title(t[0], fontsize=10); ax.grid(alpha=.25); ax.legend(fontsize=7.5, frameon=False, loc="lower left")
    plt.tight_layout()
    for ext in ("png", "pdf", "svg"): fig.savefig(f"{ROOT}/figures/fig4_vitaldb_{lang}.{ext}", dpi=300, bbox_inches="tight")
    plt.close(fig)
```
