## Supplementary material for "Non-generative readout with few-label calibration versus LoRA fine-tuning of a large language model for ASA physical status classification: accuracy, speed, cost and label efficiency in three data sources": Analysis code and aggregate results: 96_fig5_guidance.py

```
"""图 5 与补充配对差：自然语言指导（读出臂，无训练、无标签）。A：各指导下预测类别构成与原记录构成；B：κ（对本院记录），参考线为 1,000 标签校准读出与本院 LoRA 全量。
另算 T2 下 GLOC − 1000 标签校准读出、GLOC − LoRA T2@1000 的患者级配对差。输出 figures/fig5_guidance_{zh,en}.*、reports/guidance_pairs.md"""
import os, numpy as np, pandas as pd
import matplotlib; matplotlib.use("Agg")
import matplotlib.pyplot as plt
from matplotlib import font_manager as fm
from sklearn.metrics import cohen_kappa_score as ck
fm.fontManager.addfont("/usr/share/fonts/opentype/noto/NotoSansCJK-Regular.ttc")
plt.rcParams.update({"font.sans-serif": ["Noto Sans CJK SC", "DejaVu Sans"], "axes.unicode_minus": False, "axes.spines.top": False, "axes.spines.right": False, "font.size": 9, "pdf.fonttype": 42, "svg.fonttype": "none"})
D = os.path.expanduser("~/research/NE_ERAS_JEV/data/derived"); ROOT = os.path.expanduser("~/research/Anes_JEV_vs_LoRA"); PC = ["p1", "p2", "p3", "p4"]
K = lambda y, p: ck(y, p, weights="quadratic"); rd = lambda f: pd.read_csv(f"{D}/{f}.csv").set_index("op_id")
pid = pd.read_csv(f"{D}/asa_dataset_v1.csv", usecols=["operation_id", "patient_sn"]).rename(columns={"operation_id": "op_id"}).set_index("op_id").patient_sn
G = ["G0", "GOFF", "GLOC", "GSTR"]; P = {}
for tv in ("t1", "t2"):
    for g in G: P[(tv, g)] = rd(f"pred_guid_{g}_{tv}_val")
ids = P[("t1", "G0")].index; y = P[("t1", "G0")].asa4.astype(int).values; pr = {k: v.loc[ids][PC].values.argmax(1) + 1 for k, v in P.items()}
def cuts_fit(E, yy):
    cu = np.array([1.5, 2.5, 3.5]); g = np.linspace(1.0, 4.0, 121)
    for _ in range(3):
        for j in range(3): cu[j] = max(g, key=lambda v: K(yy, 1 + (E[:, None] > np.where(np.arange(3) == j, v, cu)).sum(1)))
    return cu
c = pd.read_csv(f"{D}/asa_common_local.csv", usecols=["op_id", "split", "asa4"]).set_index("op_id"); dev1k = c[c.split == "dev"].sample(1000, random_state=11).index
Ex = lambda a: (np.asarray(a) * np.arange(1, 5)).sum(1)
cal = {}
for tv, f in (("t1", "pred_llm_local"), ("t2", "pred_llm_local_t2")):
    r = rd(f); cal[tv] = 1 + (Ex(r.loc[ids][PC].values)[:, None] > cuts_fit(Ex(r.loc[dev1k][PC].values), c.loc[dev1k].asa4.astype(int).values)).sum(1)
lora1k_t2 = rd("pred_lora_LOCT2_1000_local_val").loc[ids][PC].values.argmax(1) + 1; lora_full = {"t1": rd("pred_lora_LOC4326s20261017_local_val").loc[ids][PC].values.argmax(1) + 1, "t2": rd("pred_lora_LOCT2_4326_local_val").loc[ids][PC].values.argmax(1) + 1}
pp = pid.loc[ids].values; up = np.unique(pp); ix = {q: np.where(pp == q)[0] for q in up}; rng = np.random.default_rng(13); B = [np.concatenate([ix[q] for q in rng.choice(up, len(up))]) for _ in range(1000)]
def d(a, b): v = np.array([K(y[s], a[s]) - K(y[s], b[s]) for s in B]); return f"{K(y,a)-K(y,b):+.3f}（{np.percentile(v,2.5):+.3f} ~ {np.percentile(v,97.5):+.3f}）"
out = ["# 自然语言指导：补充配对差（本院 val，n=%d）\n" % len(ids)]
for tv in ("t1", "t2"):
    out.append(f"- {tv.upper()}：GLOC（本院记录习惯，无标签）− 无指导读出 + 1000 标签校准 = {d(pr[(tv,'GLOC')], cal[tv])}")
    out.append(f"- {tv.upper()}：GLOC − 本院 LoRA 全量 = {d(pr[(tv,'GLOC')], lora_full[tv])}")
out.append(f"- T2：GLOC − 本院 LoRA T2@1000（单种子）= {d(pr[('t2','GLOC')], lora1k_t2)}")
out.append(f"- T2：GSTR − 无指导读出 + 1000 标签校准 = {d(pr[('t2','GSTR')], cal['t2'])}")
open(f"{ROOT}/analysis/reports/guidance_pairs.md", "w").write("\n".join(out)); print("\n".join(out))
orig = np.bincount(y - 1, minlength=4) / len(y) * 100
NM = {"zh": {"G0": "无指导", "GOFF": "ASA 官方定义", "GLOC": "本院记录习惯", "GSTR": "严格按指南", "orig": "原记录"}, "en": {"G0": "No guidance", "GOFF": "ASA 2020 definitions", "GLOC": "Local habits", "GSTR": "Strict guideline", "orig": "Recorded ASA"}}
CL = ["#9ecae1", "#4292c6", "#2171b5", "#08306b"]
for lang in ("zh", "en"):
    nm = NM[lang]; fig, ax = plt.subplots(1, 3, figsize=(13, 4.0), gridspec_kw=dict(width_ratios=[1.2, 1.2, 1]))
    for a, tv in zip(ax[:2], ("t1", "t2")):
        labs = [nm["orig"]] + [nm[g] for g in G]; dist = [orig] + [np.bincount(pr[(tv, g)] - 1, minlength=4) / len(y) * 100 for g in G]; bot = np.zeros(len(labs))
        for k in range(4): v = np.array([x[k] for x in dist]); a.barh(range(len(labs)), v, left=bot, color=CL[k], label=["I", "II", "III", "IV–V"][k]); bot += v
        a.set_yticks(range(len(labs))); a.set_yticklabels(labs); a.invert_yaxis(); a.set_xlim(0, 100); a.set_xlabel("%" ); a.set_title(("T1" if tv == "t1" else "T2") + (" · 预测类别构成" if lang == "zh" else " · Predicted class mix"), fontsize=10)
        for i, g in enumerate(G): a.text(101, i + 1, f"κ {K(y, pr[(tv,g)]):.3f}", va="center", fontsize=8)
    ax[0].legend(ncol=4, fontsize=7, frameon=False, loc="lower center", bbox_to_anchor=(0.5, -0.32))
    w = 0.38
    for j, (tv, col) in enumerate((("t1", "#9ecae1"), ("t2", "#2171b5"))):
        ax[2].bar(np.arange(4) + (j - 0.5) * w, [K(y, pr[(tv, g)]) for g in G], w, color=col, label=tv.upper())
        ax[2].hlines(K(y, cal[tv]), -0.5, 3.5, color=col, ls="--", lw=1); ax[2].hlines(K(y, lora_full[tv]), -0.5, 3.5, color=col, ls=":", lw=1.4)
    ax[2].set_xticks(range(4)); ax[2].set_xticklabels([nm[g] for g in G], rotation=20, fontsize=8); ax[2].set_ylabel("κ"); ax[2].set_ylim(0, 0.75); ax[2].legend(fontsize=8, frameon=False, loc="upper center", ncol=2)
    ax[2].set_title(("虚线：1,000 标签校准读出；点线：本院 LoRA 全量" if lang == "zh" else "Dashed: 1,000-label calibrated readout; dotted: full local LoRA"), fontsize=8)
    for a, l in zip(ax, "ABC"): a.text(-0.1, 1.05, l, transform=a.transAxes, fontsize=12, fontweight="bold")
    plt.tight_layout()
    for ext in ("png", "pdf", "svg"): fig.savefig(f"{ROOT}/figures/fig5_guidance_{lang}.{ext}", dpi=300, bbox_inches="tight")
    plt.close(fig)
```
