## Supplementary material for "Non-generative readout with few-label calibration versus LoRA fine-tuning of a large language model for ASA physical status classification: accuracy, speed, cost and label efficiency in three data sources": Analysis code and aggregate results: 97_token_cost.py

```
"""同底模（Qwen3.5-9B 分词器）的词元消耗：每例输入词元、生成式输出词元、推理时前向/逐词解码步数；LoRA 训练处理的词元数；校准标注的前向词元数。输出 reports/token_cost.md。
读出与生成式提示取自 07_infer_zeroshot.py / 13_generative_zeroshot.py；输入文本为 T1 模板（本院 val、MOVER 子集）。输入词元用提示全文计数；生成式输出词元不再用已保存的 raw 文本（该文本截断在 400 字符，会低估），而取 98_gen_tokens.py 的实测计数（本院 val 生成式子集前 96 台）。"""
import os, re, numpy as np, pandas as pd
from transformers import AutoTokenizer
D = os.path.expanduser("~/research/NE_ERAS_JEV/data/derived"); R = os.path.expanduser("~/research/Anes_JEV_vs_LoRA/analysis/reports")
tok = AutoTokenizer.from_pretrained("/mnt/c/backup/Qwen3.5-9B"); n = lambda s: len(tok.encode(s, add_special_tokens=False))
PR = ("You are an anesthesiologist. Based on the preoperative information below, assign the ASA Physical Status class.\n\n%s\n\nOptions:\nA. ASA I\nB. ASA II\nC. ASA III\nD. ASA IV-V\nAnswer:")
PG = ("You are an anesthesiologist. Based on the preoperative information below, assign the ASA Physical Status class (I, II, III, IV-V).\n\n%s\n\n"
      "Give a brief rationale (at most 3 sentences), then end with exactly one line: 'Final answer: ASA <I|II|III|IV-V>'.")
out = ["# 同底模（Qwen3.5-9B 分词器）的词元消耗\n"]; rows = []
for name, f, genf, idsf in (("本院 val（T1）", "asa_common_local.csv", "pred_gen_local_val.csv", None), ("MOVER 子集（T1）", "asa_common_mover.csv", "pred_gen_mover_sub.csv", "mover_test_sub4000.csv")):
    c = pd.read_csv(f"{D}/{f}").set_index("op_id"); g = pd.read_csv(f"{D}/{genf}").set_index("op_id"); g = g[g.index.isin(c.index)]
    if name.startswith("本院"): c = c[c.split == "val"]
    ids = g.index[:1000] if len(g) > 1000 else g.index; pr = np.array([n(PR % c.loc[i, "text"]) for i in ids]); pg = np.array([n(PG % c.loc[i, "text"]) for i in ids])
    rows.append((name, len(ids), pr.mean(), pg.mean()))
out += ["| 数据 | 样本数 | 读出每例输入词元 | 生成式每例输入词元 |", "|---|---|---|---|"]
out += [f"| {a} | {b} | {c:.0f} | {d:.0f} |" for a, b, c, d in rows]
T0 = pd.read_csv(f"{R}/gen_tokens_think0.csv")
out += ["", "生成式输出词元（实测，本院 val 生成式子集前 %d 台，关闭思考模式，上限 300；MOVER 未重测输出词元）：" % len(T0), "", "| 模式 | 台数 | 均值 | 中位数 | 第 10–90 百分位 | 触及上限比例 | 含 Final answer 比例 |", "|---|---|---|---|---|---|---|"]
for nm, T in (("关闭思考（本研究使用）", T0),): out.append(f"| {nm} | {len(T)} | {T.n_out.mean():.0f} | {T.n_out.median():.0f} | {T.n_out.quantile(.1):.0f}–{T.n_out.quantile(.9):.0f} | {T.hit_cap.mean()*100:.1f}% | {T.final.mean()*100:.1f}% |")
a = rows[0]; P, PGm, G = a[2], a[3], T0.n_out.mean()
out += ["", f"以本院 val 为例（读出 {P:.0f} 个输入词元，生成式 {PGm:.0f} 个输入词元 + 平均 {G:.0f} 个输出词元）：", "",
        "| 项目 | 读出 | 生成式零样本 | LoRA（推理） |", "|---|---|---|---|",
        f"| 每例处理词元（输入 + 输出） | {P:.0f} + 0 | {PGm:.0f} + {G:.0f} | {P:.0f} + 0 |", f"| 每例顺序解码步数（逐词） | 0（一次前向，取末位 4 个选项 logits） | 约 {G:.0f} | 0 |",
        f"| 1 万例处理词元合计 | {P*1e4/1e6:.2f} M | {(PGm+G)*1e4/1e6:.2f} M（其中 {G*1e4/1e6:.2f} M 为逐词生成） | {P*1e4/1e6:.2f} M |", f"| 1 万例逐词解码步数 | 0 | {G*1e4/1e6:.2f} M | 0 |",
        "", "标签相关的一次性词元消耗（本院 T1，每例 %.0f 词元；训练 1 个 epoch，需前向与反向）：" % P, "", "| 项目 | 处理词元 | 说明 |", "|---|---|---|"]
for k in (100, 1000): out.append(f"| 读出 + {k} 标签校准 | {k*P/1e6:.3f} M（仅前向） | 对 {k} 例打分；3 个切点在 CPU 上拟合，不更新模型参数 |")
for k in (300, 1000, 4326): out.append(f"| LoRA 训练 n={k} | {k*P/1e6:.3f} M（前向 + 反向 + 梯度检查点） | 更新适配器参数；每个站点各训一次并保存适配器 |")
open(f"{R}/token_cost.md", "w").write("\n".join(out)); print("\n".join(out))
```
