## Supplementary material for "Non-generative readout with few-label calibration versus LoRA fine-tuning of a large language model for ASA physical status classification: accuracy, speed, cost and label efficiency in three data sources": Analysis code and aggregate results: 98_gen_tokens.py

```
"""实测生成式零样本的输出词元数（修正：此前保存的 raw 文本截断在 400 字符，不能用来计数）。用法：98_gen_tokens.py <gpu> <think:0|1> <N> <max_new>
取本院 val 生成式子集的前 N 台（T1），提示与 13_generative_zeroshot.py 相同；记录每例实际生成词元数（到 EOS 或上限）、是否触顶、是否出现 'Final answer'。输出 reports/gen_tokens_think{0|1}.csv（仅词元计数，不作精度评估）。"""
import os, re, sys, time, numpy as np, pandas as pd, torch
from transformers import AutoTokenizer, AutoModelForCausalLM
gpu, think, N, MX = int(sys.argv[1]), int(sys.argv[2]), int(sys.argv[3]), int(sys.argv[4]); D = os.path.expanduser("~/research/NE_ERAS_JEV/data/derived"); R = os.path.expanduser("~/research/Anes_JEV_vs_LoRA/analysis/reports")
c = pd.read_csv(f"{D}/asa_common_local.csv").set_index("op_id"); ids = pd.read_csv(f"{D}/pred_gen_local_val.csv").op_id.tolist()[:N]; d = c.loc[ids]
path = "/mnt/c/backup/Qwen3.5-9B"; tok = AutoTokenizer.from_pretrained(path); tok.padding_side = "left"
m = AutoModelForCausalLM.from_pretrained(path, dtype=torch.float16, device_map={"": gpu}).eval()
Q = ("You are an anesthesiologist. Based on the preoperative information below, assign the ASA Physical Status class (I, II, III, IV-V).\n\n%s\n\n"
     "Give a brief rationale (at most 3 sentences), then end with exactly one line: 'Final answer: ASA <I|II|III|IV-V>'.")
def prompt(t): return tok.apply_chat_template([{"role": "user", "content": Q % t}], tokenize=False, add_generation_prompt=True, enable_thinking=bool(think))
rows = []; bs = 8; t0 = time.time(); eos = {tok.eos_token_id, tok.convert_tokens_to_ids("<|im_end|>")}
for i in range(0, len(d), bs):
    b = d.iloc[i:i + bs]; x = tok([prompt(t) for t in b.text], return_tensors="pt", padding=True).to(f"cuda:{gpu}")
    with torch.no_grad(): g = m.generate(**x, max_new_tokens=MX, do_sample=False)
    gen = g[:, x.input_ids.shape[1]:].cpu().numpy()
    for r, row in zip(b.itertuples(), gen):
        n = len(row)
        for j, t in enumerate(row):
            if int(t) in eos: n = j + 1; break
        s = tok.decode(row, skip_special_tokens=True); rows.append(dict(op_id=r.Index, n_out=n, hit_cap=n >= MX and int(row[-1]) not in eos, final=("Final answer" in s)))
    pd.DataFrame(rows).to_csv(f"{R}/gen_tokens_think{think}.csv", index=False); print(i + bs, f"{time.time()-t0:.0f}s", flush=True)
print("完成")
```
