## Supplementary material for "Non-generative readout with few-label calibration versus LoRA fine-tuning of a large language model for ASA physical status classification: accuracy, speed, cost and label efficiency in three data sources": Analysis code and aggregate results: 99_adjacent.py

```
"""相邻类误差：各臂的准确率、相邻准确率（|差|≤1）、跨 ≥2 级的误差比例。本院 val（T1）与 MOVER 子集；校准用与 87 脚本相同的 1,000 标签。输出 reports/adjacent_errors.md"""
import os, numpy as np, pandas as pd
D = os.path.expanduser("~/research/NE_ERAS_JEV/data/derived"); R = os.path.expanduser("~/research/Anes_JEV_vs_LoRA/analysis/reports"); PC = ["p1", "p2", "p3", "p4"]
from sklearn.metrics import cohen_kappa_score as ck
K = lambda y, p: ck(y, p, weights="quadratic"); rd = lambda f: pd.read_csv(f"{D}/{f}.csv").set_index("op_id"); Ex = lambda a: (np.asarray(a) * np.arange(1, 5)).sum(1)
def cuts_fit(E, y):
    cu = np.array([1.5, 2.5, 3.5]); g = np.linspace(1.0, 4.0, 121)
    for _ in range(3):
        for j in range(3): cu[j] = max(g, key=lambda v: K(y, 1 + (E[:, None] > np.where(np.arange(3) == j, v, cu)).sum(1)))
    return cu
def stats(y, p): d = np.abs(p - y); return (p == y).mean(), (d <= 1).mean(), (d >= 2).mean()
out = ["# 相邻类误差（准确率 / 相邻准确率 |差|≤1 / 跨 ≥2 级误差比例）\n", "| 数据 | 臂 | n | 准确率 | 相邻准确率 | 跨 ≥2 级 |", "|---|---|---|---|---|---|"]
c = pd.read_csv(f"{D}/asa_common_local.csv", usecols=["op_id", "split", "asa4"]).set_index("op_id"); val = c[c.split == "val"].index; dev = c[c.split == "dev"].sample(1000, random_state=11).index; y = c.loc[val].asa4.astype(int).values
r = rd("pred_llm_local"); cu = cuts_fit(Ex(r.loc[dev][PC].values), c.loc[dev].asa4.astype(int).values)
arms = {"读出 零样本": r.loc[val][PC].values.argmax(1) + 1, "读出 + 1,000 标签校准": 1 + (Ex(r.loc[val][PC].values)[:, None] > cu).sum(1)}
for n in (1000, 4326):
    arms[f"LoRA n={n}（种子均值）"] = [rd(f"pred_lora_LOC{n}s{s}_local_val").loc[val][PC].values.argmax(1) + 1 for s in ("20261017", "11", "12")]
for nm, p in arms.items():
    ps = p if isinstance(p, list) else [p]; m = np.mean([stats(y, q) for q in ps], 0); out.append(f"| 本院 val | {nm} | {len(y):,} | {m[0]:.3f} | {m[1]:.3f} | {m[2]:.3f} |")
sub = pd.read_csv(f"{D}/mover_test_sub4000.csv").op_id.tolist(); rt = rd("pred_llm_mover_test"); rc = rd("pred_llm_mover_cal"); ids = [i for i in sub if i in rt.index]; y = rt.loc[ids].asa4.astype(int).values
cu = cuts_fit(Ex(rc[PC].values), rc.asa4.astype(int).values)
arms = {"读出 零样本": rt.loc[ids][PC].values.argmax(1) + 1, "读出 + 1,000 标签校准": 1 + (Ex(rt.loc[ids][PC].values)[:, None] > cu).sum(1)}
for n, tag in ((1000, "curve1000"), (3000, "curve3000")):
    arms[f"LoRA n={n}（种子均值）"] = [rd(f"pred_lora_{tag}{s}_mover_sub").loc[ids][PC].values.argmax(1) + 1 for s in ("", "s11", "s12")]
arms["LoRA n=12000（种子 20261017）"] = rd("pred_lora_mover12k_mover_test").loc[ids][PC].values.argmax(1) + 1
for nm, p in arms.items():
    ps = p if isinstance(p, list) else [p]; m = np.mean([stats(y, q) for q in ps], 0); out.append(f"| MOVER 子集 | {nm} | {len(y):,} | {m[0]:.3f} | {m[1]:.3f} | {m[2]:.3f} |")
open(f"{R}/adjacent_errors.md", "w").write("\n".join(out)); print("\n".join(out))
```
