## Supplementary material for "Non-generative readout with few-label calibration versus LoRA fine-tuning of a large language model for ASA physical status classification: accuracy, speed, cost and label efficiency in three data sources": Analysis code and aggregate results: 103_perm_avg.py

```
"""审稿修改：读出臂的选项顺序置换平均与中文模板敏感性。全量评价集（本院 val 2,753 + 本院校准池 1,000；MOVER 评价子集 4,000 + 校准池 1,000），不抽样。
模式 perm：英文模板，选项按多种顺序呈现，位置概率映射回类别；模式 zh1：T1 模板译为中文、提示壳保持英文；模式 zh2：模板与提示壳均为中文（顺序为原顺序）。
用法：103_perm_avg.py <gpu> <local|mover> <perm|zh1|zh2> [order ...]  输出 pred_pa_<ds>_<mode>_<order>.csv（op_id, asa4, p1..p4；类别概率）。断点续跑。"""
import os, sys, time, numpy as np, pandas as pd, torch
from transformers import AutoTokenizer, AutoModelForCausalLM
gpu, ds, mode = int(sys.argv[1]), sys.argv[2], sys.argv[3]; D = os.path.expanduser("~/research/NE_ERAS_JEV/data/derived")
ORD = {"orig": [0, 1, 2, 3], "rev": [3, 2, 1, 0], "s1": [1, 2, 3, 0], "s2": [2, 3, 0, 1], "s3": [3, 0, 1, 2], "r5": [1, 3, 0, 2]}
orders = sys.argv[4:] or (["rev", "s1", "s2", "s3", "r5"] if mode == "perm" else ["orig"])
if ds == "local":
    c = pd.read_csv(f"{D}/asa_common_local.csv"); dev = c[c.split == "dev"].op_id.sample(1000, random_state=11); ids = list(c[c.split == "val"].op_id) + list(dev)
else:
    c = pd.read_csv(f"{D}/asa_common_mover.csv"); ids = pd.read_csv(f"{D}/mover_test_sub4000.csv").op_id.tolist() + pd.read_csv(f"{D}/pred_llm_mover_cal.csv").op_id.tolist()
d = c[c.op_id.isin(ids)].drop_duplicates("op_id").reset_index(drop=True)
if mode == "calpool": d = c[c.split == "train80"].sample(5000, random_state=21).reset_index(drop=True)   # MOVER 校准池扩充：训练划分内随机 5,000 台，用于标签抽样敏感性
def zh_text(r):
    f = lambda v, fmt: "未知" if pd.isna(v) else fmt % v; yn = lambda k: "是" if getattr(r, k) else "否"
    return ("年龄：%s 岁；性别：%s；体重：%s kg；BMI：%s；就诊类型：%s\n术前生命体征（入手术室前最近一次）：血压 %s/%s mmHg，心率 %s 次/分\n既往史：高血压%s，冠心病%s，糖尿病%s\n术前检验：肌酐 %s mg/dL，血红蛋白 %s g/dL，白蛋白 %s g/dL\n拟行手术：%s") % (
        f(r.age, "%.0f"), "女" if r.female else "男", f(r.weight_kg, "%.0f"), f(r.bmi, "%.1f"), "住院" if r.inpatient else "门诊", f(r.sbp, "%.0f"), f(r.dbp, "%.0f"), f(r.hr, "%.0f"), yn("htn"), yn("chd"), yn("dm"),
        f(r.cr_mgdl, "%.2f"), f(r.hb_gdl, "%.1f"), f(r.alb_gdl, "%.1f"), (r.procedure if isinstance(r.procedure, str) and r.procedure else "未知"))
if mode.startswith("zh"): d["text"] = [zh_text(r) for r in d.itertuples()]
path = "/mnt/c/backup/Qwen3.5-9B"; tok = AutoTokenizer.from_pretrained(path); m = AutoModelForCausalLM.from_pretrained(path, dtype=torch.float16, device_map={"": gpu}).eval()
a1 = [tok.encode(" " + x, add_special_tokens=False)[-1] for x in "ABCD"]; a2 = [tok.encode(x, add_special_tokens=False)[-1] for x in "ABCD"]
NAMES = ["ASA I", "ASA II", "ASA III", "ASA IV-V"]
def prompt(t, perm):
    opts = "\n".join(f"{'ABCD'[i]}. {NAMES[perm[i]]}" for i in range(4))
    if mode == "zh2": return f"你是一名麻醉医师。根据下列术前信息，判定患者的 ASA 分级。\n\n{t}\n\n选项：\n{opts}\n答案："
    return f"You are an anesthesiologist. Based on the preoperative information below, assign the ASA Physical Status class.\n\n{t}\n\nOptions:\n{opts}\nAnswer:"
for od in orders:
    perm = ORD[od]; out = f"{D}/pred_pa_{ds}_{mode}_{od}.csv"; done = pd.read_csv(out) if os.path.exists(out) else pd.DataFrame(columns=["op_id"])
    todo = d[~d.op_id.isin(done.op_id)]; rows = []; t0 = time.time(); print(ds, mode, od, "待做", len(todo), flush=True)
    for i, r in enumerate(todo.itertuples()):
        x = tok(prompt(r.text, perm), return_tensors="pt").to(f"cuda:{gpu}")
        with torch.no_grad(): lg = m(**x).logits[0, -1].float()
        p = torch.softmax(torch.maximum(lg[a1], lg[a2]), 0).cpu().numpy(); q = np.zeros(4); q[perm] = p
        rows.append(dict(op_id=r.op_id, asa4=r.asa4, p1=q[0], p2=q[1], p3=q[2], p4=q[3]))
        if (i + 1) % 500 == 0: pd.concat([done, pd.DataFrame(rows)]).to_csv(out, index=False); print(od, i + 1, f"{(time.time()-t0)/(i+1):.3f}s/例", flush=True)
    pd.concat([done, pd.DataFrame(rows)]).to_csv(out, index=False)
print("完成")
```
