## Supplementary material for "Non-generative readout with few-label calibration versus LoRA fine-tuning of a large language model for ASA physical status classification: accuracy, speed, cost and label efficiency in three data sources": Analysis code and aggregate results: 105_review_analyses.py

```
"""审稿意见补分析（CPU 部分）：(1) 标签池匹配核对与标签抽样敏感性；(2) 非劣效界值 δ 敏感性；(3) 校准与 LoRA 的交点（break-even，κ 与准确率）；(4) 本院验证集亚组（性别、年龄）。输出 reports/review_analyses.md 与 review_labeldraw.csv"""
import os, numpy as np, pandas as pd
from sklearn.metrics import cohen_kappa_score as ck
D = os.path.expanduser("~/research/NE_ERAS_JEV/data/derived"); R = os.path.expanduser("~/research/Anes_JEV_vs_LoRA/analysis/reports"); PC = ["p1", "p2", "p3", "p4"]
K = lambda y, p: ck(y, p, weights="quadratic"); rd = lambda f: pd.read_csv(f"{D}/{f}.csv").set_index("op_id"); Ex = lambda a: (np.asarray(a) * np.arange(1, 5)).sum(1)
def cuts_fit(E, y):
    cu = np.array([1.5, 2.5, 3.5]); g = np.linspace(1.0, 4.0, 121)
    for _ in range(3):
        for j in range(3): cu[j] = max(g, key=lambda v: K(y, 1 + (E[:, None] > np.where(np.arange(3) == j, v, cu)).sum(1)))
    return cu
out = ["# 审稿意见补分析（CPU 部分）\n"]
c = pd.read_csv(f"{D}/asa_common_local.csv", usecols=["op_id", "split", "asa4", "age", "female"]).merge(pd.read_csv(f"{D}/asa_dataset_v1.csv", usecols=["operation_id", "patient_sn"]).rename(columns={"operation_id": "op_id"}), on="op_id").set_index("op_id")
dev = c[c.split == "dev"].index; val = c[c.split == "val"].index; r = rd("pred_llm_local"); yv = c.loc[val].asa4.astype(int).values; Ev = Ex(r.loc[val][PC].values); pid = c.loc[val].patient_sn.values
lora = {sd: rd(f"pred_lora_LOC1000s{sd}_local_val").loc[val][PC].values.argmax(1) + 1 for sd in ("20261017", "11", "12")}
# ---------- (1) 标签池匹配与抽样敏感性 ----------
out.append("## 标签池匹配（本院）\n")
cal11 = set(c[c.split == "dev"].reset_index().op_id.sample(1000, random_state=11))
for sd in (11, 12, 20261017):
    s = set(c[c.split == "dev"].reset_index().sample(1000, random_state=sd).op_id); out.append(f"- LoRA 种子 {sd} 的 1,000 训练标签与校准池重合 {len(s & cal11)}/1000")
rng = np.random.default_rng(2026); devl = np.array(dev); ydev = c.loc[dev].asa4.astype(int).values; Edev = Ex(r.loc[dev][PC].values); NDRAW = 50
preds = []; kd = []; ad = []
for j in range(NDRAW):
    i = rng.choice(len(devl), 1000, replace=False); cu = cuts_fit(Edev[i], ydev[i]); p = 1 + (Ev[:, None] > cu).sum(1); preds.append(p); kd.append(K(yv, p)); ad.append((p == yv).mean())
kl = [K(yv, lora[s]) for s in lora]; al = [(lora[s] == yv).mean() for s in lora]
out += ["", f"## 标签抽样敏感性（本院；从开发集 4,326 台中独立抽取 {NDRAW} 个 1,000 标签池，各自拟合 3 切点，在验证集 2,753 台上评估）\n",
        f"- 校准读出 κ：均值 {np.mean(kd):.3f}，标签抽样 SD {np.std(kd, ddof=1):.3f}，范围 {min(kd):.3f}–{max(kd):.3f}；准确率均值 {np.mean(ad):.3f}",
        f"- LoRA@1,000 三个种子 κ：{', '.join(f'{x:.3f}' for x in kl)}（均值 {np.mean(kl):.3f}，SD {np.std(kl, ddof=1):.3f}）；准确率均值 {np.mean(al):.3f}",
        "- 注：开发集仅 4,326 台，50 个池之间有重叠，SD 低估真实的池间变异；LoRA 只有 3 个训练池。"]
up = np.unique(pid); ix = {q: np.where(pid == q)[0] for q in up}; B = 2000; diffs = []; diffs_fixed = []
for _ in range(B):
    s = np.concatenate([ix[q] for q in rng.choice(up, len(up))]); j = rng.integers(NDRAW); sd = list(lora)[rng.integers(3)]
    diffs.append(K(yv[s], preds[j][s]) - K(yv[s], lora[sd][s]))
d = np.array(diffs); out.append(f"- **含标签抽样与种子变异的 H2 差值**（每次重抽：患者簇 + 随机一个校准池 + 随机一个 LoRA 种子，B={B}）：均值 {d.mean():+.3f}，95% 区间 {np.percentile(d, 2.5):+.3f} ~ {np.percentile(d, 97.5):+.3f}；下界 > −0.05：{'是' if np.percentile(d, 2.5) > -0.05 else '否'}")
pd.DataFrame({"draw": range(NDRAW), "kappa": kd, "acc": ad}).to_csv(f"{R}/review_labeldraw.csv", index=False)
import json; json.dump(dict(draw_k_mean=float(np.mean(kd)), draw_k_sd=float(np.std(kd, ddof=1)), draw_k_min=float(min(kd)), draw_k_max=float(max(kd)), draw_acc_mean=float(np.mean(ad)), lora_k_mean=float(np.mean(kl)), comb_diff=float(d.mean()), comb_lo=float(np.percentile(d, 2.5)), comb_hi=float(np.percentile(d, 97.5))), open(f"{R}/review_numbers_105.json", "w"), indent=1)
# ---------- 2. δ 敏感性 ----------
P = pd.read_csv(f"{R}/paired_differences.csv"); h2 = P[P.hyp == "H2"]
out += ["", "## 非劣效界值 δ 敏感性（H2，差值 95% 区间下界 > −δ）\n", "| 数据 | 对比 | 下界 | δ=0.03 | δ=0.05 | δ=0.10 |", "|---|---|---|---|---|---|"]
for _, x in h2.iterrows(): out.append(f"| {x.dataset} | {x.contrast} | {x.lo:+.3f} | {'成立' if x.lo > -0.03 else '不成立'} | {'成立' if x.lo > -0.05 else '不成立'} | {'成立' if x.lo > -0.10 else '不成立'} |")
# ---------- 3. break-even ----------
out += ["", "## 校准读出与 LoRA 的交点（log n 线性插值；校准曲线取同一 1,000 标签池的子集）\n"]
def cross(ns, lora_k, cal_fn):
    dd = [cal_fn(n) - l for n, l in zip(ns, lora_k)]
    for a in range(len(ns) - 1):
        if dd[a] > 0 >= dd[a + 1]: t = dd[a] / (dd[a] - dd[a + 1]); return float(np.exp(np.log(ns[a]) + t * (np.log(ns[a + 1]) - np.log(ns[a]))))
    return None
def interp(ks, vs): return lambda n: float(np.interp(np.log(n), np.log(ks), vs))
sets = {"本院 val（T1，κ）": ([100, 300, 1000, 4326], {"20261017": [0.461, 0.541, 0.553, 0.558], "11": None, "12": None}, interp([25, 50, 100, 200, 500, 1000], [0.491, 0.516, 0.526, 0.528, 0.535, 0.540]))}
# 本院 LoRA 逐种子 κ
loc = {}
for sd in ("20261017", "11", "12"):
    loc[sd] = [K(yv, rd(f"pred_lora_LOC{n}s{sd}_local_val").loc[val][PC].values.argmax(1) + 1) for n in (100, 300, 1000, 4326)]
calL = interp([25, 50, 100, 200, 500, 1000, 4326], [0.491, 0.516, 0.526, 0.528, 0.535, 0.540, 0.540])
cs = [cross([100, 300, 1000, 4326], loc[sd], calL) for sd in loc]; out.append(f"- 本院 κ：逐种子交点 {cs}（校准曲线在 k>1,000 按平台外推）；均值曲线交点 {round(cross([100, 300, 1000, 4326], list(np.mean([loc[s] for s in loc], 0)), calL))}")
acc_cal = (1 + (Ev[:, None] > cuts_fit(Edev[[list(dev).index(i) for i in c[c.split == 'dev'].reset_index().op_id.sample(1000, random_state=11)]], ydev[[list(dev).index(i) for i in c[c.split == 'dev'].reset_index().op_id.sample(1000, random_state=11)]])).sum(1) == yv).mean()
out.append(f"- 本院 准确率：校准读出(1,000) {acc_cal:.3f}；LoRA n=100/300/1000/4326（3 种子均值）：" + "，".join(f"{np.mean([(rd(f'pred_lora_LOC{n}s{sd}_local_val').loc[val][PC].values.argmax(1) + 1 == yv).mean() for sd in loc]):.3f}" for n in (100, 300, 1000, 4326)) + "。LoRA 在 n=100 时准确率已高于校准读出，准确率交点低于 100 标签。")
open(f"{R}/review_analyses.md", "w").write("\n".join(out)); print("\n".join(out))
```
