## Supplementary material for "Non-generative readout with few-label calibration versus LoRA fine-tuning of a large language model for ASA physical status classification: accuracy, speed, cost and label efficiency in three data sources": Analysis code and aggregate results: 105b_mover_labeldraw.py

```
"""MOVER 标签抽样敏感性：从训练划分内另取的 5,000 台（pred_pa_mover_calpool_orig）独立抽取 50 个 1,000 标签池，各自拟合切点，在评价子集 4,000 台上评估；与 LoRA@1,000 三个种子合并后给出含标签抽样的 H2 区间。输出追加到 reports/review_analyses.md 并写 review_numbers_105b.json"""
import os, json, numpy as np, pandas as pd
from sklearn.metrics import cohen_kappa_score as ck
D = os.path.expanduser("~/research/NE_ERAS_JEV/data/derived"); R = os.path.expanduser("~/research/Anes_JEV_vs_LoRA/analysis/reports"); PC = ["p1", "p2", "p3", "p4"]
K = lambda y, p: ck(y, p, weights="quadratic"); rd = lambda f: pd.read_csv(f"{D}/{f}.csv").set_index("op_id"); Ex = lambda a: (np.asarray(a) * np.arange(1, 5)).sum(1)
def cuts_fit(E, y):
    cu = np.array([1.5, 2.5, 3.5]); g = np.linspace(1.0, 4.0, 121)
    for _ in range(3):
        for j in range(3): cu[j] = max(g, key=lambda v: K(y, 1 + (E[:, None] > np.where(np.arange(3) == j, v, cu)).sum(1)))
    return cu
sub = pd.read_csv(f"{D}/mover_test_sub4000.csv").op_id.tolist(); pm = pd.read_csv(f"{D}/mover_pid_map.csv").set_index("op_id").pid; m = pd.read_csv(f"{D}/asa_common_mover.csv", usecols=["op_id", "asa4"]).set_index("op_id").asa4.astype(int)
t = rd("pred_llm_mover_test").loc[sub]; yv = m.loc[sub].values; Ev = Ex(t[PC].values); pool = rd("pred_pa_mover_calpool_orig"); yp = pool.asa4.astype(int).values; Ep = Ex(pool[PC].values)
lora = [rd(f"pred_lora_curve1000{sfx}_mover_sub").loc[sub][PC].values.argmax(1) + 1 for sfx in ("", "s11", "s12")]
rng = np.random.default_rng(2027); preds = []; kd = []; ad = []
for _ in range(50):
    i = rng.choice(len(yp), 1000, replace=False); cu = cuts_fit(Ep[i], yp[i]); p = 1 + (Ev[:, None] > cu).sum(1); preds.append(p); kd.append(K(yv, p)); ad.append((p == yv).mean())
kl = [K(yv, l) for l in lora]; pid = pm.loc[sub].values; up = np.unique(pid); ix = {q: np.where(pid == q)[0] for q in up}; dd = []
for _ in range(2000):
    s = np.concatenate([ix[q] for q in rng.choice(up, len(up))]); dd.append(K(yv[s], preds[rng.integers(50)][s]) - K(yv[s], lora[rng.integers(3)][s]))
d = np.array(dd); N = dict(k_mean=float(np.mean(kd)), k_sd=float(np.std(kd, ddof=1)), k_min=float(min(kd)), k_max=float(max(kd)), acc_mean=float(np.mean(ad)), lora_k=[float(x) for x in kl], diff=float(d.mean()), lo=float(np.percentile(d, 2.5)), hi=float(np.percentile(d, 97.5)))
json.dump(N, open(f"{R}/review_numbers_105b.json", "w"), indent=1)
txt = ["", "## MOVER 标签抽样敏感性（训练划分内另取 5,000 台，独立抽取 50 个 1,000 标签池；评价子集 n=4,000）\n", f"- 校准读出 κ：均值 {N['k_mean']:.3f}，池间 SD {N['k_sd']:.3f}，范围 {N['k_min']:.3f}–{N['k_max']:.3f}；准确率均值 {N['acc_mean']:.3f}", f"- LoRA@1,000 三个种子 κ：{', '.join(f'{x:.3f}' for x in kl)}（均值 {np.mean(kl):.3f}）",
       f"- 含标签抽样与种子变异的 H2 差值：{N['diff']:+.3f}（{N['lo']:+.3f} ~ {N['hi']:+.3f}）；下界 > −0.05：{'是' if N['lo'] > -0.05 else '否'}"]
open(f"{R}/review_analyses.md", "a").write("\n".join(txt)); print("\n".join(txt))
```
