## Supplementary material for "Non-generative readout with few-label calibration versus LoRA fine-tuning of a large language model for ASA physical status classification: accuracy, speed, cost and label efficiency in three data sources": Analysis code and aggregate results: 106_subgroup.py

```
"""本院验证集亚组（探索性）：性别、年龄（<65 / ≥65）。臂：读出零样本、读出+1,000 标签校准（池 11）、LoRA@1,000（三个种子均值）。κ 与准确率，患者级 bootstrap（B=1000）。输出 reports/subgroup_local.md"""
import os, numpy as np, pandas as pd
from sklearn.metrics import cohen_kappa_score as ck
D = os.path.expanduser("~/research/NE_ERAS_JEV/data/derived"); R = os.path.expanduser("~/research/Anes_JEV_vs_LoRA/analysis/reports"); PC = ["p1", "p2", "p3", "p4"]
K = lambda y, p: ck(y, p, weights="quadratic") if len(np.unique(y)) > 1 else np.nan; rd = lambda f: pd.read_csv(f"{D}/{f}.csv").set_index("op_id"); Ex = lambda a: (np.asarray(a) * np.arange(1, 5)).sum(1)
def cuts_fit(E, y):
    cu = np.array([1.5, 2.5, 3.5]); g = np.linspace(1.0, 4.0, 121)
    for _ in range(3):
        for j in range(3): cu[j] = max(g, key=lambda v: K(y, 1 + (E[:, None] > np.where(np.arange(3) == j, v, cu)).sum(1)))
    return cu
c = pd.read_csv(f"{D}/asa_common_local.csv", usecols=["op_id", "split", "asa4", "age", "female"]).merge(pd.read_csv(f"{D}/asa_dataset_v1.csv", usecols=["operation_id", "patient_sn"]).rename(columns={"operation_id": "op_id"}), on="op_id").set_index("op_id")
val = c[c.split == "val"].index; r = rd("pred_llm_local"); cal = c[c.split == "dev"].reset_index().op_id.sample(1000, random_state=11).values
cu = cuts_fit(Ex(r.loc[cal][PC].values), c.loc[cal].asa4.astype(int).values); v = c.loc[val]; y = v.asa4.astype(int).values; Ev = Ex(r.loc[val][PC].values)
A = {"读出零样本": r.loc[val][PC].values.argmax(1) + 1, "读出+1,000 标签校准": 1 + (Ev[:, None] > cu).sum(1)}
L = [rd(f"pred_lora_LOC1000s{sd}_local_val").loc[val][PC].values.argmax(1) + 1 for sd in ("20261017", "11", "12")]
pid = v.patient_sn.values; rng = np.random.default_rng(31)
groups = {"全部": np.ones(len(v), bool), "女性": v.female.values == 1, "男性": v.female.values == 0, "年龄 <65": v.age.values < 65, "年龄 ≥65": v.age.values >= 65}
out = ["# 本院验证集亚组（探索性；κ 与准确率，95% CI 为患者级 bootstrap）\n", "| 亚组 | n | ASA I/II/III/IV–V (%) | 读出零样本 κ | 校准读出 κ | LoRA@1,000 κ（3 种子均值） | 校准读出准确率 | LoRA 准确率 |", "|---|---|---|---|---|---|---|---|"]
for g, m in groups.items():
    idx = np.where(m)[0]; yg = y[idx]; dist = " / ".join(f"{x:.1f}" for x in np.bincount(yg - 1, minlength=4)[:4] / len(yg) * 100); up = np.unique(pid[idx]); ix = {q: idx[pid[idx] == q] for q in up}
    boots = [np.concatenate([ix[q] for q in rng.choice(up, len(up))]) for _ in range(1000)]
    def ci(fn): vals = np.array([fn(s) for s in boots]); return f"{fn(idx):.3f}（{np.percentile(vals, 2.5):.3f}–{np.percentile(vals, 97.5):.3f}）"
    out.append(f"| {g} | {len(idx):,} | {dist} | " + " | ".join([ci(lambda s: K(y[s], A['读出零样本'][s])), ci(lambda s: K(y[s], A['读出+1,000 标签校准'][s])), ci(lambda s: np.mean([K(y[s], p[s]) for p in L])), ci(lambda s: (y[s] == A['读出+1,000 标签校准'][s]).mean()), ci(lambda s: np.mean([(y[s] == p[s]).mean() for p in L]))]) + " |")
out.append("\n注：≥65 岁组的 ASA III 占 78.2%、无 I 级，组内 κ 受取值范围限制，不宜解读，宜以准确率为准。")
open(f"{R}/subgroup_local.md", "w").write("\n".join(out)); print("\n".join(out))
```
