## Supplementary material for "Non-generative readout with few-label calibration versus LoRA fine-tuning of a large language model for ASA physical status classification: accuracy, speed, cost and label efficiency in three data sources": Analysis code and aggregate results: 107_fig_flow.py

```
"""图 2：三个数据源的纳入与划分流程（STARD/TRIPOD-LLM 16a 式）。数字来自 reports/asa_dataset_v1.md、asa_mover_v1.md 与 vitaldb 构建记录。输出 figures/fig_flow_{zh,en}.*"""
import os
import matplotlib; matplotlib.use("Agg")
import matplotlib.pyplot as plt
from matplotlib import font_manager as fm
from matplotlib.patches import FancyBboxPatch
fm.fontManager.addfont("/usr/share/fonts/opentype/noto/NotoSansCJK-Regular.ttc")
plt.rcParams.update({"font.sans-serif": ["Noto Sans CJK SC", "DejaVu Sans"], "axes.unicode_minus": False, "pdf.fonttype": 42, "svg.fonttype": "none"})
ROOT = os.path.expanduser("~/research/Anes_JEV_vs_LoRA")
D = {"zh": dict(
  cols=["本院", "MOVER", "VitalDB"],
  boxes=[["手术麻醉概览\n8,852 台", "MOVER 患者信息表\n64,354 台", "VitalDB 病例\n6,388 例"],
         ["有麻醉开始时间与 ASA\n7,348 台（6,828 人）", "有 ASA I–V（去重后）\n57,504 台", "ASA 为 1–5\n6,242 台（5,961 人）"],
         ["年龄 ≥18 岁\n7,332 台", "年龄 ≥18 岁\n57,473 台（38,245 人）", ""],
         ["术前 30 天内有麻醉前访视\n7,080 台", "", ""],
         ["分析集 7,079 台（6,804 人）", "按患者划分", "按患者划分"],
         ["开发集 2021–24\n4,326 台", "开发 80%\n45,994 台", "训练 70%\n4,357 台"],
         ["时间外验证集 2025–26\n2,753 台（2,640 人）", "测试 20%\n11,479 台；评价子集 4,000 台（2,606 人）", "测试 30%\n1,885 台（1,788 人）"]],
  excl=["排除 1,504 台\n（缺开始时间或 ASA）", "排除 6,850 台\n（缺 ASA 等）", "排除 146 例\n（ASA 缺失或 6 级）", "排除 16 台（<18 岁）", "排除 31 台（<18 岁）", "排除 252 台\n（无术前 30 天内访视）", "排除 1 台（开发期患者的\n验证期再手术）"]),
 "en": dict(
  cols=["Local hospital", "MOVER", "VitalDB"],
  boxes=[["Anesthesia overview\n8,852 operations", "MOVER patient table\n64,354 operations", "VitalDB cases\n6,388"],
         ["With anesthesia start and ASA\n7,348 (6,828 patients)", "With ASA I–V (deduplicated)\n57,504", "ASA 1–5\n6,242 (5,961 patients)"],
         ["Age ≥18 y\n7,332", "Age ≥18 y\n57,473 (38,245 patients)", ""],
         ["Preoperative visit within 30 d\n7,080", "", ""],
         ["Analysis set 7,079 (6,804 patients)", "Split by patient", "Split by patient"],
         ["Development 2021–24\n4,326", "Development 80%\n45,994", "Training 70%\n4,357"],
         ["Temporal validation 2025–26\n2,753 (2,640 patients)", "Test 20%: 11,479\nEvaluation subset 4,000 (2,606 patients)", "Test 30%\n1,885 (1,788 patients)"]],
  excl=["Excluded 1,504\n(no start time or ASA)", "Excluded 6,850\n(no ASA etc.)", "Excluded 146\n(ASA missing or 6)", "Excluded 16 (<18 y)", "Excluded 31 (<18 y)", "Excluded 252\n(no visit within 30 d)", "Excluded 1 (re-operation of a\ndevelopment-period patient)"])}
for lang, t in D.items():
    fig, ax = plt.subplots(figsize=(11.5, 8.6)); ax.axis("off"); ax.set_xlim(0, 100); ax.set_ylim(0, 100)
    xs = [17, 50, 83]; w = 28; h = 7.0; ys = [92, 79, 66, 53, 40, 27, 14]
    for ci, (cx, cn) in enumerate(zip(xs, t["cols"])): ax.text(cx, 99, cn, ha="center", fontsize=11, fontweight="bold")
    for ri, row in enumerate(t["boxes"]):
        for ci, txt in enumerate(row):
            if not txt: continue
            fc = "#eef3fa" if ri < 5 else "#eef7ee"
            ax.add_patch(FancyBboxPatch((xs[ci] - w / 2, ys[ri] - h / 2), w, h, boxstyle="round,pad=0.2,rounding_size=1", fc=fc, ec="#2c5aa0" if ri < 5 else "#3f6a1e", lw=1))
            ax.text(xs[ci], ys[ri], txt, ha="center", va="center", fontsize=8.2, linespacing=1.3)
    # 竖直箭头（同列相邻非空盒）
    for ci in range(3):
        last = None
        for ri in range(7):
            if t["boxes"][ri][ci]:
                if last is not None:
                    ax.annotate("", xy=(xs[ci], ys[ri] + h / 2), xytext=(xs[ci], ys[last] - h / 2), arrowprops=dict(arrowstyle="-|>", color="#555", lw=1))
                last = ri
    # 排除说明：放在箭头右侧的小字
    ex_pos = [(0, 1), (1, 1), (2, 1), (0, 2), (1, 2), (0, 3), (0, 4)]
    for (ci, ri), txt in zip(ex_pos, t["excl"]):
        ax.text(xs[ci] + 1.8, (ys[ri - 1] + ys[ri]) / 2, txt.replace("\n", " "), ha="left", va="center", fontsize=6.6, color="#a33")
    fig.savefig(f"{ROOT}/figures/fig_flow_{lang}.png", dpi=300, bbox_inches="tight"); fig.savefig(f"{ROOT}/figures/fig_flow_{lang}.pdf", bbox_inches="tight"); fig.savefig(f"{ROOT}/figures/fig_flow_{lang}.svg", bbox_inches="tight"); plt.close(fig)
print("ok")
```
