## Supplementary material for "Non-generative readout with few-label calibration versus LoRA fine-tuning of a large language model for ASA physical status classification: accuracy, speed, cost and label efficiency in three data sources": Analysis code and aggregate results: 108_perm_analysis.py

```
"""审稿修改：选项顺序置换平均读出、中文 T1 模板、MOVER 标签抽样、break-even。读取 pred_pa_* 与既有预测，输出 reports/review_perm_zh.md 与 review_numbers.json（正文引用的数字）。"""
import os, json, numpy as np, pandas as pd
from sklearn.metrics import cohen_kappa_score as ck, roc_auc_score
D = os.path.expanduser("~/research/NE_ERAS_JEV/data/derived"); R = os.path.expanduser("~/research/Anes_JEV_vs_LoRA/analysis/reports"); PC = ["p1", "p2", "p3", "p4"]
K = lambda y, p: ck(y, p, weights="quadratic"); rd = lambda f: pd.read_csv(f"{D}/{f}.csv").set_index("op_id"); Ex = lambda a: (np.asarray(a) * np.arange(1, 5)).sum(1)
def cuts_fit(E, y):
    cu = np.array([1.5, 2.5, 3.5]); g = np.linspace(1.0, 4.0, 121)
    for _ in range(3):
        for j in range(3): cu[j] = max(g, key=lambda v: K(y, 1 + (E[:, None] > np.where(np.arange(3) == j, v, cu)).sum(1)))
    return cu
def ece(P, y, nb=10):
    conf = P.max(1); pred = P.argmax(1) + 1; b = np.minimum((conf * nb).astype(int), nb - 1); e = 0
    for k in range(nb):
        m = b == k
        if m.any(): e += m.mean() * abs((pred[m] == y[m]).mean() - conf[m].mean())
    return e
out = ["# 选项顺序置换平均与中文模板（审稿修改）\n"]; N = {}
OR = ["orig", "rev", "s1", "s2", "s3", "r5"]
def block(ds, val_ids, cal_ids, y_all, pid_all, lora_fn, label):
    global out
    P = {}
    for od in OR:
        f = rd("pred_llm_local" if ds == "local" else "pred_llm_mover_test") if od == "orig" else rd(f"pred_pa_{ds}_perm_{od}")
        if od == "orig":
            fc = rd("pred_llm_mover_cal") if ds == "mover" else f
            P[od] = (f.loc[val_ids][PC].values, fc.loc[cal_ids][PC].values)
        else: P[od] = (f.loc[val_ids][PC].values, f.loc[cal_ids][PC].values)
    yv = y_all.loc[val_ids].values; yc = y_all.loc[cal_ids].values; pids = pid_all.loc[val_ids].values
    rows = []; pv = {}
    for od in OR:
        v, cc = P[od]; cu = cuts_fit(Ex(cc), yc); raw = v.argmax(1) + 1; cal = 1 + (Ex(v)[:, None] > cu).sum(1); pv[od] = cal
        rows.append((od, K(yv, raw), K(yv, cal), np.mean([roc_auc_score((yv == k + 1).astype(int), v[:, k]) for k in range(4)]), (raw == (P['orig'][0].argmax(1) + 1)).mean()))
    Pv = np.mean([P[o][0] for o in OR], 0); Pc = np.mean([P[o][1] for o in OR], 0); cu = cuts_fit(Ex(Pc), yc); rawA = Pv.argmax(1) + 1; calA = 1 + (Ex(Pv)[:, None] > cu).sum(1)
    aucA = np.mean([roc_auc_score((yv == k + 1).astype(int), Pv[:, k]) for k in range(4)])
    out += [f"## {label}（评价 n={len(yv):,}，校准池 {len(yc):,}）\n", "| 顺序 | 未校准 κ | 校准 κ | 宏 AUC | 与原顺序预测一致率 |", "|---|---|---|---|---|"]
    out += [f"| {o} | {a:.3f} | {b:.3f} | {c:.3f} | {d:.3f} |" for o, a, b, c, d in rows]
    ks = [r[1] for r in rows]; kc = [r[2] for r in rows]
    out += [f"| **6 种顺序平均概率** | **{K(yv, rawA):.3f}** | **{K(yv, calA):.3f}** | {aucA:.3f} | — |", "",
            f"- 单一顺序未校准 κ 范围 {min(ks):.3f}–{max(ks):.3f}（均值 {np.mean(ks):.3f}）；校准后范围 {min(kc):.3f}–{max(kc):.3f}（均值 {np.mean(kc):.3f}）。",
            f"- 平均概率的 ECE：{ece(Pv, yv):.3f}；原顺序：{ece(P['orig'][0], yv):.3f}。"]
    N[ds] = dict(single_raw=[round(x, 3) for x in ks], single_cal=[round(x, 3) for x in kc], avg_raw=round(K(yv, rawA), 3), avg_cal=round(K(yv, calA), 3), auc_avg=round(aucA, 3), auc_orig=round(rows[0][3], 3), ece_avg=round(ece(Pv, yv), 3), ece_orig=round(ece(P['orig'][0], yv), 3))
    # H2：平均读出（校准）减 LoRA@1000（三种子均值），patient bootstrap
    L = lora_fn(val_ids); up = np.unique(pids); ix = {q: np.where(pids == q)[0] for q in up}; rng = np.random.default_rng(77); dd = []; d0 = []
    for _ in range(1000):
        s = np.concatenate([ix[q] for q in rng.choice(up, len(up))]); kl = np.mean([K(yv[s], l[s]) for l in L]); dd.append(K(yv[s], calA[s]) - kl); d0.append(K(yv[s], pv["orig"][s]) - kl)
    pt = K(yv, calA) - np.mean([K(yv, l) for l in L])
    out += [f"- **H2（顺序平均的校准读出 − LoRA@1,000 三种子均值）**：{pt:+.3f}（{np.percentile(dd, 2.5):+.3f} ~ {np.percentile(dd, 97.5):+.3f}）；对照（原顺序）：{K(yv, pv['orig']) - np.mean([K(yv, l) for l in L]):+.3f}（{np.percentile(d0, 2.5):+.3f} ~ {np.percentile(d0, 97.5):+.3f}）", ""]
    N[ds]["h2_avg"] = [round(pt, 3), round(np.percentile(dd, 2.5), 3), round(np.percentile(dd, 97.5), 3)]
# ---- 本院 ----
c = pd.read_csv(f"{D}/asa_common_local.csv", usecols=["op_id", "split", "asa4"]).merge(pd.read_csv(f"{D}/asa_dataset_v1.csv", usecols=["operation_id", "patient_sn"]).rename(columns={"operation_id": "op_id"}), on="op_id").set_index("op_id")
val = list(c[c.split == "val"].index); cal = list(c[c.split == "dev"].reset_index().op_id.sample(1000, random_state=11))
def full(f, n): return os.path.exists(f) and len(pd.read_csv(f)) >= n
if all(full(f"{D}/pred_pa_local_perm_{o}.csv", 3753) for o in OR[1:]):
    block("local", val, cal, c.asa4.astype(int), c.patient_sn, lambda ids: [rd(f"pred_lora_LOC1000s{sd}_local_val").loc[ids][PC].values.argmax(1) + 1 for sd in ("20261017", "11", "12")], "本院验证集（T1 英文）")
# ---- MOVER ----
if all(full(f"{D}/pred_pa_mover_perm_{o}.csv", 5000) for o in OR[1:]):
    sub = pd.read_csv(f"{D}/mover_test_sub4000.csv").op_id.tolist(); pm = pd.read_csv(f"{D}/mover_pid_map.csv").set_index("op_id").pid; m = pd.read_csv(f"{D}/asa_common_mover.csv", usecols=["op_id", "asa4"]).set_index("op_id").asa4.astype(int)
    calm = pd.read_csv(f"{D}/pred_llm_mover_cal.csv").op_id.tolist()
    def mlora(ids): return [rd(f"pred_lora_curve1000{sfx}_mover_sub").loc[ids][PC].values.argmax(1) + 1 for sfx in ("", "s11", "s12")]
    block("mover", sub, calm, m, pm, mlora, "MOVER 评价子集（T1 英文）")
# ---- 中文模板 ----
yv = c.loc[val].asa4.astype(int).values; yc = c.loc[cal].asa4.astype(int).values; pids = c.loc[val].patient_sn.values
out += ["## 中文 T1 模板（本院验证集，n=2,753；校准池 1,000，同池 11）\n", "| 条件 | 未校准 κ | 校准 κ | 宏 AUC |", "|---|---|---|---|"]
r0 = rd("pred_llm_local"); r2 = rd("pred_llm_local_t2"); zh = {}
cond = {"T1 英文模板 + 英文提示壳": (r0.loc[val][PC].values, r0.loc[cal][PC].values), "T2 中文原始访视文本 + 英文提示壳": (r2.loc[val][PC].values, r2.loc[cal][PC].values)}
for md, nm in (("zh1", "T1 中文模板 + 英文提示壳"), ("zh2", "T1 中文模板 + 中文提示壳")):
    f = f"{D}/pred_pa_local_{md}_orig.csv"
    if full(f, 3753): z = rd(f"pred_pa_local_{md}_orig"); cond[nm] = (z.loc[val][PC].values, z.loc[cal][PC].values)
for nm, (v, cc) in cond.items():
    cu = cuts_fit(Ex(cc), yc); raw = v.argmax(1) + 1; calp = 1 + (Ex(v)[:, None] > cu).sum(1); auc = np.mean([roc_auc_score((yv == k + 1).astype(int), v[:, k]) for k in range(4)])
    out.append(f"| {nm} | {K(yv, raw):.3f} | {K(yv, calp):.3f} | {auc:.3f} |"); zh[nm] = [round(K(yv, raw), 3), round(K(yv, calp), 3), round(auc, 3)]
N["zh"] = zh
open(f"{R}/review_perm_zh.md", "w").write("\n".join(out)); json.dump(N, open(f"{R}/review_numbers.json", "w"), ensure_ascii=False, indent=1); print("\n".join(out))
```
