## Supplementary material for "Non-generative readout with few-label calibration versus LoRA fine-tuning of a large language model for ASA physical status classification: accuracy, speed, cost and label efficiency in three data sources": Analysis code and aggregate results: 109_breakeven.py

```
"""break-even：校准读出与 LoRA 的 κ 交点（逐种子，log n 线性插值；校准曲线在 k>1,000 按 k=1,000 水平外推）与准确率比较。数据：本院、MOVER、VitalDB T2。输出 reports/breakeven.md"""
import os, re, glob, numpy as np, pandas as pd
from sklearn.metrics import cohen_kappa_score as ck
D = os.path.expanduser("~/research/NE_ERAS_JEV/data/derived"); R = os.path.expanduser("~/research/Anes_JEV_vs_LoRA/analysis/reports"); PC = ["p1", "p2", "p3", "p4"]
K = lambda y, p: ck(y, p, weights="quadratic"); rd = lambda f: pd.read_csv(f"{D}/{f}.csv").set_index("op_id"); Ex = lambda a: (np.asarray(a) * np.arange(1, 5)).sum(1)
def cuts_fit(E, y):
    cu = np.array([1.5, 2.5, 3.5]); g = np.linspace(1.0, 4.0, 121)
    for _ in range(3):
        for j in range(3): cu[j] = max(g, key=lambda v: K(y, 1 + (E[:, None] > np.where(np.arange(3) == j, v, cu)).sum(1)))
    return cu
def interp(ks, vs): return lambda n: float(np.interp(np.log(n), np.log(ks), vs))
def cross(ns, lk, cal):
    dd = [cal(n) - l for n, l in zip(ns, lk)]
    if dd[0] <= 0: return f"<{ns[0]}"
    for a in range(len(ns) - 1):
        if dd[a] > 0 >= dd[a + 1]: t = dd[a] / (dd[a] - dd[a + 1]); return round(float(np.exp(np.log(ns[a]) + t * (np.log(ns[a + 1]) - np.log(ns[a])))))
    return f">{ns[-1]}"
fmtd = lambda d: "；".join(f"种子 {k}：{v}" for k, v in d.items())
out = ["# Break-even（校准读出 vs LoRA）\n", "校准曲线取自同一 1,000 标签池的子集（每个 k 重复抽样 100 次的均值）；k>1,000 按 k=1,000 的水平外推；LoRA 为各种子的 κ。交点为 log n 线性插值。\n"]
# 本院
c = pd.read_csv(f"{D}/asa_common_local.csv", usecols=["op_id", "split", "asa4"]).set_index("op_id"); val = c[c.split == "val"].index; yv = c.loc[val].asa4.astype(int).values
ns = [100, 300, 1000, 4326]; L = {sd: [rd(f"pred_lora_LOC{n}s{sd}_local_val").loc[val][PC].values.argmax(1) + 1 for n in ns] for sd in ("20261017", "11", "12")}
cal = interp([25, 50, 100, 200, 500, 1000, 4326], [0.491, 0.516, 0.526, 0.528, 0.535, 0.540, 0.540])
cs = {sd: cross(ns, [K(yv, p) for p in L[sd]], cal) for sd in L}
out.append(f"- **本院（κ）**：逐种子交点 {fmtd(cs)}；均值曲线 {round(cross(ns, list(np.mean([[K(yv, p) for p in L[sd]] for sd in L], 0)), cal))}")
accL = [float(np.mean([(L[sd][i] == yv).mean() for sd in L])) for i in range(4)]; out.append(f"- 本院 LoRA 准确率（3 种子均值）n={ns}：{[round(a, 3) for a in accL]}；校准读出（池 11）0.666，（50 个池均值）0.693。")
# MOVER
m = pd.read_csv(f"{D}/asa_common_mover.csv", usecols=["op_id", "asa4"]).set_index("op_id").asa4.astype(int); sub = pd.read_csv(f"{D}/mover_test_sub4000.csv").op_id.tolist(); ym = m.loc[sub].values
nm = [100, 300, 1000, 3000, 12000]
def mp(n, sfx): return rd(f"pred_lora_curve{n}{sfx}_mover_sub" if n != 12000 or sfx else "pred_lora_mover12k_mover_test").loc[sub][PC].values.argmax(1) + 1
LM = {}
for sfx, nmx in (("", "20261017"), ("s11", "11"), ("s12", "12")):
    LM[nmx] = [rd(f"pred_lora_curve{n}{sfx}_mover_sub").loc[sub][PC].values.argmax(1) + 1 if os.path.exists(f"{D}/pred_lora_curve{n}{sfx}_mover_sub.csv") else rd("pred_lora_mover12k_mover_test").loc[sub][PC].values.argmax(1) + 1 for n in nm]
calm = interp([25, 50, 100, 200, 500, 1000, 12000], [0.467, 0.495, 0.508, 0.517, 0.521, 0.524, 0.524])
csm = {sd: cross(nm, [K(ym, p) for p in LM[sd]], calm) for sd in LM}
out.append(f"- **MOVER（κ）**：逐种子交点 {fmtd(csm)}（校准曲线在 k>1,000 外推）")
out.append(f"- MOVER LoRA 准确率（3 种子均值）n={nm}：{[round(float(np.mean([(LM[sd][i] == ym).mean() for sd in LM])), 3) for i in range(5)]}；校准读出（池 11）0.515。")
# VitalDB T2
V = pd.read_csv(f"{D}/asa_common_vitaldb_t2.csv", usecols=["op_id", "split", "asa4"]).set_index("op_id"); tid = V[V.split == "test"].index; yt = V.loc[tid].asa4.astype(int).values
nv = [100, 300, 1000, 3000]; LV = {}
for f in glob.glob(f"{D}/pred_vdb_lora_VDB_t2_*_test.csv"):
    mm = re.search(r"VDB_t2_(\d+)_(\d+)_test", f); LV.setdefault(mm.group(2), {})[int(mm.group(1))] = rd(os.path.basename(f)[:-4]).loc[tid][PC].values.argmax(1) + 1
calv = interp([25, 50, 100, 200, 500, 1000, 3000], [0.509, 0.533, 0.549, 0.555, 0.563, 0.567, 0.567])
csv_ = {sd: cross(nv, [K(yt, LV[sd][n]) for n in nv], calv) for sd in LV}
out.append(f"- **VitalDB T2（κ）**：逐种子交点 {fmtd(csv_)}")
out.append(f"- VitalDB T2 LoRA 准确率（3 种子均值）n={nv}：{[round(float(np.mean([(LV[sd][n] == yt).mean() for sd in LV])), 3) for n in nv]}")
open(f"{R}/breakeven.md", "w").write("\n".join(out)); print("\n".join(out))
```
