## Supplementary material for "Non-generative readout with few-label calibration versus LoRA fine-tuning of a large language model for ASA physical status classification: accuracy, speed, cost and label efficiency in three data sources": Analysis code and aggregate results: 111_lora_fp16_xcard.py

```
"""审稿修改：fp16 LoRA 的跨卡补跑（本院 n=1,000，种子 11，T1，与 4-bit LoRA 同一批标签）。底模为 fp16（无量化），按层切分到两张卡（device_map="auto"，模型并行，非张量并行）；
超参与 08_lora.py 一致（r=16, α=32, dropout 0.05，lr 1e-4 单周期余弦，1 epoch，有效批 16，损失为四个选项字母 logits 的交叉熵），改用 fp16 autocast + GradScaler（SM75 无 bf16）。
训练后在本院 val 全量上推理，并记录批量 1 的延迟与各卡峰值显存。输出：models/LOC1000s11fp16x2、pred_lora_LOC1000s11fp16x2_local_val.csv、reports/lora_fp16_xcard.json"""
import os, sys, time, json, numpy as np, pandas as pd, torch
from transformers import AutoTokenizer, AutoModelForCausalLM
from peft import LoraConfig, get_peft_model
SEED = 11; torch.manual_seed(SEED); D = os.path.expanduser("~/research/NE_ERAS_JEV/data/derived"); M = os.path.expanduser("~/research/Anes_JEV_vs_LoRA/models"); R = os.path.expanduser("~/research/Anes_JEV_vs_LoRA/analysis/reports")
path = "/mnt/c/backup/Qwen3.5-9B"; tag = "LOC1000s11fp16x2"; N = 1000
P = "You are an anesthesiologist. Based on the preoperative information below, assign the ASA Physical Status class.\n\n%s\n\nOptions:\nA. ASA I\nB. ASA II\nC. ASA III\nD. ASA IV-V\nAnswer:"
tok = AutoTokenizer.from_pretrained(path); tok.padding_side = "left"; ids = torch.tensor([tok.encode(" " + c, add_special_tokens=False)[-1] for c in "ABCD"])
c = pd.read_csv(f"{D}/asa_common_local.csv"); dev = c[c.split == "dev"]; d = dev.sample(N, random_state=SEED).reset_index(drop=True)
t_load = time.time()
m = AutoModelForCausalLM.from_pretrained(path, dtype=torch.float16, device_map="auto", max_memory={0: "20GiB", 1: "20GiB"})
print("设备映射（各卡参数量）:", {str(k): sum(1 for v in m.hf_device_map.values() if v == k) for k in set(m.hf_device_map.values())}, f"载入 {time.time()-t_load:.0f}s", flush=True)
m.gradient_checkpointing_enable(gradient_checkpointing_kwargs={"use_reentrant": False}); m.enable_input_require_grads()
m = get_peft_model(m, LoraConfig(r=16, lora_alpha=32, lora_dropout=0.05, target_modules=["q_proj", "k_proj", "v_proj", "o_proj", "gate_proj", "up_proj", "down_proj"], task_type="CAUSAL_LM")); m.print_trainable_parameters()
first = next(m.parameters()).device; first = torch.device("cuda:0")
def fwd(texts):
    x = tok([P % t for t in texts], return_tensors="pt", padding=True).to(first)
    return m(**x, logits_to_keep=1).logits[:, -1].float()[:, ids.to(m.device if hasattr(m, "device") else first)] if False else m(**x, logits_to_keep=1).logits[:, -1].float().cpu()[:, ids]
params = [p for p in m.parameters() if p.requires_grad]; opt = torch.optim.AdamW(params, lr=1e-4, weight_decay=0.0); bs, acc = 4, 4; steps = N // (bs * acc)
sched = torch.optim.lr_scheduler.OneCycleLR(opt, max_lr=1e-4, total_steps=steps, pct_start=0.05, anneal_strategy="cos"); scaler = torch.amp.GradScaler("cuda")
m.train(); t0 = time.time(); la = []
for s in range(steps):
    for a in range(acc):
        b = d.iloc[(s * acc + a) * bs:(s * acc + a + 1) * bs]
        with torch.autocast("cuda", dtype=torch.float16): lg = fwd(list(b.text))
        loss = torch.nn.functional.cross_entropy(lg, torch.tensor(b.asa4.values - 1)) / acc; scaler.scale(loss).backward(); la.append(loss.item() * acc)
    scaler.unscale_(opt); torch.nn.utils.clip_grad_norm_(params, 1.0); scaler.step(opt); scaler.update(); sched.step(); opt.zero_grad()
    if (s + 1) % 10 == 0: print(f"step {s+1}/{steps} loss {np.mean(la[-40:]):.4f} {(time.time()-t0)/(s+1):.1f}s/step", flush=True)
train_s = time.time() - t0; peak_train = [torch.cuda.max_memory_allocated(i) / 1e9 for i in range(2)]; m.save_pretrained(f"{M}/{tag}"); print("训练完成", f"{train_s:.0f}s", "峰值显存(GB)", peak_train, flush=True)
m.eval(); v = c[c.split == "val"].reset_index(drop=True); rows = []; t1 = time.time()
for i in range(0, len(v), 8):
    b = v.iloc[i:i + 8]
    with torch.no_grad(), torch.autocast("cuda", dtype=torch.float16): p = torch.softmax(fwd(list(b.text)), -1).numpy()
    for r, q in zip(b.itertuples(), p): rows.append(dict(op_id=r.op_id, asa4=r.asa4, p1=q[0], p2=q[1], p3=q[2], p4=q[3]))
    if (i // 8 + 1) % 50 == 0: print("推理", i + 8, f"{(time.time()-t1)/(i+8):.3f}s/例", flush=True)
pd.DataFrame(rows).to_csv(f"{D}/pred_lora_{tag}_local_val.csv", index=False); infer_s = time.time() - t1
lat = []
for t in list(v.text[:5]): 
    with torch.no_grad(), torch.autocast("cuda", dtype=torch.float16): fwd([t])
for t in list(v.text[5:105]):
    torch.cuda.synchronize(); t2 = time.time()
    with torch.no_grad(), torch.autocast("cuda", dtype=torch.float16): fwd([t])
    torch.cuda.synchronize(); lat.append(time.time() - t2)
J = dict(tag=tag, train_s=train_s, infer_s=infer_s, p50_ms=float(np.percentile(lat, 50) * 1000), p95_ms=float(np.percentile(lat, 95) * 1000), peak_train_gb=peak_train, peak_all_gb=[torch.cuda.max_memory_allocated(i) / 1e9 for i in range(2)], final_loss=float(np.mean(la[-16:])))
json.dump(J, open(f"{R}/lora_fp16_xcard.json", "w"), indent=1); print(J)
```
