## Supplementary material for "Non-generative readout with few-label calibration versus LoRA fine-tuning of a large language model for ASA physical status classification: accuracy, speed, cost and label efficiency in three data sources": Analysis code and aggregate results: 112_fp16_lora_eval.py

```
"""fp16 LoRA（跨卡补跑，同一批 1,000 标签，种子 11）与 4-bit LoRA、校准读出的比较（本院 val，n=2,753；患者级 bootstrap）。输出 reports/lora_fp16_xcard.md"""
import os, json, numpy as np, pandas as pd
from sklearn.metrics import cohen_kappa_score as ck, roc_auc_score
D = os.path.expanduser("~/research/NE_ERAS_JEV/data/derived"); R = os.path.expanduser("~/research/Anes_JEV_vs_LoRA/analysis/reports"); PC = ["p1", "p2", "p3", "p4"]
K = lambda y, p: ck(y, p, weights="quadratic"); rd = lambda f: pd.read_csv(f"{D}/{f}.csv").set_index("op_id"); Ex = lambda a: (np.asarray(a) * np.arange(1, 5)).sum(1)
def cuts_fit(E, y):
    cu = np.array([1.5, 2.5, 3.5]); g = np.linspace(1.0, 4.0, 121)
    for _ in range(3):
        for j in range(3): cu[j] = max(g, key=lambda v: K(y, 1 + (E[:, None] > np.where(np.arange(3) == j, v, cu)).sum(1)))
    return cu
c = pd.read_csv(f"{D}/asa_common_local.csv", usecols=["op_id", "split", "asa4"]).merge(pd.read_csv(f"{D}/asa_dataset_v1.csv", usecols=["operation_id", "patient_sn"]).rename(columns={"operation_id": "op_id"}), on="op_id").set_index("op_id")
val = c[c.split == "val"].index; y = c.loc[val].asa4.astype(int).values; pid = c.loc[val].patient_sn.values
f16 = rd("pred_lora_LOC1000s11fp16x2_local_val").loc[val][PC].values; q4 = rd("pred_lora_LOC1000s11_local_val").loc[val][PC].values
cal = c[c.split == "dev"].reset_index().op_id.sample(1000, random_state=11).values; r = rd("pred_llm_local"); cu = cuts_fit(Ex(r.loc[cal][PC].values), c.loc[cal].asa4.astype(int).values)
arms = {"fp16 LoRA（跨两卡，n=1,000，种子 11）": f16.argmax(1) + 1, "4-bit LoRA（单卡，同一批 1,000 标签）": q4.argmax(1) + 1, "fp16 读出 + 1,000 标签校准（同池）": 1 + (Ex(r.loc[val][PC].values)[:, None] > cu).sum(1)}
up = np.unique(pid); ix = {q: np.where(pid == q)[0] for q in up}; rng = np.random.default_rng(8); B = [np.concatenate([ix[q] for q in rng.choice(up, len(up))]) for _ in range(1000)]
KB = {n: np.array([K(y[s], p[s]) for s in B]) for n, p in arms.items()}; names = list(arms)
out = ["# fp16 LoRA 跨卡补跑（本院 val，n=2,753）\n", "| 臂 | κ（95% CI） | 准确率 | 宏 AUC |", "|---|---|---|---|"]
for n, p in arms.items():
    PP = {names[0]: f16, names[1]: q4}.get(n); auc = "—" if PP is None else f"{np.mean([roc_auc_score((y == k + 1).astype(int), PP[:, k]) for k in range(4)]):.3f}"
    out.append(f"| {n} | {K(y, p):.3f}（{np.percentile(KB[n], 2.5):.3f}–{np.percentile(KB[n], 97.5):.3f}） | {(p == y).mean():.3f} | {auc} |")
def d(a, b): v = KB[a] - KB[b]; return f"{K(y, arms[a]) - K(y, arms[b]):+.3f}（{np.percentile(v, 2.5):+.3f} ~ {np.percentile(v, 97.5):+.3f}）"
out += ["", f"- fp16 LoRA − 4-bit LoRA（同一批标签）：{d(names[0], names[1])}", f"- **H2（标签匹配，fp16 LoRA）**：fp16 读出校准 − fp16 LoRA = {d(names[2], names[0])}"]
J = json.load(open(f"{R}/lora_fp16_xcard.json")); out += ["", f"- 训练 {J['train_s']:.0f} s（62 步，跨两卡层切分）；全量 val 推理 {J['infer_s']:.0f} s；批量 1 延迟 p50 {J['p50_ms']:.0f} ms、p95 {J['p95_ms']:.0f} ms（含卡间传输，适配器未合并）；训练峰值显存 {J['peak_train_gb'][0]:.1f}/{J['peak_train_gb'][1]:.1f} GB（两卡）。"]
open(f"{R}/lora_fp16_xcard.md", "w").write("\n".join(out)); print("\n".join(out))
```
