## Supplementary material for "Non-generative readout with few-label calibration versus LoRA fine-tuning of a large language model for ASA physical status classification: accuracy, speed, cost and label efficiency in three data sources": Analysis code and aggregate results: 113_readout4.py

```
"""4-bit（NF4）量化的零样本读出：本院（val 2,753 + 校准池 1,000）或 MOVER（评价子集 4,000 + 校准池 1,000），自然顺序、英文 T1 模板、批量 1，与 07_infer_zeroshot.py 同一提示与读出。
用法：113_readout4.py <local|mover> <gpu>；输出 pred_q4_<ds>.csv（op_id, asa4, p1..p4）。断点续跑。"""
import os, sys, time, numpy as np, pandas as pd, torch
from transformers import AutoTokenizer, AutoModelForCausalLM, BitsAndBytesConfig
ds, gpu = sys.argv[1], int(sys.argv[2]); D = os.path.expanduser("~/research/NE_ERAS_JEV/data/derived")
if ds == "local":
    c = pd.read_csv(f"{D}/asa_common_local.csv"); cal = c[c.split == "dev"].op_id.sample(1000, random_state=11); ids = list(c[c.split == "val"].op_id) + list(cal)
else:
    c = pd.read_csv(f"{D}/asa_common_mover.csv"); ids = pd.read_csv(f"{D}/mover_test_sub4000.csv").op_id.tolist() + pd.read_csv(f"{D}/pred_llm_mover_cal.csv").op_id.tolist()
d = c[c.op_id.isin(ids)].drop_duplicates("op_id").reset_index(drop=True); out = f"{D}/pred_q4_{ds}.csv"
done = pd.read_csv(out) if os.path.exists(out) else pd.DataFrame(columns=["op_id"]); todo = d[~d.op_id.isin(done.op_id)]; print(ds, "总", len(d), "待做", len(todo), flush=True)
path = "/mnt/c/backup/Qwen3.5-9B"; tok = AutoTokenizer.from_pretrained(path)
bnb = BitsAndBytesConfig(load_in_4bit=True, bnb_4bit_quant_type="nf4", bnb_4bit_compute_dtype=torch.float16, bnb_4bit_use_double_quant=True)
m = AutoModelForCausalLM.from_pretrained(path, quantization_config=bnb, dtype=torch.float16, device_map={"": gpu}).eval()
a1 = [tok.encode(" " + x, add_special_tokens=False)[-1] for x in "ABCD"]; a2 = [tok.encode(x, add_special_tokens=False)[-1] for x in "ABCD"]
P = "You are an anesthesiologist. Based on the preoperative information below, assign the ASA Physical Status class.\n\n%s\n\nOptions:\nA. ASA I\nB. ASA II\nC. ASA III\nD. ASA IV-V\nAnswer:"
rows = []; t0 = time.time()
for i, r in enumerate(todo.itertuples()):
    x = tok(P % r.text, return_tensors="pt").to(f"cuda:{gpu}")
    with torch.no_grad(): lg = m(**x).logits[0, -1].float()
    q = torch.softmax(torch.maximum(lg[a1], lg[a2]), 0).cpu().numpy(); rows.append(dict(op_id=r.op_id, asa4=r.asa4, p1=q[0], p2=q[1], p3=q[2], p4=q[3]))
    if (i + 1) % 500 == 0: pd.concat([done, pd.DataFrame(rows)]).to_csv(out, index=False); print(i + 1, f"{(time.time()-t0)/(i+1):.3f}s/例", flush=True)
pd.concat([done, pd.DataFrame(rows)]).to_csv(out, index=False); print("完成")
```
