## Supplementary material for "Non-generative readout with few-label calibration versus LoRA fine-tuning of a large language model for ASA physical status classification: accuracy, speed, cost and label efficiency in three data sources": Analysis code and aggregate results: 114_readout4_eval.py

```
"""4-bit 读出对 fp16 读出：本院 val 与 MOVER 评价子集的 κ（未校准与 1,000 标签校准）、宏 AUC、预测一致率及配对差（患者级 bootstrap）。输出 reports/readout4_quant.md、readout4_numbers.json"""
import os, json, numpy as np, pandas as pd
from sklearn.metrics import cohen_kappa_score as ck, roc_auc_score
D = os.path.expanduser("~/research/NE_ERAS_JEV/data/derived"); R = os.path.expanduser("~/research/Anes_JEV_vs_LoRA/analysis/reports"); PC = ["p1", "p2", "p3", "p4"]
K = lambda y, p: ck(y, p, weights="quadratic"); rd = lambda f: pd.read_csv(f"{D}/{f}.csv").set_index("op_id"); Ex = lambda a: (np.asarray(a) * np.arange(1, 5)).sum(1)
def cuts_fit(E, y):
    cu = np.array([1.5, 2.5, 3.5]); g = np.linspace(1.0, 4.0, 121)
    for _ in range(3):
        for j in range(3): cu[j] = max(g, key=lambda v: K(y, 1 + (E[:, None] > np.where(np.arange(3) == j, v, cu)).sum(1)))
    return cu
out = ["# 4-bit（NF4）量化读出与 fp16 读出的比较（零样本，自然顺序，英文 T1）\n", "| 数据 | 精度 | 未校准 κ | 1,000 标签校准 κ | 宏 AUC | 与 fp16 预测一致率 |", "|---|---|---|---|---|---|"]; N = {}
def block(ds, val_ids, cal_ids, y_all, pid_all, f16_val, f16_cal, f4, label):
    yv = y_all.loc[val_ids].values; yc = y_all.loc[cal_ids].values; pid = pid_all.loc[val_ids].values
    r = {}
    for nm, (v, c_) in {"fp16": (f16_val.loc[val_ids][PC].values, f16_cal.loc[cal_ids][PC].values), "4-bit": (f4.loc[val_ids][PC].values, f4.loc[cal_ids][PC].values)}.items():
        cu = cuts_fit(Ex(c_), yc); raw = v.argmax(1) + 1; cal = 1 + (Ex(v)[:, None] > cu).sum(1); auc = np.mean([roc_auc_score((yv == k + 1).astype(int), v[:, k]) for k in range(4)]); r[nm] = (raw, cal, auc)
    agree = (r["fp16"][0] == r["4-bit"][0]).mean()
    for nm in ("fp16", "4-bit"): out.append(f"| {label} | {nm} | {K(yv, r[nm][0]):.3f} | {K(yv, r[nm][1]):.3f} | {r[nm][2]:.3f} | {'—' if nm == 'fp16' else f'{agree:.3f}'} |")
    up = np.unique(pid); ix = {q: np.where(pid == q)[0] for q in up}; rng = np.random.default_rng(4); B = [np.concatenate([ix[q] for q in rng.choice(up, len(up))]) for _ in range(1000)]
    d_raw = np.array([K(yv[s], r["4-bit"][0][s]) - K(yv[s], r["fp16"][0][s]) for s in B]); d_cal = np.array([K(yv[s], r["4-bit"][1][s]) - K(yv[s], r["fp16"][1][s]) for s in B])
    pt_raw = K(yv, r["4-bit"][0]) - K(yv, r["fp16"][0]); pt_cal = K(yv, r["4-bit"][1]) - K(yv, r["fp16"][1])
    out.append(f"|  | 4-bit − fp16 | {pt_raw:+.3f}（{np.percentile(d_raw, 2.5):+.3f} ~ {np.percentile(d_raw, 97.5):+.3f}） | {pt_cal:+.3f}（{np.percentile(d_cal, 2.5):+.3f} ~ {np.percentile(d_cal, 97.5):+.3f}） |  |  |")
    N[ds] = dict(fp16_raw=round(K(yv, r['fp16'][0]), 3), fp16_cal=round(K(yv, r['fp16'][1]), 3), q4_raw=round(K(yv, r['4-bit'][0]), 3), q4_cal=round(K(yv, r['4-bit'][1]), 3), d_raw=[round(pt_raw, 3), round(np.percentile(d_raw, 2.5), 3), round(np.percentile(d_raw, 97.5), 3)], d_cal=[round(pt_cal, 3), round(np.percentile(d_cal, 2.5), 3), round(np.percentile(d_cal, 97.5), 3)], auc16=round(r['fp16'][2], 3), auc4=round(r['4-bit'][2], 3), agree=round(agree, 3))
c = pd.read_csv(f"{D}/asa_common_local.csv", usecols=["op_id", "split", "asa4"]).merge(pd.read_csv(f"{D}/asa_dataset_v1.csv", usecols=["operation_id", "patient_sn"]).rename(columns={"operation_id": "op_id"}), on="op_id").set_index("op_id")
f4 = rd("pred_q4_local"); val = list(c[c.split == "val"].index); cal = list(c[c.split == "dev"].reset_index().op_id.sample(1000, random_state=11)); f16 = rd("pred_llm_local")
if len(f4) >= 3753: block("local", val, cal, c.asa4.astype(int), c.patient_sn, f16, f16, f4, "本院 val")
if os.path.exists(f"{D}/pred_q4_mover.csv") and len(rd("pred_q4_mover")) >= 5000:
    sub = pd.read_csv(f"{D}/mover_test_sub4000.csv").op_id.tolist(); pm = pd.read_csv(f"{D}/mover_pid_map.csv").set_index("op_id").pid; m = pd.read_csv(f"{D}/asa_common_mover.csv", usecols=["op_id", "asa4"]).set_index("op_id").asa4.astype(int)
    block("mover", sub, pd.read_csv(f"{D}/pred_llm_mover_cal.csv").op_id.tolist(), m, pm, rd("pred_llm_mover_test"), rd("pred_llm_mover_cal"), rd("pred_q4_mover"), "MOVER 子集")
open(f"{R}/readout4_quant.md", "w").write("\n".join(out)); json.dump(N, open(f"{R}/readout4_numbers.json", "w"), indent=1); print("\n".join(out))
```
