## Supplementary material for "Non-generative readout with few-label calibration versus LoRA fine-tuning of a large language model for ASA physical status classification: accuracy, speed, cost and label efficiency in three data sources": Analysis code and aggregate results: 115_precision_matched_h2.py

```
"""精度匹配的 H2：4-bit 校准读出 − 4-bit LoRA@1,000（本院：种子 11 标签匹配与三种子均值；MOVER：三种子均值），患者级 bootstrap。输出追加到 reports/readout4_quant.md，数字存 readout4_h2_numbers.json"""
import os, json, numpy as np, pandas as pd
from sklearn.metrics import cohen_kappa_score as ck
D = os.path.expanduser("~/research/NE_ERAS_JEV/data/derived"); R = os.path.expanduser("~/research/Anes_JEV_vs_LoRA/analysis/reports"); PC = ["p1", "p2", "p3", "p4"]
K = lambda y, p: ck(y, p, weights="quadratic"); rd = lambda f: pd.read_csv(f"{D}/{f}.csv").set_index("op_id"); Ex = lambda a: (np.asarray(a) * np.arange(1, 5)).sum(1)
def cf(E, y):
    cu = np.array([1.5, 2.5, 3.5]); g = np.linspace(1, 4, 121)
    for _ in range(3):
        for j in range(3): cu[j] = max(g, key=lambda v: K(y, 1 + (E[:, None] > np.where(np.arange(3) == j, v, cu)).sum(1)))
    return cu
def boot(pid, fn, seed=3):
    up = np.unique(pid); ix = {u: np.where(pid == u)[0] for u in up}; rng = np.random.default_rng(seed); return np.array([fn(np.concatenate([ix[u] for u in rng.choice(up, len(up))])) for _ in range(1000)])
res = {}; out = ["", "## 精度匹配的 H2（读出与 LoRA 同为 4-bit）", "", "| 数据 | 对比（校准读出 − LoRA@1,000，均为 4-bit） | 差值（95% CI） |", "|---|---|---|"]
c = pd.read_csv(f"{D}/asa_common_local.csv", usecols=["op_id", "split", "asa4"]).merge(pd.read_csv(f"{D}/asa_dataset_v1.csv", usecols=["operation_id", "patient_sn"]).rename(columns={"operation_id": "op_id"}), on="op_id").set_index("op_id")
val = list(c[c.split == "val"].index); cal = list(c[c.split == "dev"].reset_index().op_id.sample(1000, random_state=11)); y = c.loc[val].asa4.astype(int).values; pid = c.loc[val].patient_sn.values
q = rd("pred_q4_local"); cu = cf(Ex(q.loc[cal][PC].values), c.loc[cal].asa4.astype(int).values); cal4 = 1 + (Ex(q.loc[val][PC].values)[:, None] > cu).sum(1)
L = {sd: rd(f"pred_lora_LOC1000s{sd}_local_val").loc[val][PC].values.argmax(1) + 1 for sd in ("20261017", "11", "12")}
for nm, fn in (("本院 val，种子 11（标签匹配）", lambda s: K(y[s], cal4[s]) - K(y[s], L["11"][s])), ("本院 val，三种子均值", lambda s: K(y[s], cal4[s]) - np.mean([K(y[s], L[k][s]) for k in L]))):
    v = boot(pid, fn); pt = fn(np.arange(len(y))); res[nm] = [round(pt, 3), round(np.percentile(v, 2.5), 3), round(np.percentile(v, 97.5), 3)]; out.append(f"| {nm.split('，')[0]} | {nm.split('，')[1]} | {pt:+.3f}（{np.percentile(v, 2.5):+.3f} ~ {np.percentile(v, 97.5):+.3f}） |")
sub = pd.read_csv(f"{D}/mover_test_sub4000.csv").op_id.tolist(); pm = pd.read_csv(f"{D}/mover_pid_map.csv").set_index("op_id").pid; m = pd.read_csv(f"{D}/asa_common_mover.csv", usecols=["op_id", "asa4"]).set_index("op_id").asa4.astype(int)
qm = rd("pred_q4_mover"); calm = pd.read_csv(f"{D}/pred_llm_mover_cal.csv").op_id.tolist(); ym = m.loc[sub].values; cum = cf(Ex(qm.loc[calm][PC].values), m.loc[calm].values); calq = 1 + (Ex(qm.loc[sub][PC].values)[:, None] > cum).sum(1)
LM = [rd(f"pred_lora_curve1000{s}_mover_sub").loc[sub][PC].values.argmax(1) + 1 for s in ("", "s11", "s12")]
fn = lambda s: K(ym[s], calq[s]) - np.mean([K(ym[s], l[s]) for l in LM]); v = boot(pm.loc[sub].values, fn); pt = fn(np.arange(len(ym))); res["MOVER 三种子均值"] = [round(pt, 3), round(np.percentile(v, 2.5), 3), round(np.percentile(v, 97.5), 3)]
out.append(f"| MOVER 子集 | 三种子均值 | {pt:+.3f}（{np.percentile(v, 2.5):+.3f} ~ {np.percentile(v, 97.5):+.3f}） |")
out += ["", "对照：均为 fp16 的标签匹配对比（fp16 读出校准 − fp16 LoRA，本院，种子 11）为 −0.019（−0.045 ~ +0.006），见 fp16 LoRA 跨卡补跑。"]
json.dump(res, open(f"{R}/readout4_h2_numbers.json", "w"), ensure_ascii=False, indent=1); open(f"{R}/readout4_quant.md", "a").write("\n".join(out)); print("\n".join(out))
```
