## Supplementary material for "Non-generative readout with few-label calibration versus LoRA fine-tuning of a large language model for ASA physical status classification: accuracy, speed, cost and label efficiency in three data sources": Analysis code and aggregate results: 116_h1_subset_acc.py

```
"""审稿修改：(1) H1 生成式臂子集与未纳入病例的可比性（本院随机 1,000 台；MOVER 评价子集前 2,000 台 vs 后 2,000 台）；(2) 准确率的配对差与 95% CI（LoRA 对校准读出，n=100/300/1000；本院与 MOVER）。输出 reports/h1_subset_acc.md、h1_subset_acc.json"""
import os, json, numpy as np, pandas as pd
from sklearn.metrics import cohen_kappa_score as ck
D = os.path.expanduser("~/research/NE_ERAS_JEV/data/derived"); R = os.path.expanduser("~/research/Anes_JEV_vs_LoRA/analysis/reports"); PC = ["p1", "p2", "p3", "p4"]
K = lambda y, p: ck(y, p, weights="quadratic"); rd = lambda f: pd.read_csv(f"{D}/{f}.csv").set_index("op_id"); Ex = lambda a: (np.asarray(a) * np.arange(1, 5)).sum(1)
def cf(E, y):
    cu = np.array([1.5, 2.5, 3.5]); g = np.linspace(1, 4, 121)
    for _ in range(3):
        for j in range(3): cu[j] = max(g, key=lambda v: K(y, 1 + (E[:, None] > np.where(np.arange(3) == j, v, cu)).sum(1)))
    return cu
def boot(pid, fn, B=1000, seed=6):
    up = np.unique(pid); ix = {u: np.where(pid == u)[0] for u in up}; rng = np.random.default_rng(seed); return np.array([fn(np.concatenate([ix[u] for u in rng.choice(up, len(up))])) for _ in range(B)])
dist = lambda a: " / ".join(f"{x:.1f}" for x in np.bincount(a - 1, minlength=4)[:4] / len(a) * 100)
out = ["# H1 子集的可比性与准确率配对差\n"]; N = {}
# ---- 本院 ----
c = pd.read_csv(f"{D}/asa_common_local.csv", usecols=["op_id", "split", "asa4", "age", "female"]).merge(pd.read_csv(f"{D}/asa_dataset_v1.csv", usecols=["operation_id", "patient_sn"]).rename(columns={"operation_id": "op_id"}), on="op_id").set_index("op_id")
val = c[c.split == "val"].index; g = rd("pred_gen_local_val"); gsub = [i for i in g.index if i in set(val)]; r = rd("pred_llm_local")
rows = []
for nm, ids in (("H1 子集（随机 1,000）", gsub), ("未纳入", [i for i in val if i not in set(gsub)]), ("全部验证集", list(val))):
    v = c.loc[ids]; y = v.asa4.astype(int).values; p = r.loc[ids][PC].values.argmax(1) + 1; rows.append((nm, len(ids), f"{v.age.median():.0f}", f"{100 * (v.female == 1).mean():.1f}%", dist(y), f"{K(y, p):.3f}"))
out += ["## 本院验证集", "", "| 组 | n | 年龄中位数 | 女性 | ASA I/II/III/IV–V (%) | 读出 κ（未校准） |", "|---|---|---|---|---|---|"] + ["| " + " | ".join(map(str, x)) + " |" for x in rows]
gp = g.dropna(subset=["pred"]); N["local_gen_parsable"] = int(len(gp)); N["local_gen_n"] = int(len(g)); N["local_rows"] = rows
# ---- MOVER ----
m = pd.read_csv(f"{D}/asa_common_mover.csv", usecols=["op_id", "asa4", "age", "female"]).set_index("op_id"); sub = pd.read_csv(f"{D}/mover_test_sub4000.csv").op_id.tolist(); t = rd("pred_llm_mover_test"); gm = rd("pred_gen_mover_sub")
rows = []
for nm, ids in (("H1 子集（前 2,000）", sub[:2000]), ("未纳入（后 2,000）", sub[2000:]), ("评价子集 4,000", sub)):
    v = m.loc[ids]; y = v.asa4.astype(int).values; p = t.loc[ids][PC].values.argmax(1) + 1; rows.append((nm, len(ids), f"{v.age.median():.0f}", f"{100 * (v.female == 1).mean():.1f}%", dist(y), f"{K(y, p):.3f}"))
out += ["", "## MOVER 评价子集", "", "| 组 | n | 年龄中位数 | 女性 | ASA I/II/III/IV–V (%) | 读出 κ（未校准） |", "|---|---|---|---|---|---|"] + ["| " + " | ".join(map(str, x)) + " |" for x in rows]
N["mover_rows"] = rows; N["mover_gen_parsable"] = int(gm.dropna(subset=["pred"]).shape[0]); N["mover_gen_n"] = int(len(gm))
# ---- 准确率配对差 ----
out += ["", "## 准确率：LoRA（3 种子均值）− 1,000 标签校准读出（池 11），95% CI 为患者级 bootstrap", "", "| 数据 | LoRA n | LoRA 准确率 | 校准读出准确率 | 差值（95% CI） |", "|---|---|---|---|---|"]
cal = list(c[c.split == "dev"].reset_index().op_id.sample(1000, random_state=11)); yv = c.loc[val].asa4.astype(int).values; cu = cf(Ex(r.loc[cal][PC].values), c.loc[cal].asa4.astype(int).values); calp = 1 + (Ex(r.loc[val][PC].values)[:, None] > cu).sum(1); pid = c.loc[val].patient_sn.values
N["acc"] = {}
for n in (100, 300, 1000):
    L = [rd(f"pred_lora_LOC{n}s{sd}_local_val").loc[val][PC].values.argmax(1) + 1 for sd in ("20261017", "11", "12")]
    fn = lambda s: np.mean([(yv[s] == l[s]).mean() for l in L]) - (yv[s] == calp[s]).mean(); v = boot(pid, fn); pt = fn(np.arange(len(yv)))
    out.append(f"| 本院 | {n} | {np.mean([(yv == l).mean() for l in L]):.3f} | {(yv == calp).mean():.3f} | {pt:+.3f}（{np.percentile(v, 2.5):+.3f} ~ {np.percentile(v, 97.5):+.3f}） |"); N["acc"][f"local_{n}"] = [round(pt, 3), round(np.percentile(v, 2.5), 3), round(np.percentile(v, 97.5), 3)]
pm = pd.read_csv(f"{D}/mover_pid_map.csv").set_index("op_id").pid; ym = m.loc[sub].asa4.astype(int).values; calm = pd.read_csv(f"{D}/pred_llm_mover_cal.csv").op_id.tolist(); yc = m.loc[calm].asa4.astype(int).values
cum = cf(Ex(rd("pred_llm_mover_cal").loc[calm][PC].values), yc); calq = 1 + (Ex(t.loc[sub][PC].values)[:, None] > cum).sum(1); pidm = pm.loc[sub].values
for n in (100, 300, 1000):
    L = [rd(f"pred_lora_curve{n}{sfx}_mover_sub").loc[sub][PC].values.argmax(1) + 1 for sfx in ("", "s11", "s12")]
    fn = lambda s: np.mean([(ym[s] == l[s]).mean() for l in L]) - (ym[s] == calq[s]).mean(); v = boot(pidm, fn); pt = fn(np.arange(len(ym)))
    out.append(f"| MOVER 子集 | {n} | {np.mean([(ym == l).mean() for l in L]):.3f} | {(ym == calq).mean():.3f} | {pt:+.3f}（{np.percentile(v, 2.5):+.3f} ~ {np.percentile(v, 97.5):+.3f}） |"); N["acc"][f"mover_{n}"] = [round(pt, 3), round(np.percentile(v, 2.5), 3), round(np.percentile(v, 97.5), 3)]
open(f"{R}/h1_subset_acc.md", "w").write("\n".join(out)); json.dump(N, open(f"{R}/h1_subset_acc.json", "w"), ensure_ascii=False, indent=1, default=str); print("\n".join(out))
```
