## Supplementary material for "Non-generative readout with few-label calibration versus LoRA fine-tuning of a large language model for ASA physical status classification: accuracy, speed, cost and label efficiency in three data sources": Analysis code and aggregate results: dl_mover_parallel.py

```
"""MOVER 分段并行下载（走本机代理，服务器限制单连接速度）。凭据读 netrc，不打印。公钥固定替代证书校验（服务器缺根证书）。
用法：dl_mover_parallel.py <文件名> <目标目录> <总字节> <期望md5> [并行数=16] [分段MB=128]"""
import os, subprocess, sys, time, hashlib, concurrent.futures as cf
name, dest, total, md5x = sys.argv[1], sys.argv[2], int(sys.argv[3]), sys.argv[4]; W = int(sys.argv[5]) if len(sys.argv) > 5 else 16
CHMB = int(sys.argv[6]) if len(sys.argv) > 6 else 128; CH = CHMB * 1024 * 1024; parts = f"{dest}/parts_{name}_{CHMB}m"; os.makedirs(parts, exist_ok=True)
PIN = open("/tmp/a4s/mover.pin").read().strip(); NETRC = os.path.expanduser("~/research/NE_ERAS_JEV/data/private/mover.netrc")
URL = f"https://mover-download.ics.uci.edu/{name}"
chunks = [(i, i * CH, min(total, (i + 1) * CH) - 1) for i in range((total + CH - 1) // CH)]
def fetch(c):
    i, a, b = c; fp = f"{parts}/{i:04d}"; need = b - a + 1
    for t in range(200):
        have = os.path.getsize(fp) if os.path.exists(fp) else 0
        if have == need: return i
        if have > need: os.remove(fp); have = 0
        subprocess.run(["curl", "-s", "-m", "300", "-k", "--pinnedpubkey", PIN, "--netrc-file", NETRC, "-r", f"{a + have}-{b}", "-o", fp + ".tmp", URL], timeout=330)
        if os.path.exists(fp + ".tmp"):
            with open(fp, "ab") as o, open(fp + ".tmp", "rb") as s: o.write(s.read())
            os.remove(fp + ".tmp")
        time.sleep(1)
    raise RuntimeError(f"chunk {i} failed")
t0 = time.time()
with cf.ThreadPoolExecutor(W) as ex:
    done = 0
    for _ in ex.map(fetch, chunks):
        done += 1
        if done % 5 == 0 or done == len(chunks): print(f"{done}/{len(chunks)} chunks, {time.time() - t0:.0f}s", flush=True)
out = f"{dest}/{name}"; h = hashlib.md5()
with open(out, "wb") as o:
    for c in chunks:
        with open(f"{parts}/{c[0]:04d}", "rb") as s:
            while b := s.read(1 << 24): o.write(b); h.update(b)
print("size", os.path.getsize(out), "expected", total, "| md5", h.hexdigest(), "OK" if h.hexdigest() == md5x else "MISMATCH", flush=True)
if h.hexdigest() == md5x: import shutil; shutil.rmtree(parts)
```
