## Supplementary material for "Non-generative readout with few-label calibration versus LoRA fine-tuning of a large language model for ASA physical status classification: accuracy, speed, cost and label efficiency in three data sources": Analysis code and aggregate results: hf.py

```
"""Hugging Face ``transformers`` backend.

Every candidate completion of every question in a request is scored in one
batched forward pass (chunked only when it would not fit the token budget).
Because all questions share the same *state*, the common token prefix is run
once and its KV-cache is broadcast across the batch, so adding questions adds
roughly the cost of their (short) suffixes rather than another pass over the
state.

No token is ever sampled: the answer is read straight off the log-probabilities
of the closed candidate set, which is the whole point of a System One model.
"""

from __future__ import annotations

import copy
import logging
from dataclasses import dataclass
from typing import Any, Sequence

import torch

from ..prompting import Prompt, disambiguate_candidate_tokens
from .base import ScoringResult, ScoringTask

logger = logging.getLogger(__name__)


@dataclass
class _Seq:
    task: int
    cand: int
    prefix: list[int]
    cand_ids: list[int]

    @property
    def full(self) -> list[int]:
        return self.prefix + self.cand_ids


class HFPromptRenderer:
    """Turn :class:`Prompt` messages and candidate labels into token ids for a given tokenizer.

    Shared by the inference backend and the training script so both see exactly
    the same token sequences.
    """

    def __init__(self, tokenizer) -> None:
        self.tokenizer = tokenizer
        self.chat = bool(getattr(tokenizer, "chat_template", None))
        self._system_ok = True
        self.terminator = self._pick_terminator()

    def _pick_terminator(self) -> int | None:
        """Token appended to candidates when one label is a prefix of another ("1" vs "12")."""
        if self.chat:
            # Probe the template for its end-of-turn token: whatever follows an assistant message.
            try:
                probe = self.tokenizer.apply_chat_template(
                    [{"role": "user", "content": "u"}, {"role": "assistant", "content": "a"}],
                    tokenize=False,
                )
                head = self.tokenizer.apply_chat_template(
                    [{"role": "user", "content": "u"}], tokenize=False, add_generation_prompt=True
                )
                if probe.startswith(head):
                    tail = probe[len(head):]
                    if tail.startswith("a"):
                        ids = self.tokenizer(tail[1:], add_special_tokens=False).input_ids
                        if ids:
                            return ids[0]
            except Exception:  # pragma: no cover - template quirks
                pass
        if self.tokenizer.eos_token_id is not None:
            return self.tokenizer.eos_token_id
        nl = self.tokenizer("\n", add_special_tokens=False).input_ids
        return nl[0] if nl else None

    def render_prompt(self, prompt: Prompt) -> list[int]:
        if self.chat:
            messages = [{"role": "system", "content": prompt.system}, {"role": "user", "content": prompt.user}]
            if not self._system_ok:
                messages = [{"role": "user", "content": f"{prompt.system}\n\n{prompt.user}"}]
            try:
                text = self.tokenizer.apply_chat_template(messages, tokenize=False, add_generation_prompt=True, enable_thinking=False)
            except Exception:
                # Templates without a system role (e.g. some Gemma builds) raise; fold it into the user turn.
                self._system_ok = False
                messages = [{"role": "user", "content": f"{prompt.system}\n\n{prompt.user}"}]
                text = self.tokenizer.apply_chat_template(messages, tokenize=False, add_generation_prompt=True, enable_thinking=False)
            return self.tokenizer(text, add_special_tokens=False).input_ids
        text = f"{prompt.system}\n\n{prompt.user}\nAnswer:"
        return self.tokenizer(text, add_special_tokens=True).input_ids

    def candidate_ids(self, candidates: Sequence[str]) -> list[list[int]]:
        ids = [
            self.tokenizer(c if self.chat else " " + c, add_special_tokens=False).input_ids
            for c in candidates
        ]
        return disambiguate_candidate_tokens(ids, self.terminator)


class HFBackend:
    def __init__(
        self,
        model_id: str,
        *,
        device: str | None = None,
        dtype: torch.dtype | str | None = None,
        device_map: str | dict | None = None,
        trust_remote_code: bool = False,
        max_batch_tokens: int | None = None,
        share_prefix: bool = True,
        min_shared_prefix: int = 8,
        tokenizer_id: str | None = None,
        adapter: str | None = None,
        quantization_config: Any = None,   # [本项目补丁] 传给 from_pretrained，用于 4/8-bit 量化加载
    ) -> None:
        from transformers import AutoModelForCausalLM, AutoTokenizer

        self.name = model_id
        self.share_prefix = share_prefix
        self.min_shared_prefix = min_shared_prefix

        if device is None:
            device = "cuda" if torch.cuda.is_available() else "cpu"
        self.device = torch.device(device)

        if isinstance(dtype, str):
            dtype = getattr(torch, dtype)
        if dtype is None:
            if self.device.type == "cuda" and torch.cuda.is_bf16_supported():
                dtype = torch.bfloat16
            else:
                dtype = torch.float32
        self.dtype = dtype

        if max_batch_tokens is None:
            max_batch_tokens = 16384 if self.device.type == "cuda" else 2048
        self.max_batch_tokens = max_batch_tokens

        self.tokenizer = AutoTokenizer.from_pretrained(tokenizer_id or model_id, trust_remote_code=trust_remote_code)
        if self.tokenizer.pad_token_id is None:
            self.tokenizer.pad_token = self.tokenizer.eos_token

        load_kwargs: dict[str, Any] = {"trust_remote_code": trust_remote_code}
        if quantization_config is not None: load_kwargs["quantization_config"] = quantization_config
        try:
            load_kwargs["dtype"] = dtype
            self.model = AutoModelForCausalLM.from_pretrained(model_id, device_map=device_map, **load_kwargs)
        except TypeError:  # older transformers spell it torch_dtype
            load_kwargs.pop("dtype")
            self.model = AutoModelForCausalLM.from_pretrained(model_id, device_map=device_map, torch_dtype=dtype, **load_kwargs)
        if adapter:
            # A LoRA adapter produced by scripts/train_calibrated.py; merged so scoring stays a plain forward.
            from peft import PeftModel

            self.model = PeftModel.from_pretrained(self.model, adapter).merge_and_unload()
            self.name = f"{model_id}+{adapter}"
        if device_map is None:
            self.model.to(self.device)
        else:
            # With a device map the inputs go wherever the embedding layer lives.
            self.device = self.model.get_input_embeddings().weight.device
        self.model.eval()

        self.renderer = HFPromptRenderer(self.tokenizer)

    def render_prompt(self, prompt: Prompt) -> list[int]:
        return self.renderer.render_prompt(prompt)

    def candidate_ids(self, candidates: Sequence[str]) -> list[list[int]]:
        return self.renderer.candidate_ids(candidates)

    # ------------------------------------------------------------------ scoring

    @torch.inference_mode()
    def score(self, tasks: Sequence[ScoringTask]) -> list[ScoringResult]:
        seqs: list[_Seq] = []
        prompt_tokens: list[int] = []
        for ti, task in enumerate(tasks):
            prefix = self.render_prompt(task.prompt)
            prompt_tokens.append(len(prefix))
            for ci, cand in enumerate(self.candidate_ids(task.candidates)):
                seqs.append(_Seq(task=ti, cand=ci, prefix=prefix, cand_ids=cand))

        shared = self._shared_prefix_len(seqs) if self.share_prefix else 0
        if shared:
            try:
                logprobs = self._score_with_shared_prefix(seqs, shared)
            except Exception as exc:  # cache API differences across transformers versions
                logger.warning("shared-prefix scoring failed (%s); falling back to full sequences", exc)
                logprobs = self._score_full(seqs)
        else:
            logprobs = self._score_full(seqs)

        results: list[ScoringResult] = []
        for ti, task in enumerate(tasks):
            lp = [0.0] * len(task.candidates)
            ntok = 0
            for s, v in zip(seqs, logprobs):
                if s.task == ti:
                    lp[s.cand] = v
                    ntok += len(s.cand_ids)
            results.append(ScoringResult(logprobs=lp, prompt_tokens=prompt_tokens[ti], candidate_tokens=ntok))
        return results

    def _shared_prefix_len(self, seqs: list[_Seq]) -> int:
        if len(seqs) < 2:
            return 0
        first = seqs[0].prefix
        n = min(len(s.prefix) for s in seqs)
        for i in range(n):
            tok = first[i]
            if any(s.prefix[i] != tok for s in seqs):
                n = i
                break
        # Keep at least the final prefix token in the suffix so the logits that predict
        # the first candidate token are produced in the suffix pass.
        n = min(n, min(len(s.prefix) for s in seqs) - 1)
        return n if n >= self.min_shared_prefix else 0

    def _batches(self, seqs: list[_Seq], length_of) -> list[list[int]]:
        order = sorted(range(len(seqs)), key=lambda i: length_of(seqs[i]))
        batches: list[list[int]] = []
        cur: list[int] = []
        cur_max = 0
        for i in order:
            n = length_of(seqs[i])
            new_max = max(cur_max, n)
            if cur and new_max * (len(cur) + 1) > self.max_batch_tokens:
                batches.append(cur)
                cur, cur_max = [], 0
                new_max = n
            cur.append(i)
            cur_max = new_max
        if cur:
            batches.append(cur)
        return batches

    def _pad(self, rows: list[list[int]]) -> tuple[torch.Tensor, torch.Tensor]:
        width = max(len(r) for r in rows)
        pad = self.tokenizer.pad_token_id
        ids = torch.full((len(rows), width), pad, dtype=torch.long)
        mask = torch.zeros((len(rows), width), dtype=torch.long)
        for i, r in enumerate(rows):
            ids[i, : len(r)] = torch.tensor(r, dtype=torch.long)
            mask[i, : len(r)] = 1
        return ids.to(self.device), mask.to(self.device)

    def _hidden_then_logits(
        self,
        input_ids: torch.Tensor,
        attention_mask: torch.Tensor,
        gather: list[tuple[int, list[int]]],
        *,
        position_ids: torch.Tensor | None = None,
        past_key_values=None,
    ) -> list[torch.Tensor]:
        """Run the decoder and apply the LM head only at the positions we need.

        ``gather`` lists, per batch row, the positions whose next-token logits are
        required. Applying the head to a handful of rows instead of the whole
        ``batch x seq x vocab`` tensor keeps memory flat regardless of vocab size.
        """
        decoder = self.model.get_decoder() if hasattr(self.model, "get_decoder") else None
        head = self.model.get_output_embeddings()
        kwargs: dict[str, Any] = {"attention_mask": attention_mask}
        if position_ids is not None:
            kwargs["position_ids"] = position_ids
        if past_key_values is not None:
            kwargs["past_key_values"] = past_key_values
            kwargs["use_cache"] = True
        else:
            kwargs["use_cache"] = False

        if decoder is not None and head is not None and not getattr(self.model.config, "final_logit_softcapping", None):
            hidden = decoder(input_ids=input_ids, **kwargs).last_hidden_state
            out = []
            for row, positions in gather:
                out.append(head(hidden[row, positions]).float().log_softmax(-1))
            return out
        logits = self.model(input_ids=input_ids, **kwargs).logits
        return [logits[row, positions].float().log_softmax(-1) for row, positions in gather]

    def _score_full(self, seqs: list[_Seq]) -> list[float]:
        out = [0.0] * len(seqs)
        for batch in self._batches(seqs, lambda s: len(s.full)):
            rows = [seqs[i].full for i in batch]
            ids, mask = self._pad(rows)
            gather = []
            for r, i in enumerate(batch):
                s = seqs[i]
                start = len(s.prefix) - 1
                gather.append((r, list(range(start, start + len(s.cand_ids)))))
            lps = self._hidden_then_logits(ids, mask, gather)
            for r, i in enumerate(batch):
                targets = torch.tensor(seqs[i].cand_ids, device=lps[r].device)
                out[i] = float(lps[r].gather(1, targets[:, None]).sum())
        return out

    def _score_with_shared_prefix(self, seqs: list[_Seq], shared: int) -> list[float]:
        prefix_ids = torch.tensor([seqs[0].prefix[:shared]], dtype=torch.long, device=self.device)
        prefix_out = self.model(input_ids=prefix_ids, use_cache=True)
        cache = prefix_out.past_key_values

        out = [0.0] * len(seqs)
        for batch in self._batches(seqs, lambda s: len(s.full) - shared):
            rows = [seqs[i].full[shared:] for i in batch]
            ids, mask = self._pad(rows)
            b, width = ids.shape
            full_mask = torch.cat([torch.ones((b, shared), dtype=mask.dtype, device=mask.device), mask], dim=1)
            position_ids = (shared + torch.arange(width, device=self.device))[None, :].expand(b, -1)
            gather = []
            for r, i in enumerate(batch):
                s = seqs[i]
                start = len(s.prefix) - 1 - shared
                gather.append((r, list(range(start, start + len(s.cand_ids)))))
            lps = self._hidden_then_logits(
                ids, full_mask, gather, position_ids=position_ids, past_key_values=_expand_cache(cache, b)
            )
            for r, i in enumerate(batch):
                targets = torch.tensor(seqs[i].cand_ids, device=lps[r].device)
                out[i] = float(lps[r].gather(1, targets[:, None]).sum())
        return out


def _expand_cache(cache, batch: int):
    """Broadcast a batch-1 KV cache to ``batch`` rows without touching the original."""
    if batch == 1:
        return copy.deepcopy(cache)
    if hasattr(cache, "batch_repeat_interleave"):
        expanded = copy.deepcopy(cache)
        expanded.batch_repeat_interleave(batch)
        return expanded
    if hasattr(cache, "to_legacy_cache"):
        from transformers import DynamicCache

        legacy = cache.to_legacy_cache()
        expanded = tuple(tuple(t.expand(batch, *t.shape[1:]).contiguous() for t in layer) for layer in legacy)
        return DynamicCache.from_legacy_cache(expanded)
    if isinstance(cache, tuple):
        return tuple(tuple(t.expand(batch, *t.shape[1:]).contiguous() for t in layer) for layer in cache)
    raise TypeError(f"unsupported cache type {type(cache)!r}")
```
