## Supplementary material for "Non-generative readout with few-label calibration versus LoRA fine-tuning of a large language model for ASA physical status classification: accuracy, speed, cost and label efficiency in three data sources": Analysis code and aggregate results: PATCH_NOTES.md

Upstream: GitHub30/OpenJev, commit aae11e1b56e9472c1e67b231a717e70709f6a3a6 (2026-09-20), MIT (see LICENSE_OpenJev_MIT).
File: src/openjev/backends/hf.py (patched copy).
Patch 1: HFBackend.__init__ gains a quantization_config argument passed to from_pretrained (4-bit loading).
Patch 2: render_prompt passes enable_thinking=False to apply_chat_template (the Qwen3.5 template otherwise opens with a thinking marker, which degrades the option-label probabilities).
