## Supplementary material for "Non-generative readout with few-label calibration versus LoRA fine-tuning of a large language model for ASA physical status classification: accuracy, speed, cost and label efficiency in three data sources": Analysis code and aggregate results: README.md

# Code and aggregate results

Analysis code for "Non-generative readout with few-label calibration versus LoRA fine-tuning of a large language model for ASA physical status classification: accuracy, speed, cost and label efficiency in three data sources".

## What is and is not included
- **Included**: all analysis scripts (`analysis_scripts/`), the patched file of the OpenJev implementation (`openjev_patch/`), aggregate result tables (`aggregate_results/reports/`), the SHA-256 checksums of the Qwen3.5-9B weight files, and the 40 manually checked rationales with hashed operation identifiers.
- **Not included**: patient-level data of the local hospital (cannot be shared), MOVER data (requires a data use agreement from the providers: https://doi.org/10.1093/jamiaopen/ooad084), VitalDB data (public, https://doi.org/10.13026/czw8-9p62; CC BY-NC-SA 4.0), model weights and prediction files.
- Paths: scripts read their inputs from a project directory (`~/research/Anes_JEV_vs_LoRA` and sibling data directories) that must be recreated by the user after obtaining the data.

## Environment
See `requirements.txt` (versions used). Hardware: two NVIDIA GeForce RTX 2080 Ti with 22 GB each (compute capability 7.5, fp16 only). Base model: Hugging Face `Qwen/Qwen3.5-9B` (Apache-2.0), downloaded 2026-10-16.

## Order of the pipeline (script prefix = order of development)
| Step | Scripts |
|---|---|
| Build datasets and the T1/T2 templates | 01–05, 17, 36, 60 |
| Zero-shot readout and generative zero-shot | 06, 07, 13, 21 |
| Task LoRA (4-bit QLoRA) | 08, 111, 112 |
| XGBoost and kNN arms | 09, 15 |
| OpenJev tuning and frozen runs | 30–34, 50–55 |
| Cutpoint calibration and curves | 35, 41, 82 |
| Evaluation, paired differences and bootstrap | 10, 14, 20, 22, 87, 87b |
| Speed, tokens, latency | 18, 97, 98 |
| Robustness: option order, label format, permutations, 4-bit readout, label matching | 24, 26, 38, 54, 103, 105, 105b, 108, 113–115 |
| Subgroups, break-even, adjacent errors, H1 subset | 99, 106, 109, 116 |
| Natural-language guidance | 83, 85 |
| Hallucination screen | 81 |
| Figures and tables | 80, 86, 93–96, 107 |

## OpenJev
Upstream: https://github.com/GitHub30/OpenJev, commit aae11e1b56e9472c1e67b231a717e70709f6a3a6 (2026-09-20), MIT license. Two patches were applied to `src/openjev/backends/hf.py` (file provided in `openjev_patch/`): (1) `HFBackend.__init__` accepts a `quantization_config` argument and passes it to `from_pretrained`; (2) `render_prompt` calls `apply_chat_template` with `enable_thinking=False`, because the Qwen3.5 template otherwise starts with a thinking marker and degrades the output. Clone the upstream repository at the commit above and replace this file.

## Prompts
The full prompt texts of every arm are in the supplementary material (S6) and in scripts 07, 13, 33 and 83.
